# The gut microbiota and colonic pH map to the peripheral immune landscape in humans

**DOI:** 10.64898/2025.12.02.25341510

**Authors:** Evany Dinakis, Dakota Rhys-Jones, Leticia Camargo Tavares, Chaoran Yang, Liang Xie, CK Yao, Daniel So, Dovile Anderson, Darren Creek, Stephen J. Turner, Peter R. Gibson, Charles R. Mackay, Joanne A. O’Donnell, Jane Muir, Francine Z. Marques

## Abstract

**Background:** The regional physicochemical environment of the gut, particularly colonic pH, is a key determinant of microbial composition and metabolic activity. However, how these localised intraluminal physicochemical dynamics interface with systemic immunity in humans remains unknown.

**Objective:** We aimed to investigate the relationship between colonic pH, microbial metabolism, and immune tone.

**Design:** In a cross-sectional study of 54 healthy adults, we integrated four complementary datasets to enable a systems-level analysis of gut–immune interactions: *in vivo* real-time colonic pH and transit dynamics via the SmartPill wireless pH–motility capsule; microbial community structure via faecal metagenomic sequencing; microbial metabolic activity via plasma short-chain fatty acid (SCFA) quantification; and 121 circulating immune subsets via deep immunophenotyping of peripheral blood mononuclear cells (PBMCs).

**Results:** Lower colonic pH was associated with circulating cytokine-producing CD8^+^ T cells. Conversely, higher pH correlated with an increase in circulating mature low-density neutrophils (LDNs) and a distinct shift towards PD1-expressing exhaustion-like CD8^+^ T cells. Metagenomic profiling identified species-level associations within the genus *Eubacterium*, linked to colonic pH and distinct peripheral immune profiles.

**Conclusion:** These findings suggest that gut’s physicochemical microenvironment, driven by pH, may act alongside microbial activity and metabolism to jointly shape systemic immune tone in humans. This supports a functional intraluminal-peripheral axis, suggesting that targeting gut pH and fermentation dynamics could offer novel avenues for exploring immune-targeted interventions.

**What is already known on this topic:**

- Colonic pH is a critical driver of microbial niche selection and metabolic activity *in vitro*, but how this localised physicochemical environment influences systemic immunity in humans remains unknown.

**What this study adds?:**

- By pairing *in vivo* real-time wireless pH-capsule transit data with metagenomics, plasma SCFAs, and deep immunophenotyping, we demonstrate specific associations between colonic pH and the peripheral immune landscape, mapping gut–immune interactions in humans
- Lower colonic pH associates with robust, cytokine-producing CD8^+^ T cells, whereas a higher pH correlates with a shift toward PD1 exhaustion-like CD8^+^ T cell phenotypes and elevated systemic mature LDNs.

**How this study might affect research, practice or policy:**

- These findings establish a novel physicochemical-gut-immune axis, suggesting that therapeutic manipulation of colonic pH (via diet, prebiotics, or targeted delivery systems) could serve as a strategy to modulate or support systemic immune tone in inflammatory or metabolic diseases.

## Introduction

The gastrointestinal (GI) tract orchestrates tightly regulated physiological gradients, among which pH plays a central role in shaping microbial^1^ and immune dynamics^2^. pH dynamics across the GI tract are markedly heterogenous^2,3^. The large intestine presents a mildly acidic to neutral pH (5.5–7), with the proximal colon maintaining a more acidic environment (pH ∼5.5-6.0), whereas the distal colonic luminal pH is typically more alkaline (∼6.5-7.0)^4,5^. This spatial variation parallels the distribution of the gut microbiota^6–8^. Microbial density is low in the upper GI tract but peaks in the colon, which harbours the most diverse and metabolically active communities, thus serving as a key site for host-microbiota interactions^9,10^.

Colonic pH is shaped by microbial metabolic activity^11,12^, while pH itself influences the composition and activity of the gut microbiota^13,14^. This bidirectional interaction, therefore, tightly links microbial function with the physicochemical environment of the colon.^1^ Diet is a key modulator of this interaction; diets rich in complex carbohydrates such as fibre, for example, enhance microbial diversity and fermentation, leading to the production of short-chain fatty acids (SCFAs)^15,16^. These acidic metabolites lower colonic luminal and interstitial fluid pH^17^, making colonic pH a dynamic parameter responsive to both microbial and dietary inputs.

The GI tract is also a major immunological organ^18^, with extensive crosstalk between resident microbes and host immunity. Studies using germ-free animal models revealed that the gut microbiota is essential for the maturation of both intestinal and systemic immunity^19,20^, leading to impaired lymphoid tissue development and T cell function, and reduced immunoglobulin production^21–23^. Yet, most mechanistic insights have largely centred on mucosal immunity^24^ and experimental models.^6^ The extent to which the gut microbiota shapes the peripheral immune system in humans remains poorly understood.

Here, we present an integrated systems-level analysis of host microbiota–intestinal pH–immune interactions in a human cohort, investigating whether colonic pH shapes systemic immune tone through microbial metabolic pathways. We hypothesised that lower colonic pH shapes systemic immune tone through microbial functional pathways and functionally distinct immune phenotypes arising from microbiota□derived metabolic activity. By combining faecal shotgun metagenomic sequencing, *in vivo* colonic intraluminal pH profiling using the SmartPill wireless pH-motility capsule, and deep immunophenotyping of peripheral blood mononuclear cells (PBMCs), our findings indicate that colonic pH may indeed have an important effect on peripheral immune tone. In particular, *Eubacterium* species and their metabolic input in amino acid and nucleotide metabolism and biosynthesis pathways were associated with differences in peripheral immune profiles, suggesting that colonic pH may reflect a functional interface between the microbiota and systemic immunity.

## Results

### Mapping colonic pH gradients using a wireless pH-motility capsule (SmartPill)

Fifty-four healthy participants completed the trial, of whom 41% were female. Participants had a mean age of 49 ± 12.9 years and body mass index (BMI) of 27.0 ± 3.7 kg□m^-2^ (Figure 1, Table S1). The SmartPill software provides median, minimum, and maximum pH values – these provide proxy values for different intestinal sites. The minimum colonic value indicates the ascending colon, while the maximum colonic pH indicates distal colonic and sigmoid/rectum pH (Figure 2A-B). To map the heterogeneity of luminal pH across different colonic regions, we divided the colon into four equal quartiles, as previously reported^10^. This was performed based on transit time by dividing the total time in the colon into four equal quartiles, with the assumption that transit time would have been the same across all segments of the colon. These quartiles are thought to approximately correspond to the main regions of the large intestine (Figure 2A). Our data fitted the expected pattern of pH distribution along the colon (Figure 2B), whereby colonic minimum pH was significantly correlated with first and second quartile intraluminal pH (Figures 2C-D), colonic median pH was significantly correlated with first to third quartiles (Figures 2E-G), and colonic maximum pH was correlated with second to fourth quartiles (Figures 2H-J).

**Figure 1.**
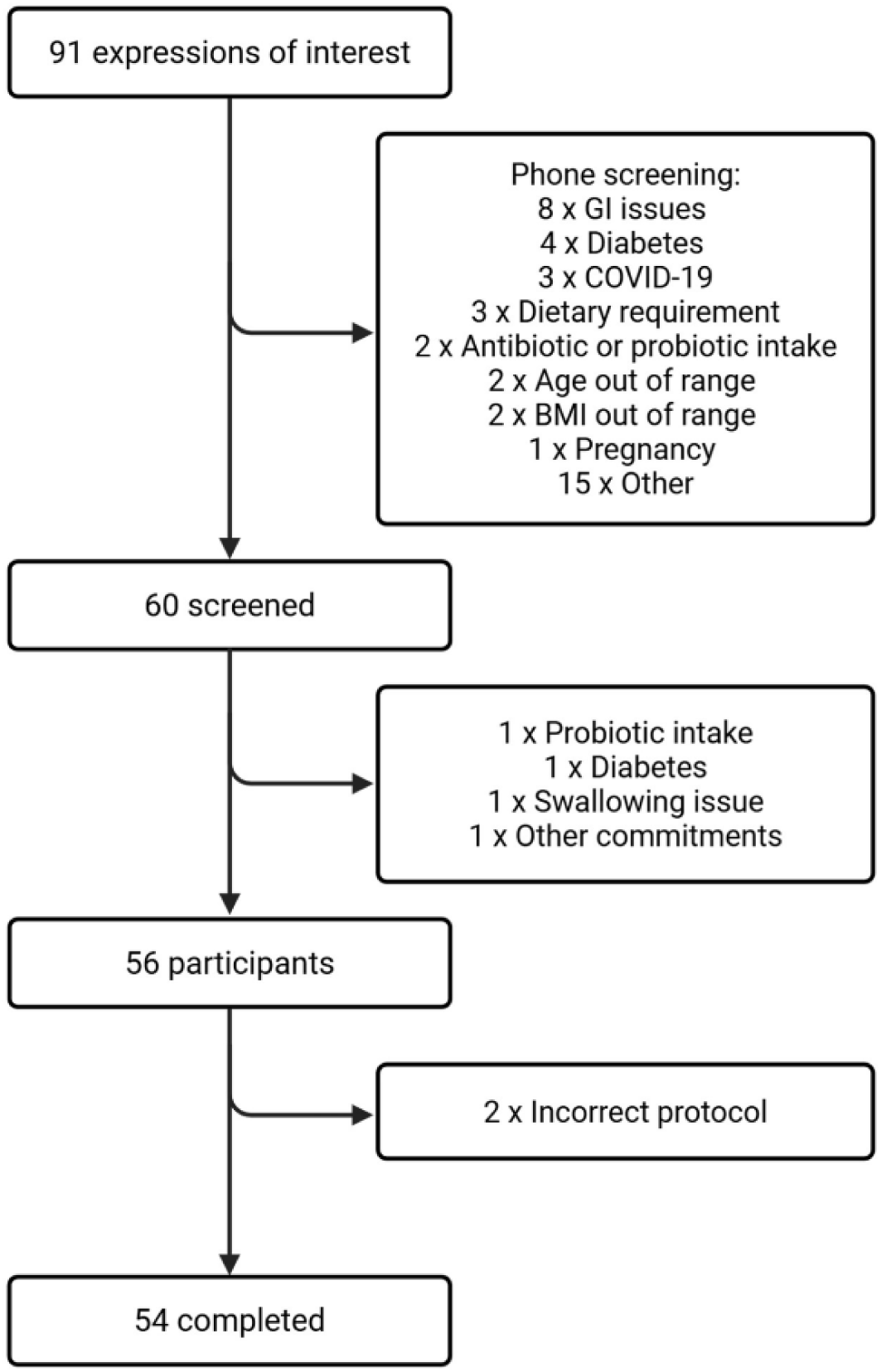
Recruitment and cohort characteristics. Recruitment summary of the pH of Intestines and Blood-pressure Regulation (pHibre) study; out of 91 expressions of interest, 54 participants finished the trial. GI, gastrointestine; COVID-19, coronavirus disease; BMI, body mass index.

**Figure 2.**
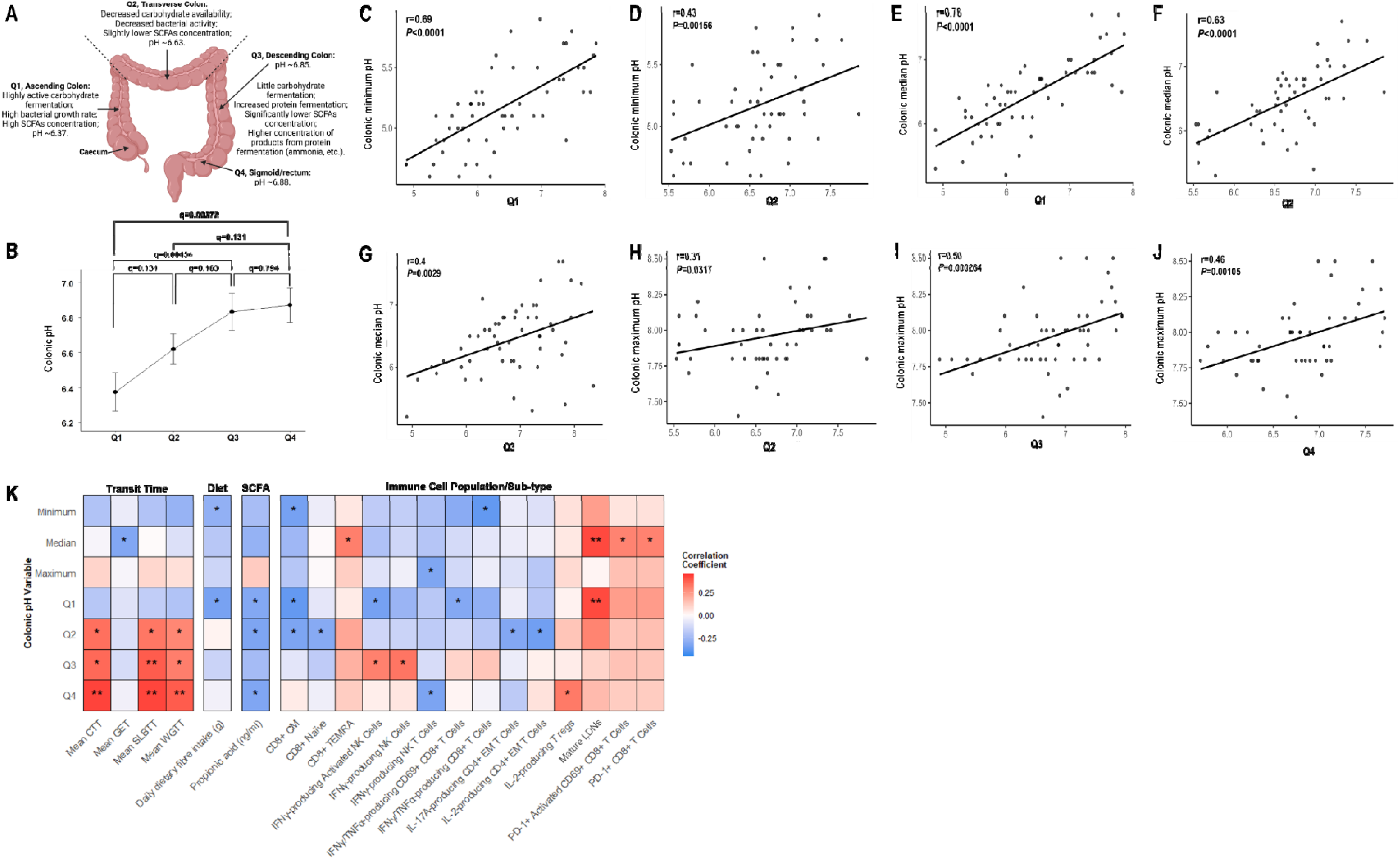
Colonic pH and associations with immune cells, dietary fibre and short-chain fatty acids (SCFAs). **(A)** Graphic scheme of the regional pH profile in the colon and respective bacterial activity and SCFA concentration, adapted from^11,93^ and created on BioRender. **(B)** Regional pH profile in the colon of the pHibre cohort generated by the SmartPill. **(C-J)** Spearman’s rank correlations between colonic minimum, median and maximum pH and quartile data. **(K)** Spearman’s rank correlations as heatmaps between colonic pH variables and transit time, diet, plasma SCFAs and immune cell populations/sub-types. Comparisons between quartiles were performed using a one-way analysis of variance (ANOVA) followed by pairwise t-tests. Data presented as mean ± standard error of mean (SEM). Spearman’s rank correlations were performed on non-normally distributed continuous variables. The colour gradient represents the strength and direction of correlation (red = positive, blue = negative). Raw unadjusted P-values are presented in heatmaps; *P<0.05, **P<0.01. n=41-54. CTT, colonic transit time; GET, gastrointestinal emptying time; SLBTT, small and large bowel transit time; WGTT, whole gut transit time; SCFA, short-chain fatty acid; CM, central memory; TEMRA, terminally differentiated effector memory T cells; IFN-γ, interferon-γ; NK, natural killer; TNF-α, tumour necrosis factor-α; IL-, interleukin-; EM, effector memory; T regs, T regulatory cells; LDNs, low-density neutrophils; PD-1, programmed cell death protein 1.

### Colonic pH correlates with peripheral immune cells and SCFAs

Gut transit time is an important physiological variable that integrates nutrient absorption and microbial fermentation^25^, thus, its link with intraluminal pH represents an interesting avenue for investigation. Longer whole-gut (WGTT), small and large bowel (SLBTT) and colonic transit time (CTT) were associated with higher (more basic) pH in quartiles two through four, but not quartile one (Figure 2K). Higher daily dietary fibre intake was associated with lower, or more acidic, minimum and quartile one colonic pH (Figure 2K). The main by-product of fermentable fibre produced by the gut microbiota inhabiting the large intestine is SCFAs^26^, acidic immunomodulatory metabolites that can profoundly influence physiological outcomes such as colonic pH^17^. Only higher plasma propionic acid levels were associated with lower quartile one, two and four colonic pH (Figure 2K).

We further analysed potential relationships between colonic pH and 121 peripheral immune cell populations and subtypes, including activation and cytokine expression across these cell types. We identified varying associations with 14 different peripheral immune cell subsets across all seven colonic pH variables (Figure 2K). Overall, these suggest that colonic pH may be associated with peripheral immunomodulation. Ten associations were with the T cell lineage; for example, elevated numbers of CD8^+^ central memory T cells and a dual cytokine-producing (interferon-γ [IFNγ]/ tumour necrosis factor-α [TNFα]) population of activated CD8^+^ T cells were associated with lower colonic minimum pH. Conversely, programmed cell death protein-1 (PD-1)^+^ CD8^+^, activated PD-1^+^ CD8^+^ T cells, IFNγ-producing natural killer (NK) cells and mature LDNs^27^ were associated with more basic colonic pH.

Correlation analyses between plasma SCFA levels and the peripheral immune system further revealed 17 significant associations, of which 14 were negatively associated with isovaleric acid and were comprised almost entirely of CD4^+^ T cell subsets (Figure S1). Plasma butanoic acid was positively associated with B cells and PD-1^+^ CD8^+^ T cells, whilst lower plasma acetic and butanoic acid were associated with the production of both interleukin (IL)-2 and TNFα by CD161^+^ CD4^+^ T cells (Figure S1).

### A distinct subset of PD-1-expressing, exhaustion-like CD8^+^ T cells consistently underpins gut microbiota and colonic pH associations

Upon persistent antigen stimulation, such as during chronic infection or cancer, activated T cells can enter a state of functional decline known as exhaustion^28,29^. This transition is marked by sustained expression of inhibitory receptors, including PD-1 and TIM-3, and diminished proliferative capacity, often linked to impaired IL-2 signalling and reduced cytokine production (i.e., TNFα and IFNγ)^30–32^ (Figure S2A). For example, in melanoma patients, TIM-3^+^ PD-1^+^ tumour-specific CD8^+^T cells were more dysfunctional than TIM-3- PD-1^+^ and TIM-3- PD-1-T cells, producing less IFN-γ, TNF-α and IL-2^31^. However, T cells can enter a transitional pre-exhausted state, typically marked by reduced IL-2 production and consequently diminished proliferative capacity^33^. This state is characterized by elevated PD-1 expression in the absence of other inhibitory receptors such as TIM-3^34^, while retaining functional competence and the ability to produce effector cytokines^35,36^ (Figure S2A). We performed a sensitivity analysis focusing on the functionality of PD-1^+^ CD8^+^ T cells and found that these cells retained a pre-exhausted phenotype, as evidenced by a lack of TIM-3 and IL-2 expression (Figure S2B). Given the broader aim of integrating immune phenotypes with gut microbial features, we next examined whether this pre□exhausted CD8□T cell subset was associated with variation in gut microbial composition. Indeed, taxonomic profiling identified 14 differentially abundant taxa specifically positively associated with these pre-exhausted CD8^+^ T cells (Figure S2C).

### Colonic pH across anatomical segments is associated with the gut microbiota and associated metabolic pathways

To assess differences in the gut microbiota in relation to colonic pH measurements, we calculated α- and β-diversity metrics from shotgun metagenome data. We found no association between α-diversity and colonic pH (Table S2). However, β-diversity analyses revealed that quartile colonic pH was associated with differences in gut microbiota composition (Table S2). Based on unweighted UniFrac distances, quartile one colonic pH explained 3.5% of the observed variation in gut microbiota composition (P=0.014, Table S2), suggesting compositional differences characterised by the presence/absence of unique microbial taxa.

Taxonomic profiling analysis revealed that one bacterial species, *Blautia schinkii*, was associated with colonic pH in quartile four (q=0.0143; Figure S3). Functional profiling analysis revealed 169 positive associations between differentially abundant species-specific metabolic pathways and quartile four (Figure 3A). Eighty metabolic pathways, categorised by biological function, were stratified across eight bacterial species (Tables S3-S4, Figure 3A). We did not identify any significant *Blautia schinkii*-specific metabolic pathways associations with quartile four colonic pH. Our analysis instead showed that *Bifidobacterium pseudocatenulatum,* a species that thrives on complex carbohydrate fermentation^37^, exhibited the strongest associations across all seven classifications of metabolic pathways, particularly with pathways involved in amino acid biosynthesis.

**Figure 3.**
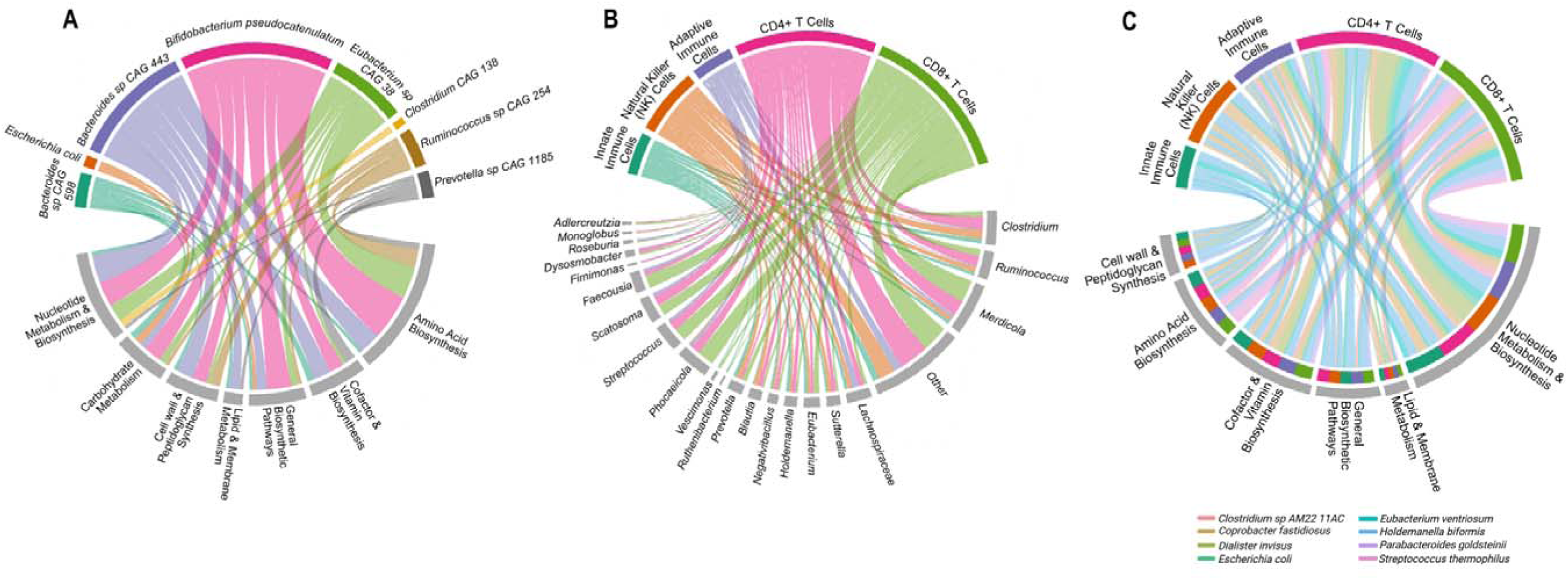
Associations between colonic pH, immune cells, the gut microbiota and species-specific metabolic pathways. D fferentially abundant associations between **(A)** colonic pH in the fourth quartile (Q4) and species-specific metabolic pathways (grouped by function), **(B)** immune cell populations/sub-types (grouped into categories) and species (grouped by genus) and **(C)** immune cell populations/sub-types (grouped into categories) and species-specific metabolic pathways (grouped by function). Differential abundance analyses were conducted using the R package MaAsLin2 (v1.15.1) at both the species and pathway levels, employing linear regressions adjusted for age, sex, and BMI. Relative abundances were normalised using the centred log-ratio (CLR) method. Adjustments were performed using the Benjamini–Hochberg False FDR method. Only significant associations (q<0.05) with two or more associations are depicted in the chord diagrams. Line thickness is scaled to the significance of the association (−log10(FDR)). Figures were generated with the circlize R package (v0.4.16). n=48-53.

### Gut microbial species and metabolic pathways are associated with the peripheral immune system

Next, we sought to identify specific taxa and microbial pathways associated with the peripheral immune system. Sixty two taxa, grouped at the genus level, were associated with 64 immune cell populations/sub-types, stratified by immune subset categories, resulting in 380 associations in total (Figure 3B, Tables S5-S7). Although associations were present across all immune groups, associations stemming from the CD4^+^ and CD8^+^ T cell categories prevailed. These clusters were predominantly composed of single and double pro-inflammatory cytokine-producing early and late-activated populations (CD69^+^ and human leukocyte antigen-DR isotype [HLA-DR]^+^, respectively), as well as PD-1^+^ expressing cells amongst others (Tables S5 and S7). We also identified that differentially abundant taxa classified as ‘Other’ exhibited the strongest associations with both CD4^+^ and CD8^+^ T cell clusters, followed by the genus *Merdicola*, comprising two unique, genetically similar bacterial taxa *Merdicola sp001915925* and *Merdicola sp900555085* (Table S5). Taxa grouped into the ‘Other’ category also predominated in associations with the peripheral immune system overall, comprising taxa yet to be characterised (Tables S5-S6). Of the 80 associations in total, GGB3612 SGB4882 (COE1 sp001916965) is the most prevalent, with 16 associations spanning innate, NK, CD4□, and CD8□T cell categories (Table S5). At the functional level, 945 species-specific metabolic pathway associations were identified with the peripheral immune system, linked by eight bacterial species (Figure 3C and Table S8). Thirty-five metabolic pathways, categorised by biological function, were associated with 60 peripheral immune cell populations/subtypes (Tables S4, S7 and S8). CD4^+^ and CD8^+^ T cell clusters showed consistently the largest and strongest associations, again predominated by subsets including single (i.e., IFNγ, IL-17A and TNFα) and double (i.e., IFNγ/TNFα IL-2/TNFα) pro-inflammatory cytokine-producing activated cell types (Figure 3C & Tables S7-S8). Moreover, pathways involved in nucleotide metabolism and biosynthesis showed the most pronounced associations within the peripheral immune system (Figure 3C, Tables S4, S7, and S8). Among the species identified as differentially abundant in the taxonomic profiling analysis, four remained consistent, linking the peripheral immune system and bacterial metabolic output: *Clostridium sp AM22 11AC*, *Eubacterium ventriosum*, *Holdemanella biformis* and *Streptococcus thermophilus*. We found that *Holdemanella biformis* exhibited the highest functional metabolic contribution across all pathway groups (excluding lipid and membrane metabolism) and had the greatest number of associations with the peripheral immune system (Figure 3C).

### *Holdemanella biformis* and its metabolic pathways are associated with the peripheral immune system

To delineate this further, we explored the relationship between *Holdemanella biformis,* formally *Eubacterium biforme*^38^, which was prevalent in 18.5% of the cohort, and the peripheral immune system more closely, revealing positive associations with 11 peripheral immune cell types (Figure 4A). More than 50% of associated immune cells were T cell subsets, specifically IL-17A-producing and PD-1^+^ CD4^+^ and CD8^+^ T cells. We identified two natural killer (NK) cell populations that produce TNFα, a well-characterised pro-inflammatory cytokine that reportedly governs whether Holdemanella biformis drives inflammation or resolution^39^. We demonstrated that the functional metabolic activity of *Holdemanella biformis* spans 37 pathways and exhibited varying positive associations with 21 peripheral immune cell populations and sub-types, 11 of which correspond to the immune subsets identified via taxonomic profiling (Figure 4B). Among these, four pathways consistently prevailed with the strongest associations across the suite of immune cell subsets; the methylerythritol phosphate pathway I, superpathway of L-tyrosine biosynthesis and superpathway of thiamine diphosphate biosynthesis III, as well as the flavin biosynthesis I pathway being associated with 18 and 19 subsets, respectively (Figure 4B).

**Figure 4.**
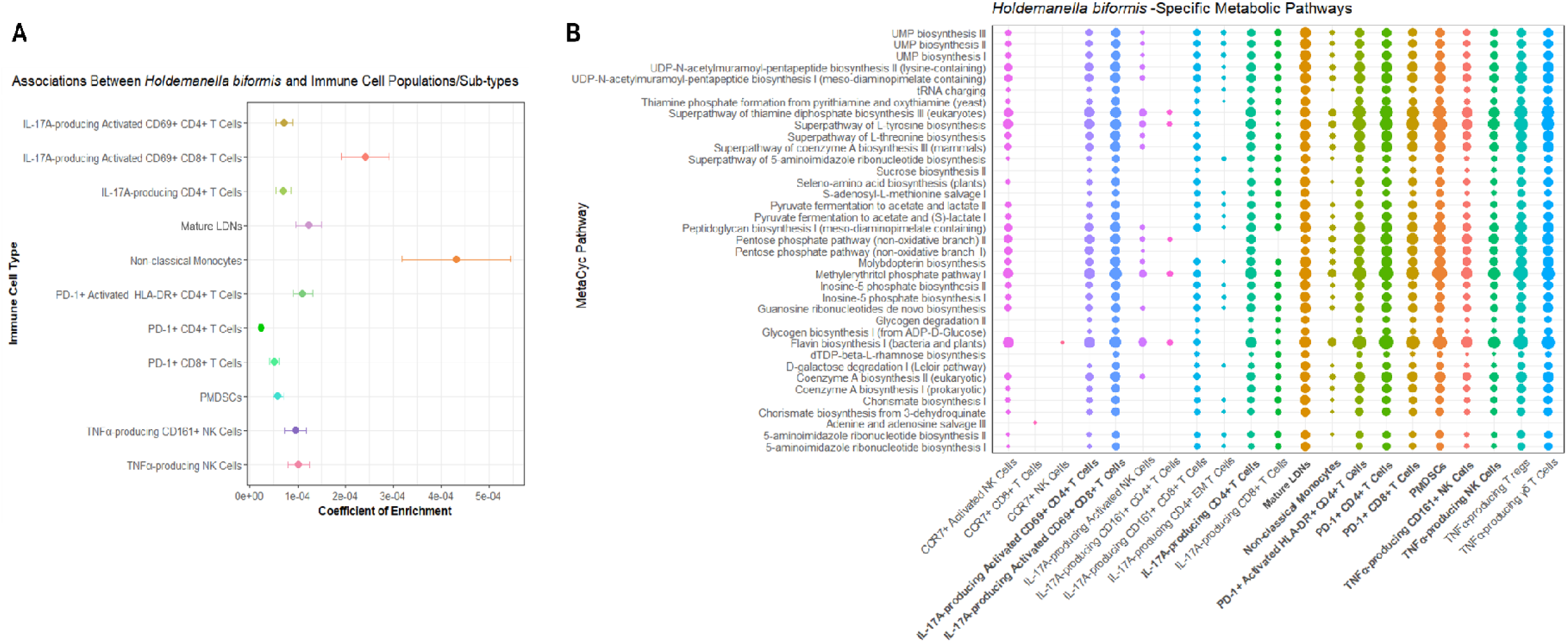
Associations between *Holdemanella biformis* and its pathways with the systemic immune system. Differentially abundant associations between the bacterial species Holdemanella biformis and **(A)** immune cell populations/sub-types, and **(B)** metabolic pathways. Differential abundance analyses were conducted using the R package MaAsLin2 (v1.15.1) at both the species and pathway levels, employing linear regressions adjusted for age, sex, and BMI. Relative abundances were normalised using the centred log-ratio (CLR) method. Adjustments were performed using the Benjamini–Hochberg False FDR method. Only significant associations (q<0.05) are depicted. In panel A, coefficient of enrichment (β) quantifies the direction and strength of the association, with standard errors (SE) shown as horizontal error bars. In panel B, the size of the points corresponds to the significance of the association (−log10(FDR)), and the point colour intensity reflects the scaled effect size (coefficient, β), with stronger colours indicating larger effect sizes. Immune cell types in bold overlap in both analyses. Figures were generated with the ggplot2 R package (v3.5.2). n=48-53. IL-, interleukin-; LDNs, low-density neutrophils; PD-1, programmed cell death protein 1; HLA-DR, human leukocyte antigen-DR isotype; PMDSCs, polymorphonuclear myeloid-derived suppressor cells; TNF-α, tumour necrosis factor-α; NK, natural killer.

### *Eubacterium*-specific metabolic pathways emerged as the shared feature of colonic pH in quartile four and the peripheral immune system

To investigate the interplay between colonic pH, gut microbiota, and the peripheral immune system, we compared the taxonomic and functional profiling results to identify any shared features. This revealed a genus-level convergence centred on *Eubacterium*, with distinct species showing differential associations across biological domains (Figure 5). *Eubacterium ventriosum*, which had a prevalence of 25.9% in the cohort, was consistently linked to peripheral immune cell profiles, suggesting a potential relationship with systemic immune modulation. In contrast, *Eubacterium rectale* and *Eubacterium sp CAG 38* were associated with colonic pH in quartile four. Taxonomic differential abundance analysis showed that three immune cell types, C-C chemokine receptor type 7 (CCR7)^+^, central memory (CM) CD8^+^ T cells, and non-classical monocytes, were positively associated with *Eubacterium ventriosum* (Figure 5A). These remained prominent among the 13 other peripheral immune cell types positively associated with the metabolic pathways of *Eubacterium ventriosum* (Figure 5B). Non-classical monocytes exhibited the strongest and largest associations with 15 of the 16 metabolic pathways, whilst the purine ribonucleoside degradation pathway had the highest number of associations across the suite of immune cell subsets, predominated by pro-inflammatory T cell subsets including IL-17A-producing and PD-1 expressing CD4^+^ and CD8^+^ T cells, as well as TNFα-producing NK and T cell populations (Figure 5B).

**Figure 5.**
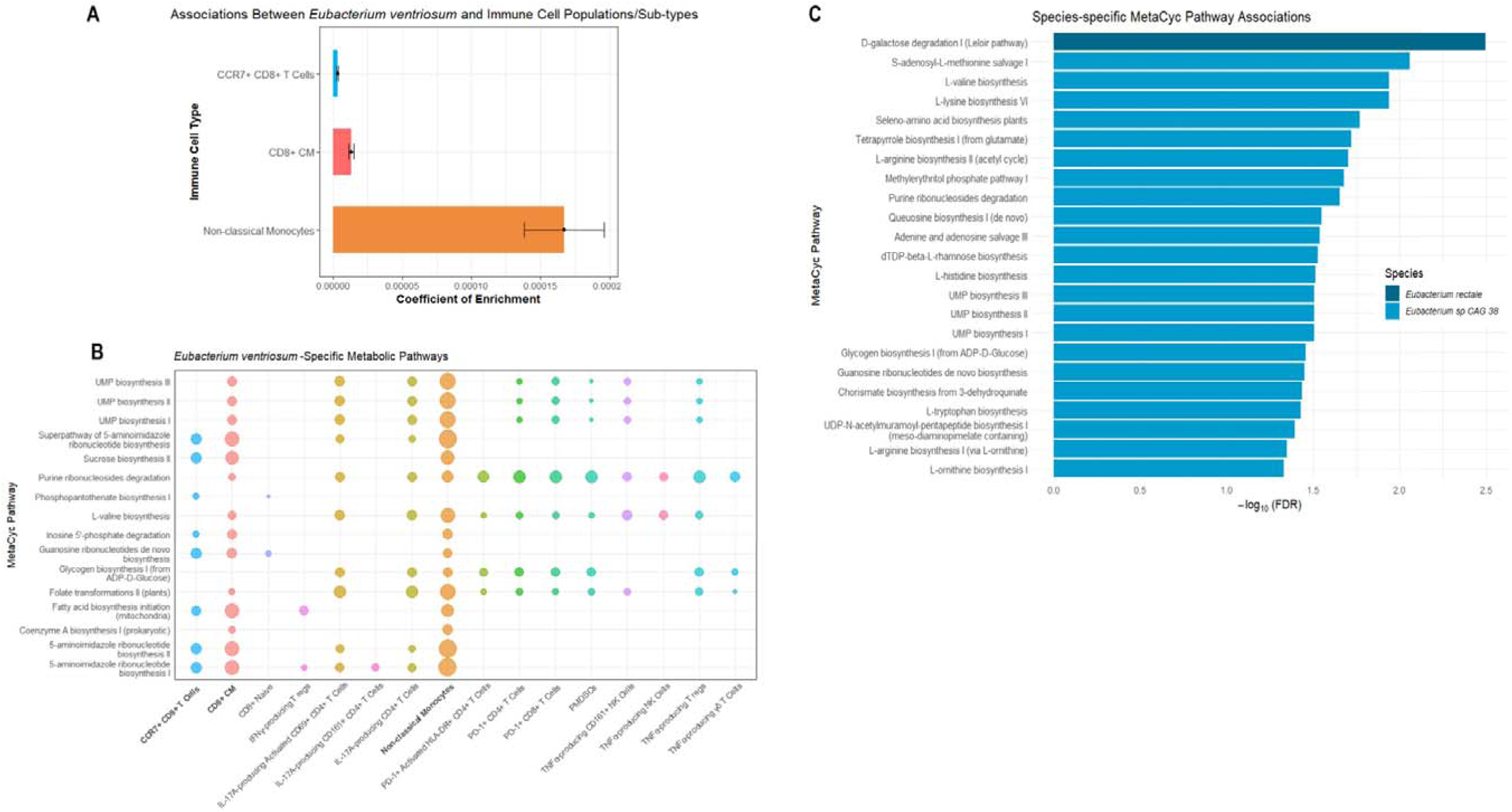
*Eubacterium* species are a key link between the gut microbiota and the systemic immune system. Differentially abundant associations between the bacterial species Eubacterium ventriosum and **(A)** immune cell populations/sub-types, and **(B)** metabolic pathways. Differentially abundant associations between **(C)** quartile 4 (Q4) colonic pH and Eubacterium rectale-and Eubacterium sp CAG 38-specific metabolic pathways. Differential abundance analyses were conducted using the R package MaAsLin2 (v1.15.1) at both the species and pathway levels, employing linear regressions adjusted for age, sex, and BMI. Relative abundances were normalised using the centred log-ratio (CLR) method. Adjustments were performed using the Benjamini–Hochberg False FDR method. Only significant associations (q<0.05) are depicted. In panel A, coefficient of enrichment (β) quantifies the direction and strength of the association, with standard errors (SE) shown as horizontal error bars. In panel B, the size of the points corresponds to the significance of the association (−log10 (FDR)), and the point colour intensity reflects the scaled effect size (coefficient, β), with stronger colours indicating larger effect sizes. Immune cell types in bold overlap in both analyses. In panel C, association strength is represented by the weighted –log10(FDR). Figures were generated with the ggplot2 R package (v3.5.2). n=48-53. CCR7, C-C chemokine receptor type 7; CM, central memory; IFN-γ, interferon-γ; T regs, T regulatory cells; IL-, interleukin-; PD-1, programmed cell death protein 1; HLA-DR, human leukocyte antigen-DR isotype; PMDSCs, polymorphonuclear myeloid-derived suppressor cells; TNF-α, tumour necrosis factor-α; NK, natural killer.

Whilst we did not identify any *Eubacterium ventriosum*-specific metabolic pathway associated with colonic pH, we profiled another two metabolically active *Eubacterium* species (Figure 5C). *Eubacterium rectale* facilitated the positive association between colonic pH in quartile four and D-galactose degradation (the Leloir pathway). In contrast, *Eubacterium sp CAG 38*-specific pathways, including those involved in amino acid and nucleotide biosynthesis, accounted for the remaining positive associations with quartile four (Figure 5C).

To better elucidate the biological relevance of these findings, we examined overlapping pathways across both functional analyses and found that *Eubacterium sp CAG 38* and *Eubacterium ventriosum* contributed to the L-valine (amino acid) biosynthesis pathway and the purine ribonucleoside degradation and UMP biosynthesis II and III pathways, classed as nucleotide metabolism and biosynthesis pathways respectively, linking colonic pH in the fourth quartile and the peripheral immune system (Figure 6). Twelve cell types prevailed, reinforcing a more pro-inflammatory profile characterised by IL-17A-producing CD4^+^ T cells, non-classical monocytes and TNFα-producing NK and T cell subsets, all of which have been consistently reported throughout. Additionally, PD-1-expressing T cells, particularly CD8^+^ T cells, repeatedly emerged as a recurrent feature.

**Figure 6.**
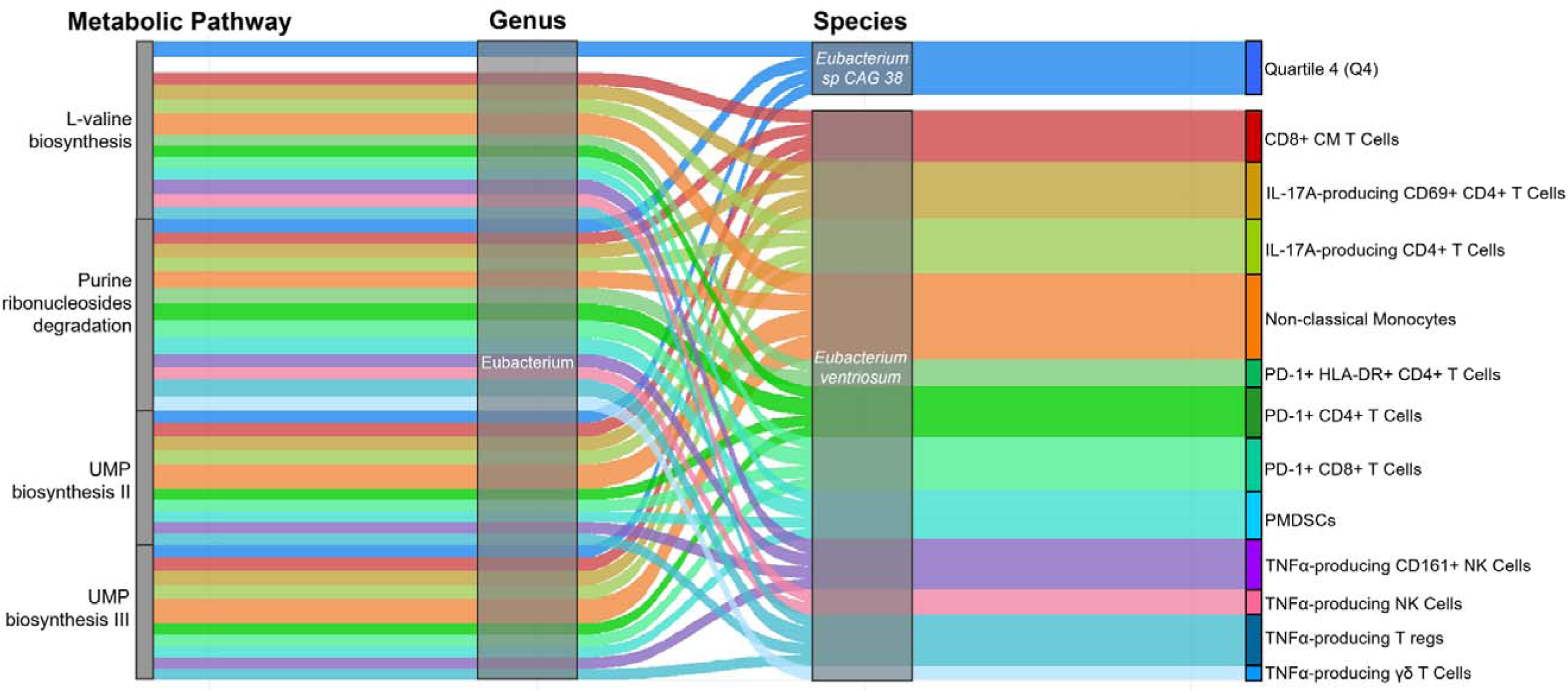
*Eubacterium* species connect microbiota, colonic pH and the systemic immune system. Alluvial diagram showing the connections between significant MetaCyc pathways, bacterial genus and associated species and biological variables, including quartile 4 (Q4) colonic pH and specific immune cell populations/sub-types. Link widths are weighted by the strength of association (−log10(FDR)). Out of all significant associations detected, these immune cell types, Q4, and metabolic pathways are linked through the Eubacterium genus. Differential abundance analyses were conducted using the R package MaAsLin2 (v1.15.1) at both the species and pathway levels, employing linear regressions adjusted for age, sex, and BMI. Relative abundances were normalised using the centred log-ratio (CLR) method. Adjustments were performed using the Benjamini–Hochberg False FDR method. Only significant associations (q<0.05) are depicted. Figure was generated using the ggplot2 (v3.5.2) and ggalluvial (v0.12.5) R packages. n=48-53. CM, central memory; IL-, interleukin-; PD-1, programmed cell death protein 1; HLA-DR, human leukocyte antigen-DR isotype; TNF-α, tumour necrosis factor-α; NK, natural killer; T regs, T regulatory cells.

## Discussion

While the gut microbiota is known to influence mucosal immunity, its impact on systemic immune tone via physicochemical cues such as colonic pH remains poorly defined in humans. A pilot study using 16S ribosomal RNA sequencing showed that baseline frequencies of 10 circulating T cell subsets were associated with shifts in bacterial genera over 6 weeks, suggesting links between peripheral immunity and microbial dynamics^40^. Building on this, we performed in-depth phenotyping of gastrointestinal pH using real-time monitoring with the SmartPill device, the gut microbiota using metagenomic sequencing, and the immune system using flow cytometry to characterise 121 subtypes and activation markers. We demonstrated that both colonic luminal and regional pH were associated with differences in peripheral immune profiles. Lower colonic luminal pH was associated with the enrichment of T cell populations, particularly CD8^+^ T cells, including cytokine-producing and pre-exhausted phenotypes. A more acidic colonic pH, primarily driven by SCFA production from fibre fermentation^17^, may foster a metabolically active gut environment^1^ that supports functional CD8^+^ T cell responses. These cells, traditionally recognised for their role in antiviral and cytotoxic responses^41,42^, are highly responsive to environmental cues, including microbial-derived metabolites and inflammatory cytokines^43–45^. These environmental cues can potentiate CD8^+^ T cell effector responses and enhance the secretion of key pro-inflammatory mediators including IFNγ and TNFα^46–48^. In contrast, a more alkaline pH, typically associated with depletion of fibre substrates for fermentation and reduced SCFA levels^17^, may favour PD-1^+^ exhausted-like CD8^+^ phenotypes, linked to chronic diseases such as cancer^29,49^ and high blood pressure^50^. Additionally, higher mature LDN counts were associated with a more alkaline colonic pH^51^, commonly observed in metabolic syndrome^52^. Together, these findings suggest that colonic pH, shaped by the gut microbiota and microbial fermentation, may serve as a functional cue for peripheral immune activation, linking gut metabolic tone to systemic immune outcomes.

A key finding was *Eubacterium* species represented a shared microbial feature linked to variation in both colonic pH and peripheral immune profiles in humans. This genus, a core component of the human gut microbiota, exhibits widespread colonisation across diverse populations spanning Africa^53^, Australia^54^, Europe^55^, Asia^56^ and the Americas^57,58^. Our analyses revealed that, while functionally distinct, *Eubacterium sp CAG 38* and *Eubacterium ventriosum* metabolically converge in the L-valine biosynthesis pathway, which emerged as the only shared metabolic feature associated with colonic pH (i.e., quartile 4) and 11 peripheral immune cells, 7 of which are T cell subtypes. Valine, an essential branched-chain amino acid, serves as a fundamental protein building block and an immunomodulatory metabolite^59^. L-valine enhances PI3K/Akt1 signalling in macrophages^60^, key antigen-presenting cells of the innate immune system^61^. Pharmacological inhibition of PI3K partially attenuates L-valine-induced bacterial phagocytosis^60^, aligning with prior evidence that PI3K/Akt signalling governs macrophage phagocytic function^62,63^. We observed a strong association with non-classical monocytes, precursors of macrophages, which also exhibit antigen-processing capabilities under inflammatory conditions^64^. Moreover, altered valine tRNA biogenesis augments mitochondrial energy production in T cell acute lymphoblastic leukemia^65^, underscoring valine’s broader role in supporting immune cell bioenergetics across both innate and adaptive compartments. These findings suggest that *Eubacterium*-derived valine metabolism may represent a mechanistic bridge between gut physicochemical gradients and systemic immune tone.

In parallel, both species showed strong associations with the purine ribonucleoside degradation pathway. Intestinal uric acid, the end product of purine metabolism^66^, is predominantly host-derived; however, systemic levels can increase in response to diets high in purine-rich foods (i.e., red meat, seafood) or low in fibre, shifting microbial activity toward increased uric acid production^67^. As a recognised damage-associated molecular pattern^68,69^, uric acid activates antigen-presenting cells – including monocytes/macrophages and dendritic cells – prompting the upregulation of co-stimulatory molecules such as CD80 and CD86^70^. This enhanced expression facilitates the activation of CD8^+^ T cells, driving their proliferation and effector function^71,72^. Conversely, pharmacological inhibition of uric acid production suppresses CD8^+^ T cell activation and effector functions, as evidenced by reduced CD69 and IFN-γ expression, respectively, and proliferative capacity due to diminished secretion of IL-2^73^. These findings suggest that sustained upregulation of *Eubacterium ventriosum*-mediated purine catabolism may elevate uric acid levels, contributing to the altered CD8^+^ T cell landscape we observed, including loss of IL-2 production in PD-1^+^ CD8^+^ T cells, consistent with a pre-exhausted phenotype.

Under physiological conditions, PD-1 is transiently induced upon T-cell activation to limit excessive T-cell receptor signalling and maintain immune homeostasis^74^. Under conditions of chronic infection and cancer, however, persistent antigen exposure drives sustained PD-1 expression on CD8^+^ T cells, contributing to their progressive functional exhaustion^28,29,75^. In cancer, exhausted T cells accumulate within the tumour microenvironment and are a key target of immune checkpoint blockade^49^. For example, therapeutic inhibition of PD-1 and its ligand PD-L1, also known as the PD-1/PD-L1 axis^76^, has shown clinical efficacy across numerous cancer types and acts primarily by reinvigorating effector function and restoring anti-tumour immunity^77,78^. Butyrate-producing bacteria, such as those in the *Eubacterium* genus^85,86^, including *Eubacterium R sp003526845*, which we identified as differentially abundant, can modulate the epigenetic and metabolic programming of CD8^+^ T cells, thereby influencing the expression of exhaustion markers such as PD-1^79^. Through enhanced histone acetylation and improved mitochondrial function, butyrate supports effector activity and mitigates terminal exhaustion, ultimately enhancing responsiveness to PD-1 checkpoint blockade^80–82^. This may underlie the positive correlation we observed between PD-1^+^ CD8^+^ T cells and plasma butanoic acid levels and may explain why these profiled PD-1^+^ CD8^+^ T cells remain functional (i.e., IFNγ-producing) and are not terminally exhausted. Butyrate also plays a potent role in regulating neutrophil activity by suppressing inflammation through pathways such as GPR43 signalling^83,84^. Consequently, reduced SCFA levels, often indicated by a more alkaline colonic pH, may contribute to the elevated mature LDN counts observed.

Another key finding concerned *Holdemanella*, which has been associated with improved outcomes in inflammatory bowel disease and immune checkpoint inhibitor-associated colitis^85^. *Holdemanella biformis* is a well-characterised producer of SCFAs, particularly butyrate, and has been reported to exhibit both anti-inflammatory and anti-tumorigenic properties in the gut^86,87^. However, our analysis revealed a strong functional association with the L-tyrosine biosynthesis pathway, a metabolic pathway recently implicated in microbial adaptation to dietary fibre deficiency^88,89^. Importantly, increased L-tyrosine biosynthesis enhances microbial tyrosine fermentation, leading to elevated *p*-Cresol production^90^. This microbial metabolite is subsequently conjugated in the host to form uremic toxins such as *p*-Cresol sulphate and *p*-Cresol glucuronide, both of which have been implicated in systemic toxicity, chronic kidney disease^91,92^, and elevated blood pressure in humans^88^.

While these findings offer important insights, certain limitations cannot be overlooked, most notably, the small cohort size. The colonic quartiles were calculated based on transit time, not on geographical locations; thus, they represent proxy sites rather than the exact colonic regions we attempted to characterise. This underscores the need for novel technologies capable of capturing more precise, spatially resolved pH profiles across both luminal and tissue microenvironments. Following on from this, while faecal samples remain the most readily obtainable specimen for evaluating the human gut microbiota, they do not accurately reflect region-specific microbial communities; thus, such devices would also ideally enable concurrent sampling of region-specific microbiota, facilitating a more integrated understanding of host–microbiota interactions within distinct gut niches. Therefore, our findings regarding quartile four regional colonic pH were not unexpected, as quartile four is most directly reflective of the rectum and, therefore, of the faecal microbiota from which metagenomic sequencing was performed. Nonetheless, our study has several strengths, including a balanced representation of males and females and the use of an *in vivo* system to capture detailed pH measurements in the human GI tract. The device employed in this study is no longer commercially available, and no current alternatives offer comparable resolution or functionality. This renders our dataset both unique and timely, providing a valuable reference point for future investigations into gut physicochemical-immune axes in humans.

In conclusion, our findings indicate a potential physiological axis linking gut microbial composition, colonic intraluminal pH, and the production of fibre-derived metabolites to systemic immune modulation in humans. This coordinated interplay supports the idea that local gut environmental cues, particularly those shaped by microbial fermentation and physicochemical gradients, can exert far-reaching effects on peripheral immune tone. These insights advance our understanding of how the gut microbiota, associated microbial metabolism, and gut pH converge to regulate human immunity, with implications for the design of microbiota-targeted interventions for immune-mediated and inflammatory diseases.

## Materials and Methods

All materials and methods are detailed in the Supplementary file.

## Acknowledgements

The authors thank Monash University FlowCore, the Monash Statistical Consulting Service, the Monash eResearch capabilities for providing access to M3 servers, and the Australian Genome Research Facility. The authors also thank Dr Matthew Snelson and Michael Nakai for their guided assistance with data analysis tools and experimental work respectively, as well as Dr Meena Madhur and Dr Charles Duncan Smart for their intellectual input and methodological guidance.

## Author Contributions

E.D. designed, planned and performed the human immunoprofiling flow cytometry experiment, data analyses, and wrote the manuscript. D.R.J. secured human ethics approval, recruited the participants, and analysed the SmartPill^TM^ data, verified by C.K.Y and D.S. L.C.T. and C.Y. contributed to the development of analysis tools and data analysis. D.A. and D.C. measured SCFAs. P.R.G. contributed to the interpretation of intestinal pH. C.R.M. contributed towards the central hypothesis and securing funding. J.O.D. provided intellectual input and assisted in the planning and execution of the human immunoprofiling flow cytometry experiment. J.M. contributed towards trial design, supervision, and data collection and analysis. F.Z.M. conceived and designed the study, secured funding, provided intellectual input and contributed to writing the article. All authors approved and contributed to the final version of the manuscript.

## Data and Code Availability

The codes supporting this study’s findings are available at https://github.com/fzmarques/Dinakis-et-al_pH-microbiome-and-immune-system. Microbiome data are available at https://doi.org/10.5281/zenodo.17645792. We did not obtain patient consent for all the data to be made publicly available. However, the data underlying this article can be shared for selected research questions upon reasonable request to the corresponding author. Please email F.Z.M. at, who will respond within 4□weeks.

