## Supplementary material for "The gut microbiota and colonic pH map to the peripheral immune landscape in humans": Online supplementary methods, figures and tables

**Short title:** Gut microbiota, pH and immune system

Evany Dinakis^1,2^, Dakota Rhys-Jones^3^, Leticia Camargo Tavares^1,2^, Chaoran Yang^1,2^, Liang Xie^1,4^, CK Yao^3^, Daniel So^3^, Dovile Anderson^5^, Darren Creek^6^, Stephen J. Turner^7^, Peter R. Gibson^3^, Charles R. Mackay^8,9^, Joanne A. O’Donnell^1,2^, Jane Muir^3^, Francine Z. Marques^1,2,10*^

^1^Department of Pharmacology, Biomedical Discovery Institute, Faculty of Medicine, Nursing and Health Sciences, Monash University, Clayton VIC 3800, Australia; ^2^Victorian Heart Hospital, Monash University, Clayton VIC, 3168, Australia; ^3^Department of Gastroenterology, Central Clinical School, Monash University, Melbourne, 3000, Victoria, Australia; ^4^Precision Medicine Translational Research Programme, Department of Obstetrics & Gynaecology, Yong Loo Lin School of Medicine, National University of Singapore, Singapore; ^5^Monash Proteomics and Metabolomics Facility, Monash Institute of Pharmaceutical Sciences, Monash University, Parkville, VIC 3052, Australia; ^6^Monash Biomedical Imaging, Monash University, Clayton, VIC 3168, Australia; ^7^Department of Microbiology, Immunity Theme, Biomedical Discovery Institute, Monash University, Clayton, VIC, Australia; ^8^Department of Microbiology, Biomedicine Discovery Institute, Monash University, Clayton, VIC 3800, Australia; ^9^School of Pharmaceutical Sciences, Shandong Analysis and Test Center, Qilu University of Technology (Shandong Academy of Sciences), Jinan, 250014, China; ^10^Baker Heart and Diabetes Institute, Melbourne, VIC 3000, Australia.

***Corresponding author**: Prof Francine Marques, Hypertension Research Laboratory, Victorian Heart Institute, Level 2, Victorian Heart Hospital, 631 Blackburn Road, Clayton, VIC 3168, Monash University, Melbourne, Australia, Phone: ^+^61-03-9905 6958.

### **Materials and Methods**

**The pH of Intestines and Blood-pressure Regulation (pHibre) cohort**

The pH of Intestines and Blood-pressure Regulation (pHibre) study was approved by the Monash University Human Research Ethics Committee (Study ID: 23336) and followed the Declaration of Helsinki. All participants provided written consent. The study was registered with the Australian New Zealand Clinical Trials Registry under registration number ACTRN12620000284965. Recruitment of male and female volunteers was conducted between June 2020 and October 2021 through study advertisement. Inclusion criteria included body mass index (BMI) between 18.5–35 kg/m^2^ and age between 18-70 years old. Exclusion criteria included non-English speaking; gastrointestinal disorders and diseases (including history of intestinal surgery, lactose intolerance, inflammatory bowel disease, celiac disease, chronic pancreatitis or other malabsorption disorder); gastrointestinal surgery in the last 3 months; type 1 or 2 diabetes; chronic kidney disease; swallowing disorders or severe dysphagia to food or pills; having implanted or portable electro-mechanical devices; and recent use of antibiotics (< 3 months) or probiotics (< 6 weeks). The recruitment summary is outlined in Figure 1A. Study outcomes are described in detail in the following sections. Briefly, the study included one visit during which a fasted blood sample was collected for subsequent plasma SCFA quantification and PBMC isolation, and measurement of fasting blood glucose (Table S1), followed by ingestion of the wireless pH-motility capsule (SmartPill) to measure transit time and pH^1^. The mean whole-gut transit time was 32.8 ± 17.6 hours. Participants also provided a faecal sample and self-assessed stool consistency using the Bristol Stool Form Scale (BSS)^2^, with a median reported BSS stool classification of type 4 (Table S1).

**Habitual dietary records**

Participants completed a three-day food diary leading up to the appointment, on one weekend day, and on two weekdays to assess habitual intake. The dietary information was then analysed through a nutritional analysis software, FoodWorks 10 (Xyris, Queensland), using an Australian food composition database for energy, macro- and micronutrient intake (Table S1).

**Wireless pH-motility (SmartPill) capsule data collection**

Fasting participants consumed a standardised low-fat and low-fibre muesli bar to facilitate gastric motility and support capsule passage, followed by ingestion of the SmartPill™ Motility Testing System (Medtronic). After being swallowed, the SmartPill™ measured in real-time, the intraluminal pH, pressure, and temperature as it travelled through the gastrointestinal tract. The information was transmitted wirelessly to a portable receiver worn by the participants until excretion, indicated by a signal loss from the device coinciding with the passage of stools. Because the SmartPill was ingested once, each participant contributes one complete transit profile, from which GI transit times and regional pH characteristics were independently determined by at least two trained investigators.

**Peripheral blood mononuclear cell (PBMC) isolation & flow cytometric preparation**

PBMCs were isolated by density gradient centrifugation. Briefly, whole blood was diluted at a 1:1 volume ratio with Dulbecco’s phosphate buffered saline (dPBS, 1X, Sigma-Aldrich, D8537) supplemented with 2% foetal bovine serum (FBS, Sigma-Aldrich, 12007C). The diluted blood was slowly overlayed onto Ficoll-Paque (GE Healthcare, 17-1440-03) at a 1:2 volume ratio to create an interface. Density gradient centrifugation was run at 700g (1800RPM) for 20 minutes at 20°C with the deceleration (brake) off. Following centrifugation, the plasma layer was aspirated, and the PBMC layer was harvested using a sterile transfer pipette. PBMCs were washed with cold 1x PBS and centrifuged at 400g (1400RPM) for 10 minutes. The cell pellet was then resuspended in 10ml of cold 1x PBS. Cell counts were performed manually using a haemocytometer and Trypan blue (0.4%; Invitrogen, T10282). Samples were cryopreserved until the time of the experiment. Briefly, isolated PBMCs were resuspended in freezing medium (10% DMSO; Sigma-Aldrich, D8418 and 90% FBS). Cryovials were then placed into a Mr. Frosty (ThermoFisher, 5100-0001) and into a -80°C freezer. Cryovials were transferred to liquid nitrogen storage the following day.

Frozen PBMC samples were retrieved, slowly thawed in a 37°C water bath and resuspended in pre-warmed RPMI 1640 (Biomedicine Learning and Teaching Building & Media Services Facility) supplemented with 10% FBS. Cells were centrifuged at 400g for 5 minutes. Cell counts were performed manually using a haemocytometer and Trypan blue (0.4%; Invitrogen, T10282). 2.5x10^6^ cells/mL per sample were stimulated with 50ng/mL phorbol 12-myristate 13-acetate (PMA; Sigma-Aldrich, 16561-29-8) and 1μg/mL ionomycin (Sigma-Aldrich, I0634) in a total of 3mL (7. 5x10^6^ cells total) and incubated at 5% CO2 at 37°C for 60 minutes. Cells were then treated with brefeldin A (5μg/mL, eBioscience™ Brefeldin A Solution (1000X), 00-4506-51) and monensin (5μg/mL, eBioscience™ Monensin Solution (1000X), 00-4505-51) and incubated for 5 hours and refrigerated overnight. Cells were washed and spun down at 400g for 5 minutes and 1x10^6^ viable cells per sample were subjected to flow cytometric staining. 1x10^6^ cells per sample were not subjected to the stimulation protocol to allow accurate staining and analysis of immune cell populations that are particularly sensitive to stimulation and at higher risk of cell death, such as mature low-density neutrophils (LDNs).

**Flow cytometry**

Cells were stained with ViaDye Red Fixable Viability Dye (1:500, Cytek Biosciences, R7-60008), incubated for 20 minutes at RT in the dark, washed with staining buffer, comprised of 1X dPBS supplemented with 2% bovine serum albumin (BSA, Sigma-Aldrich, A7906) and spun at 400g for 5 minutes at RT. Cells were blocked with True-Stain Monocyte Blocker (BioLegend, 426102) and diluted in Brilliant Stain Buffer Plus (BD Biosciences, 568264). The antibody panel designed incorporated a custom human immunoprofiling assay panel with 14 made-to-order cFluor reagents (Cytek Biosciences, RC-00685) and 15 externally sourced antibodies (Table S9). Surface antibodies TCRγδ and CCR7 were added to all stimulated samples, incubated for 10 minutes at RT in the dark, followed by the addition of all other surface antibody markers (Table S9). Unstimulated cells were incubated with a separate antibody cocktail containing CD14, CD15 and CD16 surface antibodies. Cells were incubated for 20 minutes at RT in the dark, washed with staining buffer and spun at 400g for 5 minutes at RT. Cells were fixed with the Foxp3/Transcription Factor Staining Buffer Kit (Sapphire Bioscience, TNB-0607-KIT), washed with 1X Perm Buffer, spun at 400g for 5 minutes at RT and at this time, unstimulated cells were ready for acquisition. Stimulated cells were stained with an antibody cocktail containing intracellular antibody markers as described in Table S9 for 60 minutes at RT. Samples were washed with 1X Perm Buffer, spun at 400g for 5 minutes at RT, and this time, stimulated cells were ready for acquisition. Samples were acquired on a Cytek Aurora 5 Laser Spectral Analyser at the FlowCore Facility of Monash University. Data was analysed using FlowJo v10.10.0, and gating strategies are outlined in Table S10.

**Plasma short-chain fatty acid (SCFA) measurements**

Fasting blood was collected in the morning, and plasma SCFAs were quantified using mass spectrometry as previously described^3^. Briefly, 20 µl of plasma was analysed in duplicates in a Q-Exactive Orbitrap mass spectrometer (Thermo Fisher Scientific) in conjunction with a Dionex UltiMate 3000 RS high-performance liquid chromatography (HPLC) system (Thermo Fisher Scientific). We accepted a coefficient of variability <15%. Standard curves were constructed using the area ratio of the target analyte, and the internal standard in the range of each analyte was used. The levels of acetic (n=1), butanoic (n=3), isobutanoic (n=2), valeric (n=1), isovaleric (1) and propionic acid (n=9) were below detection level of the standard curve; in this case, they were considered half of the lowest measurable value (equivalent to 0.025 ng ml−1).

**Faecal DNA extraction and high-throughput shotgun-metagenomic sequencing**

The STORMS (Strengthening the Organization and Reporting of Microbiome Studies) reporting guidelines were followed throughout^4,5^. Participants collected stool samples at home in tubes containing DNA/RNA Shield (Zymo Research, R1100) for microbial DNA extraction. Tubes were brought to the clinics immediately or were stored at −20°C for less than 24 hours and then brought to the clinic, where they were stored at −80^o^C until further processing. DNA was extracted using the DNeasy PowerSoil DNA isolation kit (Qiagen, 12888). Libraries were constructed using the Illumina DNA Prep (M) Tagmentation Kit (Illumina, 20018705) with IDT for Illumina DNA/RNA UD Index Sets A-D (Illumina, 20027213-16) according to the manufacturer’s instructions, with a volume adjustment to accommodate processing in a 384-plate format. Resulting libraries were assessed using a high-sensitivity dsDNA fluorometric assay (QuantIT, ThermoFisher, Q33120) and individual libraries were visualised with capillary gel electrophoresis using the QIAxcel DNA High Resolution Kit (Qiagen, 929002). Libraries were required to meet minimum criteria for average size, smallest and largest fragment gating, and concentration. Individual libraries were pooled in equimolar amounts to create a sequencing pool, which was assessed using a high-sensitivity dsDNA fluorometric assay (QuantIT, ThermoFisher, Q33120) and visualised by capillary gel electrophoresis with the QIAxcel DNA High Resolution Kit (Qiagen, 929002). Pools were required to meet minimum criteria for average size, smallest and largest fragment gating, and concentration. Sequencing pools were loaded and sequenced on the Illumina NovaSeq™ 6000 using v1.5 300 bp PE sequencing reagents, according to the manufacturer’s instructions, to a target depth of 3GB. Sequences were demultiplexed, sequencer adaptors were trimmed, and files were converted from bcl to fastq format. QC processing of raw stool shotgun-metagenomic forward and reverse reads in fastq format was performed using KneadData7 as a wrapper. Raw reads were initially quality trimmed to remove base calls with a Phred score lower than 33 and a minimum read length of 50 nucleotides (SLIDINGWINDOW:4:20 MINLEN:50) using Trimmomatic v0.38.8 Trimmed reads were then host-filtered if they mapped against the human (GRCh37 and GRCh38) or the Escherichia phage phiX174 (NCBI accession: NC_001422) genomes using Bowtie29 v.2.3.5 very-sensitive pre-set parameters (-D 20 -R 3 -N 0 -L 20 -i S,1,0.50). QC visualisation was performed with fastQC v.0.12.1 and multiQC10 v1.19. The sequencing depth was 3 million reads per sample.

**Microbial taxonomic and functional profiling**

Taxonomic profiling per sample was performed using MetaPhlAn11 v4.1 and the mpa_vJun23_CHOCOPhlAnSGB_202307 database, which spans 36,822 species-level genome bins (SGB) for bacteria, archaea, and eukaryote, relying on over 5 million unique clade-specific marker genes. The CHOCOPhlAn database leverages systematically organised and annotated microbial genomes and gene family clusters based on the NCBI and UniProt/UniRef reference databases. For taxonomic profiling, by default, MetaPhlAn searched for high-quality reads matching at least 20% of the unique clade-specific marker genes available for a species using nucleotide BLAST (blastn) with an e-value (the number of alignments expected by chance) threshold of 1x10^-6^, indicating a strong similarity between the query sequence and the subject sequence in the database. When multiple matches occurred between a read and markers from different clades, only the top hit was considered, in a way that a read could only be mapped to a single clade. The obtained reads per kilobase (RPK) were normalised by the total number of reads per sample in the metagenome to ensure fair comparisons of abundances across samples. Abundance per sample was calculated based on the coverage of the species-specific marker genes multiplied by the clade’s average genome size. Relative abundances were then calculated as local clade abundance divided by the sum of all the local abundances for that sample.

Functional read profiling was performed using HUMAnN7 v3.9, with default settings. For functional profiling, HUMAnN uses Bowtie29 (for sequence alignment to pangenomes), Prodigal12 (for ORF identification), and Diamond13 (for BLAST local sequence alignment) against the UniProt Reference Clusters database at 90% identity (UniRef90). The abundances of metaCyc14 pathways were obtained from total coverage depth (RPK) values per sample, which were then normalised to relative abundances to enable comparisons between samples with different sequencing depths. Reads that mapped to gene families but could not be assigned to any known metaCyc pathway were grouped as ‘unintegrated’, whereas reads that did not map to any gene family were categorised as ‘unmapped’. These were excluded before functional differential abundance analyses.

**Microbial and metabolic pathway differential abundance analyses**

Differential abundance analyses were conducted using the R package MaAsLin2 (v1.15.1) at both the species and pathway levels, employing linear regressions adjusted for age, sex, and BMI. Relative abundances were normalised using the centred log-ratio (CLR) method and used as input. Species were included if present in at least one individual in the sample (totalling 1678 species). Additionally, metaCyc pathways were analysed at the species level, as provided by the HuManN pipeline and were included if present in at least one individual (totalling 15,173 species-level metaCyc pathways). Variables demonstrating fewer than two significant associations were omitted.

**Colonic pH diversity metrics analysis**

Both α and β diversity metrics were calculated based on colonic minimum, median, and maximum pH values, as well as quartile pH data. Specifically, α diversity, which estimates species diversity within a sample, was calculated using the Shannon and Simpson indices using the microbiome R package. β diversity was calculated at the species‑level genome bin (SGB) resolution using both unweighted and weighted UniFrac metrics, using the rbiom R package and the Species Genome Bin (SGB) phylogenetic tree version mpa_vJun23_CHOCOPhlAnSGB_202307.nwk provided by MetaPhlAn v4.1.

**Statistical analyses**

All statistical analyses were conducted in R 4.3.2. Normality was assessed using the Shapiro-Wilk test. To minimise the influence of extreme values, outliers were removed using the interquartile range (IQR) method. For each variable, the first quartile (Q1) and third quartile (Q3) were calculated, and the IQR was defined as the difference between Q3 and Q1. Data points falling below Q1 – 1.5 × IQR or above Q3 ^+^ 1.5 × IQR were excluded from further analysis. Comparisons between quartiles were performed using a one-way analysis of variance (ANOVA) followed by pairwise t-tests. Data presented as mean ± standard error of mean (SEM). Spearman’s rank correlations were performed on non-normally distributed continuous variables. A significance threshold of P<0.05 was used. Adjustments were performed using the Benjamini–Hochberg False FDR method, and q<0.05 was considered statistically significant. For alpha diversity, linear regressions (on log-transformed α metrics) were used for continuous variables. For beta diversity, Permutational multivariate analyses of variance (PERMANOVA) using 9,999 permutations were conducted with the adonis2 function from the vegan15 R package.

**Supplementary Figures**

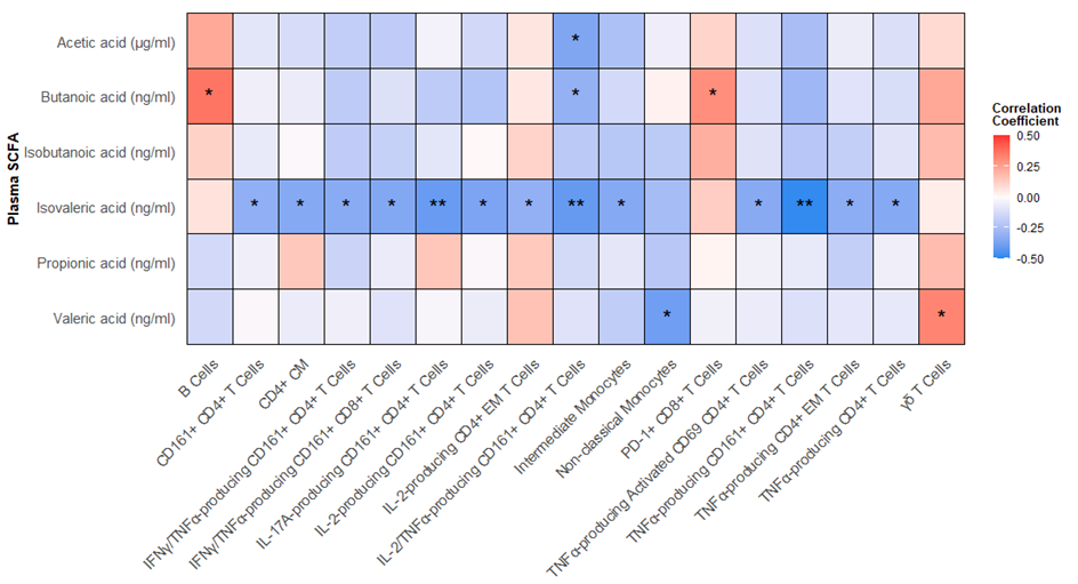

**Figure S1. Associations between plasma short-chain fatty acids (SCFAs) and peripheral immune cells.** Spearman’s rank correlations as a heatmap between plasma SCFA levels and peripheral immune cell populations/sub-types. Spearman’s rank correlations were performed on non-normally distributed continuous variables. The colour gradient represents the strength and direction of correlation (red = positive, blue = negative). Raw unadjusted P-values are presented in heatmaps; *P<0.05, **P<0.01. n=41-54. SCFA, short-chain fatty acid; CM, central memory; IFN-γ, interferon- γ; TNF-α, tumour necrosis factor- α; IL-, interleukin-; EM, effector memory; PD-1, programmed cell death protein 1.

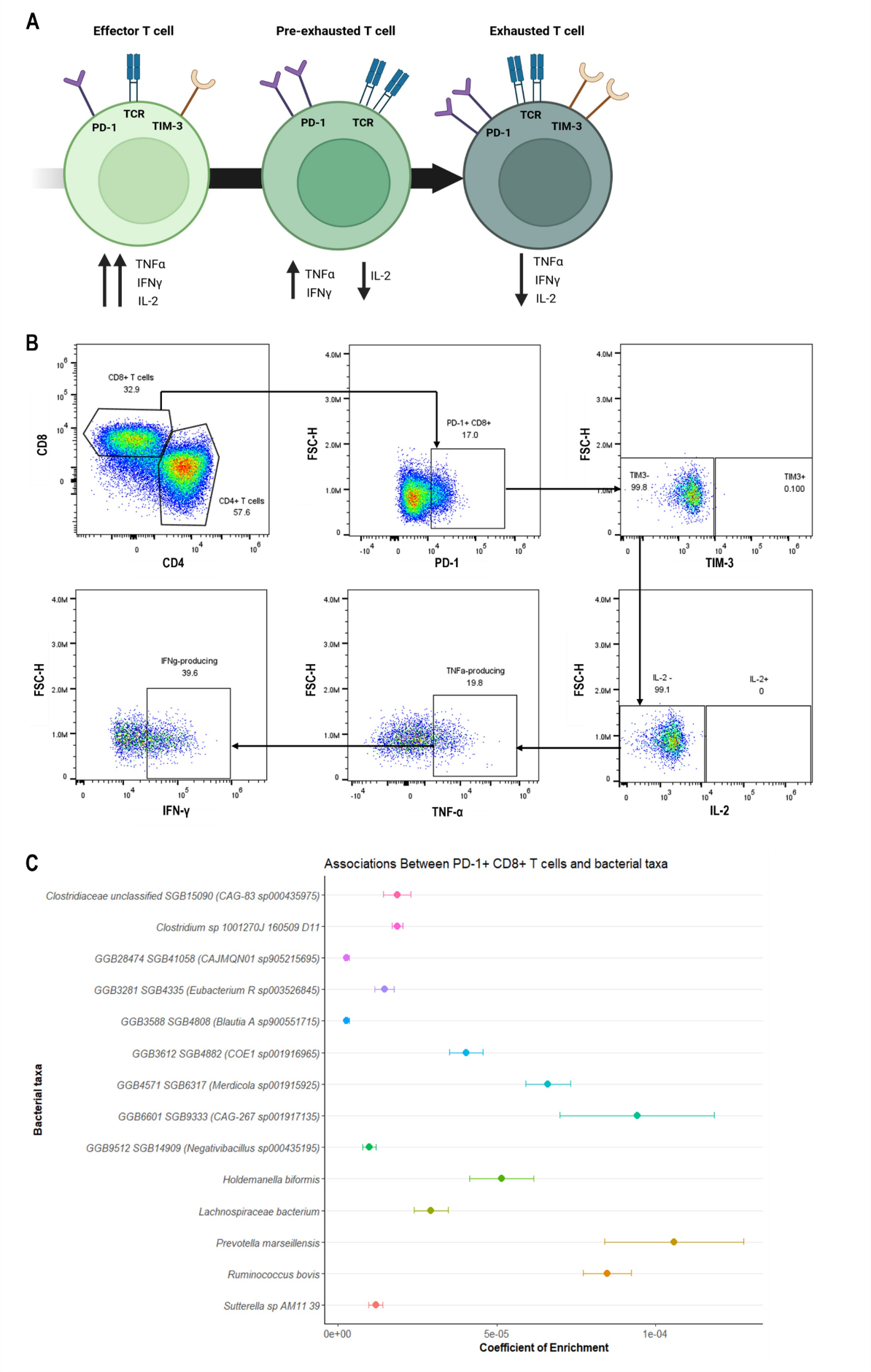
**Figure S2. CD8^+^ T cell transition from pre-exhausted to exhausted phenotype**. (A) Schematic of the transition of effector CD8^+^ T cells from pre-exhausted to exhausted. (B) Gating strategy used to identify the pre-exhausted subset of CD8^+^ T cells using FlowJo v 10.10.0. (C) Differential abundance analysis between PD-1^+^ CD8^+^ T cells and the gut microbiome using MaAsLin2.

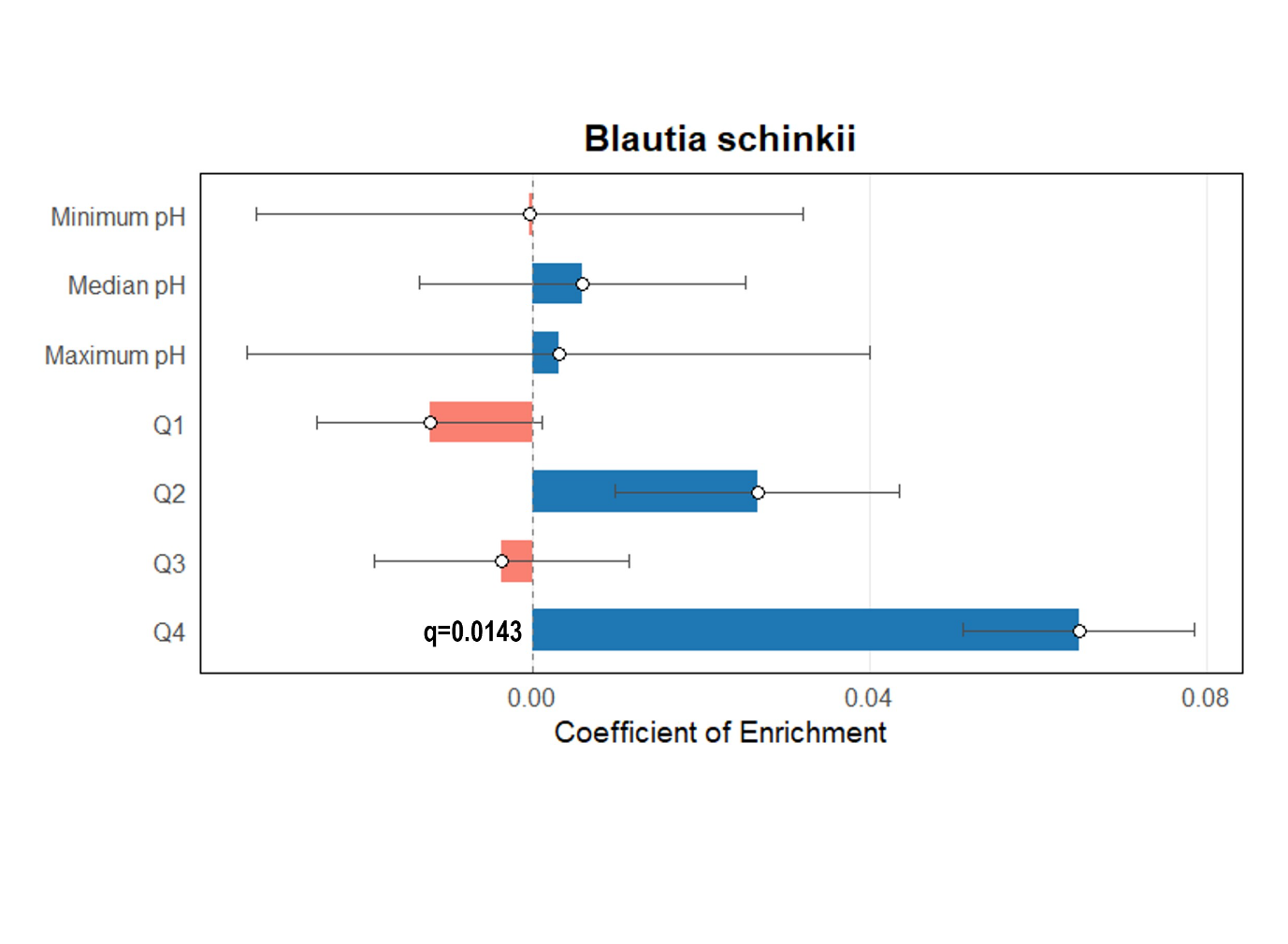

#### **Figure S3. Differential abundance analysis between colonic pH measures and the gut microbiome.** Analysis was performed using MaAsLin2 to identify microbial taxa associated with colonic pH. Only one bacterial species, *Blautia schinkii*, showed a statistically significant association with one of the colonic pH measures, quartile 4 (Q4) (FDR-adjusted q = 0.0143).

### **Supplementary Tables**

| **Cohort Characteristics** | **Participants (n=54)** | |
| --- | --- | --- |
|  | **Mean ± SD** | **N** |
| Sex (% female) | 22 (41%) | 54 |
| Age (years) | 49 ± 12.9 | 54 |
| BMI (kg m^−2^) | 27.0 ± 3.7 | 54 |
| Fasting glucose (mmol/L) | 4.8 ± 0.5 | 48 |
| Whole gut transit time (hours) | 32.8 ± 17.6 | 46 |
| Stool consistency, BSS (median) | 4 | 46 |
| **Habitual dietary intake** |  |  |
| Total energy (kJ) | 8713.75 ± 2379.4 | 53 |
| Carbohydrate (g) | 197.4 ± 67.62 | 53 |
| Protein (g) | 94.84 ± 29.29 | 53 |
| Total fat (g) | 88.36 ± 30.67 | 53 |
| Monounsaturated fat (g) | 33.38 ± 12.98 | 53 |
| Polyunsaturated fat (g) | 14.51 ± 6.82 | 53 |
| Saturated fat (g) | 31.98 ± 14.3 | 53 |
| Sugars (g) | 79.38 ± 32.37 | 53 |
| Total dietary fibre intake (g) | 25.86 ± 9.44 | 53 |
| **Plasma short-chain fatty acids (SCFAs)** |  |  |
| Acetic acid (μg/mL) | 3.91 ± 4.04 | 54 |
| Butanoic acid (ng/mL) | 57.72 ± 35.22 | 54 |
| Isobutanoic acid (ng/mL) | 24.36 ± 16.59 | 54 |
| Isovaleric acid (ng/mL) | 61.4 ± 39.89 | 54 |
| Propionic acid (ng/mL) | 414.53 ± 352.02 | 54 |
| Valeric acid (ng/mL) | 26.81 ± 11.99 | 54 |

**Table S1. Cohort Characteristics.**

**Table S2. α- and β-diversity associations on colonic pH variables.**

| **Colonic pH variables** | **α-diversity** | | | | **β-diversity** | | | |
| --- | --- | --- | --- | --- | --- | --- | --- | --- |
|  | **Shannon** | | **Simpson** | | **Unweighted** | | **Weighted** | |
|  | **Effect** | ***P* value** | **Effect** | ***P* value** | **R^2^** | ***P* value** | **R^2^** | ***P* value** |
| Colonic minimum pH | -0.075 | 0.40 | -0.040 | 0.29 | 0.027 | 0.070 | 0.017 | 0.43 |
| Colonic median pH | -0.044 | 0.40 | -0.022 | 0.32 | 0.027 | 0.076 | 0.018 | 0.39 |
| Colonic maximum pH | -0.086 | 0.36 | -0.025 | 0.53 | 0.027 | 0.068 | 0.028 | 0.17 |
| Quartile 1 | -0.020 | 0.58 | -0.0086 | 0.58 | **0.035** | **0.014** | 0.021 | 0.31 |
| Quartile 2 | -0.012 | 0.79 | -0.0066 | 0.74 | 0.016 | 0.76 | 0.010 | 0.77 |
| Quartile 3 | -0.00087 | 0.98 | -0.00026 | 0.99 | 0.017 | 0.66 | 0.010 | 0.78 |
| Quartile 4 | 0.019 | 0.65 | 0.0085 | 0.63 | 0.021 | 0.27 | 0.017 | 0.44 |

Associations with α-diversity were assessed using linear regression, reporting β coefficients (Effect) for log₁₀-transformed Shannon and Simpson indexes. For β-diversity, variance explained (R²) and *P* values were obtained from PERMANOVA (9,999 permutations). All models were adjusted for age, sex, and BMI. Significant associations (*P*<0.05) are shown in bold.

**Table S3. Differential abundance analysis for microbial metabolic pathway (metacyc) at the species level in relation to quartile 4 colonic pH.**

| **Functional pathway** | |  |  |  |  |
| --- | --- | --- | --- | --- | --- |
| **metaCyc pathway** | **Category** | **Species (HumanN pipeline)** | **β coefficient** | **SE** | **q-value (FDR)** |
| L-ornithine biosynthesis II | Amino Acid Biosynthesis | *Bacteroides sp CAG 598* | 2.01E-06 | 4.02E-07 | 0.00233 |
| Superpathway of coenzyme A biosynthesis III (mammals) | Cofactor & Vitamin Biosynthesis | *Bacteroides sp CAG 598* | 1.60E-06 | 3.18E-07 | 0.00233 |
| Coenzyme A biosynthesis I (prokaryotic) | Cofactor & Vitamin Biosynthesis | *Bacteroides sp CAG 598* | 1.78E-06 | 3.55E-07 | 0.00233 |
| L-histidine biosynthesis | Amino Acid Biosynthesis | *Bacteroides sp CAG 598* | 2.06E-06 | 4.11E-07 | 0.00233 |
| Methylerythritol phosphate pathway I | General Biosynthetic Pathways | *Bacteroides sp CAG 598* | 1.67E-06 | 3.32E-07 | 0.00233 |
| Phosphopantothenate biosynthesis I | Lipid & Membrane Metabolism | *Bacteroides sp CAG 598* | 1.86E-06 | 3.72E-07 | 0.00233 |
| Superpathway of coenzyme A biosynthesis I (bacteria) | Cofactor & Vitamin Biosynthesis | *Bacteroides sp CAG 598* | 1.71E-06 | 3.41E-07 | 0.00233 |
| Peptidoglycan biosynthesis I (meso diaminopimelate containing) | Cell wall & Peptidoglycan Synthesis | *Bacteroides sp CAG 598* | 1.84E-06 | 3.67E-07 | 0.00233 |
| L-lysine biosynthesis VI | Amino Acid Biosynthesis | *Bacteroides sp CAG 598* | 1.74E-06 | 3.46E-07 | 0.00233 |
| Mannosylglycerate biosynthesis I | Carbohydrate Metabolism | *Escherichia coli* | 5.25E-07 | 9.94E-08 | 0.00233 |
| CDP diacylglycerol biosynthesis I | Lipid & Membrane Metabolism | *Bacteroides sp CAG 598* | 1.41E-06 | 2.81E-07 | 0.00233 |
| UMP biosynthesis I | Nucleotide Metabolism & Biosynthesis | *Bacteroides sp CAG 598* | 1.89E-06 | 3.76E-07 | 0.00233 |
| Inosine-5-phosphate degradation | Nucleotide Metabolism & Biosynthesis | *Bacteroides sp CAG 598* | 2.06E-06 | 4.11E-07 | 0.00233 |
| Cis vaccenate biosynthesis | Carbohydrate Metabolism | *Bacteroides sp CAG 598* | 1.64E-06 | 3.27E-07 | 0.00233 |
| Inosine-5-phosphate biosynthesis I | Nucleotide Metabolism & Biosynthesis | *Bacteroides sp CAG 598* | 1.60E-06 | 3.18E-07 | 0.00233 |
| Inosine-5-phosphate biosynthesis II | Nucleotide Metabolism & Biosynthesis | *Bacteroides sp CAG 598* | 1.56E-06 | 3.11E-07 | 0.00233 |
| S-adenosyl-L-methionine salvage I | Amino Acid Biosynthesis | *Bacteroides sp CAG 598* | 1.49E-06 | 2.97E-07 | 0.00233 |
| Chorismate biosynthesis from 3 dehydroquinate | General Biosynthetic Pathways | *Bacteroides sp CAG 598* | 1.78E-06 | 3.55E-07 | 0.00233 |
| Peptidoglycan biosynthesis III (mycobacteria) | Cell wall & Peptidoglycan Synthesis | *Bacteroides sp CAG 598* | 1.84E-06 | 3.67E-07 | 0.00233 |
| UDP-N-acetylmuramoyl pentapeptide biosynthesis II (lysine containing) | Cell wall & Peptidoglycan Synthesis | *Bacteroides sp CAG 598* | 1.80E-06 | 3.60E-07 | 0.00233 |
| UDP-N-acetylmuramoyl pentapeptide biosynthesis I (meso diaminopimelate containing) | Cell wall & Peptidoglycan Synthesis | *Bacteroides sp CAG 598* | 1.82E-06 | 3.62E-07 | 0.00233 |
| Adenine and adenosine salvage III | Nucleotide Metabolism & Biosynthesis | *Bacteroides sp CAG 598* | 2.01E-06 | 4.02E-07 | 0.00233 |
| Queuosine biosynthesis I (de novo) | Nucleotide Metabolism & Biosynthesis | *Bacteroides sp CAG 598* | 2.25E-06 | 4.51E-07 | 0.00233 |
| PreQ0 biosynthesis | Nucleotide Metabolism & Biosynthesis | *Bacteroides sp CAG 598* | 1.47E-06 | 2.93E-07 | 0.00233 |
| Pyrimidine deoxyribonucleosides salvage | Nucleotide Metabolism & Biosynthesis | *Bacteroides sp CAG 598* | 1.68E-06 | 3.34E-07 | 0.00233 |
| Guanosine ribonucleotides de novo biosynthesis | Nucleotide Metabolism & Biosynthesis | *Bacteroides sp CAG 598* | 1.44E-06 | 2.86E-07 | 0.00233 |
| Gondoate biosynthesis (anaerobic) | Carbohydrate Metabolism | *Bacteroides sp CAG 598* | 1.72E-06 | 3.44E-07 | 0.00233 |
| UMP biosynthesis II | Nucleotide Metabolism & Biosynthesis | *Bacteroides sp CAG 598* | 1.89E-06 | 3.76E-07 | 0.00233 |
| UMP biosynthesis III | Nucleotide Metabolism & Biosynthesis | *Bacteroides sp CAG 598* | 1.89E-06 | 3.76E-07 | 0.00233 |
| Coenzyme A biosynthesis II (eukaryotic) | Cofactor & Vitamin Biosynthesis | *Bacteroides sp CAG 598* | 1.54E-06 | 3.07E-07 | 0.00233 |
| UDP-N-acetylmuramoyl pentapeptide biosynthesis III (meso diaminopimelate containing) | Cell wall & Peptidoglycan Synthesis | *Bacteroides sp CAG 598* | 1.72E-06 | 3.44E-07 | 0.00233 |
| L-methionine biosynthesis IV | Amino Acid Biosynthesis | *Bacteroides sp CAG 598* | 1.70E-06 | 3.39E-07 | 0.00233 |
| CDP-diacylglycerol biosynthesis II | Lipid & Membrane Metabolism | *Bacteroides sp CAG 598* | 1.41E-06 | 2.81E-07 | 0.00233 |
| Fatty acid biosynthesis initiation (mitochondria) | Lipid & Membrane Metabolism | *Bacteroides sp CAG 443* | 1.45E-06 | 2.88E-07 | 0.00233 |
| NAD de novo biosynthesis I (from aspartate) | Nucleotide Metabolism & Biosynthesis | *Bacteroides sp CAG 598* | 1.69E-06 | 3.37E-07 | 0.00233 |
| L-rhamnose degradation I | Carbohydrate Metabolism | *Bacteroides sp CAG 598* | 1.69E-06 | 3.37E-07 | 0.00233 |
| L-valine biosynthesis | Amino Acid Biosynthesis | *Bacteroides sp CAG 598* | 1.69E-06 | 3.37E-07 | 0.00233 |
| Folate transformations II (plants) | Cofactor & Vitamin Biosynthesis | *Bacteroides sp CAG 598* | 2.53E-06 | 5.07E-07 | 0.00234 |
| dTDP-beta-L-rhamnose biosynthesis | Carbohydrate Metabolism | *Bacteroides sp CAG 598* | 3.87E-06 | 7.80E-07 | 0.00249 |
| Superpathway of aromatic amino acid biosynthesis | Amino Acid Biosynthesis | *Bifidobacterium pseudocatenulatum* | 5.69E-07 | 1.16E-07 | 0.00301 |
| Folate transformations II (plants) | Cofactor & Vitamin Biosynthesis | *Bifidobacterium pseudocatenulatum* | 4.97E-07 | 1.03E-07 | 0.00380 |
| Superpathway of L-phenylalanine biosynthesis | Amino Acid Biosynthesis | *Bifidobacterium pseudocatenulatum* | 4.90E-07 | 1.02E-07 | 0.00384 |
| Chorismate biosynthesis from 3 dehydroquinate | General Biosynthetic Pathways | *Bifidobacterium pseudocatenulatum* | 5.21E-07 | 1.10E-07 | 0.00449 |
| Chorismate biosynthesis I | General Biosynthetic Pathways | *Bifidobacterium pseudocatenulatum* | 5.28E-07 | 1.12E-07 | 0.00473 |
| Methylerythritol phosphate pathway I | General Biosynthetic Pathways | *Bifidobacterium pseudocatenulatum* | 4.39E-07 | 9.31E-08 | 0.00489 |
| Superpathway of L-tryptophan biosynthesis | Amino Acid Biosynthesis | *Bifidobacterium pseudocatenulatum* | 4.16E-07 | 8.92E-08 | 0.00561 |
| dTDP-beta-L-rhamnose biosynthesis | Carbohydrate Metabolism | *Bacteroides sp CAG 443* | 6.57E-06 | 1.43E-06 | 0.00651 |
| S-adenosyl-L-methionine salvage I | Amino Acid Biosynthesis | *Bifidobacterium pseudocatenulatum* | 7.23E-07 | 1.57E-07 | 0.00651 |
| Guanosine ribonucleotides de novo biosynthesis | Nucleotide Metabolism & Biosynthesis | *Bifidobacterium pseudocatenulatum* | 4.66E-07 | 1.01E-07 | 0.00662 |
| Coenzyme A biosynthesis II (eukaryotic) | Cofactor & Vitamin Biosynthesis | *Bifidobacterium pseudocatenulatum* | 4.81E-07 | 1.05E-07 | 0.00697 |
| Queuosine biosynthesis I (de novo) | Nucleotide Metabolism & Biosynthesis | *Bacteroides sp CAG 443* | 3.78E-06 | 8.29E-07 | 0.00712 |
| S-adenosyl-L-methionine salvage I | Amino Acid Biosynthesis | *Eubacterium sp CAG 38* | 1.04E-06 | 2.32E-07 | 0.00878 |
| Superpathway of coenzyme A biosynthesis III (mammals) | Cofactor & Vitamin Biosynthesis | *Bifidobacterium pseudocatenulatum* | 4.53E-07 | 1.01E-07 | 0.00922 |
| Sucrose degradation IV (sucrose phosphorylase) | Carbohydrate Metabolism | *Bifidobacterium pseudocatenulatum* | 6.27E-07 | 1.41E-07 | 0.00922 |
| Chitin degradation II (Vibrio) | Carbohydrate Metabolism | *Bifidobacterium pseudocatenulatum* | 4.68E-07 | 1.05E-07 | 0.00922 |
| Inosine-5-phosphate biosynthesis II | Nucleotide Metabolism & Biosynthesis | *Bifidobacterium pseudocatenulatum* | 3.90E-07 | 8.82E-08 | 0.01034 |
| Glycogen biosynthesis I (from ADP D Glucose) | Carbohydrate Metabolism | *Bifidobacterium pseudocatenulatum* | 5.96E-07 | 1.36E-07 | 0.01065 |
| Glycolysis IV | Carbohydrate Metabolism | *Bacteroides sp CAG 443* | 4.64E-06 | 1.06E-06 | 0.01065 |
| UDP-N-acetylmuramoyl pentapeptide biosynthesis II (lysine containing) | Cell wall & Peptidoglycan Synthesis | *Bifidobacterium pseudocatenulatum* | 3.35E-07 | 7.63E-08 | 0.01065 |
| UMP biosynthesis II | Nucleotide Metabolism & Biosynthesis | *Bacteroides sp CAG 443* | 3.67E-06 | 8.38E-07 | 0.01065 |
| UMP biosynthesis III | Nucleotide Metabolism & Biosynthesis | *Bacteroides sp CAG 443* | 3.67E-06 | 8.38E-07 | 0.01065 |
| Gamma glutamyl cycle | General Biosynthetic Pathways | *Escherichia coli* | 5.61E-07 | 1.28E-07 | 0.01067 |
| L-valine biosynthesis | Amino Acid Biosynthesis | *Bacteroides sp CAG 443* | 2.77E-06 | 6.34E-07 | 0.01067 |
| Bifidobacterium shunt | Carbohydrate Metabolism | *Bifidobacterium pseudocatenulatum* | 4.83E-07 | 1.11E-07 | 0.01104 |
| Superpathway of coenzyme A biosynthesis I (bacteria) | Cofactor & Vitamin Biosynthesis | *Bacteroides sp CAG 443* | 2.94E-06 | 6.76E-07 | 0.01109 |
| Folate transformations II (plants) | Cofactor & Vitamin Biosynthesis | *Bacteroides sp CAG 443* | 3.12E-06 | 7.18E-07 | 0.01149 |
| L-valine biosynthesis | Amino Acid Biosynthesis | *Eubacterium sp CAG 38* | 7.68E-07 | 1.77E-07 | 0.01149 |
| L-lysine biosynthesis VI | Amino Acid Biosynthesis | *Eubacterium sp CAG 38* | 7.79E-07 | 1.80E-07 | 0.01152 |
| L-lysine biosynthesis VI | Amino Acid Biosynthesis | *Bacteroides sp CAG 443* | 3.16E-06 | 7.32E-07 | 0.01167 |
| Peptidoglycan maturation (meso diaminopimelate containing) | Cell wall & Peptidoglycan Synthesis | *Bifidobacterium pseudocatenulatum* | 7.23E-07 | 1.67E-07 | 0.01167 |
| Folate transformations III (E coli) | Cofactor & Vitamin Biosynthesis | *Bacteroides sp CAG 443* | 2.92E-06 | 6.87E-07 | 0.01206 |
| Coenzyme A biosynthesis I (prokaryotic) | Cofactor & Vitamin Biosynthesis | *Bacteroides sp CAG 443* | 3.56E-06 | 8.30E-07 | 0.01206 |
| CDP diacylglycerol biosynthesis I | Lipid & Membrane Metabolism | *Bacteroides sp CAG 443* | 2.79E-06 | 6.52E-07 | 0.01206 |
| UMP biosynthesis I | Nucleotide Metabolism & Biosynthesis | *Bacteroides sp CAG 443* | 3.64E-06 | 8.46E-07 | 0.01206 |
| Inosine-5-phosphate biosynthesis II | Nucleotide Metabolism & Biosynthesis | *Bacteroides sp CAG 443* | 3.21E-06 | 7.48E-07 | 0.01206 |
| PreQ0 biosynthesis | Nucleotide Metabolism & Biosynthesis | *Bacteroides sp CAG 443* | 4.19E-06 | 9.80E-07 | 0.01206 |
| Inosine-5-phosphate biosynthesis III | Nucleotide Metabolism & Biosynthesis | *Bifidobacterium pseudocatenulatum* | 3.77E-07 | 8.78E-08 | 0.01206 |
| UMP biosynthesis II | Nucleotide Metabolism & Biosynthesis | *Bifidobacterium pseudocatenulatum* | 4.37E-07 | 1.03E-07 | 0.01206 |
| UMP biosynthesis III | Nucleotide Metabolism & Biosynthesis | *Bifidobacterium pseudocatenulatum* | 4.37E-07 | 1.03E-07 | 0.01206 |
| CDP-diacylglycerol biosynthesis II | Lipid & Membrane Metabolism | *Bacteroides sp CAG 443* | 2.79E-06 | 6.52E-07 | 0.01206 |
| tRNA charging | Amino Acid Biosynthesis | *Bacteroides sp CAG 443* | 3.26E-06 | 7.63E-07 | 0.01206 |
| UDP-N-acetyl-D-glucosamine biosynthesis I | Cell wall & Peptidoglycan Synthesis | *Bifidobacterium pseudocatenulatum* | 4.78E-07 | 1.12E-07 | 0.01206 |
| Inosine-5-phosphate biosynthesis I | Nucleotide Metabolism & Biosynthesis | *Bifidobacterium pseudocatenulatum* | 4.01E-07 | 9.43E-08 | 0.01207 |
| Peptidoglycan biosynthesis I (meso diaminopimelate containing) | Cell wall & Peptidoglycan Synthesis | *Bacteroides sp CAG 443* | 3.15E-06 | 7.42E-07 | 0.01211 |
| Inosine-5-phosphate biosynthesis I | Nucleotide Metabolism & Biosynthesis | *Bacteroides sp CAG 443* | 3.27E-06 | 7.70E-07 | 0.01211 |
| L-histidine biosynthesis | Amino Acid Biosynthesis | *Bacteroides sp CAG 443* | 2.74E-06 | 6.47E-07 | 0.01211 |
| Guanosine ribonucleotides de novo biosynthesis | Nucleotide Metabolism & Biosynthesis | *Bacteroides sp CAG 443* | 3.07E-06 | 7.25E-07 | 0.01211 |
| O-antigen building blocks biosynthesis (E coli) | General Biosynthetic Pathways | *Bifidobacterium pseudocatenulatum* | 4.01E-07 | 9.47E-08 | 0.01216 |
| Chorismate biosynthesis from 3 dehydroquinate | General Biosynthetic Pathways | *Bacteroides sp CAG 443* | 2.58E-06 | 6.10E-07 | 0.01219 |
| Phosphopantothenate biosynthesis I | Lipid & Membrane Metabolism | *Bacteroides sp CAG 443* | 2.41E-06 | 5.71E-07 | 0.01219 |
| Coenzyme A biosynthesis II (eukaryotic) | Cofactor & Vitamin Biosynthesis | *Bacteroides sp CAG 443* | 3.53E-06 | 8.35E-07 | 0.01219 |
| L-isoleucine biosynthesis I (from threonine) | Amino Acid Biosynthesis | *Bifidobacterium pseudocatenulatum* | 5.96E-07 | 1.42E-07 | 0.01264 |
| L-isoleucine biosynthesis III | Amino Acid Biosynthesis | *Bifidobacterium pseudocatenulatum* | 5.77E-07 | 1.37E-07 | 0.01264 |
| Chorismate biosynthesis I | General Biosynthetic Pathways | *Bacteroides sp CAG 443* | 2.55E-06 | 6.07E-07 | 0.01268 |
| Peptidoglycan biosynthesis III (mycobacteria) | Cell wall & Peptidoglycan Synthesis | *Bacteroides sp CAG 443* | 3.14E-06 | 7.49E-07 | 0.01268 |
| Pyruvate fermentation to acetate and S-lactate I | General Biosynthetic Pathways | *Bifidobacterium pseudocatenulatum* | 3.56E-07 | 8.50E-08 | 0.01291 |
| Pyruvate fermentation to acetate and lactate II | General Biosynthetic Pathways | *Bifidobacterium pseudocatenulatum* | 3.56E-07 | 8.50E-08 | 0.01291 |
| UDP-N-acetylmuramoyl pentapeptide biosynthesis I (meso diaminopimelate containing) | Cell wall & Peptidoglycan Synthesis | *Bacteroides sp CAG 443* | 3.09E-06 | 7.38E-07 | 0.01291 |
| Superpathway of L-threonine biosynthesis | Amino Acid Biosynthesis | *Bifidobacterium pseudocatenulatum* | 5.58E-07 | 1.33E-07 | 0.01292 |
| Glycogen degradation II | Carbohydrate Metabolism | *Bifidobacterium pseudocatenulatum* | 6.66E-07 | 1.60E-07 | 0.01334 |
| L-methionine biosynthesis IV | Amino Acid Biosynthesis | *Bacteroides sp CAG 443* | 3.20E-06 | 7.68E-07 | 0.01345 |
| UDP-N-acetylmuramoyl pentapeptide biosynthesis II (lysine containing) | Cell wall & Peptidoglycan Synthesis | *Bacteroides sp CAG 443* | 3.09E-06 | 7.43E-07 | 0.01360 |
| Sucrose biosynthesis II | General Biosynthetic Pathways | *Bifidobacterium pseudocatenulatum* | 6.87E-07 | 1.65E-07 | 0.01360 |
| Superpathway of branched chain amino acid biosynthesis | Amino Acid Biosynthesis | *Bifidobacterium pseudocatenulatum* | 5.85E-07 | 1.41E-07 | 0.01360 |
| 5-aminoimidazole ribonucleotide biosynthesis II | Nucleotide Metabolism & Biosynthesis | *Bacteroides sp CAG 443* | 2.90E-06 | 7.00E-07 | 0.01360 |
| Superpathway of 5-aminoimidazole ribonucleotide biosynthesis | Nucleotide Metabolism & Biosynthesis | *Bacteroides sp CAG 443* | 2.90E-06 | 7.00E-07 | 0.01360 |
| Coenzyme A biosynthesis I (prokaryotic) | Cofactor & Vitamin Biosynthesis | *Bifidobacterium pseudocatenulatum* | 5.15E-07 | 1.25E-07 | 0.01382 |
| UDP-N-acetylmuramoyl pentapeptide biosynthesis III (meso diaminopimelate containing) | Cell wall & Peptidoglycan Synthesis | *Bacteroides sp CAG 443* | 2.80E-06 | 6.78E-07 | 0.01403 |
| L-ornithine biosynthesis II | Amino Acid Biosynthesis | *Bacteroides sp CAG 443* | 2.90E-06 | 7.05E-07 | 0.01450 |
| UMP biosynthesis I | Nucleotide Metabolism & Biosynthesis | *Bifidobacterium pseudocatenulatum* | 4.30E-07 | 1.05E-07 | 0.01589 |
| L-valine biosynthesis | Amino Acid Biosynthesis | *Bifidobacterium pseudocatenulatum* | 5.90E-07 | 1.45E-07 | 0.01651 |
| Seleno amino acid biosynthesis (plants) | Amino Acid Biosynthesis | *Eubacterium sp CAG 38* | 8.65E-07 | 2.13E-07 | 0.01694 |
| Inosine-5-phosphate degradation | Nucleotide Metabolism & Biosynthesis | *Bacteroides sp CAG 443* | 2.89E-06 | 7.13E-07 | 0.01694 |
| 5-aminoimidazole ribonucleotide biosynthesis I | Nucleotide Metabolism & Biosynthesis | *Bacteroides sp CAG 443* | 2.88E-06 | 7.11E-07 | 0.01712 |
| 5-aminoimidazole ribonucleotide biosynthesis I | Nucleotide Metabolism & Biosynthesis | *Clostridium sp CAG 138* | 6.87E-07 | 1.70E-07 | 0.01768 |
| L-arginine biosynthesis II (acetyl cycle) | Amino Acid Biosynthesis | *Bifidobacterium pseudocatenulatum* | 3.28E-07 | 8.16E-08 | 0.01804 |
| Glucose and glucose 1 phosphate degradation | Carbohydrate Metabolism | *Escherichia coli* | 4.12E-07 | 1.02E-07 | 0.01804 |
| dTDP-beta-L-rhamnose biosynthesis | Carbohydrate Metabolism | *Bifidobacterium pseudocatenulatum* | 3.05E-07 | 7.59E-08 | 0.01821 |
| Methylerythritol phosphate pathway I | General Biosynthetic Pathways | *Bacteroides sp CAG 443* | 3.23E-06 | 8.06E-07 | 0.01870 |
| Tetrapyrrole biosynthesis I (from glutamate) | Cofactor & Vitamin Biosynthesis | *Eubacterium sp CAG 38* | 6.16E-07 | 1.54E-07 | 0.01912 |
| L-arginine biosynthesis II (acetyl cycle) | Amino Acid Biosynthesis | *Eubacterium sp CAG 38* | 9.33E-07 | 2.34E-07 | 0.01985 |
| Sucrose biosynthesis II | General Biosynthetic Pathways | *Ruminococcus sp CAG 254* | 2.15E-06 | 5.41E-07 | 0.02072 |
| Methylerythritol phosphate pathway I | General Biosynthetic Pathways | *Eubacterium sp CAG 38* | 7.02E-07 | 1.77E-07 | 0.02094 |
| 5-aminoimidazole ribonucleotide biosynthesis I | Nucleotide Metabolism & Biosynthesis | *Bifidobacterium pseudocatenulatum* | 3.12E-07 | 7.89E-08 | 0.02159 |
| Purine ribonucleosides degradation | Nucleotide Metabolism & Biosynthesis | *Eubacterium sp CAG 38* | 7.93E-07 | 2.01E-07 | 0.02229 |
| Phosphopantothenate biosynthesis I | Lipid & Membrane Metabolism | *Prevotella sp CAG 1185* | 6.45E-06 | 1.64E-06 | 0.02247 |
| L-tryptophan biosynthesis | Amino Acid Biosynthesis | *Bifidobacterium pseudocatenulatum* | 2.94E-07 | 7.53E-08 | 0.02414 |
| L-histidine degradation I | Amino Acid Biosynthesis | *Prevotella sp CAG 1185* | 5.58E-06 | 1.43E-06 | 0.02445 |
| 5-aminoimidazole ribonucleotide biosynthesis II | Nucleotide Metabolism & Biosynthesis | *Bifidobacterium pseudocatenulatum* | 3.36E-07 | 8.67E-08 | 0.02614 |
| Superpathway of 5-aminoimidazole ribonucleotide biosynthesis | Nucleotide Metabolism & Biosynthesis | *Bifidobacterium pseudocatenulatum* | 3.36E-07 | 8.67E-08 | 0.02614 |
| Adenine and adenosine salvage III | Nucleotide Metabolism & Biosynthesis | *Bacteroides sp CAG 443* | 3.22E-06 | 8.32E-07 | 0.02614 |
| Queuosine biosynthesis I (de novo) | Nucleotide Metabolism & Biosynthesis | *Eubacterium sp CAG 38* | 7.28E-07 | 1.89E-07 | 0.02828 |
| L-lysine biosynthesis I | Amino Acid Biosynthesis | *Bifidobacterium pseudocatenulatum* | 4.59E-07 | 1.20E-07 | 0.02854 |
| Adenine and adenosine salvage III | Nucleotide Metabolism & Biosynthesis | *Eubacterium sp CAG 38* | 7.14E-07 | 1.86E-07 | 0.02898 |
| L-valine biosynthesis | Amino Acid Biosynthesis | *Prevotella sp CAG 1185* | 6.32E-06 | 1.65E-06 | 0.02931 |
| 5-aminoimidazole ribonucleotide biosynthesis II | Nucleotide Metabolism & Biosynthesis | *Clostridium sp CAG 138* | 6.62E-07 | 1.73E-07 | 0.02960 |
| Superpathway of 5-aminoimidazole ribonucleotide biosynthesis | Nucleotide Metabolism & Biosynthesis | *Clostridium sp CAG 138* | 6.62E-07 | 1.73E-07 | 0.02960 |
| dTDP-beta-L-rhamnose biosynthesis | Carbohydrate Metabolism | *Eubacterium sp CAG 38* | 8.22E-07 | 2.15E-07 | 0.02985 |
| L-valine biosynthesis | Amino Acid Biosynthesis | *Ruminococcus sp CAG 254* | 1.37E-06 | 3.60E-07 | 0.03076 |
| L-histidine biosynthesis | Amino Acid Biosynthesis | *Eubacterium sp CAG 38* | 6.34E-07 | 1.67E-07 | 0.03076 |
| UMP biosynthesis I | Nucleotide Metabolism & Biosynthesis | *Eubacterium sp CAG 38* | 8.04E-07 | 2.12E-07 | 0.03119 |
| UMP biosynthesis II | Nucleotide Metabolism & Biosynthesis | *Eubacterium sp CAG 38* | 8.04E-07 | 2.12E-07 | 0.03119 |
| UMP biosynthesis III | Nucleotide Metabolism & Biosynthesis | *Eubacterium sp CAG 38* | 8.04E-07 | 2.12E-07 | 0.03119 |
| UDP-N-acetylmuramoyl pentapeptide biosynthesis I (meso diaminopimelate containing) | Cell wall & Peptidoglycan Synthesis | *Ruminococcus sp CAG 254* | 1.21E-06 | 3.22E-07 | 0.03422 |
| Superpathway of coenzyme A biosynthesis III (mammals) | Cofactor & Vitamin Biosynthesis | *Bacteroides sp CAG 443* | 1.48E-06 | 3.94E-07 | 0.03480 |
| Glycogen biosynthesis I (from ADP D Glucose) | Carbohydrate Metabolism | *Eubacterium sp CAG 38* | 9.51E-07 | 2.54E-07 | 0.03488 |
| Guanosine ribonucleotides de novo biosynthesis | Nucleotide Metabolism & Biosynthesis | *Eubacterium sp CAG 38* | 7.04E-07 | 1.88E-07 | 0.03546 |
| Glycogen biosynthesis I (from ADP D Glucose) | Carbohydrate Metabolism | *Ruminococcus sp CAG 254* | 1.30E-06 | 3.48E-07 | 0.03591 |
| Chorismate biosynthesis from 3 dehydroquinate | General Biosynthetic Pathways | *Eubacterium sp CAG 38* | 8.92E-07 | 2.40E-07 | 0.03657 |
| Superpathway of coenzyme A biosynthesis I (bacteria) | Cofactor & Vitamin Biosynthesis | *Prevotella sp CAG 1185* | 6.29E-06 | 1.69E-06 | 0.03709 |
| Folate transformations II (plants) | Cofactor & Vitamin Biosynthesis | *Prevotella sp CAG 1185* | 6.07E-06 | 1.63E-06 | 0.03709 |
| L-tryptophan biosynthesis | Amino Acid Biosynthesis | *Eubacterium sp CAG 38* | 6.79E-07 | 1.83E-07 | 0.03719 |
| L-isoleucine biosynthesis I (from threonine) | Amino Acid Biosynthesis | *Ruminococcus sp CAG 254* | 1.25E-06 | 3.40E-07 | 0.04035 |
| Pyruvate fermentation to isobutanol (engineered) | General Biosynthetic Pathways | *Bifidobacterium pseudocatenulatum* | 3.48E-07 | 9.47E-08 | 0.04036 |
| UDP-N-acetylmuramoyl pentapeptide biosynthesis I (meso diaminopimelate containing) | Cell wall & Peptidoglycan Synthesis | *Eubacterium sp CAG 38* | 8.40E-07 | 2.28E-07 | 0.04055 |
| Superpathway of L-serine and glycine biosynthesis I | Amino Acid Biosynthesis | *Bifidobacterium pseudocatenulatum* | 4.99E-07 | 1.37E-07 | 0.04369 |
| Peptidoglycan biosynthesis III (mycobacteria) | Cell wall & Peptidoglycan Synthesis | *Bifidobacterium pseudocatenulatum* | 3.25E-07 | 8.92E-08 | 0.04386 |
| L-lysine biosynthesis VI | Amino Acid Biosynthesis | *Ruminococcus sp CAG 254* | 8.39E-07 | 2.31E-07 | 0.04468 |
| S-adenosyl-L-methionine salvage I | Amino Acid Biosynthesis | *Ruminococcus sp CAG 254* | 1.25E-06 | 3.45E-07 | 0.04468 |
| Superpathway of branched chain amino acid biosynthesis | Amino Acid Biosynthesis | *Ruminococcus sp CAG 254* | 1.31E-06 | 3.60E-07 | 0.04478 |
| L-arginine biosynthesis I (via L-ornithine) | Amino Acid Biosynthesis | *Eubacterium sp CAG 38* | 7.71E-07 | 2.12E-07 | 0.04502 |
| Superpathway of coenzyme A biosynthesis III (mammals) | Cofactor & Vitamin Biosynthesis | *Prevotella sp CAG 1185* | 6.19E-06 | 1.71E-06 | 0.04613 |
| L-isoleucine biosynthesis III | Amino Acid Biosynthesis | *Ruminococcus sp CAG 254* | 1.16E-06 | 3.20E-07 | 0.04613 |
| Superpathway of L-lysine, L-threonine and L-methionine biosynthesis II | Amino Acid Biosynthesis | *Bifidobacterium pseudocatenulatum* | 3.54E-07 | 9.79E-08 | 0.04613 |
| UDP-N-acetylmuramoyl pentapeptide biosynthesis II (lysine containing) | Cell wall & Peptidoglycan Synthesis | *Ruminococcus sp CAG 254* | 1.29E-06 | 3.58E-07 | 0.04647 |
| Coenzyme A biosynthesis II (eukaryotic) | Cofactor & Vitamin Biosynthesis | *Prevotella sp CAG 1185* | 6.03E-06 | 1.67E-06 | 0.04649 |
| L-ornithine biosynthesis I | Amino Acid Biosynthesis | *Eubacterium sp CAG 38* | 5.76E-07 | 1.60E-07 | 0.04689 |
| Peptidoglycan biosynthesis I (meso diaminopimelate containing) | Cell wall & Peptidoglycan Synthesis | *Ruminococcus sp CAG 254* | 1.12E-06 | 3.11E-07 | 0.04747 |
| Superpathway of adenosine nucleotides de novo biosynthesis I | Nucleotide Metabolism & Biosynthesis | *Prevotella sp CAG 1185* | 6.64E-06 | 1.85E-06 | 0.04778 |

Differential abundance analyses were conducted using the R package MaAsLin2 (v1.15.1), employing linear regressions adjusted for age, sex, and BMI. Relative abundances were normalised using the centred log-ratio (CLR) method. Adjustments were performed using the Benjamini–Hochberg False FDR method, and all results presented meet the significance threshold of q<0.05.

**Table S4. Metabolic pathway categories and corresponding individual pathways as referenced in Figure 3.**

|  | **Individual metabolic pathway** | |
| --- | --- | --- |
| **Metabolic pathway category** | **Figure 3A** | **Figure 3C** |
| **Amino Acid Biosynthesis** | L-arginine biosynthesis I (via L-ornithine) |  |
|  | L-arginine biosynthesis II (acetyl cycle) |  |
|  | L-histidine biosynthesis | L-histidine biosynthesis |
|  | L-histidine degradation I |  |
|  | L-isoleucine biosynthesis I (from threonine) |  |
|  | L-isoleucine biosynthesis III |  |
|  | L-lysine biosynthesis I |  |
|  | L-lysine biosynthesis VI | L-lysine biosynthesis VI |
|  | L-methionine biosynthesis IV |  |
|  | L-ornithine biosynthesis I |  |
|  | L-ornithine biosynthesis II |  |
|  | L-tryptophan biosynthesis |  |
|  | L-valine biosynthesis | L-valine biosynthesis |
|  | S-adenosyl-L-methionine salvage I | S-adenosyl-L-methionine salvage I |
|  | Seleno amino acid biosynthesis (plants) |  |
|  | Superpathway of aromatic amino acid biosynthesis |  |
|  | Superpathway of branched chain amino acid biosynthesis |  |
|  | Superpathway of L-lysine, L-threonine and L-methionine biosynthesis II |  |
|  | Superpathway of L-phenylalanine biosynthesis |  |
|  | Superpathway of L-serine and glycine biosynthesis I |  |
|  | Superpathway of L-threonine biosynthesis | Superpathway of L-threonine biosynthesis |
|  | Superpathway of L-tryptophan biosynthesis |  |
|  | tRNA charging | tRNA charging |
| **Carbohydrate Metabolism** | Bifidobacterium shunt |  |
|  | Chitin degradation II (Vibrio) |  |
|  | Cis vaccenate biosynthesis |  |
|  | dTDP-beta-L-rhamnose biosynthesis |  |
|  | Glucose and glucose 1 phosphate degradation |  |
|  | Glycogen biosynthesis I (from ADP D Glucose) |  |
|  | Glycogen degradation II |  |
|  | Glycolysis IV |  |
|  | Gondoate biosynthesis (anaerobic) |  |
|  | L-rhamnose degradation I |  |
|  | Mannosylglycerate biosynthesis I |  |
|  | Sucrose degradation IV (sucrose phosphorylase) |  |
| **Cell wall & Peptidoglycan Synthesis** | Peptidoglycan biosynthesis I (meso diaminopimelate containing) | Peptidoglycan biosynthesis I (meso-diaminopimelate containing) |
|  | Peptidoglycan biosynthesis III (mycobacteria) | Peptidoglycan biosynthesis III (mycobacteria) |
|  | Peptidoglycan maturation (meso diaminopimelate containing) |  |
|  | UDP-N-acetyl-D-glucosamine biosynthesis I |  |
|  | UDP-N-acetylmuramoyl pentapeptide biosynthesis I (meso diaminopimelate containing) | UDP-N-acetylmuramoyl-pentapeptide biosynthesis I (meso-diaminopimelate containing) |
|  | UDP-N-acetylmuramoyl pentapeptide biosynthesis II (lysine containing) | UDP-N-acetylmuramoyl-pentapeptide biosynthesis II (lysine-containing) |
|  | UDP-N-acetylmuramoyl pentapeptide biosynthesis III (meso diaminopimelate containing) |  |
| **Cofactor & Vitamin Biosynthesis** | Coenzyme A biosynthesis I (prokaryotic) | Coenzyme A biosynthesis I (prokaryotic) |
|  | Coenzyme A biosynthesis II (eukaryotic) | Coenzyme A biosynthesis II (eukaryotic) |
|  |  | Flavin biosynthesis I (bacteria and plants) |
|  | Folate transformations II (plants) | Folate transformations II (plants) |
|  | Folate transformations III (E coli) |  |
|  | Superpathway of coenzyme A biosynthesis I (bacteria) |  |
|  | Superpathway of coenzyme A biosynthesis III (mammals) | Superpathway of coenzyme A biosynthesis III (mammals) |
|  | Tetrapyrrole biosynthesis I (from glutamate) |  |
| **General Biosynthetic Pathways** | Chorismate biosynthesis from 3 dehydroquinate | Chorismate biosynthesis from 3-dehydroquinate |
|  | Chorismate biosynthesis I | Chorismate biosynthesis I |
|  | Gamma glutamyl cycle |  |
|  |  | Glycogen biosynthesis I (from ADP-D-Glucose) |
|  | Methylerythritol phosphate pathway I | Methylerythritol phosphate pathway I |
|  | O-antigen building blocks biosynthesis (E coli) |  |
|  | Pyruvate fermentation to acetate and lactate II |  |
|  | Pyruvate fermentation to acetate and S-lactate I |  |
|  | Pyruvate fermentation to isobutanol (engineered) |  |
|  | Sucrose biosynthesis II | Sucrose biosynthesis II |
| **Lipid & Membrane Metabolism** | CDP diacylglycerol biosynthesis I | CDP diacylglycerol biosynthesis I |
|  | CDP-diacylglycerol biosynthesis II | CDP diacylglycerol biosynthesis II |
|  | Fatty acid biosynthesis initiation (mitochondria) | Fatty acid biosynthesis initiation (mitochondria) |
|  | Phosphopantothenate biosynthesis I | Phosphopantothenate biosynthesis I |
| **Nucleotide Metabolism & Biosynthesis** | 5-aminoimidazole ribonucleotide biosynthesis I | 5-aminoimidazole ribonucleotide biosynthesis I |
|  | 5-aminoimidazole ribonucleotide biosynthesis II | 5-aminoimidazole ribonucleotide biosynthesis II |
|  | Adenine and adenosine salvage III | Adenine and adenosine salvage III |
|  | Guanosine ribonucleotides de novo biosynthesis | Guanosine ribonucleotides de novo biosynthesis |
|  | Inosine-5-phosphate biosynthesis I |  |
|  | Inosine-5-phosphate biosynthesis II |  |
|  | Inosine-5-phosphate biosynthesis III |  |
|  | Inosine-5-phosphate degradation | Inosine 5-phosphate degradation |
|  | NAD de novo biosynthesis I (from aspartate) |  |
|  | PreQ0 biosynthesis |  |
|  | Purine ribonucleosides degradation | Purine ribonucleosides degradation |
|  | Pyrimidine deoxyribonucleosides salvage |  |
|  | Queuosine biosynthesis I (de novo) | Queuosine biosynthesis I (de novo) |
|  | Superpathway of 5-aminoimidazole ribonucleotide biosynthesis | Superpathway of 5-aminoimidazole ribonucleotide biosynthesis |
|  | Superpathway of adenosine nucleotides de novo biosynthesis I |  |
|  | UMP biosynthesis I | UMP biosynthesis I |
|  | UMP biosynthesis II | UMP biosynthesis II |
|  | UMP biosynthesis III | UMP biosynthesis III |

**Table S5. Differential abundance analysis at the microbial species level in relation to the peripheral immune system.**

| **Species** | |  |  |  |  |  |
| --- | --- | --- | --- | --- | --- | --- |
| **Metaphlan/NCBI (GTDB taxa)** | **Genus** | **Immune cell population/subtype** | **Immune category** | **β coefficient** | **SE** | **q-value (FDR)** |
| *GGB4583 SGB6334 (Merdicola sp900555085)* | *Merdicola* | IFNγ-producing Natural Killer T Cells | Adaptive Immune Cells | 1.41E-05 | 3.40E-07 | 6.80E-33 |
| *GGB4583 SGB6334 (Merdicola sp900555085)* | *Merdicola* | Natural Killer T Cells | Adaptive Immune Cells | 9.50E-06 | 3.80E-07 | 4.18E-24 |
| *Clostridium sp 1001270J 160509 D11* | *Clostridium* | PMDSCs | Innate Immune Cells | 2.22E-05 | 1.50E-06 | 1.43E-15 |
| *Clostridium sp 1001270J 160509 D11* | *Clostridium* | Mature LDNs | Innate Immune Cells | 5.14E-05 | 3.50E-06 | 2.44E-15 |
| *Clostridium sp 1001270J 160509 D11* | *Clostridium* | PD-1^+^ CD4^+^ T Cells | CD4^+^ T Cells | 9.09E-06 | 6.40E-07 | 6.49E-15 |
| *Ruminococcus bovis* | *Ruminococcus* | TNFα-producing CD161^+^ NK Cells | Natural Killer (NK) Cells | 1.85E-04 | 1.30E-05 | 2.17E-14 |
| *Ruminococcus bovis* | *Ruminococcus* | PMDSCs | Innate Immune Cells | 9.75E-05 | 7.20E-06 | 2.69E-14 |
| *Ruminococcus bovis* | *Ruminococcus* | TNFα-producing NK Cells | Natural Killer (NK) Cells | 1.93E-04 | 1.40E-05 | 2.89E-14 |
| *Clostridium sp 1001270J 160509 D11* | *Clostridium* | PD-1^+^ Activated HLA-DR^+^ CD4^+^ T Cells | CD4^+^ T Cells | 4.13E-05 | 3.00E-06 | 4.27E-14 |
| *Ruminococcus bovis* | *Ruminococcus* | PD-1^+^ CD4^+^ T Cells | CD4^+^ T Cells | 3.99E-05 | 3.10E-06 | 7.07E-14 |
| *Ruminococcus bovis* | *Ruminococcus* | PD-1^+^ Activated HLA-DR^+^ CD4^+^ T Cells | CD4^+^ T Cells | 1.78E-04 | 1.50E-05 | 3.93E-12 |
| *Ruminococcus bovis* | *Ruminococcus* | IL-17A-producing CD4^+^ T Cells | CD4^+^ T Cells | 1.30E-04 | 1.10E-05 | 7.02E-12 |
| *Ruminococcus bovis* | *Ruminococcus* | PD-1^+^ CD8^+^ T Cells | CD8^+^ T Cells | 8.48E-05 | 7.60E-06 | 3.06E-11 |
| *Ruminococcus bovis* | *Ruminococcus* | IL-17A-producing Activated CD69^+^ CD4^+^ T Cells | CD4^+^ T Cells | 1.35E-04 | 1.20E-05 | 3.61E-11 |
| *Ruminococcus bovis* | *Ruminococcus* | Non-classical Monocytes | Innate Immune Cells | 8.71E-04 | 7.90E-05 | 3.71E-11 |
| *Clostridium sp 1001270J 160509 D11* | *Clostridium* | IL-17A-producing Activated CD69^+^ CD8^+^ T Cells | CD8^+^ T Cells | 9.11E-05 | 8.40E-06 | 5.88E-11 |
| *Clostridium sp 1001270J 160509 D11* | *Clostridium* | TNFα-producing NK Cells | Natural Killer (NK) Cells | 4.08E-05 | 3.90E-06 | 6.37E-11 |
| *GGB4583 SGB6334 (Merdicola sp900555085)* | *Merdicola* | TNFα-producing Natural Killer T Cells | Adaptive Immune Cells | 3.17E-05 | 2.90E-06 | 7.63E-11 |
| *Clostridium sp 1001270J 160509 D11* | *Clostridium* | PD-1^+^ CD8^+^ T Cells | CD8^+^ T Cells | 1.87E-05 | 1.80E-06 | 7.73E-11 |
| *Streptococcus thermophilus* | *Streptococcus* | IFNγ-producing Natural Killer T Cells | Adaptive Immune Cells | 3.81E-06 | 3.70E-07 | 1.11E-10 |
| *Phocaeicola plebeius* | *Phocaeicola* | IFNγ-producing Natural Killer T Cells | Adaptive Immune Cells | 4.75E-05 | 4.80E-06 | 4.54E-10 |
| *Clostridium sp 1001270J 160509 D11* | *Clostridium* | IL-17A-producing CD4^+^ T Cells | CD4^+^ T Cells | 2.78E-05 | 2.80E-06 | 4.73E-10 |
| *GGB6601 SGB9333 (CAG-267 sp001917135)* | *Other* | CD4^+^ TEMRA | CD4^+^ T Cells | 9.84E-05 | 9.70E-06 | 4.98E-10 |
| *Phocaeicola plebeius* | *Phocaeicola* | Natural Killer T Cells | Adaptive Immune Cells | 3.26E-05 | 3.30E-06 | 5.42E-10 |
| *Streptococcus thermophilus* | *Streptococcus* | Natural Killer T Cells | Adaptive Immune Cells | 2.58E-06 | 2.60E-07 | 5.42E-10 |
| *Clostridium sp 1001270J 160509 D11* | *Clostridium* | TNFα-producing CD161^+^ NK Cells | Natural Killer (NK) Cells | 3.81E-05 | 3.90E-06 | 6.93E-10 |
| *Ruminococcus bovis* | *Ruminococcus* | Mature LDNs | Innate Immune Cells | 2.08E-04 | 2.20E-05 | 1.49E-09 |
| *GGB4571 SGB6317 (Merdicola sp001915925)* | *Merdicola* | PD-1^+^ CD4^+^ T Cells | CD4^+^ T Cells | 3.00E-05 | 3.20E-06 | 1.57E-09 |
| *GGB4571 SGB6317 (Merdicola sp001915925)* | *Merdicola* | PD-1^+^ CD8^+^ T Cells | CD8^+^ T Cells | 6.61E-05 | 7.10E-06 | 1.70E-09 |
| *GGB4571 SGB6317 (Merdicola sp001915925)* | *Merdicola* | PMDSCs | Innate Immune Cells | 7.29E-05 | 7.80E-06 | 1.78E-09 |
| *GGB9063 SGB13982 (Scatosoma sp003150575)* | *Scatosoma* | IFNγ-producing Natural Killer T Cells | Adaptive Immune Cells | 8.75E-06 | 9.50E-07 | 2.24E-09 |
| *GGB3612 SGB4882 (COE1 sp001916965)* | *Other* | PMDSCs | Innate Immune Cells | 4.74E-05 | 5.20E-06 | 3.16E-09 |
| *GGB9063 SGB13982 (Scatosoma sp003150575)* | *Scatosoma* | Natural Killer T Cells | Adaptive Immune Cells | 5.97E-06 | 6.60E-07 | 4.16E-09 |
| *Prevotella marseillensis* | *Prevotella* | IL-17A-producing Activated CD69^+^ CD8^+^ T Cells | CD8^+^ T Cells | 6.89E-04 | 7.70E-05 | 9.60E-09 |
| *GGB4571 SGB6317 (Merdicola sp001915925)* | *Merdicola* | PD-1^+^ Activated HLA-DR^+^ CD4^+^ T Cells | CD4^+^ T Cells | 1.35E-04 | 1.50E-05 | 9.83E-09 |
| *Clostridium sp 1001270J 160509 D11* | *Clostridium* | CCR7^+^ Activated NK Cells | Natural Killer (NK) Cells | 1.54E-04 | 1.70E-05 | 1.09E-08 |
| *GGB3612 SGB4882 (COE1 sp001916965)* | *Other* | IL-17A-producing Activated CD69^+^ CD8^+^ T Cells | CD8^+^ T Cells | 2.04E-04 | 2.40E-05 | 1.59E-08 |
| *Ruminococcus bovis* | *Ruminococcus* | CCR7^+^ Activated NK Cells | Natural Killer (NK) Cells | 6.76E-04 | 7.70E-05 | 1.84E-08 |
| *Ruminococcus bovis* | *Ruminococcus* | IL-17A-producing Activated CD69^+^ CD8^+^ T Cells | CD8^+^ T Cells | 3.76E-04 | 4.40E-05 | 1.90E-08 |
| *GGB3612 SGB4882 (COE1 sp001916965)* | *Other* | PD-1^+^ CD4^+^ T Cells | CD4^+^ T Cells | 1.90E-05 | 2.30E-06 | 2.42E-08 |
| *Clostridium sp 1001270J 160509 D11* | *Clostridium* | IL-17A-producing Activated CD69^+^ CD4^+^ T Cells | CD4^+^ T Cells | 2.79E-05 | 3.30E-06 | 5.01E-08 |
| *GGB4583 SGB6334 (Merdicola sp900555085)* | *Merdicola* | CD8^+^ TEMRA | CD8^+^ T Cells | 6.20E-06 | 7.20E-07 | 6.60E-08 |
| *GGB4583 SGB6334 (Merdicola sp900555085)* | *Merdicola* | IFNγ-producing CD161^+^ CD4^+^ T Cells | CD4^+^ T Cells | 1.46E-05 | 1.70E-06 | 7.30E-08 |
| *GGB4583 SGB6334 (Merdicola sp900555085)* | *Merdicola* | IFNγ-producing Activated CD69^+^ CD4^+^ T Cells | CD4^+^ T Cells | 6.57E-06 | 7.70E-07 | 8.03E-08 |
| *GGB4583 SGB6334 (Merdicola sp900555085)* | *Merdicola* | IFNγ-producing CD4^+^ EM T Cells | CD4^+^ T Cells | 1.51E-05 | 1.80E-06 | 8.38E-08 |
| *GGB4571 SGB6317 (Merdicola sp001915925)* | *Merdicola* | TNFα-producing CD161^+^ NK Cells | Natural Killer (NK) Cells | 1.30E-04 | 1.60E-05 | 1.11E-07 |
| *GGB4583 SGB6334 (Merdicola sp900555085)* | *Merdicola* | IFNγ/TNFα-producing CD161^+^ CD8^+^ T Cells | CD8^+^ T Cells | 1.16E-04 | 1.40E-05 | 1.39E-07 |
| *GGB4571 SGB6317 (Merdicola sp001915925)* | *Merdicola* | TNFα-producing NK Cells | Natural Killer (NK) Cells | 1.36E-04 | 1.70E-05 | 1.44E-07 |
| *GGB3612 SGB4882 (COE1 sp001916965)* | *Other* | PD-1^+^ Activated HLA-DR^+^ CD4^+^ T Cells | CD4^+^ T Cells | 8.46E-05 | 1.10E-05 | 1.81E-07 |
| *Ruminococcus bovis* | *Ruminococcus* | IL-17A-producing CD161^+^ CD4^+^ T Cells | CD4^+^ T Cells | 3.34E-04 | 4.10E-05 | 1.87E-07 |
| *GGB3612 SGB4882 (COE1 sp001916965)* | *Other* | Mature LDNs | Innate Immune Cells | 1.04E-04 | 1.30E-05 | 2.25E-07 |
| *GGB3612 SGB4882 (COE1 sp001916965)* | *Other* | PD-1^+^ CD8^+^ T Cells | CD8^+^ T Cells | 4.03E-05 | 5.30E-06 | 2.89E-07 |
| *GGB4583 SGB6334 (Merdicola sp900555085)* | *Merdicola* | CD4^+^ TEMRA | CD4^+^ T Cells | 2.06E-05 | 2.70E-06 | 5.21E-07 |
| *Lachnospiraceae bacterium* | *Lachnospiraceae* | Mature LDNs | Innate Immune Cells | 8.78E-05 | 1.20E-05 | 6.72E-07 |
| *GGB4571 SGB6317 (Merdicola sp001915925)* | *Merdicola* | IL-17A-producing CD4^+^ T Cells | CD4^+^ T Cells | 9.11E-05 | 1.20E-05 | 1.15E-06 |
| *GGB6601 SGB9333 (CAG-267 sp001917135)* | *Other* | TNFα-producing Natural Killer T Cells | Adaptive Immune Cells | 1.21E-04 | 1.60E-05 | 1.34E-06 |
| *GGB3612 SGB4882 (COE1 sp001916965)* | *Other* | TNFα-producing CD161^+^ NK Cells | Natural Killer (NK) Cells | 8.12E-05 | 1.10E-05 | 1.69E-06 |
| *GGB4583 SGB6334 (Merdicola sp900555085)* | *Merdicola* | IFNγ/TNFα-producing CD8^+^ T Cells | CD8^+^ T Cells | 1.77E-05 | 2.40E-06 | 1.85E-06 |
| *GGB3612 SGB4882 (COE1 sp001916965)* | *Other* | TNFα-producing NK Cells | Natural Killer (NK) Cells | 8.45E-05 | 1.20E-05 | 2.32E-06 |
| *GGB4571 SGB6317 (Merdicola sp001915925)* | *Merdicola* | IL-17A-producing Activated CD69^+^ CD4^+^ T Cells | CD4^+^ T Cells | 9.49E-05 | 1.30E-05 | 2.49E-06 |
| *GGB4583 SGB6334 (Merdicola sp900555085)* | *Merdicola* | IFNγ/TNFα-producing CD161^+^ CD4^+^ T Cells | CD4^+^ T Cells | 2.35E-05 | 3.20E-06 | 2.72E-06 |
| *GGB3612 SGB4882 (COE1 sp001916965)* | *Other* | CCR7^+^ Activated NK Cells | Natural Killer (NK) Cells | 3.33E-04 | 4.70E-05 | 3.13E-06 |
| *Prevotella marseillensis* | *Prevotella* | IL-17A-producing CD161^+^ CD8^+^ T Cells | CD8^+^ T Cells | 2.18E-03 | 3.00E-04 | 3.22E-06 |
| *GGB4571 SGB6317 (Merdicola sp001915925)* | *Merdicola* | Mature LDNs | Innate Immune Cells | 1.49E-04 | 2.20E-05 | 3.38E-06 |
| *GGB4571 SGB6317 (Merdicola sp001915925)* | *Merdicola* | IL-17A-producing Activated CD69^+^ CD8^+^ T Cells | CD8^+^ T Cells | 2.81E-04 | 4.10E-05 | 3.41E-06 |
| *GGB9063 SGB13982 (Scatosoma sp003150575)* | *Scatosoma* | TNFα-producing Natural Killer T Cells | Adaptive Immune Cells | 2.04E-05 | 2.90E-06 | 3.58E-06 |
| *Phocaeicola plebeius* | *Phocaeicola* | CD8^+^ TEMRA | CD8^+^ T Cells | 2.29E-05 | 3.20E-06 | 3.68E-06 |
| *GGB4583 SGB6334 (Merdicola sp900555085)* | *Merdicola* | IFNγ/TNFα-producing CD69^+^ CD8^+^ T Cells | CD8^+^ T Cells | 4.53E-05 | 6.20E-06 | 4.35E-06 |
| *GGB3612 SGB4882 (COE1 sp001916965)* | *Other* | IL-17A-producing CD4^+^ T Cells | CD4^+^ T Cells | 5.79E-05 | 8.50E-06 | 4.54E-06 |
| *Streptococcus thermophilus* | *Streptococcus* | TNFα-producing Natural Killer T Cells | Adaptive Immune Cells | 8.42E-06 | 1.20E-06 | 5.27E-06 |
| *Phocaeicola plebeius* | *Phocaeicola* | TNFα-producing Natural Killer T Cells | Adaptive Immune Cells | 1.06E-04 | 1.60E-05 | 5.70E-06 |
| *Eubacterium ventriosum* | *Eubacterium* | CD8^+^ CM | CD8^+^ T Cells | 1.32E-05 | 1.80E-06 | 6.54E-06 |
| *Lachnospiraceae bacterium* | *Lachnospiraceae* | PMDSCs | Innate Immune Cells | 3.59E-05 | 5.40E-06 | 7.32E-06 |
| *Clostridium sp 1001270J 160509 D11* | *Clostridium* | IL-17A-producing CD161^+^ CD8^+^ T Cells | CD8^+^ T Cells | 2.64E-04 | 3.80E-05 | 7.94E-06 |
| *GGB4583 SGB6334 (Merdicola sp900555085)* | *Merdicola* | IFNγ-producing Activated CD69^+^ CD8^+^ T Cells | CD8^+^ T Cells | 4.66E-06 | 6.70E-07 | 1.11E-05 |
| *GGB4583 SGB6334 (Merdicola sp900555085)* | *Merdicola* | TNFα-producing Activated CD69^+^ CD8^+^ T Cells | CD8^+^ T Cells | 1.26E-05 | 1.80E-06 | 1.22E-05 |
| *Prevotella marseillensis* | *Prevotella* | IL-17A-producing CD8^+^ T Cells | CD8^+^ T Cells | 5.73E-04 | 8.30E-05 | 1.34E-05 |
| *Clostridium sp 1001270J 160509 D11* | *Clostridium* | IL-17A-producing Activated NK Cells | Natural Killer (NK) Cells | 4.13E-04 | 6.00E-05 | 1.43E-05 |
| *GGB4583 SGB6334 (Merdicola sp900555085)* | *Merdicola* | TNFα-producing CD8^+^ T Cells | CD8^+^ T Cells | 1.57E-05 | 2.30E-06 | 1.43E-05 |
| *GGB4583 SGB6334 (Merdicola sp900555085)* | *Merdicola* | IFNγ-producing CD8^+^ T Cells | CD8^+^ T Cells | 4.39E-06 | 6.40E-07 | 1.56E-05 |
| *GGB3612 SGB4882 (COE1 sp001916965)* | *Other* | IL-17A-producing Activated CD69^+^ CD4^+^ T Cells | CD4^+^ T Cells | 5.92E-05 | 9.30E-06 | 2.10E-05 |
| *GGB4583 SGB6334 (Merdicola sp900555085)* | *Merdicola* | TNFα-producing Activated HLA-DR^+^ CD8^+^ T Cells | CD8^+^ T Cells | 4.78E-05 | 7.10E-06 | 2.49E-05 |
| *GGB4571 SGB6317 (Merdicola sp001915925)* | *Merdicola* | Non-classical Monocytes | Innate Immune Cells | 5.85E-04 | 9.00E-05 | 2.82E-05 |
| *GGB4583 SGB6334 (Merdicola sp900555085)* | *Merdicola* | IFNγ/TNFα-producing HLA-DR^+^ CD8^+^ T Cells | CD8^+^ T Cells | 1.50E-04 | 2.30E-05 | 3.00E-05 |
| *GGB4571 SGB6317 (Merdicola sp001915925)* | *Merdicola* | CCR7^+^ Activated NK Cells | Natural Killer (NK) Cells | 4.78E-04 | 7.70E-05 | 3.31E-05 |
| *GGB4583 SGB6334 (Merdicola sp900555085)* | *Merdicola* | IFNγ-producing Activated HLA-DR^+^ CD4^+^ T Cells | CD4^+^ T Cells | 4.97E-05 | 7.50E-06 | 3.51E-05 |
| *Clostridium sp 1001270J 160509 D11* | *Clostridium* | IL-17A-producing CD8^+^ T Cells | CD8^+^ T Cells | 6.86E-05 | 1.10E-05 | 4.03E-05 |
| *Lachnospiraceae bacterium* | *Lachnospiraceae* | PD-1^+^ CD4^+^ T Cells | CD4^+^ T Cells | 1.41E-05 | 2.30E-06 | 4.96E-05 |
| *Sutterella sp AM11 39* | *Sutterella* | PMDSCs | Innate Immune Cells | 1.39E-05 | 2.40E-06 | 9.11E-05 |
| *GGB4583 SGB6334 (Merdicola sp900555085)* | *Merdicola* | IFNγ-producing Activated HLA-DR^+^ CD8^+^ T Cells | CD8^+^ T Cells | 1.67E-05 | 2.60E-06 | 9.20E-05 |
| *Sutterella sp AM11 39* | *Sutterella* | PD-1^+^ CD4^+^ T Cells | CD4^+^ T Cells | 5.70E-06 | 9.80E-07 | 9.45E-05 |
| *Sutterella sp AM11 39* | *Sutterella* | PD-1^+^ Activated HLA-DR^+^ CD4^+^ T Cells | CD4^+^ T Cells | 2.62E-05 | 4.50E-06 | 9.86E-05 |
| *GGB4583 SGB6334 (Merdicola sp900555085)* | *Merdicola* | IFNγ/TNFα-producing CD69^+^ CD4^+^ T Cells | CD4^+^ T Cells | 9.38E-06 | 1.50E-06 | 1.19E-04 |
| *GGB3612 SGB4882 (COE1 sp001916965)* | *Other* | Non-classical Monocytes | Innate Immune Cells | 3.61E-04 | 6.30E-05 | 1.54E-04 |
| *Clostridium sp 1001270J 160509 D11* | *Clostridium* | Non-classical Monocytes | Innate Immune Cells | 1.49E-04 | 2.60E-05 | 1.54E-04 |
| *Eubacterium ventriosum* | *Eubacterium* | Non-classical Monocytes | Innate Immune Cells | 1.67E-04 | 2.90E-05 | 1.54E-04 |
| *GGB3588 SGB4808 (Blautia A sp900551715)* | *Blautia* | PD-1^+^ Activated HLA-DR^+^ CD4^+^ T Cells | CD4^+^ T Cells | 7.24E-06 | 1.30E-06 | 1.54E-04 |
| *Lachnospiraceae bacterium* | *Lachnospiraceae* | PD-1^+^ Activated HLA-DR^+^ CD4^+^ T Cells | CD4^+^ T Cells | 6.23E-05 | 1.10E-05 | 1.54E-04 |
| *Streptococcus thermophilus* | *Streptococcus* | IFNγ-producing CD161^+^ CD4^+^ T Cells | CD4^+^ T Cells | 3.89E-06 | 6.50E-07 | 1.70E-04 |
| *Streptococcus thermophilus* | *Streptococcus* | IFNγ-producing CD4^+^ EM T Cells | CD4^+^ T Cells | 4.00E-06 | 6.90E-07 | 1.71E-04 |
| *GGB9063 SGB13982 (Scatosoma sp003150575)* | *Scatosoma* | IFNγ-producing CD4^+^ EM T Cells | CD4^+^ T Cells | 9.58E-06 | 1.60E-06 | 1.71E-04 |
| *Lachnospiraceae bacterium* | *Lachnospiraceae* | TNFα-producing CD161^+^ NK Cells | Natural Killer (NK) Cells | 6.23E-05 | 1.10E-05 | 1.78E-04 |
| *Streptococcus thermophilus* | *Streptococcus* | IFNγ/TNFα-producing CD161^+^ CD8^+^ T Cells | CD8^+^ T Cells | 3.12E-05 | 5.30E-06 | 1.95E-04 |
| *Streptococcus thermophilus* | *Streptococcus* | IFNγ-producing Activated CD69^+^ CD4^+^ T Cells | CD4^+^ T Cells | 1.74E-06 | 3.00E-07 | 2.28E-04 |
| *Phocaeicola plebeius* | *Phocaeicola* | IFNγ-producing Activated CD69^+^ CD4^+^ T Cells | CD4^+^ T Cells | 2.16E-05 | 3.90E-06 | 2.62E-04 |
| *GGB9063 SGB13982 (Scatosoma sp003150575)* | *Scatosoma* | IFNγ-producing Activated CD69^+^ CD4^+^ T Cells | CD4^+^ T Cells | 4.07E-06 | 7.20E-07 | 2.62E-04 |
| *Streptococcus thermophilus* | *Streptococcus* | CD8^+^ TEMRA | CD8^+^ T Cells | 1.61E-06 | 2.80E-07 | 2.68E-04 |
| *Holdemanella biformis* | *Holdemanella* | PD-1^+^ CD4^+^ T Cells | CD4^+^ T Cells | 2.43E-05 | 4.50E-06 | 2.71E-04 |
| *Phocaeicola plebeius* | *Phocaeicola* | IFNγ-producing CD4^+^ EM T Cells | CD4^+^ T Cells | 4.96E-05 | 8.90E-06 | 2.77E-04 |
| *Holdemanella biformis* | *Holdemanella* | PMDSCs | Innate Immune Cells | 5.88E-05 | 1.10E-05 | 2.99E-04 |
| *Holdemanella biformis* | *Holdemanella* | PD-1^+^ Activated HLA-DR^+^ CD4^+^ T Cells | CD4^+^ T Cells | 1.10E-04 | 2.10E-05 | 3.07E-04 |
| *GGB9063 SGB13982 (Scatosoma sp003150575)* | *Scatosoma* | IFNγ-producing CD161^+^ CD4^+^ T Cells | CD4^+^ T Cells | 9.00E-06 | 1.60E-06 | 3.09E-04 |
| *Lachnospiraceae bacterium* | *Lachnospiraceae* | TNFα-producing NK Cells | Natural Killer (NK) Cells | 6.41E-05 | 1.20E-05 | 3.26E-04 |
| *GGB9063 SGB13982 (Scatosoma sp003150575)* | *Scatosoma* | CD4^+^ TEMRA | CD4^+^ T Cells | 1.32E-05 | 2.40E-06 | 3.43E-04 |
| *Lachnospiraceae bacterium* | *Lachnospiraceae* | PD-1^+^ CD8^+^ T Cells | CD8^+^ T Cells | 2.93E-05 | 5.40E-06 | 3.77E-04 |
| *Lachnospiraceae bacterium* | *Lachnospiraceae* | CCR7^+^ Activated NK Cells | Natural Killer (NK) Cells | 2.49E-04 | 4.70E-05 | 4.61E-04 |
| *Sutterella sp AM11 39* | *Sutterella* | PD-1^+^ CD8^+^ T Cells | CD8^+^ T Cells | 1.19E-05 | 2.30E-06 | 5.34E-04 |
| *Lachnospiraceae bacterium* | *Lachnospiraceae* | IL-17A-producing CD4^+^ T Cells | CD4^+^ T Cells | 4.38E-05 | 8.20E-06 | 5.42E-04 |
| *Streptococcus thermophilus* | *Streptococcus* | IFNγ/TNFα-producing CD69^+^ CD8^+^ T Cells | CD8^+^ T Cells | 1.25E-05 | 2.20E-06 | 5.62E-04 |
| *GGB9063 SGB13982 (Scatosoma sp003150575)* | *Scatosoma* | CD8^+^ TEMRA | CD8^+^ T Cells | 3.72E-06 | 6.90E-07 | 5.63E-04 |
| *GGB4583 SGB6334 (Merdicola sp900555085)* | *Merdicola* | IFNγ-producing CD161^+^ CD8^+^ T Cells | CD8^+^ T Cells | 2.48E-05 | 4.30E-06 | 5.95E-04 |
| *Phocaeicola plebeius* | *Phocaeicola* | TNFα-producing Activated HLA-DR^+^ CD8^+^ T Cells | CD8^+^ T Cells | 1.72E-04 | 3.10E-05 | 6.97E-04 |
| *Streptococcus thermophilus* | *Streptococcus* | IFNγ/TNFα-producing CD8^+^ T Cells | CD8^+^ T Cells | 4.75E-06 | 8.60E-07 | 7.34E-04 |
| *Holdemanella biformis* | *Holdemanella* | PD-1^+^ CD8^+^ T Cells | CD8^+^ T Cells | 5.16E-05 | 1.00E-05 | 7.58E-04 |
| *Phocaeicola plebeius* | *Phocaeicola* | IFNγ-producing CD161^+^ CD4^+^ T Cells | CD4^+^ T Cells | 4.60E-05 | 8.70E-06 | 7.92E-04 |
| *GGB3588 SGB4808 (Blautia A sp900551715)* | *Blautia* | PD-1^+^ CD4^+^ T Cells | CD4^+^ T Cells | 1.49E-06 | 2.90E-07 | 8.02E-04 |
| *Clostridiaceae unclassified SGB15090 (CAG-83 sp000435975)* | *Other* | PD-1^+^ CD4^+^ T Cells | CD4^+^ T Cells | 9.20E-06 | 1.90E-06 | 8.74E-04 |
| *GGB9512 SGB14909 (Negativibacillus sp000435195)* | *Negativibacillus* | PD-1^+^ CD4^+^ T Cells | CD4^+^ T Cells | 4.70E-06 | 9.50E-07 | 8.74E-04 |
| *GGB9699 SGB15216 (Faecousia sp003525905)* | *Faecousia* | IFNγ-producing Activated CD69^+^ CD8^+^ T Cells | CD8^+^ T Cells | 3.38E-06 | 6.20E-07 | 8.83E-04 |
| *Clostridium sp 1001270J 160509 D11* | *Clostridium* | IL-17A-producing CD4^+^ EM T Cells | CD4^+^ T Cells | 7.46E-05 | 1.30E-05 | 8.89E-04 |
| *Phocaeicola plebeius* | *Phocaeicola* | IFNγ-producing Activated CD69^+^ CD8^+^ T Cells | CD8^+^ T Cells | 1.61E-05 | 3.00E-06 | 9.04E-04 |
| *Streptococcus thermophilus* | *Streptococcus* | IFNγ/TNFα-producing CD161^+^ CD4^+^ T Cells | CD4^+^ T Cells | 6.32E-06 | 1.20E-06 | 9.16E-04 |
| *GGB9699 SGB15216 (Faecousia sp003525905)* | *Faecousia* | IFNγ-producing CD8^+^ T Cells | CD8^+^ T Cells | 3.19E-06 | 5.90E-07 | 9.95E-04 |
| *Phocaeicola plebeius* | *Phocaeicola* | IFNγ-producing CD8^+^ T Cells | CD8^+^ T Cells | 1.52E-05 | 2.90E-06 | 0.00103 |
| *GGB9512 SGB14909 (Negativibacillus sp000435195)* | *Negativibacillus* | PMDSCs | Innate Immune Cells | 1.14E-05 | 2.30E-06 | 0.00103 |
| *GGB4583 SGB6334 (Merdicola sp900555085)* | *Merdicola* | IFNγ-producing CD4^+^ T Cells | CD4^+^ T Cells | 3.89E-06 | 7.00E-07 | 0.00111 |
| *Phocaeicola plebeius* | *Phocaeicola* | IFNγ/TNFα-producing CD8^+^ T Cells | CD8^+^ T Cells | 5.86E-05 | 1.10E-05 | 0.00114 |
| *GGB9063 SGB13982 (Scatosoma sp003150575)* | *Scatosoma* | IFNγ/TNFα-producing CD161^+^ CD4^+^ T Cells | CD4^+^ T Cells | 1.47E-05 | 2.80E-06 | 0.00119 |
| *GGB28474 SGB41058 (CAJMQN01 sp905215695)* | *Other* | PD-1^+^ Activated HLA-DR^+^ CD4^+^ T Cells | CD4^+^ T Cells | 6.81E-06 | 1.40E-06 | 0.00120 |
| *Phocaeicola plebeius* | *Phocaeicola* | CD4^+^ TEMRA | CD4^+^ T Cells | 6.65E-05 | 1.30E-05 | 0.00120 |
| *Streptococcus thermophilus* | *Streptococcus* | CD4^+^ TEMRA | CD4^+^ T Cells | 5.21E-06 | 1.00E-06 | 0.00120 |
| *GGB3612 SGB4882 (COE1 sp001916965)* | *Other* | IL-17A-producing CD161^+^ CD8^+^ T Cells | CD8^+^ T Cells | 5.43E-04 | 1.10E-04 | 0.00127 |
| *Ruminococcus bovis* | *Ruminococcus* | IL-17A-producing CD161^+^ CD8^+^ T Cells | CD8^+^ T Cells | 1.00E-03 | 2.00E-04 | 0.00127 |
| *Phocaeicola plebeius* | *Phocaeicola* | TNFα-producing Activated CD69^+^ CD8^+^ T Cells | CD8^+^ T Cells | 4.37E-05 | 8.20E-06 | 0.00134 |
| *GGB9699 SGB15216 (Faecousia sp003525905)* | *Faecousia* | IFNγ-producing Natural Killer T Cells | Adaptive Immune Cells | 7.21E-06 | 1.40E-06 | 0.00136 |
| *Phocaeicola plebeius* | *Phocaeicola* | TNFα-producing CD8^+^ T Cells | CD8^+^ T Cells | 5.45E-05 | 1.00E-05 | 0.00140 |
| *Prevotella marseillensis* | *Prevotella* | PD-1^+^ CD8^+^ T Cells | CD8^+^ T Cells | 1.06E-04 | 2.20E-05 | 0.00140 |
| *GGB3281 SGB4335 (Eubacterium R sp003526845)* | *Eubacterium* | PD-1^+^ CD8^+^ T Cells | CD8^+^ T Cells | 1.46E-05 | 3.00E-06 | 0.00140 |
| *Streptococcus thermophilus* | *Streptococcus* | IFNγ-producing Activated CD69^+^ CD8^+^ T Cells | CD8^+^ T Cells | 1.24E-06 | 2.40E-07 | 0.00146 |
| *Clostridiaceae unclassified SGB15090 (CAG-83 sp000435975)* | *Other* | PD-1^+^ Activated HLA-DR^+^ CD4^+^ T Cells | CD4^+^ T Cells | 4.12E-05 | 8.60E-06 | 0.00154 |
| *GGB3588 SGB4808 (Blautia A sp900551715)* | *Blautia* | PMDSCs | Innate Immune Cells | 3.50E-06 | 7.30E-07 | 0.00159 |
| *Clostridiaceae unclassified SGB15090 (CAG-83 sp000435975)* | *Other* | PMDSCs | Innate Immune Cells | 2.18E-05 | 4.60E-06 | 0.00161 |
| *Streptococcus thermophilus* | *Streptococcus* | TNFα-producing Activated CD69^+^ CD8^+^ T Cells | CD8^+^ T Cells | 3.37E-06 | 6.60E-07 | 0.00161 |
| *GGB6601 SGB9333 (CAG-267 sp001917135)* | *Other* | IFNγ-producing Natural Killer T Cells | Adaptive Immune Cells | 3.74E-05 | 7.60E-06 | 0.00166 |
| *Phocaeicola plebeius* | *Phocaeicola* | IFNγ/TNFα-producing CD161^+^ CD8^+^ T Cells | CD8^+^ T Cells | 3.65E-04 | 7.10E-05 | 0.00168 |
| *Lachnospiraceae bacterium* | *Lachnospiraceae* | IL-17A-producing Activated CD69^+^ CD8^+^ T Cells | CD8^+^ T Cells | 1.32E-04 | 2.70E-05 | 0.00170 |
| *Holdemanella biformis* | *Holdemanella* | IL-17A-producing Activated CD69^+^ CD8^+^ T Cells | CD8^+^ T Cells | 2.41E-04 | 5.00E-05 | 0.00170 |
| *GGB9512 SGB14909 (Negativibacillus sp000435195)* | *Negativibacillus* | PD-1^+^ Activated HLA-DR^+^ CD4^+^ T Cells | CD4^+^ T Cells | 2.08E-05 | 4.40E-06 | 0.00172 |
| *Streptococcus thermophilus* | *Streptococcus* | TNFα-producing CD8^+^ T Cells | CD8^+^ T Cells | 4.20E-06 | 8.20E-07 | 0.00175 |
| *GGB3281 SGB4335 (Eubacterium R sp003526845)* | *Eubacterium* | PMDSCs | Innate Immune Cells | 1.58E-05 | 3.40E-06 | 0.00177 |
| *Streptococcus thermophilus* | *Streptococcus* | IFNγ-producing CD8^+^ T Cells | CD8^+^ T Cells | 1.16E-06 | 2.30E-07 | 0.00179 |
| *GGB9063 SGB13982 (Scatosoma sp003150575)* | *Scatosoma* | IFNγ/TNFα-producing CD161^+^ CD8^+^ T Cells | CD8^+^ T Cells | 6.76E-05 | 1.40E-05 | 0.00183 |
| *Sutterella sp AM11 39* | *Sutterella* | TNFα-producing NK Cells | Natural Killer (NK) Cells | 2.46E-05 | 5.10E-06 | 0.00189 |
| *Sutterella sp AM11 39* | *Sutterella* | IL-17A-producing Activated CD69^+^ CD8^+^ T Cells | CD8^+^ T Cells | 5.39E-05 | 1.10E-05 | 0.00193 |
| *Prevotella marseillensis* | *Prevotella* | PMDSCs | Innate Immune Cells | 1.13E-04 | 2.40E-05 | 0.00194 |
| *GGB3281 SGB4335 (Eubacterium R sp003526845)* | *Eubacterium* | PD-1^+^ CD4^+^ T Cells | CD4^+^ T Cells | 6.45E-06 | 1.40E-06 | 0.00215 |
| *GGB9699 SGB15216 (Faecousia sp003525905)* | *Faecousia* | TNFα-producing Natural Killer T Cells | Adaptive Immune Cells | 1.82E-05 | 3.80E-06 | 0.00230 |
| *Phocaeicola plebeius* | *Phocaeicola* | IFNγ-producing Activated HLA-DR^+^ CD8^+^ T Cells | CD8^+^ T Cells | 5.93E-05 | 1.20E-05 | 0.00242 |
| *GGB39037 SGB4393 (UBA737 sp002431945)* | *Other* | γδ T Cells | Adaptive Immune Cells | 2.34E-06 | 4.70E-07 | 0.00250 |
| *GGB9172 SGB14110 (Fimimonas sp003537755)* | *Fimimonas* | γδ T Cells | Adaptive Immune Cells | 1.16E-05 | 2.40E-06 | 0.00250 |
| *GGB3303 SGB4364 (CAG-177 sp000431775)* | *Other* | γδ T Cells | Adaptive Immune Cells | 4.82E-06 | 9.20E-07 | 0.00250 |
| *GGB9618 SGB15064 (Dysosmobacter sp900763685)* | *Dysosmobacter* | γδ T Cells | Adaptive Immune Cells | 2.75E-06 | 5.60E-07 | 0.00250 |
| *GGB4571 SGB6317 (Merdicola sp001915925)* | *Merdicola* | IL-17A-producing Activated NK Cells | Natural Killer (NK) Cells | 1.28E-03 | 2.50E-04 | 0.00254 |
| *Ruminococcus bovis* | *Ruminococcus* | IL-17A-producing Activated NK Cells | Natural Killer (NK) Cells | 1.54E-03 | 3.10E-04 | 0.00254 |
| *GGB9512 SGB14909 (Negativibacillus sp000435195)* | *Negativibacillus* | PD-1^+^ CD8^+^ T Cells | CD8^+^ T Cells | 9.87E-06 | 2.10E-06 | 0.00255 |
| *Lachnospiraceae bacterium* | *Lachnospiraceae* | IL-17A-producing Activated CD69^+^ CD4^+^ T Cells | CD4^+^ T Cells | 4.36E-05 | 9.00E-06 | 0.00262 |
| *Prevotella marseillensis* | *Prevotella* | PD-1^+^ CD4^+^ T Cells | CD4^+^ T Cells | 4.59E-05 | 1.00E-05 | 0.00275 |
| *GGB9088 SGB14017 (UMGS1795 sp900555125)* | *Other* | IL-2-producing Activated HLA-DR^+^ CD8^+^ T Cells | CD8^+^ T Cells | 1.70E-04 | 3.20E-05 | 0.00279 |
| *GGB28474 SGB41058 (CAJMQN01 sp905215695)* | *Other* | PD-1^+^ CD4^+^ T Cells | CD4^+^ T Cells | 1.41E-06 | 3.10E-07 | 0.00291 |
| *Streptococcus thermophilus* | *Streptococcus* | TNFα-producing Activated HLA-DR^+^ CD8^+^ T Cells | CD8^+^ T Cells | 1.27E-05 | 2.60E-06 | 0.00295 |
| *GGB9762 SGB15377 (SFMI01 sp004556155)* | *Other* | γδ T Cells | Adaptive Immune Cells | 4.06E-06 | 8.60E-07 | 0.00310 |
| *Lachnospiraceae bacterium OM04 12BH* | *Lachnospiraceae* | γδ T Cells | Adaptive Immune Cells | 1.34E-05 | 2.80E-06 | 0.00310 |
| *Phocaeicola plebeius* | *Phocaeicola* | IFNγ/TNFα-producing CD69^+^ CD8^+^ T Cells | CD8^+^ T Cells | 1.47E-04 | 3.00E-05 | 0.00311 |
| *Streptococcus thermophilus* | *Streptococcus* | IFNγ/TNFα-producing HLA-DR^+^ CD8^+^ T Cells | CD8^+^ T Cells | 4.05E-05 | 8.00E-06 | 0.00312 |
| *Clostridium sp 1001270J 160509 D11* | *Clostridium* | IL-17A-producing CD161^+^ CD4^+^ T Cells | CD4^+^ T Cells | 5.85E-05 | 1.20E-05 | 0.00312 |
| *Sutterella sp AM11 39* | *Sutterella* | Mature LDNs | Innate Immune Cells | 2.83E-05 | 6.00E-06 | 0.00333 |
| *Sutterella sp AM11 39* | *Sutterella* | CCR7^+^ Activated NK Cells | Natural Killer (NK) Cells | 9.46E-05 | 2.00E-05 | 0.00354 |
| *Clostridium SGB6179 (Clostridium sp900540255)* | *Clostridium* | PD-1^+^ Activated HLA-DR^+^ CD4^+^ T Cells | CD4^+^ T Cells | 1.54E-05 | 3.50E-06 | 0.00355 |
| *GGB3281 SGB4335 (Eubacterium R sp003526845)* | *Eubacterium* | PD-1^+^ Activated HLA-DR^+^ CD4^+^ T Cells | CD4^+^ T Cells | 2.87E-05 | 6.50E-06 | 0.00355 |
| *Adlercreutzia equolifaciens* | *Adlercreutzia* | CD8^+^ CM | CD8^+^ T Cells | 1.07E-05 | 2.10E-06 | 0.00363 |
| *GGB3475 SGB4638 (V9D3004 sp900760345)* | *Other* | CD8^+^ Naive | CD8^+^ T Cells | 2.23E-06 | 4.30E-07 | 0.00388 |
| *GGB27106 SGB6188 (Clostridium sp900539375)* | *Clostridium* | γδ T Cells | Adaptive Immune Cells | 1.83E-06 | 4.00E-07 | 0.00404 |
| *Holdemanella biformis* | *Holdemanella* | Mature LDNs | Innate Immune Cells | 1.23E-04 | 2.70E-05 | 0.00404 |
| *GGB9699 SGB15216 (Faecousia sp003525905)* | *Faecousia* | Natural Killer T Cells | Adaptive Immune Cells | 4.71E-06 | 1.00E-06 | 0.00408 |
| *Prevotella marseillensis* | *Prevotella* | PD-1^+^ Activated HLA-DR^+^ CD4^+^ T Cells | CD4^+^ T Cells | 2.05E-04 | 4.70E-05 | 0.00414 |
| *Lachnospira sp NSJ 43* | *Lachnospiraceae* | IFNγ-producing Natural Killer T Cells | Adaptive Immune Cells | 2.44E-05 | 5.30E-06 | 0.00422 |
| *GGB3303 SGB4364 (CAG-177 sp000431775)* | *Other* | IL-2-producing CD4^+^ EM T Cells | CD4^+^ T Cells | 2.20E-05 | 4.30E-06 | 0.00449 |
| *GGB9618 SGB15064 (Dysosmobacter sp900763685)* | *Dysosmobacter* | IL-2-producing CD4^+^ EM T Cells | CD4^+^ T Cells | 1.27E-05 | 2.60E-06 | 0.00449 |
| *GGB3588 SGB4808 (Blautia A sp900551715)* | *Blautia* | IL-17A-producing CD4^+^ T Cells | CD4^+^ T Cells | 4.66E-06 | 1.00E-06 | 0.00457 |
| *GGB9699 SGB15216 (Faecousia sp003525905)* | *Faecousia* | CD4^+^ TEMRA | CD4^+^ T Cells | 1.28E-05 | 2.80E-06 | 0.00458 |
| *GGB4571 SGB6317 (Merdicola sp001915925)* | *Merdicola* | IL-17A-producing CD161^+^ CD4^+^ T Cells | CD4^+^ T Cells | 2.07E-04 | 4.30E-05 | 0.00476 |
| *Phocaeicola plebeius* | *Phocaeicola* | IFNγ/TNFα-producing CD161^+^ CD4^+^ T Cells | CD4^+^ T Cells | 7.33E-05 | 1.60E-05 | 0.00488 |
| *Sutterella sp AM11 39* | *Sutterella* | TNFα-producing CD161^+^ NK Cells | Natural Killer (NK) Cells | 2.25E-05 | 4.90E-06 | 0.00511 |
| *GGB9059 SGB13976 (CAG-349 sp003539515)* | *Other* | γδ T Cells | Adaptive Immune Cells | 3.38E-05 | 7.60E-06 | 0.00551 |
| *Clostridiaceae unclassified SGB15090 (CAG-83 sp000435975)* | *Other* | PD-1^+^ CD8^+^ T Cells | CD8^+^ T Cells | 1.86E-05 | 4.30E-06 | 0.00572 |
| *GGB6601 SGB9333 (CAG-267 sp001917135)* | *Other* | Natural Killer T Cells | Adaptive Immune Cells | 2.42E-05 | 5.40E-06 | 0.00574 |
| *GGB3612 SGB4882 (COE1 sp001916965)* | *Other* | IL-17A-producing Activated NK Cells | Natural Killer (NK) Cells | 7.92E-04 | 1.70E-04 | 0.00595 |
| *GGB3588 SGB4808 (Blautia A sp900551715)* | *Blautia* | IL-17A-producing Activated CD69^+^ CD4^+^ T Cells | CD4^+^ T Cells | 4.87E-06 | 1.10E-06 | 0.00605 |
| *Ruminococcus sp AF13 28* | *Ruminococcus* | γδ T Cells | Adaptive Immune Cells | 2.27E-06 | 5.20E-07 | 0.00627 |
| *GGB39037 SGB4393 (UBA737 sp002431945)* | *Other* | IL-2-producing CD4^+^ EM T Cells | CD4^+^ T Cells | 1.04E-05 | 2.20E-06 | 0.00629 |
| *Phocaeicola faecalis* | *Phocaeicola* | CCR7^+^ CD8^+^ T Cells | CD8^+^ T Cells | 8.38E-06 | 1.80E-06 | 0.00650 |
| *GGB3475 SGB4638 (V9D3004 sp900760345)* | *Other* | CCR7^+^ CD8^+^ T Cells | CD8^+^ T Cells | 1.59E-06 | 3.40E-07 | 0.00650 |
| *GGB9724 SGB15278 (Fournierella excrementavium)* | *Other* | CCR7^+^ CD8^+^ T Cells | CD8^+^ T Cells | 4.42E-06 | 9.60E-07 | 0.00650 |
| *Vescimonas coprocola* | *Vescimonas* | CCR7^+^ CD8^+^ T Cells | CD8^+^ T Cells | 2.25E-06 | 4.80E-07 | 0.00650 |
| *GGB9260 SGB14208 (CAG-448 sp000433415)* | *Other* | IL-17A-producing CD161^+^ NK Cells | Natural Killer (NK) Cells | 5.56E-05 | 1.10E-05 | 0.00655 |
| *GGB3281 SGB4335 (Eubacterium R sp003526845)* | *Eubacterium* | TNFα-producing NK Cells | Natural Killer (NK) Cells | 2.98E-05 | 6.80E-06 | 0.00663 |
| *Holdemanella biformis* | *Holdemanella* | TNFα-producing NK Cells | Natural Killer (NK) Cells | 1.02E-04 | 2.30E-05 | 0.00663 |
| *Phocaeicola plebeius* | *Phocaeicola* | IFNγ/TNFα-producing HLA-DR^+^ CD8^+^ T Cells | CD8^+^ T Cells | 4.90E-04 | 1.00E-04 | 0.00675 |
| *GGB3281 SGB4335 (Eubacterium R sp003526845)* | *Eubacterium* | Mature LDNs | Innate Immune Cells | 3.49E-05 | 8.00E-06 | 0.00689 |
| *GGB28474 SGB41058 (CAJMQN01 sp905215695)* | *Other* | PMDSCs | Innate Immune Cells | 3.27E-06 | 7.80E-07 | 0.00691 |
| *GGB3612 SGB4882 (COE1 sp001916965)* | *Other* | IL-17A-producing CD8^+^ T Cells | CD8^+^ T Cells | 1.38E-04 | 2.90E-05 | 0.00695 |
| *Streptococcus thermophilus* | *Streptococcus* | IFNγ-producing Activated HLA-DR^+^ CD4^+^ T Cells | CD4^+^ T Cells | 1.30E-05 | 2.70E-06 | 0.00725 |
| *Streptococcus thermophilus* | *Streptococcus* | IFNγ/TNFα-producing CD69^+^ CD4^+^ T Cells | CD4^+^ T Cells | 2.49E-06 | 5.30E-07 | 0.00758 |
| *GGB9063 SGB13982 (Scatosoma sp003150575)* | *Scatosoma* | IFNγ/TNFα-producing CD69^+^ CD4^+^ T Cells | CD4^+^ T Cells | 5.91E-06 | 1.30E-06 | 0.00758 |
| *Phocaeicola plebeius* | *Phocaeicola* | IFNγ-producing Activated HLA-DR^+^ CD4^+^ T Cells | CD4^+^ T Cells | 1.62E-04 | 3.50E-05 | 0.00783 |
| *GGB9063 SGB13982 (Scatosoma sp003150575)* | *Scatosoma* | IFNγ/TNFα-producing CD8^+^ T Cells | CD8^+^ T Cells | 9.93E-06 | 2.20E-06 | 0.00836 |
| *GGB9699 SGB15216 (Faecousia sp003525905)* | *Faecousia* | IFNγ/TNFα-producing CD8^+^ T Cells | CD8^+^ T Cells | 1.10E-05 | 2.40E-06 | 0.00836 |
| *GGB9512 SGB14909 (Negativibacillus sp000435195)* | *Negativibacillus* | Mature LDNs | Innate Immune Cells | 2.39E-05 | 5.60E-06 | 0.00845 |
| *GGB3588 SGB4808 (Blautia A sp900551715)* | *Blautia* | TNFα-producing NK Cells | Natural Killer (NK) Cells | 6.38E-06 | 1.50E-06 | 0.00859 |
| *Lachnospira sp NSJ 43* | *Lachnospiraceae* | Natural Killer T Cells | Adaptive Immune Cells | 1.61E-05 | 3.70E-06 | 0.00863 |
| *Streptococcus thermophilus* | *Streptococcus* | IFNγ-producing Activated HLA-DR^+^ CD8^+^ T Cells | CD8^+^ T Cells | 4.36E-06 | 9.50E-07 | 0.00880 |
| *GGB3281 SGB4335 (Eubacterium R sp003526845)* | *Eubacterium* | CCR7^+^ Activated NK Cells | Natural Killer (NK) Cells | 1.16E-04 | 2.70E-05 | 0.00891 |
| *Holdemanella biformis* | *Holdemanella* | IL-17A-producing CD4^+^ T Cells | CD4^+^ T Cells | 6.99E-05 | 1.60E-05 | 0.00958 |
| *GGB6601 SGB9333 (CAG-267 sp001917135)* | *Other* | IL-17A-producing CD4^+^ T Cells | CD4^+^ T Cells | 1.52E-04 | 3.60E-05 | 0.00976 |
| *Lachnospira eligens* | *Lachnospiraceae* | IFNγ-producing Natural Killer T Cells | Adaptive Immune Cells | 2.67E-05 | 6.40E-06 | 0.01058 |
| *Holdemanella biformis* | *Holdemanella* | TNFα-producing CD161^+^ NK Cells | Natural Killer (NK) Cells | 9.55E-05 | 2.20E-05 | 0.01078 |
| *GGB9172 SGB14110 (Fimimonas sp003537755)* | *Fimimonas* | IL-2-producing CD4^+^ EM T Cells | CD4^+^ T Cells | 4.94E-05 | 1.10E-05 | 0.01084 |
| *Lachnospiraceae bacterium OM04 12BH* | *Lachnospiraceae* | IL-2-producing CD4^+^ EM T Cells | CD4^+^ T Cells | 5.89E-05 | 1.30E-05 | 0.01084 |
| *GGB9762 SGB15377 (SFMI01 sp004556155)* | *Other* | IL-2-producing CD4^+^ EM T Cells | CD4^+^ T Cells | 1.76E-05 | 4.10E-06 | 0.01109 |
| *GGB27106 SGB6188 (Clostridium sp900539375)* | *Clostridium* | IL-2-producing CD4^+^ EM T Cells | CD4^+^ T Cells | 8.02E-06 | 1.90E-06 | 0.01109 |
| *GGB3281 SGB4335 (Eubacterium R sp003526845)* | *Eubacterium* | TNFα-producing CD161^+^ NK Cells | Natural Killer (NK) Cells | 2.77E-05 | 6.60E-06 | 0.01115 |
| *Sutterella sp AM11 39* | *Sutterella* | IL-17A-producing CD4^+^ T Cells | CD4^+^ T Cells | 1.54E-05 | 3.70E-06 | 0.01130 |
| *GGB9063 SGB13982 (Scatosoma sp003150575)* | *Scatosoma* | IFNγ/TNFα-producing CD69^+^ CD8^+^ T Cells | CD8^+^ T Cells | 2.57E-05 | 5.80E-06 | 0.01173 |
| *GGB9063 SGB13982 (Scatosoma sp003150575)* | *Scatosoma* | IFNγ-producing Activated CD69^+^ CD8^+^ T Cells | CD8^+^ T Cells | 2.66E-06 | 6.10E-07 | 0.01212 |
| *GGB4583 SGB6334 (Merdicola sp900555085)* | *Merdicola* | IFNγ-producing CD8^+^ EM T Cells | CD8^+^ T Cells | 1.90E-05 | 3.90E-06 | 0.01214 |
| *Ruthenibacterium lactatiformans* | *Ruthenibacterium* | Classical Monocytes | Innate Immune Cells | 3.22E-06 | 6.70E-07 | 0.01339 |
| *GGB28474 SGB41058 (CAJMQN01 sp905215695)* | *Other* | IL-17A-producing Activated NK Cells | Natural Killer (NK) Cells | 7.61E-05 | 1.80E-05 | 0.01359 |
| *Clostridium SGB6179 (Clostridium sp900540255)* | *Clostridium* | IL-17A-producing Activated NK Cells | Natural Killer (NK) Cells | 1.86E-04 | 4.30E-05 | 0.01359 |
| *GGB3588 SGB4808 (Blautia A sp900551715)* | *Blautia* | IL-17A-producing Activated NK Cells | Natural Killer (NK) Cells | 7.42E-05 | 1.80E-05 | 0.01359 |
| *Lachnospiraceae bacterium* | *Lachnospiraceae* | IL-17A-producing Activated NK Cells | Natural Killer (NK) Cells | 6.43E-04 | 1.50E-04 | 0.01359 |
| *Clostridium SGB6179 (Clostridium sp900540255)* | *Clostridium* | PD-1^+^ CD4^+^ T Cells | CD4^+^ T Cells | 3.11E-06 | 7.80E-07 | 0.01361 |
| *Ruthenibacterium lactatiformans* | *Ruthenibacterium* | CCR7^+^ CD8^+^ T Cells | CD8^+^ T Cells | 1.57E-06 | 3.70E-07 | 0.01362 |
| *Eubacterium ventriosum* | *Eubacterium* | CCR7^+^ CD8^+^ T Cells | CD8^+^ T Cells | 3.24E-06 | 7.70E-07 | 0.01370 |
| *Lachnospira sp NSJ 43* | *Lachnospiraceae* | TNFα-producing Natural Killer T Cells | Adaptive Immune Cells | 5.96E-05 | 1.40E-05 | 0.01378 |
| *GGB9699 SGB15216 (Faecousia sp003525905)* | *Faecousia* | IFNγ-producing Activated CD69^+^ CD4^+^ T Cells | CD4^+^ T Cells | 3.77E-06 | 8.80E-07 | 0.01400 |
| *Holdemanella biformis* | *Holdemanella* | IL-17A-producing Activated CD69^+^ CD4^+^ T Cells | CD4^+^ T Cells | 7.19E-05 | 1.70E-05 | 0.01401 |
| *GGB6601 SGB9333 (CAG-267 sp001917135)* | *Other* | IL-17A-producing Activated CD69^+^ CD4^+^ T Cells | CD4^+^ T Cells | 1.59E-04 | 3.80E-05 | 0.01401 |
| *Phocaeicola plebeius* | *Phocaeicola* | IFNγ/TNFα-producing CD69^+^ CD4^+^ T Cells | CD4^+^ T Cells | 3.01E-05 | 6.90E-06 | 0.01414 |
| *GGB9063 SGB13982 (Scatosoma sp003150575)* | *Scatosoma* | IFNγ-producing CD8^+^ T Cells | CD8^+^ T Cells | 2.50E-06 | 5.80E-07 | 0.01422 |
| *GGB4583 SGB6334 (Merdicola sp900555085)* | *Merdicola* | TNFα-producing CD161^+^ CD8^+^ T Cells | CD8^+^ T Cells | 4.60E-05 | 9.90E-06 | 0.01459 |
| *Ruminococcus bovis* | *Ruminococcus* | TNFα-producing CD161^+^ CD8^+^ T Cells | CD8^+^ T Cells | 1.06E-04 | 2.30E-05 | 0.01459 |
| *GGB3588 SGB4808 (Blautia A sp900551715)* | *Blautia* | PD-1^+^ CD8^+^ T Cells | CD8^+^ T Cells | 2.79E-06 | 7.00E-07 | 0.01527 |
| *Phocaeicola plebeius* | *Phocaeicola* | Activated HLA-DR^+^ CD8^+^ T Cells | CD8^+^ T Cells | 2.95E-05 | 6.20E-06 | 0.01527 |
| *GGB4583 SGB6334 (Merdicola sp900555085)* | *Merdicola* | Activated HLA-DR^+^ CD8^+^ T Cells | CD8^+^ T Cells | 7.19E-06 | 1.60E-06 | 0.01527 |
| *Sutterella sp AM11 39* | *Sutterella* | IL-17A-producing Activated CD69^+^ CD4^+^ T Cells | CD4^+^ T Cells | 1.61E-05 | 3.90E-06 | 0.01531 |
| *Sutterella sp AM11 39* | *Sutterella* | IL-17A-producing Activated NK Cells | Natural Killer (NK) Cells | 2.59E-04 | 6.30E-05 | 0.01549 |
| *GGB9260 SGB14208 (CAG-448 sp000433415)* | *Other* | IL-17A-producing NK Cells | Natural Killer (NK) Cells | 4.65E-05 | 9.70E-06 | 0.01558 |
| *Roseburia lenta* | *Roseburia* | Natural Killer T Cells | Adaptive Immune Cells | 1.95E-06 | 4.80E-07 | 0.01614 |
| *Clostridiaceae unclassified SGB15090 (CAG-83 sp000435975)* | *Other* | TNFα-producing NK Cells | Natural Killer (NK) Cells | 3.82E-05 | 9.50E-06 | 0.01742 |
| *Phocaeicola faecalis* | *Phocaeicola* | CD8^+^ Naive | CD8^+^ T Cells | 1.06E-05 | 2.40E-06 | 0.01761 |
| *GGB9724 SGB15278 (Fournierella excrementavium)* | *Other* | CD8^+^ Naive | CD8^+^ T Cells | 5.61E-06 | 1.30E-06 | 0.01761 |
| *Clostridium SGB6179 (Clostridium sp900540255)* | *Clostridium* | PMDSCs | Innate Immune Cells | 7.42E-06 | 1.90E-06 | 0.01791 |
| *GGB9699 SGB15216 (Faecousia sp003525905)* | *Faecousia* | IFNγ-producing Activated HLA-DR^+^ CD8^+^ T Cells | CD8^+^ T Cells | 1.09E-05 | 2.50E-06 | 0.01816 |
| *Clostridium sp AM22 11AC* | *Clostridium* | CCR7^+^ CD8^+^ T Cells | CD8^+^ T Cells | 6.67E-06 | 1.60E-06 | 0.01833 |
| *Dysosmobacter welbionis* | *Dysosmobacter* | CCR7^+^ CD8^+^ T Cells | CD8^+^ T Cells | 5.81E-06 | 1.50E-06 | 0.01860 |
| *GGB28474 SGB41058 (CAJMQN01 sp905215695)* | *Other* | TNFα-producing NK Cells | Natural Killer (NK) Cells | 6.15E-06 | 1.60E-06 | 0.01919 |
| *GGB6601 SGB9333 (CAG-267 sp001917135)* | *Other* | PD-1^+^ CD8^+^ T Cells | CD8^+^ T Cells | 9.42E-05 | 2.40E-05 | 0.01968 |
| *Streptococcus thermophilus* | *Streptococcus* | IFNγ-producing CD161^+^ CD8^+^ T Cells | CD8^+^ T Cells | 6.68E-06 | 1.50E-06 | 0.01975 |
| *Lachnospiraceae bacterium* | *Lachnospiraceae* | Non-classical Monocytes | Innate Immune Cells | 2.52E-04 | 6.10E-05 | 0.02061 |
| *Prevotella marseillensis* | *Prevotella* | TNFα-producing NK Cells | Natural Killer (NK) Cells | 1.97E-04 | 5.10E-05 | 0.02136 |
| *GGB9512 SGB14909 (Negativibacillus sp000435195)* | *Negativibacillus* | TNFα-producing NK Cells | Natural Killer (NK) Cells | 1.89E-05 | 4.90E-06 | 0.02136 |
| *GGB3588 SGB4808 (Blautia A sp900551715)* | *Blautia* | IL-17A-producing Activated CD69^+^ CD8^+^ T Cells | CD8^+^ T Cells | 1.34E-05 | 3.40E-06 | 0.02278 |
| *GGB9595 SGB15019 (Faecousia sp900540635)* | *Faecousia* | CTLA-4^+^ Activated HLA-DR^+^ CD8^+^ T Cells | CD8^+^ T Cells | 1.21E-04 | 2.60E-05 | 0.02288 |
| *Prevotella marseillensis* | *Prevotella* | Non-classical Monocytes | Innate Immune Cells | 9.58E-04 | 2.40E-04 | 0.02339 |
| *Clostridiaceae unclassified SGB15090 (CAG-83 sp000435975)* | *Other* | IL-17A-producing CD4^+^ T Cells | CD4^+^ T Cells | 2.61E-05 | 6.70E-06 | 0.02362 |
| *Lachnospira eligens* | *Lachnospiraceae* | Natural Killer T Cells | Adaptive Immune Cells | 1.74E-05 | 4.50E-06 | 0.02396 |
| *Monoglobus pectinilyticus* | *Monoglobus* | IL-17A-producing CD4^+^ EM T Cells | CD4^+^ T Cells | 4.04E-05 | 9.70E-06 | 0.02466 |
| *Pseudoruminococcus massiliensis* | *Ruminococcus* | IL-17A-producing CD4^+^ EM T Cells | CD4^+^ T Cells | 2.02E-04 | 4.90E-05 | 0.02466 |
| *Ruminococcus bovis* | *Ruminococcus* | IL-17A-producing CD4^+^ EM T Cells | CD4^+^ T Cells | 2.75E-04 | 6.60E-05 | 0.02466 |
| *GGB6601 SGB9333 (CAG-267 sp001917135)* | *Other* | IL-17A-producing CD4^+^ EM T Cells | CD4^+^ T Cells | 5.19E-04 | 1.20E-04 | 0.02466 |
| *GGB9699 SGB15216 (Faecousia sp003525905)* | *Faecousia* | CD8^+^ TEMRA | CD8^+^ T Cells | 3.44E-06 | 8.40E-07 | 0.02499 |
| *GGB9699 SGB15216 (Faecousia sp003525905)* | *Faecousia* | TNFα-producing Activated CD69^+^ CD8^+^ T Cells | CD8^+^ T Cells | 7.72E-06 | 1.80E-06 | 0.02506 |
| *GGB4717 SGB6533 (Scybalousia sp900542735)* | *Other* | IL-17A-producing Activated HLA-DR^+^ CD8^+^ T Cells | CD8^+^ T Cells | 8.72E-05 | 1.90E-05 | 0.02521 |
| *Monoglobus pectinilyticus* | *Monoglobus* | IL-17A-producing Activated HLA-DR^+^ CD8^+^ T Cells | CD8^+^ T Cells | 6.84E-05 | 1.60E-05 | 0.02521 |
| *Prevotella marseillensis* | *Prevotella* | Mature LDNs | Innate Immune Cells | 2.30E-04 | 6.00E-05 | 0.02528 |
| *Clostridiaceae unclassified SGB15090 (CAG-83 sp000435975)* | *Other* | Mature LDNs | Innate Immune Cells | 4.36E-05 | 1.10E-05 | 0.02528 |
| *GGB6601 SGB9333 (CAG-267 sp001917135)* | *Other* | IFNγ/TNFα-producing CD161^+^ CD8^+^ T Cells | CD8^+^ T Cells | 3.05E-04 | 7.40E-05 | 0.02543 |
| *GGB28474 SGB41058 (CAJMQN01 sp905215695)* | *Other* | IL-17A-producing Activated CD69^+^ CD8^+^ T Cells | CD8^+^ T Cells | 1.35E-05 | 3.50E-06 | 0.02544 |
| *Clostridiaceae unclassified SGB15090 (CAG-83 sp000435975)* | *Other* | TNFα-producing CD161^+^ NK Cells | Natural Killer (NK) Cells | 3.58E-05 | 9.20E-06 | 0.02550 |
| *GGB3588 SGB4808 (Blautia A sp900551715)* | *Blautia* | TNFα-producing CD161^+^ NK Cells | Natural Killer (NK) Cells | 5.69E-06 | 1.50E-06 | 0.02550 |
| *GGB4583 SGB6334 (Merdicola sp900555085)* | *Merdicola* | Activated CD69^+^ CD8^+^ T Cells | CD8^+^ T Cells | 2.50E-06 | 5.40E-07 | 0.02577 |
| *GGB28474 SGB41058 (CAJMQN01 sp905215695)* | *Other* | PD-1^+^ CD8^+^ T Cells | CD8^+^ T Cells | 2.73E-06 | 7.30E-07 | 0.02589 |
| *GGB3612 SGB4882 (COE1 sp001916965)* | *Other* | IL-17A-producing CD161^+^ CD4^+^ T Cells | CD4^+^ T Cells | 1.24E-04 | 3.00E-05 | 0.02658 |
| *GGB9063 SGB13982 (Scatosoma sp003150575)* | *Scatosoma* | IFNγ/TNFα-producing HLA-DR^+^ CD8^+^ T Cells | CD8^+^ T Cells | 8.49E-05 | 2.00E-05 | 0.02693 |
| *GGB3281 SGB4335 (Eubacterium R sp003526845)* | *Eubacterium* | IL-17A-producing Activated CD69^+^ CD8^+^ T Cells | CD8^+^ T Cells | 5.95E-05 | 1.60E-05 | 0.02712 |
| *GGB9512 SGB14909 (Negativibacillus sp000435195)* | *Negativibacillus* | IL-17A-producing Activated CD69^+^ CD8^+^ T Cells | CD8^+^ T Cells | 4.15E-05 | 1.10E-05 | 0.02712 |
| *Clostridium sp 1001270J 160509 D11* | *Clostridium* | CCR7^+^ NK Cells | Natural Killer (NK) Cells | 1.58E-04 | 3.60E-05 | 0.02726 |
| *Ruminococcus bovis* | *Ruminococcus* | CCR7^+^ NK Cells | Natural Killer (NK) Cells | 7.01E-04 | 1.60E-04 | 0.02726 |
| *Vescimonas coprocola* | *Vescimonas* | Classical Monocytes | Innate Immune Cells | 4.02E-06 | 9.20E-07 | 0.02828 |
| *Adlercreutzia equolifaciens* | *Adlercreutzia* | IFNγ-producing CD4^+^ T Cells | CD4^+^ T Cells | 1.87E-06 | 4.40E-07 | 0.02834 |
| *Streptococcus thermophilus* | *Streptococcus* | IFNγ-producing CD4^+^ T Cells | CD4^+^ T Cells | 1.03E-06 | 2.40E-07 | 0.02834 |
| *Pseudoruminococcus massiliensis* | *Ruminococcus* | IL-17A-producing Activated HLA-DR^+^ CD8^+^ T Cells | CD8^+^ T Cells | 3.34E-04 | 7.90E-05 | 0.02883 |
| *GGB6612 SGB9346 (Scatacola A faecigallinarum)* | *Other* | IL-17A-producing Activated HLA-DR^+^ CD8^+^ T Cells | CD8^+^ T Cells | 6.70E-04 | 1.60E-04 | 0.02883 |
| *Clostridiaceae unclassified SGB15090 (CAG-83 sp000435975)* | *Other* | IL-17A-producing Activated CD69^+^ CD4^+^ T Cells | CD4^+^ T Cells | 2.72E-05 | 7.10E-06 | 0.02893 |
| *Prevotella marseillensis* | *Prevotella* | CCR7^+^ Activated NK Cells | Natural Killer (NK) Cells | 7.81E-04 | 2.00E-04 | 0.02949 |
| *GGB9063 SGB13982 (Scatosoma sp003150575)* | *Scatosoma* | IFNγ-producing Activated HLA-DR^+^ CD8^+^ T Cells | CD8^+^ T Cells | 9.52E-06 | 2.40E-06 | 0.02959 |
| *GGB9699 SGB15216 (Faecousia sp003525905)* | *Faecousia* | IFNγ/TNFα-producing CD161^+^ CD8^+^ T Cells | CD8^+^ T Cells | 6.33E-05 | 1.60E-05 | 0.03062 |
| *GGB4583 SGB6334 (Merdicola sp900555085)* | *Merdicola* | CD8^+^ T Cells | CD8^+^ T Cells | 2.46E-06 | 5.40E-07 | 0.03071 |
| *Roseburia lenta* | *Roseburia* | IFNγ-producing Natural Killer T Cells | Adaptive Immune Cells | 2.71E-06 | 7.10E-07 | 0.03088 |
| *GGB6601 SGB9333 (CAG-267 sp001917135)* | *Other* | PD-1^+^ Activated HLA-DR^+^ CD4^+^ T Cells | CD4^+^ T Cells | 1.88E-04 | 5.10E-05 | 0.03114 |
| *Ruminococcus sp AF13 28* | *Ruminococcus* | IL-2-producing CD4^+^ EM T Cells | CD4^+^ T Cells | 9.62E-06 | 2.50E-06 | 0.03188 |
| *GGB9063 SGB13982 (Scatosoma sp003150575)* | *Scatosoma* | TNFα-producing Activated CD69^+^ CD8^+^ T Cells | CD8^+^ T Cells | 6.84E-06 | 1.70E-06 | 0.03219 |
| *GGB2998 SGB3988 (CAG-238 sp900542245)* | *Other* | IL-17A-producing Activated CD69^+^ CD8^+^ T Cells | CD8^+^ T Cells | 1.56E-05 | 4.20E-06 | 0.03240 |
| *Roseburia lenta* | *Roseburia* | CD8^+^ TEMRA | CD8^+^ T Cells | 1.51E-06 | 3.90E-07 | 0.03250 |
| *Prevotella marseillensis* | *Prevotella* | TNFα-producing CD161^+^ NK Cells | Natural Killer (NK) Cells | 1.83E-04 | 4.90E-05 | 0.03266 |
| *GGB9512 SGB14909 (Negativibacillus sp000435195)* | *Negativibacillus* | TNFα-producing CD161^+^ NK Cells | Natural Killer (NK) Cells | 1.77E-05 | 4.70E-06 | 0.03266 |
| *Vescimonas coprocola* | *Vescimonas* | CD8^+^ Naive | CD8^+^ T Cells | 2.65E-06 | 6.60E-07 | 0.03297 |
| *GGB4682 SGB6472 (CAG-302 sp900543825)* | *Other* | IL-17A-producing CD161^+^ NK Cells | Natural Killer (NK) Cells | 1.18E-04 | 2.70E-05 | 0.03332 |
| *Adlercreutzia equolifaciens* | *Adlercreutzia* | IL-2-producing CD161^+^ CD8^+^ T Cells | CD8^+^ T Cells | 2.00E-04 | 4.40E-05 | 0.03374 |
| *GGB9699 SGB15216 (Faecousia sp003525905)* | *Faecousia* | TNFα-producing CD8^+^ T Cells | CD8^+^ T Cells | 9.46E-06 | 2.30E-06 | 0.03491 |
| *GGB6601 SGB9333 (CAG-267 sp001917135)* | *Other* | PD-1^+^ CD4^+^ T Cells | CD4^+^ T Cells | 4.07E-05 | 1.10E-05 | 0.03502 |
| *GGB29535 SGB42321 (UBA7185 sp910585405)* | *Other* | γδ T Cells | Adaptive Immune Cells | 1.17E-06 | 3.10E-07 | 0.03544 |
| *Prevotella marseillensis* | *Prevotella* | IL-17A-producing Activated NK Cells | Natural Killer (NK) Cells | 2.29E-03 | 6.10E-04 | 0.03559 |
| *Phocaeicola plebeius* | *Phocaeicola* | Activated CD69^+^ CD8^+^ T Cells | CD8^+^ T Cells | 9.46E-06 | 2.20E-06 | 0.03581 |
| *GGB9699 SGB15216 (Faecousia sp003525905)* | *Faecousia* | Activated CD69^+^ CD8^+^ T Cells | CD8^+^ T Cells | 1.92E-06 | 4.60E-07 | 0.03581 |
| *GGB9699 SGB15216 (Faecousia sp003525905)* | *Faecousia* | IFNγ-producing CD161^+^ CD4^+^ T Cells | CD4^+^ T Cells | 7.91E-06 | 2.00E-06 | 0.03615 |
| *GGB39037 SGB4393 (UBA737 sp002431945)* | *Other* | IL-2-producing CD161^+^ CD4^+^ T Cells | CD4^+^ T Cells | 1.44E-05 | 3.30E-06 | 0.03635 |
| *GGB3303 SGB4364 (CAG-177 sp000431775)* | *Other* | IL-2-producing CD161^+^ CD4^+^ T Cells | CD4^+^ T Cells | 2.71E-05 | 6.70E-06 | 0.03635 |
| *GGB27106 SGB6188 (Clostridium sp900539375)* | *Clostridium* | IL-2-producing CD161^+^ CD4^+^ T Cells | CD4^+^ T Cells | 1.13E-05 | 2.70E-06 | 0.03635 |
| *GGB9618 SGB15064 (Dysosmobacter sp900763685)* | *Dysosmobacter* | IL-2-producing CD161^+^ CD4^+^ T Cells | CD4^+^ T Cells | 1.65E-05 | 3.90E-06 | 0.03635 |
| *GGB3281 SGB4335 (Eubacterium R sp003526845)* | *Eubacterium* | IL-17A-producing CD4^+^ T Cells | CD4^+^ T Cells | 1.84E-05 | 4.90E-06 | 0.03689 |
| *Clostridiaceae unclassified SGB15090 (CAG-83 sp000435975)* | *Other* | IL-17A-producing Activated CD69^+^ CD8^+^ T Cells | CD8^+^ T Cells | 7.88E-05 | 2.20E-05 | 0.03709 |
| *Oscillospiraceae bacterium CLA AA H250* | *Other* | IL-17A-producing Activated CD69^+^ CD8^+^ T Cells | CD8^+^ T Cells | 2.02E-04 | 5.60E-05 | 0.03709 |
| *Pseudoruminococcus massiliensis* | *Ruminococcus* | IL-17A-producing CD8^+^ EM T Cells | CD8^+^ T Cells | 3.25E-04 | 7.60E-05 | 0.03831 |
| *GGB6612 SGB9346 (Scatacola A faecigallinarum)* | *Other* | IL-17A-producing CD8^+^ EM T Cells | CD8^+^ T Cells | 6.71E-04 | 1.50E-04 | 0.03831 |
| *GGB1627 SGB2230 (Cryptobacteroides sp000433355)* | *Other* | IL-2-producing Activated CD69^+^ CD4^+^ T Cells | CD4^+^ T Cells | 3.64E-04 | 8.80E-05 | 0.03837 |
| *GGB39037 SGB4393 (UBA737 sp002431945)* | *Other* | IL-2-producing Activated CD69^+^ CD4^+^ T Cells | CD4^+^ T Cells | 2.20E-05 | 5.20E-06 | 0.03837 |
| *GGB3303 SGB4364 (CAG-177 sp000431775)* | *Other* | IL-2-producing Activated CD69^+^ CD4^+^ T Cells | CD4^+^ T Cells | 4.31E-05 | 1.10E-05 | 0.03837 |
| *GGB27106 SGB6188 (Clostridium sp900539375)* | *Clostridium* | IL-2-producing Activated CD69^+^ CD4^+^ T Cells | CD4^+^ T Cells | 1.71E-05 | 4.40E-06 | 0.03837 |
| *GGB9618 SGB15064 (Dysosmobacter sp900763685)* | *Dysosmobacter* | IL-2-producing Activated CD69^+^ CD4^+^ T Cells | CD4^+^ T Cells | 2.48E-05 | 6.30E-06 | 0.03837 |
| *Ruminococcus sp AF13 28* | *Ruminococcus* | IL-2-producing Activated CD69^+^ CD4^+^ T Cells | CD4^+^ T Cells | 2.25E-05 | 5.60E-06 | 0.03837 |
| *GGB6601 SGB9333 (CAG-267 sp001917135)* | *Other* | TNFα-producing CD161^+^ CD8^+^ T Cells | CD8^+^ T Cells | 1.84E-04 | 4.50E-05 | 0.03865 |
| *GGB9063 SGB13982 (Scatosoma sp003150575)* | *Scatosoma* | TNFα-producing CD8^+^ T Cells | CD8^+^ T Cells | 8.45E-06 | 2.10E-06 | 0.03881 |
| *GGB4571 SGB6317 (Merdicola sp001915925)* | *Merdicola* | IL-17A-producing CD161^+^ CD8^+^ T Cells | CD8^+^ T Cells | 6.93E-04 | 1.70E-04 | 0.03884 |
| *Phocaeicola plebeius* | *Phocaeicola* | IFNγ-producing CD4^+^ T Cells | CD4^+^ T Cells | 1.26E-05 | 3.10E-06 | 0.03929 |
| *Blautia SGB4815 (Blautia A sp900066355)* | *Blautia* | IL-17A-producing CD161^+^ NK Cells | Natural Killer (NK) Cells | 3.67E-05 | 8.90E-06 | 0.03931 |
| *GGB4717 SGB6533 (Scybalousia sp900542735)* | *Other* | IL-17A-producing CD4^+^ EM T Cells | CD4^+^ T Cells | 4.84E-05 | 1.20E-05 | 0.03939 |
| *GGB3278 SGB4328 (Eubacterium R sp000434995)* | *Eubacterium* | IL-2-producing CD8^+^ EM T Cells | CD8^+^ T Cells | 1.81E-04 | 4.10E-05 | 0.04096 |
| *Clostridium SGB6179 (Clostridium sp900540255)* | *Clostridium* | TNFα-producing NK Cells | Natural Killer (NK) Cells | 1.38E-05 | 3.80E-06 | 0.04191 |
| *Lachnospira sp NSJ 43* | *Lachnospiraceae* | IFNγ/TNFα-producing CD161^+^ CD8^+^ T Cells | CD8^+^ T Cells | 2.22E-04 | 5.80E-05 | 0.04202 |
| *Phocaeicola plebeius* | *Phocaeicola* | CD8^+^ T Cells | CD8^+^ T Cells | 9.34E-06 | 2.20E-06 | 0.04212 |
| *GGB28474 SGB41058 (CAJMQN01 sp905215695)* | *Other* | TNFα-producing CD161^+^ NK Cells | Natural Killer (NK) Cells | 5.51E-06 | 1.50E-06 | 0.04238 |
| *Clostridium SGB6173 (Clostridium sp000435835)* | *Clostridium* | IL-17A-producing NK Cells | Natural Killer (NK) Cells | 2.74E-05 | 6.50E-06 | 0.04248 |
| *Holdemanella biformis* | *Holdemanella* | Non-classical Monocytes | Innate Immune Cells | 4.32E-04 | 1.10E-04 | 0.04316 |
| *GGB9059 SGB13976 (CAG-349 sp003539515)* | *Other* | IL-2-producing CD4^+^ EM T Cells | CD4^+^ T Cells | 1.37E-04 | 3.70E-05 | 0.04332 |
| *Phocaeicola dorei* | *Phocaeicola* | IFNγ-producing CD161^+^ NK Cells | Natural Killer (NK) Cells | 2.94E-04 | 6.90E-05 | 0.04387 |
| *GGB9699 SGB15216 (Faecousia sp003525905)* | *Faecousia* | CD8^+^ T Cells | CD8^+^ T Cells | 1.88E-06 | 4.60E-07 | 0.04394 |
| *Dysosmobacter welbionis* | *Dysosmobacter* | CD8^+^ Naive | CD8^+^ T Cells | 7.41E-06 | 1.90E-06 | 0.04516 |
| *GGB4583 SGB6334 (Merdicola sp900555085)* | *Merdicola* | TNFα-producing CD4^+^ EM T Cells | CD4^+^ T Cells | 1.34E-05 | 3.00E-06 | 0.04556 |
| *GGB9063 SGB13982 (Scatosoma sp003150575)* | *Scatosoma* | TNFα-producing CD4^+^ EM T Cells | CD4^+^ T Cells | 9.72E-06 | 2.30E-06 | 0.04556 |
| *GGB9063 SGB13982 (Scatosoma sp003150575)* | *Scatosoma* | IFNγ-producing Activated HLA-DR^+^ CD4^+^ T Cells | CD4^+^ T Cells | 2.73E-05 | 6.90E-06 | 0.04572 |
| *GGB9063 SGB13982 (Scatosoma sp003150575)* | *Scatosoma* | IFNγ-producing CD4^+^ T Cells | CD4^+^ T Cells | 2.31E-06 | 5.90E-07 | 0.04666 |
| *GGB9512 SGB14909 (Negativibacillus sp000435195)* | *Negativibacillus* | IL-17A-producing CD4^+^ T Cells | CD4^+^ T Cells | 1.25E-05 | 3.50E-06 | 0.04677 |
| *GGB4604 SGB6369 (CAG-269 sp000438255)* | *Other* | IL-17A-producing CD161^+^ NK Cells | Natural Killer (NK) Cells | 2.07E-05 | 5.40E-06 | 0.04695 |
| *GGB4715 SGB6529 (CAG-533 sp900553855)* | *Other* | IL-17A-producing CD161^+^ NK Cells | Natural Killer (NK) Cells | 1.36E-04 | 3.50E-05 | 0.04695 |
| *GGB4689 SGB6486 (CAG-628 sp003524085)* | *Other* | IL-17A-producing CD161^+^ NK Cells | Natural Killer (NK) Cells | 1.34E-04 | 3.50E-05 | 0.04695 |
| *GGB51269 SGB5062 (CAG-882 sp003486385)* | *Other* | IL-17A-producing Activated HLA-DR^+^ CD8^+^ T Cells | CD8^+^ T Cells | 1.87E-04 | 4.80E-05 | 0.04733 |
| *Clostridiales bacterium* | *Other* | CCR7^+^ CD8^+^ T Cells | CD8^+^ T Cells | 1.92E-06 | 5.30E-07 | 0.04878 |
| *GGB9699 SGB15216 (Faecousia sp003525905)* | *Faecousia* | IFNγ/TNFα-producing HLA-DR^+^ CD8^+^ T Cells | CD8^+^ T Cells | 8.89E-05 | 2.30E-05 | 0.04936 |
| *GGB6601 SGB9333 (CAG-267 sp001917135)* | *Other* | IL-17A-producing Activated CD69^+^ CD8^+^ T Cells | CD8^+^ T Cells | 4.20E-04 | 1.20E-04 | 0.04980 |

Differential abundance analyses were conducted using the R package MaAsLin2 (v1.15.1), employing linear regressions adjusted for age, sex, and BMI. Relative abundances were normalised using the centred log-ratio (CLR) method. Adjustments were performed using the Benjamini–Hochberg False FDR method, and all results presented meet the significance threshold of q<0.05. CCR7, C-C chemokine receptor type 7; CM, central memory; EM, effector memory; HLA-DR, human leukocyte antigen-DR isotype; IFN-γ, interferon-γ; IL-, interleukin-; NK, natural killer; PD-1, programmed cell death protein 1; PMDSCs, polymorphonuclear myeloid-derived suppressor cells; T regs, T regulatory cells; TEMRA, terminally differentiated effector memory T cells; TNF-α, tumour necrosis factor-α.

**Table S6. Genus- and corresponding species-level bacteria as referenced in Figure 3B.**

| **Genus** | **Species** |
| --- | --- |
| ***Adlercreutzia*** | *Adlercreutzia equolifaciens* |
| ***Blautia*** | *GGB3588 SGB4808 (Blautia A sp900551715)* |
|  | *Blautia SGB4815 (Blautia A sp900066355)* |
| ***Clostridium*** | *Clostridium sp 1001270J 160509 D11* |
|  | *Clostridium SGB6179 (Clostridium sp900540255)* |
|  | *Clostridium SGB6173 (Clostridium sp000435835)* |
|  | *GGB27106 SGB6188 (Clostridium sp900539375)* |
|  | *Clostridium sp AM22 11AC* |
| ***Dysosmobacter*** | *GGB9618 SGB15064 (Dysosmobacter sp900763685)* |
|  | *Dysosmobacter welbionis* |
| ***Eubacterium*** | *Eubacterium ventriosum* |
|  | *GGB3281 SGB4335 (Eubacterium R sp003526845)* |
|  | *GGB3278 SGB4328 (Eubacterium R sp000434995)* |
| ***Faecousia*** | *GGB9699 SGB15216 (Faecousia sp003525905)* |
|  | *GGB9595 SGB15019 (Faecousia sp900540635)* |
| ***Fimimonas*** | *GGB9172 SGB14110 (Fimimonas sp003537755)* |
| ***Holdemanella*** | *Holdemanella biformis* |
| ***Lachnospiraceae*** | *Lachnospiraceae bacterium* |
|  | *Lachnospiraceae bacterium OM04 12BH* |
|  | *Lachnospira sp NSJ 43* |
|  | *Lachnospira eligens* |
| ***Merdicola*** | *GGB4571 SGB6317 (Merdicola sp001915925)* |
|  | *GGB4583 SGB6334 (Merdicola sp900555085)* |
| ***Monoglobus*** | *Monoglobus pectinilyticus* |
| ***Negativibacillus*** | *GGB9512 SGB14909 (Negativibacillus sp000435195)* |
| **Other** | *GGB3612 SGB4882 (COE1 sp001916965)* |
|  | *Clostridiaceae unclassified SGB15090 (CAG-83 sp000435975)* |
|  | *GGB28474 SGB41058 (CAJMQN01 sp905215695)* |
|  | *GGB9260 SGB14208 (CAG-448 sp000433415)* |
|  | *GGB4682 SGB6472 (CAG-302 sp900543825)* |
|  | *GGB4604 SGB6369 (CAG-269 sp000438255)* |
|  | *GGB4715 SGB6529 (CAG-533 sp900553855)* |
|  | *GGB4689 SGB6486 (CAG-628 sp003524085)* |
|  | *GGB6601 SGB9333 (CAG-267 sp001917135)* |
|  | *GGB39037 SGB4393 (UBA737 sp002431945)* |
|  | *GGB3303 SGB4364 (CAG-177 sp000431775)* |
|  | *GGB9762 SGB15377 (SFMI01 sp004556155)* |
|  | *GGB9059 SGB13976 (CAG-349 sp003539515)* |
|  | *GGB29535 SGB42321 (UBA7185 sp910585405)* |
|  | *GGB1627 SGB2230 (Cryptobacteroides sp000433355)* |
|  | *GGB4717 SGB6533 (Scybalousia sp900542735)* |
|  | *GGB3475 SGB4638 (V9D3004 sp900760345)* |
|  | *GGB9724 SGB15278 (Fournierella excrementavium)* |
|  | *Clostridiales bacterium* |
|  | *GGB9088 SGB14017 (UMGS1795 sp900555125)* |
|  | *GGB6612 SGB9346 (Scatacola A faecigallinarum)* |
|  | *GGB2998 SGB3988 (CAG-238 sp900542245)* |
|  | *Oscillospiraceae bacterium CLA AA H250* |
|  | *GGB51269 SGB5062 (CAG-882 sp003486385)* |
| ***Phocaeicola*** | *Phocaeicola dorei* |
|  | *Phocaeicola plebeius* |
|  | *Phocaeicola faecalis* |
| ***Prevotella*** | *Prevotella marseillensis* |
| ***Roseburia*** | *Roseburia lenta* |
| ***Ruminococcus*** | *Ruminococcus bovis* |
|  | *Ruminococcus sp AF13 28* |
|  | *Pseudoruminococcus massiliensis* |
| ***Ruthenibacterium*** | *Ruthenibacterium lactatiformans* |
| ***Scatosoma*** | *GGB9063 SGB13982 (Scatosoma sp003150575)* |
| ***Streptococcus*** | *Streptococcus thermophilus* |
| ***Sutterella*** | *Sutterella sp AM11 39* |
| ***Vescimonas*** | *Vescimonas coprocola* |

**Table S7. Immune cell categories and corresponding individual cell types as referenced in Figure 3.**

|  | **Individual cell type** | |
| --- | --- | --- |
| **Immune cell category** | **Figure 3B** | **Figure 3C** |
| **Innate Immune Cells** | Classical Monocytes |  |
|  | Non-classical monocytes | Non-classical monocytes |
|  | Mature LDNs | Mature LDNs |
|  | PMDSCs | PMDSCs |
| **Natural Killer (NK) Cells** | CCR7^+^ Activated NK Cells | CCR7^+^ Activated NK Cells |
|  | CCR7^+^ NK Cells |  |
|  | IFNγ-producing CD161^+^ NK Cells | IFNγ-producing CD161^+^ NK Cells |
|  |  | IFNγ-producing Activated NK Cells |
|  |  | IFNγ-producing NK Cells |
|  | IL-17A-producing Activated NK Cells | IL-17A-producing Activated NK Cells |
|  | IL-17A-producing CD161^+^ NK Cells |  |
|  | IL-17A-producing NK Cells |  |
|  | TNFα-producing CD161^+^ NK Cells | TNFα-producing CD161^+^ NK Cells |
|  | TNFα-producing NK Cells | TNFα-producing NK Cells |
| **Adaptive Immune Cells** | γδ T Cells | γδ T Cells |
|  | IFNγ-producing Natural Killer T Cells | IFNγ-producing Natural Killer T Cells |
|  |  | IFNγ-producing T regulatory Cells |
|  |  | IL-2-producing γδ T Cells |
|  | Natural Killer T Cells | Natural Killer T Cells |
|  |  | TNFα-producing γδ T Cells |
|  | TNFα-producing Natural Killer T Cells | TNFα-producing Natural Killer T Cells |
|  |  | TNFα-producing T regulatory Cells |
| **CD4^+^ T Cells** | CD4^+^ TEMRA | CD4^+^ TEMRA |
|  | IFNγ/TNFα-producing CD161^+^ CD4^+^ T Cells | IFNγ/TNFα-producing CD161^+^ CD4^+^ T Cells |
|  | IFNγ/TNFα-producing CD69^+^ CD4^+^ T Cells | IFNγ/TNFα-producing CD69^+^ CD4^+^ T Cells |
|  | IFNγ-producing Activated CD69^+^ CD4^+^ T Cells | IFNγ-producing Activated CD69^+^ CD4^+^ T Cells |
|  | IFNγ-producing Activated HLA-DR^+^ CD4^+^ T Cells | IFNγ-producing Activated HLA-DR^+^ CD4^+^ T Cells |
|  | IFNγ-producing CD161^+^ CD4^+^ T Cells | IFNγ-producing CD161^+^ CD4^+^ T Cells |
|  | IFNγ-producing CD4^+^ EM T Cells | IFNγ-producing CD4^+^ EM T Cells |
|  | IFNγ-producing CD4^+^ T Cells | IFNγ-producing CD4^+^ T Cells |
|  | IL-17A-producing Activated CD69^+^ CD4^+^ T Cells | IL-17A-producing Activated CD69^+^ CD4^+^ T Cells |
|  | IL-17A-producing CD161^+^ CD4^+^ T Cells | IL-17A-producing CD161^+^ CD4^+^ T Cells |
|  | IL-17A-producing CD4^+^ EM T Cells | IL-17A-producing CD4^+^ EM T Cells |
|  | IL-17A-producing CD4^+^ T Cells | IL-17A-producing CD4^+^ T Cells |
|  |  | IL-2/TNFα-producing CD161^+^ CD4^+^ T Cells |
|  |  | IL-2/TNFα-producing CD4^+^ EM T Cells |
|  |  | IL-2/TNFα-producing CD4^+^ T Cells |
|  |  | IL-2/TNFα-producing CD69^+^ CD4^+^ T Cells |
|  | IL-2-producing Activated CD69^+^ CD4^+^ T Cells | IL-2-producing Activated CD69^+^ CD4^+^ T Cells |
|  | IL-2-producing CD161^+^ CD4^+^ T Cells | IL-2-producing CD161^+^ CD4^+^ T Cells |
|  | IL-2-producing CD4^+^ EM T Cells | IL-2-producing CD4^+^ EM T Cells |
|  | PD-1^+^ Activated HLA-DR^+^ CD4^+^ T Cells | PD-1^+^ Activated HLA-DR^+^ CD4^+^ T Cells |
|  | PD-1^+^ CD4^+^ T Cells | PD-1^+^ CD4^+^ T Cells |
|  | TNFα-producing CD4^+^ EM T Cells |  |
|  |  | TNFα-producing CD161^+^ CD4^+^ T Cells |
| **CD8^+^ T Cells** | Activated CD69^+^ CD8^+^ T Cells |  |
|  | Activated HLA-DR^+^ CD8^+^ T Cells |  |
|  | CCR7^+^ CD8^+^ T Cells | CCR7^+^ CD8^+^ T Cells |
|  | CD8^+^ CM | CD8^+^ CM |
|  | CD8^+^ Naive | CD8^+^ Naive |
|  | CD8^+^ T Cells |  |
|  | CD8^+^ TEMRA | CD8^+^ TEMRA |
|  | CTLA-4^+^ Activated HLA-DR^+^ CD8^+^ T Cells |  |
|  | IFNγ/TNFα-producing CD161^+^ CD8^+^ T Cells | IFNγ/TNFα-producing CD161^+^ CD8^+^ T Cells |
|  | IFNγ/TNFα-producing CD69^+^ CD8^+^ T Cells | IFNγ/TNFα-producing CD69^+^ CD8^+^ T Cells |
|  | IFNγ/TNFα-producing CD8^+^ T Cells | IFNγ/TNFα-producing CD8^+^ T Cells |
|  | IFNγ/TNFα-producing HLA-DR^+^ CD8^+^ T Cells | IFNγ/TNFα-producing HLA-DR^+^ CD8^+^ T Cells |
|  | IFNγ-producing Activated CD69^+^ CD8^+^ T Cells | IFNγ-producing Activated CD69^+^ CD8^+^ T Cells |
|  | IFNγ-producing Activated HLA-DR^+^ CD8^+^ T Cells | IFNγ-producing Activated HLA-DR^+^ CD8^+^ T Cells |
|  | IFNγ-producing CD161^+^ CD8^+^ T Cells | IFNγ-producing CD161^+^ CD8^+^ T Cells |
|  | IFNγ-producing CD8^+^ EM T Cells |  |
|  | IFNγ-producing CD8^+^ T Cells | IFNγ-producing CD8^+^ T Cells |
|  | IL-17A-producing Activated CD69^+^ CD8^+^ T Cells | IL-17A-producing Activated CD69^+^ CD8^+^ T Cells |
|  | IL-17A-producing Activated HLA-DR^+^ CD8^+^ T Cells | |
|  | IL-17A-producing CD161^+^ CD8^+^ T Cells | IL-17A-producing CD161^+^ CD8^+^ T Cells |
|  | IL-17A-producing CD8^+^ EM T Cells |  |
|  | IL-17A-producing CD8^+^ T Cells | IL-17A-producing CD8^+^ T Cells |
|  | IL2-producing Activated HLA-DR^+^ CD8^+^ T Cells | |
|  | IL2-producing CD161^+^ CD8^+^ T Cells | IL-2-producing CD161^+^ CD8^+^ T Cells |
|  | IL2-producing CD8^+^ EM T Cells |  |
|  | PD-1^+^ CD8^+^ T Cells | PD-1^+^ CD8^+^ T Cells |
|  | TNFα-producing Activated CD69^+^ CD8^+^ T Cells | TNFα-producing Activated CD69^+^ CD8^+^ T Cells |
|  | TNFα-producing Activated HLA-DR^+^ CD8^+^ T Cells | TNFα-producing Activated HLA-DR^+^ CD8^+^ T Cells |
|  | TNFα-producing CD161^+^ CD8^+^ T Cells |  |
|  | TNFα-producing CD8^+^ T Cells | TNFα-producing CD8^+^ T Cells |

CCR7, C-C chemokine receptor type 7; CM, central memory; EM, effector memory; HLA-DR, human leukocyte antigen-DR isotype; IFN-γ, interferon-γ; IL-, interleukin-; NK, natural killer; PD-1, programmed cell death protein 1; PMDSCs, polymorphonuclear myeloid-derived suppressor cells; T regs, T regulatory cells; TEMRA, terminally differentiated effector memory T cells; TNF-α, tumour necrosis factor-α.

**Table S8. Differential abundance analysis for microbial metabolic pathway (metacyc) at the species level in relation to the peripheral immune system.**

| **Functional pathway** | | **Taxonomy** | |  |  | |  |  | |
| --- | --- | --- | --- | --- | --- | --- | --- | --- | --- |
| **metaCyc pathway** | **Category** | ***Genus*** | ***Species*** | **Immune cell population/**  **subtype** | **Immune category** | **β coefficient** | **SE** | | **q-value (FDR)** |
| Methylerythritol phosphate pathway I | General Biosynthetic Pathways | *Holdemanella* | *Holdemanella biformis* | TNFα-producing T regs | Adaptive Immune Cells | 8.53E-10 | 7.09E-11 | | 3.29E-13 |
| Methylerythritol phosphate pathway I | General Biosynthetic Pathways | *Holdemanella* | *Holdemanella biformis* | PMDSCs | Innate Immune Cells | 9.72E-10 | 8.22E-11 | | 5.40E-13 |
| Methylerythritol phosphate pathway I | General Biosynthetic Pathways | *Holdemanella* | *Holdemanella biformis* | PD-1^+^ CD4^+^ T Cells | CD4^+^ T Cells | 4.00E-10 | 3.39E-11 | | 7.12E-13 |
| Flavin biosynthesis I (bacteria and plants) | Cofactor & Vitamin Biosynthesis | *Holdemanella* | *Holdemanella biformis* | TNFα-producing T regs | Adaptive Immune Cells | 8.01E-10 | 6.85E-11 | | 8.04E-13 |
| Flavin biosynthesis I (bacteria and plants) | Cofactor & Vitamin Biosynthesis | *Holdemanella* | *Holdemanella biformis* | PD-1^+^ CD4^+^ T Cells | CD4^+^ T Cells | 3.78E-10 | 3.23E-11 | | 9.06E-13 |
| Methylerythritol phosphate pathway I | General Biosynthetic Pathways | *Holdemanella* | *Holdemanella biformis* | TNFα-producing gd T Cells | Adaptive Immune Cells | 3.69E-10 | 3.19E-11 | | 1.17E-12 |
| Flavin biosynthesis I (bacteria and plants) | Cofactor & Vitamin Biosynthesis | *Holdemanella* | *Holdemanella biformis* | PMDSCs | Innate Immune Cells | 9.12E-10 | 7.97E-11 | | 1.48E-12 |
| Flavin biosynthesis I (bacteria and plants) | Cofactor & Vitamin Biosynthesis | *Holdemanella* | *Holdemanella biformis* | PD-1^+^ Activated HLA-DR^+^ CD4^+^ T Cells | CD4^+^ T Cells | 1.72E-09 | 1.51E-10 | | 2.98E-12 |
| Flavin biosynthesis I (bacteria and plants) | Cofactor & Vitamin Biosynthesis | *Holdemanella* | *Holdemanella biformis* | TNFα-producing gd T Cells | Adaptive Immune Cells | 3.46E-10 | 3.11E-11 | | 3.92E-12 |
| Methylerythritol phosphate pathway I | General Biosynthetic Pathways | *Holdemanella* | *Holdemanella biformis* | PD-1^+^ Activated HLA-DR^+^ CD4^+^ T Cells | CD4^+^ T Cells | 1.81E-09 | 1.62E-10 | | 5.34E-12 |
| Flavin biosynthesis I (bacteria and plants) | Cofactor & Vitamin Biosynthesis | *Holdemanella* | *Holdemanella biformis* | PD-1^+^ CD8^+^ T Cells | CD8^+^ T Cells | 8.19E-10 | 7.50E-11 | | 1.08E-11 |
| Queuosine biosynthesis I (de novo) | Nucleotide Metabolism & Biosynthesis | *Coprobacter* | *Coprobacter fastidiosus* | IFNγ-producing NK T Cells | Adaptive Immune Cells | 5.10E-11 | 4.74E-12 | | 2.25E-11 |
| Methylerythritol phosphate pathway I | General Biosynthetic Pathways | *Holdemanella* | *Holdemanella biformis* | PD-1^+^ CD8^+^ T Cells | CD8^+^ T Cells | 8.53E-10 | 8.23E-11 | | 4.68E-11 |
| Queuosine biosynthesis I (de novo) | Nucleotide Metabolism & Biosynthesis | *Coprobacter* | *Coprobacter fastidiosus* | NK T Cells | Adaptive Immune Cells | 3.46E-11 | 3.36E-12 | | 9.76E-11 |
| Purine ribonucleosides degradation | Nucleotide Metabolism & Biosynthesis | *Streptococcus* | *Streptococcus thermophilus* | IFNγ-producing NK T Cells | Adaptive Immune Cells | 3.67E-11 | 3.59E-12 | | 1.03E-10 |
| L-valine biosynthesis | Amino Acid Biosynthesis | *Streptococcus* | *Streptococcus thermophilus* | IFNγ-producing NK T Cells | Adaptive Immune Cells | 3.43E-11 | 3.56E-12 | | 5.11E-10 |
| Methylerythritol phosphate pathway I | General Biosynthetic Pathways | *Holdemanella* | *Holdemanella biformis* | Mature LDNs | Innate Immune Cells | 2.10E-09 | 2.25E-10 | | 9.68E-10 |
| S-adenosyl-L-methionine salvage I | Amino Acid Biosynthesis | *Streptococcus* | *Streptococcus thermophilus* | IFNγ-producing NK T Cells | Adaptive Immune Cells | 3.82E-11 | 4.08E-12 | | 9.68E-10 |
| Methylerythritol phosphate pathway I | General Biosynthetic Pathways | *Holdemanella* | *Holdemanella biformis* | IL-17A-producing Activated CD69^+^ CD8^+^ T Cells | CD8^+^ T Cells | 3.95E-09 | 4.30E-10 | | 1.32E-09 |
| Purine ribonucleosides degradation | Nucleotide Metabolism & Biosynthesis | *Streptococcus* | *Streptococcus thermophilus* | NK T Cells | Adaptive Immune Cells | 2.45E-11 | 2.64E-12 | | 1.95E-09 |
| Flavin biosynthesis I (bacteria and plants) | Cofactor & Vitamin Biosynthesis | *Holdemanella* | *Holdemanella biformis* | TNFα-producing NK Cells | Natural Killer (NK) Cells | 1.67E-09 | 1.88E-10 | | 3.75E-09 |
| Methylerythritol phosphate pathway I | General Biosynthetic Pathways | *Holdemanella* | *Holdemanella biformis* | TNFα-producing NK Cells | Natural Killer (NK) Cells | 1.75E-09 | 2.00E-10 | | 4.97E-09 |
| L-valine biosynthesis | Amino Acid Biosynthesis | *Streptococcus* | *Streptococcus thermophilus* | NK T Cells | Adaptive Immune Cells | 2.28E-11 | 2.61E-12 | | 7.66E-09 |
| Flavin biosynthesis I (bacteria and plants) | Cofactor & Vitamin Biosynthesis | *Holdemanella* | *Holdemanella biformis* | TNFα-producing CD161^+^ NK Cells | Natural Killer (NK) Cells | 1.57E-09 | 1.83E-10 | | 8.87E-09 |
| Methylerythritol phosphate pathway I | General Biosynthetic Pathways | *Holdemanella* | *Holdemanella biformis* | TNFα-producing CD161^+^ NK Cells | Natural Killer (NK) Cells | 1.65E-09 | 1.96E-10 | | 1.38E-08 |
| S-adenosyl-L-methionine salvage I | Amino Acid Biosynthesis | *Streptococcus* | *Streptococcus thermophilus* | NK T Cells | Adaptive Immune Cells | 2.53E-11 | 2.99E-12 | | 1.56E-08 |
| Flavin biosynthesis I (bacteria and plants) | Cofactor & Vitamin Biosynthesis | *Holdemanella* | *Holdemanella biformis* | IL-17A-producing Activated CD69^+^ CD8^+^ T Cells | CD8^+^ T Cells | 3.59E-09 | 4.38E-10 | | 2.36E-08 |
| Flavin biosynthesis I (bacteria and plants) | Cofactor & Vitamin Biosynthesis | *Holdemanella* | *Holdemanella biformis* | Mature LDNs | Innate Immune Cells | 1.90E-09 | 2.31E-10 | | 2.48E-08 |
| Methylerythritol phosphate pathway I | General Biosynthetic Pathways | *Holdemanella* | *Holdemanella biformis* | IL-17A-producing CD4^+^ T Cells | CD4^+^ T Cells | 1.19E-09 | 1.45E-10 | | 4.43E-08 |
| Peptidoglycan biosynthesis I (meso-diaminopimelate containing) | Cell wall & Peptidoglycan Synthesis | *Holdemanella* | *Holdemanella biformis* | TNFα-producing T regs | Adaptive Immune Cells | 6.82E-10 | 8.60E-11 | | 5.71E-08 |
| UDP-N-acetylmuramoyl-pentapeptide biosynthesis I (meso-diaminopimelate containing) | Cell wall & Peptidoglycan Synthesis | *Holdemanella* | *Holdemanella biformis* | TNFα-producing T regs | Adaptive Immune Cells | 6.70E-10 | 8.46E-11 | | 5.86E-08 |
| UDP-N-acetylmuramoyl-pentapeptide biosynthesis II (lysine-containing) | Cell wall & Peptidoglycan Synthesis | *Holdemanella* | *Holdemanella biformis* | Mature LDNs | Innate Immune Cells | 1.87E-09 | 2.35E-10 | | 6.22E-08 |
| Peptidoglycan biosynthesis I (meso-diaminopimelate containing) | Cell wall & Peptidoglycan Synthesis | *Holdemanella* | *Holdemanella biformis* | PMDSCs | Innate Immune Cells | 7.77E-10 | 9.89E-11 | | 7.42E-08 |
| UDP-N-acetylmuramoyl-pentapeptide biosynthesis I (meso-diaminopimelate containing) | Cell wall & Peptidoglycan Synthesis | *Holdemanella* | *Holdemanella biformis* | PMDSCs | Innate Immune Cells | 7.64E-10 | 9.74E-11 | | 7.71E-08 |
| UDP-N-acetylmuramoyl-pentapeptide biosynthesis II (lysine-containing) | Cell wall & Peptidoglycan Synthesis | *Holdemanella* | *Holdemanella biformis* | TNFα-producing T regs | Adaptive Immune Cells | 7.01E-10 | 8.99E-11 | | 8.57E-08 |
| Peptidoglycan biosynthesis I (meso-diaminopimelate containing) | Cell wall & Peptidoglycan Synthesis | *Holdemanella* | *Holdemanella biformis* | Mature LDNs | Innate Immune Cells | 1.79E-09 | 2.29E-10 | | 9.73E-08 |
| UDP-N-acetylmuramoyl-pentapeptide biosynthesis II (lysine-containing) | Cell wall & Peptidoglycan Synthesis | *Holdemanella* | *Holdemanella biformis* | PMDSCs | Innate Immune Cells | 7.99E-10 | 1.03E-10 | | 1.08E-07 |
| UDP-N-acetylmuramoyl-pentapeptide biosynthesis I (meso-diaminopimelate containing) | Cell wall & Peptidoglycan Synthesis | *Holdemanella* | *Holdemanella biformis* | Mature LDNs | Innate Immune Cells | 1.76E-09 | 2.26E-10 | | 1.08E-07 |
| Coenzyme A biosynthesis II (eukaryotic) | Cofactor & Vitamin Biosynthesis | *Holdemanella* | *Holdemanella biformis* | TNFα-producing T regs | Adaptive Immune Cells | 7.11E-10 | 9.20E-11 | | 1.08E-07 |
| Peptidoglycan biosynthesis I (meso-diaminopimelate containing) | Cell wall & Peptidoglycan Synthesis | *Holdemanella* | *Holdemanella biformis* | PD-1^+^ CD4^+^ T Cells | CD4^+^ T Cells | 3.18E-10 | 4.11E-11 | | 1.12E-07 |
| UDP-N-acetylmuramoyl-pentapeptide biosynthesis I (meso-diaminopimelate containing) | Cell wall & Peptidoglycan Synthesis | *Holdemanella* | *Holdemanella biformis* | PD-1^+^ CD4^+^ T Cells | CD4^+^ T Cells | 3.13E-10 | 4.05E-11 | | 1.12E-07 |
| Flavin biosynthesis I (bacteria and plants) | Cofactor & Vitamin Biosynthesis | *Holdemanella* | *Holdemanella biformis* | IL-17A-producing CD4^+^ T Cells | CD4^+^ T Cells | 1.11E-09 | 1.41E-10 | | 1.15E-07 |
| Peptidoglycan biosynthesis I (meso-diaminopimelate containing) | Cell wall & Peptidoglycan Synthesis | *Holdemanella* | *Holdemanella biformis* | TNFα-producing gd T Cells | Adaptive Immune Cells | 2.95E-10 | 3.82E-11 | | 1.19E-07 |
| UDP-N-acetylmuramoyl-pentapeptide biosynthesis I (meso-diaminopimelate containing) | Cell wall & Peptidoglycan Synthesis | *Holdemanella* | *Holdemanella biformis* | TNFα-producing gd T Cells | Adaptive Immune Cells | 2.90E-10 | 3.76E-11 | | 1.21E-07 |
| Superpathway of coenzyme A biosynthesis III (mammals) | Cofactor & Vitamin Biosynthesis | *Holdemanella* | *Holdemanella biformis* | TNFα-producing T regs | Adaptive Immune Cells | 7.25E-10 | 9.45E-11 | | 1.29E-07 |
| Coenzyme A biosynthesis II (eukaryotic) | Cofactor & Vitamin Biosynthesis | *Holdemanella* | *Holdemanella biformis* | PMDSCs | Innate Immune Cells | 8.09E-10 | 1.06E-10 | | 1.56E-07 |
| UDP-N-acetylmuramoyl-pentapeptide biosynthesis II (lysine-containing) | Cell wall & Peptidoglycan Synthesis | *Holdemanella* | *Holdemanella biformis* | PD-1^+^ CD4^+^ T Cells | CD4^+^ T Cells | 3.27E-10 | 4.30E-11 | | 1.63E-07 |
| Coenzyme A biosynthesis II (eukaryotic) | Cofactor & Vitamin Biosynthesis | *Holdemanella* | *Holdemanella biformis* | TNFα-producing gd T Cells | Adaptive Immune Cells | 3.09E-10 | 4.06E-11 | | 1.64E-07 |
| UDP-N-acetylmuramoyl-pentapeptide biosynthesis II (lysine-containing) | Cell wall & Peptidoglycan Synthesis | *Holdemanella* | *Holdemanella biformis* | TNFα-producing gd T Cells | Adaptive Immune Cells | 3.03E-10 | 3.99E-11 | | 1.72E-07 |
| Superpathway of coenzyme A biosynthesis III (mammals) | Cofactor & Vitamin Biosynthesis | *Holdemanella* | *Holdemanella biformis* | PMDSCs | Innate Immune Cells | 8.26E-10 | 1.09E-10 | | 1.78E-07 |
| Superpathway of L-threonine biosynthesis | Amino Acid Biosynthesis | *Holdemanella* | *Holdemanella biformis* | TNFα-producing T regs | Adaptive Immune Cells | 6.74E-10 | 8.90E-11 | | 1.81E-07 |
| Coenzyme A biosynthesis II (eukaryotic) | Cofactor & Vitamin Biosynthesis | *Holdemanella* | *Holdemanella biformis* | PD-1^+^ CD4^+^ T Cells | CD4^+^ T Cells | 3.31E-10 | 4.41E-11 | | 2.15E-07 |
| Superpathway of coenzyme A biosynthesis III (mammals) | Cofactor & Vitamin Biosynthesis | *Holdemanella* | *Holdemanella biformis* | PD-1^+^ CD4^+^ T Cells | CD4^+^ T Cells | 3.38E-10 | 4.51E-11 | | 2.26E-07 |
| UDP-N-acetylmuramoyl-pentapeptide biosynthesis I (meso-diaminopimelate containing) | Cell wall & Peptidoglycan Synthesis | *Holdemanella* | *Holdemanella biformis* | PD-1^+^ Activated HLA-DR^+^ CD4^+^ T Cells | CD4^+^ T Cells | 1.42E-09 | 1.89E-10 | | 2.37E-07 |
| Superpathway of coenzyme A biosynthesis III (mammals) | Cofactor & Vitamin Biosynthesis | *Holdemanella* | *Holdemanella biformis* | TNFα-producing gd T Cells | Adaptive Immune Cells | 3.14E-10 | 4.19E-11 | | 2.38E-07 |
| Peptidoglycan biosynthesis I (meso-diaminopimelate containing) | Cell wall & Peptidoglycan Synthesis | *Holdemanella* | *Holdemanella biformis* | PD-1^+^ Activated HLA-DR^+^ CD4^+^ T Cells | CD4^+^ T Cells | 1.44E-09 | 1.92E-10 | | 2.39E-07 |
| Superpathway of L-threonine biosynthesis | Amino Acid Biosynthesis | *Holdemanella* | *Holdemanella biformis* | TNFα-producing gd T Cells | Adaptive Immune Cells | 2.93E-10 | 3.92E-11 | | 2.43E-07 |
| Superpathway of L-threonine biosynthesis | Amino Acid Biosynthesis | *Holdemanella* | *Holdemanella biformis* | PMDSCs | Innate Immune Cells | 7.66E-10 | 1.03E-10 | | 2.50E-07 |
| Methylerythritol phosphate pathway I | General Biosynthetic Pathways | *Holdemanella* | *Holdemanella biformis* | IL-17A-producing Activated CD69^+^ CD4^+^ T Cells | CD4^+^ T Cells | 1.22E-09 | 1.61E-10 | | 2.74E-07 |
| Superpathway of L-threonine biosynthesis | Amino Acid Biosynthesis | *Holdemanella* | *Holdemanella biformis* | PD-1^+^ CD4^+^ T Cells | CD4^+^ T Cells | 3.14E-10 | 4.25E-11 | | 3.02E-07 |
| Flavin biosynthesis I (bacteria and plants) | Cofactor & Vitamin Biosynthesis | *Holdemanella* | *Holdemanella biformis* | CCR7^+^ Activated NK Cells | Natural Killer (NK) Cells | 6.13E-09 | 8.22E-10 | | 3.06E-07 |
| Flavin biosynthesis I (bacteria and plants) | Cofactor & Vitamin Biosynthesis | *Holdemanella* | *Holdemanella biformis* | IL-17A-producing Activated CD69^+^ CD4^+^ T Cells | CD4^+^ T Cells | 1.15E-09 | 1.53E-10 | | 3.22E-07 |
| UDP-N-acetylmuramoyl-pentapeptide biosynthesis II (lysine-containing) | Cell wall & Peptidoglycan Synthesis | *Holdemanella* | *Holdemanella biformis* | PD-1^+^ Activated HLA-DR^+^ CD4^+^ T Cells | CD4^+^ T Cells | 1.48E-09 | 2.00E-10 | | 3.29E-07 |
| Guanosine ribonucleotides de novo biosynthesis | Nucleotide Metabolism & Biosynthesis | *Holdemanella* | *Holdemanella biformis* | Mature LDNs | Innate Immune Cells | 1.63E-09 | 2.21E-10 | | 3.84E-07 |
| Superpathway of coenzyme A biosynthesis III (mammals) | Cofactor & Vitamin Biosynthesis | *Holdemanella* | *Holdemanella biformis* | PD-1^+^ Activated HLA-DR^+^ CD4^+^ T Cells | CD4^+^ T Cells | 1.53E-09 | 2.10E-10 | | 4.52E-07 |
| UMP biosynthesis I | Nucleotide Metabolism & Biosynthesis | *Holdemanella* | *Holdemanella biformis* | Mature LDNs | Innate Immune Cells | 2.42E-09 | 3.32E-10 | | 4.76E-07 |
| UMP biosynthesis II | Nucleotide Metabolism & Biosynthesis | *Holdemanella* | *Holdemanella biformis* | Mature LDNs | Innate Immune Cells | 2.42E-09 | 3.32E-10 | | 4.76E-07 |
| UMP biosynthesis III | Nucleotide Metabolism & Biosynthesis | *Holdemanella* | *Holdemanella biformis* | Mature LDNs | Innate Immune Cells | 2.42E-09 | 3.32E-10 | | 4.76E-07 |
| Coenzyme A biosynthesis II (eukaryotic) | Cofactor & Vitamin Biosynthesis | *Holdemanella* | *Holdemanella biformis* | PD-1^+^ Activated HLA-DR^+^ CD4^+^ T Cells | CD4^+^ T Cells | 1.50E-09 | 2.06E-10 | | 4.83E-07 |
| Superpathway of L-threonine biosynthesis | Amino Acid Biosynthesis | *Holdemanella* | *Holdemanella biformis* | PD-1^+^ Activated HLA-DR^+^ CD4^+^ T Cells | CD4^+^ T Cells | 1.42E-09 | 1.98E-10 | | 6.24E-07 |
| Superpathway of coenzyme A biosynthesis III (mammals) | Cofactor & Vitamin Biosynthesis | *Holdemanella* | *Holdemanella biformis* | Mature LDNs | Innate Immune Cells | 1.85E-09 | 2.61E-10 | | 8.68E-07 |
| Methylerythritol phosphate pathway I | General Biosynthetic Pathways | *Holdemanella* | *Holdemanella biformis* | CCR7^+^ Activated NK Cells | Natural Killer (NK) Cells | 6.34E-09 | 8.93E-10 | | 9.65E-07 |
| Queuosine biosynthesis I (de novo) | Nucleotide Metabolism & Biosynthesis | *Coprobacter* | *Coprobacter fastidiosus* | TNFα-producing NK T Cells | Adaptive Immune Cells | 1.15E-10 | 1.58E-11 | | 9.76E-07 |
| Peptidoglycan biosynthesis I (meso-diaminopimelate containing) | Cell wall & Peptidoglycan Synthesis | *Holdemanella* | *Holdemanella biformis* | IL-17A-producing Activated CD69^+^ CD8^+^ T Cells | CD8^+^ T Cells | 3.23E-09 | 4.61E-10 | | 1.02E-06 |
| UDP-N-acetylmuramoyl-pentapeptide biosynthesis II (lysine-containing) | Cell wall & Peptidoglycan Synthesis | *Holdemanella* | *Holdemanella biformis* | IL-17A-producing Activated CD69^+^ CD8^+^ T Cells | CD8^+^ T Cells | 3.34E-09 | 4.78E-10 | | 1.02E-06 |
| UDP-N-acetylmuramoyl-pentapeptide biosynthesis I (meso-diaminopimelate containing) | Cell wall & Peptidoglycan Synthesis | *Holdemanella* | *Holdemanella biformis* | IL-17A-producing Activated CD69^+^ CD8^+^ T Cells | CD8^+^ T Cells | 3.17E-09 | 4.54E-10 | | 1.02E-06 |
| Superpathway of L-threonine biosynthesis | Amino Acid Biosynthesis | *Holdemanella* | *Holdemanella biformis* | Mature LDNs | Innate Immune Cells | 1.72E-09 | 2.46E-10 | | 1.26E-06 |
| Peptidoglycan biosynthesis I (meso-diaminopimelate containing) | Cell wall & Peptidoglycan Synthesis | *Holdemanella* | *Holdemanella biformis* | PD-1^+^ CD8^+^ T Cells | CD8^+^ T Cells | 6.68E-10 | 9.63E-11 | | 1.47E-06 |
| UDP-N-acetylmuramoyl-pentapeptide biosynthesis I (meso-diaminopimelate containing) | Cell wall & Peptidoglycan Synthesis | *Holdemanella* | *Holdemanella biformis* | PD-1^+^ CD8^+^ T Cells | CD8^+^ T Cells | 6.57E-10 | 9.47E-11 | | 1.47E-06 |
| Coenzyme A biosynthesis II (eukaryotic) | Cofactor & Vitamin Biosynthesis | *Holdemanella* | *Holdemanella biformis* | Mature LDNs | Innate Immune Cells | 1.79E-09 | 2.59E-10 | | 1.56E-06 |
| Guanosine ribonucleotides de novo biosynthesis | Nucleotide Metabolism & Biosynthesis | *Holdemanella* | *Holdemanella biformis* | TNFα-producing T regs | Adaptive Immune Cells | 5.96E-10 | 8.67E-11 | | 1.61E-06 |
| Guanosine ribonucleotides de novo biosynthesis | Nucleotide Metabolism & Biosynthesis | *Holdemanella* | *Holdemanella biformis* | PMDSCs | Innate Immune Cells | 6.80E-10 | 9.95E-11 | | 1.88E-06 |
| UDP-N-acetylmuramoyl-pentapeptide biosynthesis II (lysine-containing) | Cell wall & Peptidoglycan Synthesis | *Holdemanella* | *Holdemanella biformis* | PD-1^+^ CD8^+^ T Cells | CD8^+^ T Cells | 6.87E-10 | 1.01E-10 | | 2.12E-06 |
| Guanosine ribonucleotides de novo biosynthesis | Nucleotide Metabolism & Biosynthesis | *Holdemanella* | *Holdemanella biformis* | TNFα-producing gd T Cells | Adaptive Immune Cells | 2.59E-10 | 3.81E-11 | | 2.13E-06 |
| Superpathway of coenzyme A biosynthesis III (mammals) | Cofactor & Vitamin Biosynthesis | *Holdemanella* | *Holdemanella biformis* | IL-17A-producing Activated CD69^+^ CD8^+^ T Cells | CD8^+^ T Cells | 3.42E-09 | 5.07E-10 | | 2.22E-06 |
| Superpathway of coenzyme A biosynthesis III (mammals) | Cofactor & Vitamin Biosynthesis | *Holdemanella* | *Holdemanella biformis* | PD-1^+^ CD8^+^ T Cells | CD8^+^ T Cells | 7.12E-10 | 1.05E-10 | | 2.56E-06 |
| Coenzyme A biosynthesis II (eukaryotic) | Cofactor & Vitamin Biosynthesis | *Holdemanella* | *Holdemanella biformis* | IL-17A-producing Activated CD69^+^ CD8^+^ T Cells | CD8^+^ T Cells | 3.33E-09 | 4.98E-10 | | 2.71E-06 |
| Superpathway of L-threonine biosynthesis | Amino Acid Biosynthesis | *Holdemanella* | *Holdemanella biformis* | PD-1^+^ CD8^+^ T Cells | CD8^+^ T Cells | 6.63E-10 | 9.86E-11 | | 2.82E-06 |
| Coenzyme A biosynthesis II (eukaryotic) | Cofactor & Vitamin Biosynthesis | *Holdemanella* | *Holdemanella biformis* | PD-1^+^ CD8^+^ T Cells | CD8^+^ T Cells | 6.94E-10 | 1.03E-10 | | 2.85E-06 |
| Superpathway of L-threonine biosynthesis | Amino Acid Biosynthesis | *Holdemanella* | *Holdemanella biformis* | IL-17A-producing Activated CD69^+^ CD8^+^ T Cells | CD8^+^ T Cells | 3.18E-09 | 4.77E-10 | | 2.86E-06 |
| Guanosine ribonucleotides de novo biosynthesis | Nucleotide Metabolism & Biosynthesis | *Holdemanella* | *Holdemanella biformis* | PD-1^+^ CD4^+^ T Cells | CD4^+^ T Cells | 2.77E-10 | 4.15E-11 | | 3.15E-06 |
| Chorismate biosynthesis I | General Biosynthetic Pathways | *Holdemanella* | *Holdemanella biformis* | Mature LDNs | Innate Immune Cells | 1.86E-09 | 2.78E-10 | | 3.28E-06 |
| Guanosine ribonucleotides de novo biosynthesis | Nucleotide Metabolism & Biosynthesis | *Holdemanella* | *Holdemanella biformis* | PD-1^+^ Activated HLA-DR^+^ CD4^+^ T Cells | CD4^+^ T Cells | 1.25E-09 | 1.93E-10 | | 6.44E-06 |
| 5-aminoimidazole ribonucleotide biosynthesis II | Nucleotide Metabolism & Biosynthesis | *Holdemanella* | *Holdemanella biformis* | Mature LDNs | Innate Immune Cells | 2.56E-09 | 3.96E-10 | | 6.65E-06 |
| Superpathway of 5-aminoimidazole ribonucleotide biosynthesis | Nucleotide Metabolism & Biosynthesis | *Holdemanella* | *Holdemanella biformis* | Mature LDNs | Innate Immune Cells | 2.56E-09 | 3.96E-10 | | 6.65E-06 |
| Queuosine biosynthesis I (de novo) | Nucleotide Metabolism & Biosynthesis | *Coprobacter* | *Coprobacter fastidiosus* | IFNγ-producing CD161^+^ CD4^+^ T Cells | CD4^+^ T Cells | 5.52E-11 | 8.09E-12 | | 6.68E-06 |
| Chorismate biosynthesis from 3-dehydroquinate | General Biosynthetic Pathways | *Holdemanella* | *Holdemanella biformis* | Mature LDNs | Innate Immune Cells | 1.68E-09 | 2.60E-10 | | 6.70E-06 |
| UMP biosynthesis I | Nucleotide Metabolism & Biosynthesis | *Holdemanella* | *Holdemanella biformis* | TNFα-producing T regs | Adaptive Immune Cells | 8.57E-10 | 1.35E-10 | | 9.26E-06 |
| UMP biosynthesis II | Nucleotide Metabolism & Biosynthesis | *Holdemanella* | *Holdemanella biformis* | TNFα-producing T regs | Adaptive Immune Cells | 8.57E-10 | 1.35E-10 | | 9.26E-06 |
| UMP biosynthesis III | Nucleotide Metabolism & Biosynthesis | *Holdemanella* | *Holdemanella biformis* | TNFα-producing T regs | Adaptive Immune Cells | 8.57E-10 | 1.35E-10 | | 9.26E-06 |
| tRNA charging | Amino Acid Biosynthesis | *Holdemanella* | *Holdemanella biformis* | Mature LDNs | Innate Immune Cells | 1.52E-09 | 2.39E-10 | | 9.35E-06 |
| UMP biosynthesis I | Nucleotide Metabolism & Biosynthesis | *Holdemanella* | *Holdemanella biformis* | PMDSCs | Innate Immune Cells | 9.79E-10 | 1.55E-10 | | 1.04E-05 |
| UMP biosynthesis II | Nucleotide Metabolism & Biosynthesis | *Holdemanella* | *Holdemanella biformis* | PMDSCs | Innate Immune Cells | 9.79E-10 | 1.55E-10 | | 1.04E-05 |
| UMP biosynthesis III | Nucleotide Metabolism & Biosynthesis | *Holdemanella* | *Holdemanella biformis* | PMDSCs | Innate Immune Cells | 9.79E-10 | 1.55E-10 | | 1.04E-05 |
| Guanosine ribonucleotides de novo biosynthesis | Nucleotide Metabolism & Biosynthesis | *Holdemanella* | *Holdemanella biformis* | IL-17A-producing Activated CD69^+^ CD8^+^ T Cells | CD8^+^ T Cells | 2.84E-09 | 4.54E-10 | | 1.05E-05 |
| Coenzyme A biosynthesis I (prokaryotic) | Cofactor & Vitamin Biosynthesis | *Holdemanella* | *Holdemanella biformis* | Mature LDNs | Innate Immune Cells | 1.85E-09 | 2.93E-10 | | 1.08E-05 |
| Purine ribonucleosides degradation | Nucleotide Metabolism & Biosynthesis | *Streptococcus* | *Streptococcus thermophilus* | TNFα-producing NK T Cells | Adaptive Immune Cells | 7.90E-11 | 1.24E-11 | | 1.14E-05 |
| Queuosine biosynthesis I (de novo) | Nucleotide Metabolism & Biosynthesis | *Coprobacter* | *Coprobacter fastidiosus* | IFNγ/TNFα-producing CD161^+^ CD8^+^ T Cells | CD8^+^ T Cells | 4.38E-10 | 6.56E-11 | | 1.19E-05 |
| Coenzyme A biosynthesis I (prokaryotic) | Cofactor & Vitamin Biosynthesis | *Holdemanella* | *Holdemanella biformis* | TNFα-producing T regs | Adaptive Immune Cells | 6.96E-10 | 1.12E-10 | | 1.23E-05 |
| UMP biosynthesis I | Nucleotide Metabolism & Biosynthesis | *Holdemanella* | *Holdemanella biformis* | TNFα-producing gd T Cells | Adaptive Immune Cells | 3.71E-10 | 5.97E-11 | | 1.42E-05 |
| UMP biosynthesis II | Nucleotide Metabolism & Biosynthesis | *Holdemanella* | *Holdemanella biformis* | TNFα-producing gd T Cells | Adaptive Immune Cells | 3.71E-10 | 5.97E-11 | | 1.42E-05 |
| UMP biosynthesis III | Nucleotide Metabolism & Biosynthesis | *Holdemanella* | *Holdemanella biformis* | TNFα-producing gd T Cells | Adaptive Immune Cells | 3.71E-10 | 5.97E-11 | | 1.42E-05 |
| Coenzyme A biosynthesis I (prokaryotic) | Cofactor & Vitamin Biosynthesis | *Holdemanella* | *Holdemanella biformis* | PMDSCs | Innate Immune Cells | 7.92E-10 | 1.28E-10 | | 1.58E-05 |
| UMP biosynthesis I | Nucleotide Metabolism & Biosynthesis | *Holdemanella* | *Holdemanella biformis* | PD-1^+^ CD4^+^ T Cells | CD4^+^ T Cells | 3.98E-10 | 6.47E-11 | | 1.67E-05 |
| UMP biosynthesis II | Nucleotide Metabolism & Biosynthesis | *Holdemanella* | *Holdemanella biformis* | PD-1^+^ CD4^+^ T Cells | CD4^+^ T Cells | 3.98E-10 | 6.47E-11 | | 1.67E-05 |
| UMP biosynthesis III | Nucleotide Metabolism & Biosynthesis | *Holdemanella* | *Holdemanella biformis* | PD-1^+^ CD4^+^ T Cells | CD4^+^ T Cells | 3.98E-10 | 6.47E-11 | | 1.67E-05 |
| Coenzyme A biosynthesis I (prokaryotic) | Cofactor & Vitamin Biosynthesis | *Holdemanella* | *Holdemanella biformis* | TNFα-producing gd T Cells | Adaptive Immune Cells | 3.02E-10 | 4.91E-11 | | 1.70E-05 |
| Peptidoglycan biosynthesis I (meso-diaminopimelate containing) | Cell wall & Peptidoglycan Synthesis | *Holdemanella* | *Holdemanella biformis* | TNFα-producing NK Cells | Natural Killer (NK) Cells | 1.37E-09 | 2.21E-10 | | 1.70E-05 |
| UDP-N-acetylmuramoyl-pentapeptide biosynthesis I (meso-diaminopimelate containing) | Cell wall & Peptidoglycan Synthesis | *Holdemanella* | *Holdemanella biformis* | TNFα-producing NK Cells | Natural Killer (NK) Cells | 1.35E-09 | 2.17E-10 | | 1.70E-05 |
| Peptidoglycan biosynthesis I (meso-diaminopimelate containing) | Cell wall & Peptidoglycan Synthesis | *Holdemanella* | *Holdemanella biformis* | IL-17A-producing CD4^+^ T Cells | CD4^+^ T Cells | 9.55E-10 | 1.54E-10 | | 1.83E-05 |
| UDP-N-acetylmuramoyl-pentapeptide biosynthesis I (meso-diaminopimelate containing) | Cell wall & Peptidoglycan Synthesis | *Holdemanella* | *Holdemanella biformis* | IL-17A-producing CD4^+^ T Cells | CD4^+^ T Cells | 9.38E-10 | 1.52E-10 | | 1.88E-05 |
| UDP-N-acetylmuramoyl-pentapeptide biosynthesis II (lysine-containing) | Cell wall & Peptidoglycan Synthesis | *Holdemanella* | *Holdemanella biformis* | IL-17A-producing CD4^+^ T Cells | CD4^+^ T Cells | 9.86E-10 | 1.60E-10 | | 2.01E-05 |
| Coenzyme A biosynthesis I (prokaryotic) | Cofactor & Vitamin Biosynthesis | *Holdemanella* | *Holdemanella biformis* | PD-1^+^ CD4^+^ T Cells | CD4^+^ T Cells | 3.23E-10 | 5.33E-11 | | 2.12E-05 |
| UDP-N-acetylmuramoyl-pentapeptide biosynthesis II (lysine-containing) | Cell wall & Peptidoglycan Synthesis | *Holdemanella* | *Holdemanella biformis* | TNFα-producing NK Cells | Natural Killer (NK) Cells | 1.41E-09 | 2.30E-10 | | 2.16E-05 |
| UMP biosynthesis I | Nucleotide Metabolism & Biosynthesis | *Holdemanella* | *Holdemanella biformis* | IL-17A-producing Activated CD69^+^ CD8^+^ T Cells | CD8^+^ T Cells | 4.17E-09 | 6.93E-10 | | 2.42E-05 |
| UMP biosynthesis II | Nucleotide Metabolism & Biosynthesis | *Holdemanella* | *Holdemanella biformis* | IL-17A-producing Activated CD69^+^ CD8^+^ T Cells | CD8^+^ T Cells | 4.17E-09 | 6.93E-10 | | 2.42E-05 |
| UMP biosynthesis III | Nucleotide Metabolism & Biosynthesis | *Holdemanella* | *Holdemanella biformis* | IL-17A-producing Activated CD69^+^ CD8^+^ T Cells | CD8^+^ T Cells | 4.17E-09 | 6.93E-10 | | 2.42E-05 |
| L-valine biosynthesis | Amino Acid Biosynthesis | *Streptococcus* | *Streptococcus thermophilus* | TNFα-producing NK T Cells | Adaptive Immune Cells | 7.34E-11 | 1.20E-11 | | 2.47E-05 |
| 5-aminoimidazole ribonucleotide biosynthesis I | Nucleotide Metabolism & Biosynthesis | *Holdemanella* | *Holdemanella biformis* | Mature LDNs | Innate Immune Cells | 2.29E-09 | 3.80E-10 | | 2.57E-05 |
| UMP biosynthesis I | Nucleotide Metabolism & Biosynthesis | *Holdemanella* | *Holdemanella biformis* | PD-1^+^ Activated HLA-DR^+^ CD4^+^ T Cells | CD4^+^ T Cells | 1.80E-09 | 3.00E-10 | | 2.68E-05 |
| UMP biosynthesis II | Nucleotide Metabolism & Biosynthesis | *Holdemanella* | *Holdemanella biformis* | PD-1^+^ Activated HLA-DR^+^ CD4^+^ T Cells | CD4^+^ T Cells | 1.80E-09 | 3.00E-10 | | 2.68E-05 |
| UMP biosynthesis III | Nucleotide Metabolism & Biosynthesis | *Holdemanella* | *Holdemanella biformis* | PD-1^+^ Activated HLA-DR^+^ CD4^+^ T Cells | CD4^+^ T Cells | 1.80E-09 | 3.00E-10 | | 2.68E-05 |
| Purine ribonucleosides degradation | Nucleotide Metabolism & Biosynthesis | *Streptococcus* | *Streptococcus thermophilus* | IFNγ-producing CD161^+^ CD4^+^ T Cells | CD4^+^ T Cells | 3.89E-11 | 6.12E-12 | | 2.79E-05 |
| S-adenosyl-L-methionine salvage I | Amino Acid Biosynthesis | *Streptococcus* | *Streptococcus thermophilus* | TNFα-producing NK T Cells | Adaptive Immune Cells | 8.22E-11 | 1.35E-11 | | 2.80E-05 |
| Guanosine ribonucleotides de novo biosynthesis | Nucleotide Metabolism & Biosynthesis | *Holdemanella* | *Holdemanella biformis* | PD-1^+^ CD8^+^ T Cells | CD8^+^ T Cells | 5.78E-10 | 9.63E-11 | | 2.83E-05 |
| Superpathway of coenzyme A biosynthesis III (mammals) | Cofactor & Vitamin Biosynthesis | *Holdemanella* | *Holdemanella biformis* | TNFα-producing NK Cells | Natural Killer (NK) Cells | 1.46E-09 | 2.41E-10 | | 2.89E-05 |
| Coenzyme A biosynthesis I (prokaryotic) | Cofactor & Vitamin Biosynthesis | *Holdemanella* | *Holdemanella biformis* | PD-1^+^ Activated HLA-DR^+^ CD4^+^ T Cells | CD4^+^ T Cells | 1.47E-09 | 2.46E-10 | | 3.12E-05 |
| Coenzyme A biosynthesis II (eukaryotic) | Cofactor & Vitamin Biosynthesis | *Holdemanella* | *Holdemanella biformis* | TNFα-producing NK Cells | Natural Killer (NK) Cells | 1.42E-09 | 2.37E-10 | | 3.19E-05 |
| Queuosine biosynthesis I (de novo) | Nucleotide Metabolism & Biosynthesis | *Coprobacter* | *Coprobacter fastidiosus* | IFNγ-producing Activated CD69^+^ CD4^+^ T Cells | CD4^+^ T Cells | 2.39E-11 | 3.78E-12 | | 3.29E-05 |
| Chorismate biosynthesis I | General Biosynthetic Pathways | *Holdemanella* | *Holdemanella biformis* | TNFα-producing T regs | Adaptive Immune Cells | 6.60E-10 | 1.12E-10 | | 3.50E-05 |
| Peptidoglycan biosynthesis I (meso-diaminopimelate containing) | Cell wall & Peptidoglycan Synthesis | *Holdemanella* | *Holdemanella biformis* | TNFα-producing CD161^+^ NK Cells | Natural Killer (NK) Cells | 1.29E-09 | 2.15E-10 | | 3.57E-05 |
| UDP-N-acetylmuramoyl-pentapeptide biosynthesis I (meso-diaminopimelate containing) | Cell wall & Peptidoglycan Synthesis | *Holdemanella* | *Holdemanella biformis* | TNFα-producing CD161^+^ NK Cells | Natural Killer (NK) Cells | 1.26E-09 | 2.11E-10 | | 3.57E-05 |
| S-adenosyl-L-methionine salvage I | Amino Acid Biosynthesis | *Holdemanella* | *Holdemanella biformis* | Mature LDNs | Innate Immune Cells | 2.12E-09 | 3.57E-10 | | 3.62E-05 |
| Chorismate biosynthesis I | General Biosynthetic Pathways | *Holdemanella* | *Holdemanella biformis* | PMDSCs | Innate Immune Cells | 7.53E-10 | 1.28E-10 | | 3.85E-05 |
| Superpathway of L-threonine biosynthesis | Amino Acid Biosynthesis | *Holdemanella* | *Holdemanella biformis* | TNFα-producing NK Cells | Natural Killer (NK) Cells | 1.35E-09 | 2.27E-10 | | 3.97E-05 |
| Superpathway of coenzyme A biosynthesis III (mammals) | Cofactor & Vitamin Biosynthesis | *Holdemanella* | *Holdemanella biformis* | IL-17A-producing CD4^+^ T Cells | CD4^+^ T Cells | 1.01E-09 | 1.69E-10 | | 4.04E-05 |
| Coenzyme A biosynthesis II (eukaryotic) | Cofactor & Vitamin Biosynthesis | *Holdemanella* | *Holdemanella biformis* | IL-17A-producing CD4^+^ T Cells | CD4^+^ T Cells | 9.82E-10 | 1.66E-10 | | 4.30E-05 |
| UDP-N-acetylmuramoyl-pentapeptide biosynthesis II (lysine-containing) | Cell wall & Peptidoglycan Synthesis | *Holdemanella* | *Holdemanella biformis* | TNFα-producing CD161^+^ NK Cells | Natural Killer (NK) Cells | 1.32E-09 | 2.24E-10 | | 4.47E-05 |
| Queuosine biosynthesis I (de novo) | Nucleotide Metabolism & Biosynthesis | *Coprobacter* | *Coprobacter fastidiosus* | IFNγ-producing CD4^+^ EM T Cells | CD4^+^ T Cells | 5.29E-11 | 8.77E-12 | | 4.49E-05 |
| Superpathway of L-threonine biosynthesis | Amino Acid Biosynthesis | *Holdemanella* | *Holdemanella biformis* | IL-17A-producing CD4^+^ T Cells | CD4^+^ T Cells | 9.35E-10 | 1.59E-10 | | 4.75E-05 |
| Chorismate biosynthesis from 3-dehydroquinate | General Biosynthetic Pathways | *Holdemanella* | *Holdemanella biformis* | TNFα-producing T regs | Adaptive Immune Cells | 5.97E-10 | 1.03E-10 | | 5.31E-05 |
| Chorismate biosynthesis I | General Biosynthetic Pathways | *Holdemanella* | *Holdemanella biformis* | TNFα-producing gd T Cells | Adaptive Immune Cells | 2.85E-10 | 4.92E-11 | | 5.32E-05 |
| Coenzyme A biosynthesis I (prokaryotic) | Cofactor & Vitamin Biosynthesis | *Holdemanella* | *Holdemanella biformis* | IL-17A-producing Activated CD69^+^ CD8^+^ T Cells | CD8^+^ T Cells | 3.32E-09 | 5.80E-10 | | 5.74E-05 |
| L-valine biosynthesis | Amino Acid Biosynthesis | *Streptococcus* | *Streptococcus thermophilus* | IFNγ-producing CD161^+^ CD4^+^ T Cells | CD4^+^ T Cells | 3.62E-11 | 5.93E-12 | | 5.84E-05 |
| Chorismate biosynthesis from 3-dehydroquinate | General Biosynthetic Pathways | *Holdemanella* | *Holdemanella biformis* | PMDSCs | Innate Immune Cells | 6.81E-10 | 1.19E-10 | | 5.85E-05 |
| Superpathway of coenzyme A biosynthesis III (mammals) | Cofactor & Vitamin Biosynthesis | *Holdemanella* | *Holdemanella biformis* | TNFα-producing CD161^+^ NK Cells | Natural Killer (NK) Cells | 1.36E-09 | 2.34E-10 | | 5.89E-05 |
| Queuosine biosynthesis I (de novo) | Nucleotide Metabolism & Biosynthesis | *Coprobacter* | *Coprobacter fastidiosus* | CD8^+^ TEMRA | CD8^+^ T Cells | 2.19E-11 | 3.64E-12 | | 6.23E-05 |
| Coenzyme A biosynthesis II (eukaryotic) | Cofactor & Vitamin Biosynthesis | *Holdemanella* | *Holdemanella biformis* | TNFα-producing CD161^+^ NK Cells | Natural Killer (NK) Cells | 1.33E-09 | 2.30E-10 | | 6.57E-05 |
| Chorismate biosynthesis I | General Biosynthetic Pathways | *Holdemanella* | *Holdemanella biformis* | PD-1^+^ CD4^+^ T Cells | CD4^+^ T Cells | 3.05E-10 | 5.34E-11 | | 6.69E-05 |
| Purine ribonucleosides degradation | Nucleotide Metabolism & Biosynthesis | *Streptococcus* | *Streptococcus thermophilus* | IFNγ-producing Activated CD69^+^ CD4^+^ T Cells | CD4^+^ T Cells | 1.70E-11 | 2.83E-12 | | 7.32E-05 |
| Purine ribonucleosides degradation | Nucleotide Metabolism & Biosynthesis | *Streptococcus* | *Streptococcus thermophilus* | IFNγ/TNFα-producing CD161^+^ CD8^+^ T Cells | CD8^+^ T Cells | 3.04E-10 | 5.03E-11 | | 7.40E-05 |
| Superpathway of L-threonine biosynthesis | Amino Acid Biosynthesis | *Holdemanella* | *Holdemanella biformis* | TNFα-producing CD161^+^ NK Cells | Natural Killer (NK) Cells | 1.26E-09 | 2.21E-10 | | 7.87E-05 |
| Purine ribonucleosides degradation | Nucleotide Metabolism & Biosynthesis | *Streptococcus* | *Streptococcus thermophilus* | CD8^+^ TEMRA | CD8^+^ T Cells | 1.58E-11 | 2.68E-12 | | 8.06E-05 |
| Chorismate biosynthesis from 3-dehydroquinate | General Biosynthetic Pathways | *Holdemanella* | *Holdemanella biformis* | TNFα-producing gd T Cells | Adaptive Immune Cells | 2.58E-10 | 4.57E-11 | | 8.24E-05 |
| Purine ribonucleosides degradation | Nucleotide Metabolism & Biosynthesis | *Streptococcus* | *Streptococcus thermophilus* | IFNγ-producing CD4^+^ EM T Cells | CD4^+^ T Cells | 3.78E-11 | 6.52E-12 | | 8.48E-05 |
| Queuosine biosynthesis I (de novo) | Nucleotide Metabolism & Biosynthesis | *Coprobacter* | *Coprobacter fastidiosus* | IFNγ/TNFα-producing CD161^+^ CD4^+^ T Cells | CD4^+^ T Cells | 8.89E-11 | 1.46E-11 | | 8.57E-05 |
| tRNA charging | Amino Acid Biosynthesis | *Holdemanella* | *Holdemanella biformis* | TNFα-producing T regs | Adaptive Immune Cells | 5.36E-10 | 9.56E-11 | | 8.57E-05 |
| Queuosine biosynthesis I (de novo) | Nucleotide Metabolism & Biosynthesis | *Coprobacter* | *Coprobacter fastidiosus* | IFNγ/TNFα-producing CD69^+^ CD8^+^ T Cells | CD8^+^ T Cells | 1.72E-10 | 2.86E-11 | | 9.00E-05 |
| Chorismate biosynthesis I | General Biosynthetic Pathways | *Holdemanella* | *Holdemanella biformis* | IL-17A-producing Activated CD69^+^ CD8^+^ T Cells | CD8^+^ T Cells | 3.19E-09 | 5.72E-10 | | 9.23E-05 |
| Chorismate biosynthesis from 3-dehydroquinate | General Biosynthetic Pathways | *Holdemanella* | *Holdemanella biformis* | PD-1^+^ CD4^+^ T Cells | CD4^+^ T Cells | 2.76E-10 | 4.94E-11 | | 9.32E-05 |
| tRNA charging | Amino Acid Biosynthesis | *Holdemanella* | *Holdemanella biformis* | PMDSCs | Innate Immune Cells | 6.11E-10 | 1.10E-10 | | 9.66E-05 |
| UMP biosynthesis I | Nucleotide Metabolism & Biosynthesis | *Holdemanella* | *Holdemanella biformis* | PD-1^+^ CD8^+^ T Cells | CD8^+^ T Cells | 8.30E-10 | 1.49E-10 | | 1.03E-04 |
| UMP biosynthesis II | Nucleotide Metabolism & Biosynthesis | *Holdemanella* | *Holdemanella biformis* | PD-1^+^ CD8^+^ T Cells | CD8^+^ T Cells | 8.30E-10 | 1.49E-10 | | 1.03E-04 |
| UMP biosynthesis III | Nucleotide Metabolism & Biosynthesis | *Holdemanella* | *Holdemanella biformis* | PD-1^+^ CD8^+^ T Cells | CD8^+^ T Cells | 8.30E-10 | 1.49E-10 | | 1.03E-04 |
| Chorismate biosynthesis I | General Biosynthetic Pathways | *Holdemanella* | *Holdemanella biformis* | PD-1^+^ Activated HLA-DR^+^ CD4^+^ T Cells | CD4^+^ T Cells | 1.38E-09 | 2.48E-10 | | 1.13E-04 |
| Flavin biosynthesis I (bacteria and plants) | Cofactor & Vitamin Biosynthesis | *Holdemanella* | *Holdemanella biformis* | IL-17A-producing Activated NK Cells | Natural Killer (NK) Cells | 1.62E-08 | 2.81E-09 | | 1.14E-04 |
| Peptidoglycan biosynthesis I (meso-diaminopimelate containing) | Cell wall & Peptidoglycan Synthesis | *Holdemanella* | *Holdemanella biformis* | IL-17A-producing Activated CD69^+^ CD4^+^ T Cells | CD4^+^ T Cells | 9.58E-10 | 1.70E-10 | | 1.16E-04 |
| UDP-N-acetylmuramoyl-pentapeptide biosynthesis I (meso-diaminopimelate containing) | Cell wall & Peptidoglycan Synthesis | *Holdemanella* | *Holdemanella biformis* | IL-17A-producing Activated CD69^+^ CD4^+^ T Cells | CD4^+^ T Cells | 9.41E-10 | 1.67E-10 | | 1.16E-04 |
| Coenzyme A biosynthesis I (prokaryotic) | Cofactor & Vitamin Biosynthesis | *Holdemanella* | *Holdemanella biformis* | PD-1^+^ CD8^+^ T Cells | CD8^+^ T Cells | 6.75E-10 | 1.23E-10 | | 1.20E-04 |
| S-adenosyl-L-methionine salvage I | Amino Acid Biosynthesis | *Streptococcus* | *Streptococcus thermophilus* | IFNγ-producing CD161^+^ CD4^+^ T Cells | CD4^+^ T Cells | 3.98E-11 | 6.79E-12 | | 1.21E-04 |
| L-valine biosynthesis | Amino Acid Biosynthesis | *Streptococcus* | *Streptococcus thermophilus* | IFNγ/TNFα-producing CD161^+^ CD8^+^ T Cells | CD8^+^ T Cells | 2.83E-10 | 4.86E-11 | | 1.23E-04 |
| tRNA charging | Amino Acid Biosynthesis | *Holdemanella* | *Holdemanella biformis* | TNFα-producing gd T Cells | Adaptive Immune Cells | 2.32E-10 | 4.21E-11 | | 1.23E-04 |
| L-valine biosynthesis | Amino Acid Biosynthesis | *Streptococcus* | *Streptococcus thermophilus* | IFNγ-producing Activated CD69^+^ CD4^+^ T Cells | CD4^+^ T Cells | 1.58E-11 | 2.74E-12 | | 1.24E-04 |
| Guanosine ribonucleotides de novo biosynthesis | Nucleotide Metabolism & Biosynthesis | *Holdemanella* | *Holdemanella biformis* | IL-17A-producing CD4^+^ T Cells | CD4^+^ T Cells | 8.39E-10 | 1.50E-10 | | 1.24E-04 |
| Queuosine biosynthesis I (de novo) | Nucleotide Metabolism & Biosynthesis | *Coprobacter* | *Coprobacter fastidiosus* | IFNγ/TNFα-producing CD8^+^ T Cells | CD8^+^ T Cells | 6.55E-11 | 1.10E-11 | | 1.26E-04 |
| Chorismate biosynthesis from 3-dehydroquinate | General Biosynthetic Pathways | *Holdemanella* | *Holdemanella biformis* | IL-17A-producing Activated CD69^+^ CD8^+^ T Cells | CD8^+^ T Cells | 2.89E-09 | 5.30E-10 | | 1.38E-04 |
| UDP-N-acetylmuramoyl-pentapeptide biosynthesis II (lysine-containing) | Cell wall & Peptidoglycan Synthesis | *Holdemanella* | *Holdemanella biformis* | IL-17A-producing Activated CD69^+^ CD4^+^ T Cells | CD4^+^ T Cells | 9.85E-10 | 1.77E-10 | | 1.40E-04 |
| Chorismate biosynthesis from 3-dehydroquinate | General Biosynthetic Pathways | *Holdemanella* | *Holdemanella biformis* | PD-1^+^ Activated HLA-DR^+^ CD4^+^ T Cells | CD4^+^ T Cells | 1.25E-09 | 2.29E-10 | | 1.41E-04 |
| 5-aminoimidazole ribonucleotide biosynthesis II | Nucleotide Metabolism & Biosynthesis | *Holdemanella* | *Holdemanella biformis* | TNFα-producing T regs | Adaptive Immune Cells | 8.80E-10 | 1.62E-10 | | 1.42E-04 |
| Superpathway of 5-aminoimidazole ribonucleotide biosynthesis | Nucleotide Metabolism & Biosynthesis | *Holdemanella* | *Holdemanella biformis* | TNFα-producing T regs | Adaptive Immune Cells | 8.80E-10 | 1.62E-10 | | 1.42E-04 |
| tRNA charging | Amino Acid Biosynthesis | *Holdemanella* | *Holdemanella biformis* | PD-1^+^ CD4^+^ T Cells | CD4^+^ T Cells | 2.48E-10 | 4.57E-11 | | 1.48E-04 |
| 5-aminoimidazole ribonucleotide biosynthesis I | Nucleotide Metabolism & Biosynthesis | *Eubacterium* | *Eubacterium ventriosum* | Non-classical Monocytes | Innate Immune Cells | 1.27E-09 | 2.20E-10 | | 1.49E-04 |
| Purine ribonucleosides degradation | Nucleotide Metabolism & Biosynthesis | *Streptococcus* | *Streptococcus thermophilus* | IFNγ/TNFα-producing CD161^+^ CD4^+^ T Cells | CD4^+^ T Cells | 6.37E-11 | 1.09E-11 | | 1.51E-04 |
| 5-aminoimidazole ribonucleotide biosynthesis II | Nucleotide Metabolism & Biosynthesis | *Holdemanella* | *Holdemanella biformis* | PMDSCs | Innate Immune Cells | 1.00E-09 | 1.85E-10 | | 1.52E-04 |
| Superpathway of 5-aminoimidazole ribonucleotide biosynthesis | Nucleotide Metabolism & Biosynthesis | *Holdemanella* | *Holdemanella biformis* | PMDSCs | Innate Immune Cells | 1.00E-09 | 1.85E-10 | | 1.52E-04 |
| Flavin biosynthesis I (bacteria and plants) | Cofactor & Vitamin Biosynthesis | *Holdemanella* | *Holdemanella biformis* | Non-classical Monocytes | Innate Immune Cells | 6.43E-09 | 1.12E-09 | | 1.53E-04 |
| L-valine biosynthesis | Amino Acid Biosynthesis | *Streptococcus* | *Streptococcus thermophilus* | CD8^+^ TEMRA | CD8^+^ T Cells | 1.46E-11 | 2.60E-12 | | 1.57E-04 |
| Methylerythritol phosphate pathway I | General Biosynthetic Pathways | *Holdemanella* | *Holdemanella biformis* | Non-classical Monocytes | Innate Immune Cells | 6.76E-09 | 1.19E-09 | | 1.68E-04 |
| L-valine biosynthesis | Amino Acid Biosynthesis | *Streptococcus* | *Streptococcus thermophilus* | IFNγ-producing CD4^+^ EM T Cells | CD4^+^ T Cells | 3.52E-11 | 6.31E-12 | | 1.70E-04 |
| S-adenosyl-L-methionine salvage I | Amino Acid Biosynthesis | *Streptococcus* | *Streptococcus thermophilus* | IFNγ/TNFα-producing CD161^+^ CD8^+^ T Cells | CD8^+^ T Cells | 3.14E-10 | 5.53E-11 | | 1.70E-04 |
| tRNA charging | Amino Acid Biosynthesis | *Holdemanella* | *Holdemanella biformis* | IL-17A-producing Activated CD69^+^ CD8^+^ T Cells | CD8^+^ T Cells | 2.62E-09 | 4.86E-10 | | 1.73E-04 |
| Superpathway of coenzyme A biosynthesis III (mammals) | Cofactor & Vitamin Biosynthesis | *Holdemanella* | *Holdemanella biformis* | IL-17A-producing Activated CD69^+^ CD4^+^ T Cells | CD4^+^ T Cells | 1.02E-09 | 1.85E-10 | | 1.77E-04 |
| Coenzyme A biosynthesis II (eukaryotic) | Cofactor & Vitamin Biosynthesis | *Holdemanella* | *Holdemanella biformis* | IL-17A-producing Activated CD69^+^ CD4^+^ T Cells | CD4^+^ T Cells | 9.94E-10 | 1.81E-10 | | 1.77E-04 |
| S-adenosyl-L-methionine salvage I | Amino Acid Biosynthesis | *Streptococcus* | *Streptococcus thermophilus* | IFNγ-producing Activated CD69^+^ CD4^+^ T Cells | CD4^+^ T Cells | 1.75E-11 | 3.12E-12 | | 1.80E-04 |
| Superpathway of L-threonine biosynthesis | Amino Acid Biosynthesis | *Holdemanella* | *Holdemanella biformis* | IL-17A-producing Activated CD69^+^ CD4^+^ T Cells | CD4^+^ T Cells | 9.48E-10 | 1.73E-10 | | 1.90E-04 |
| Methylerythritol phosphate pathway I | General Biosynthetic Pathways | *Holdemanella* | *Holdemanella biformis* | IL-17A-producing CD161^+^ CD8^+^ T Cells | CD8^+^ T Cells | 1.08E-08 | 1.93E-09 | | 1.96E-04 |
| Guanosine ribonucleotides de novo biosynthesis | Nucleotide Metabolism & Biosynthesis | *Holdemanella* | *Holdemanella biformis* | TNFα-producing NK Cells | Natural Killer (NK) Cells | 1.19E-09 | 2.18E-10 | | 1.98E-04 |
| 5-aminoimidazole ribonucleotide biosynthesis II | Nucleotide Metabolism & Biosynthesis | *Holdemanella* | *Holdemanella biformis* | TNFα-producing gd T Cells | Adaptive Immune Cells | 3.80E-10 | 7.13E-11 | | 2.13E-04 |
| Superpathway of 5-aminoimidazole ribonucleotide biosynthesis | Nucleotide Metabolism & Biosynthesis | *Holdemanella* | *Holdemanella biformis* | TNFα-producing gd T Cells | Adaptive Immune Cells | 3.80E-10 | 7.13E-11 | | 2.13E-04 |
| Queuosine biosynthesis I (de novo) | Nucleotide Metabolism & Biosynthesis | *Coprobacter* | *Coprobacter fastidiosus* | IFNγ-producing Activated HLA-DR^+^ CD4^+^ T Cells | CD4^+^ T Cells | 1.88E-10 | 3.35E-11 | | 2.15E-04 |
| tRNA charging | Amino Acid Biosynthesis | *Holdemanella* | *Holdemanella biformis* | PD-1^+^ Activated HLA-DR^+^ CD4^+^ T Cells | CD4^+^ T Cells | 1.13E-09 | 2.11E-10 | | 2.21E-04 |
| 5-aminoimidazole ribonucleotide biosynthesis II | Nucleotide Metabolism & Biosynthesis | *Holdemanella* | *Holdemanella biformis* | IL-17A-producing Activated CD69^+^ CD8^+^ T Cells | CD8^+^ T Cells | 4.33E-09 | 8.17E-10 | | 2.27E-04 |
| Superpathway of 5-aminoimidazole ribonucleotide biosynthesis | Nucleotide Metabolism & Biosynthesis | *Holdemanella* | *Holdemanella biformis* | IL-17A-producing Activated CD69^+^ CD8^+^ T Cells | CD8^+^ T Cells | 4.33E-09 | 8.17E-10 | | 2.27E-04 |
| 5-aminoimidazole ribonucleotide biosynthesis II | Nucleotide Metabolism & Biosynthesis | *Holdemanella* | *Holdemanella biformis* | PD-1^+^ CD4^+^ T Cells | CD4^+^ T Cells | 4.07E-10 | 7.71E-11 | | 2.39E-04 |
| Superpathway of 5-aminoimidazole ribonucleotide biosynthesis | Nucleotide Metabolism & Biosynthesis | *Holdemanella* | *Holdemanella biformis* | PD-1^+^ CD4^+^ T Cells | CD4^+^ T Cells | 4.07E-10 | 7.71E-11 | | 2.39E-04 |
| 5-aminoimidazole ribonucleotide biosynthesis II | Nucleotide Metabolism & Biosynthesis | *Eubacterium* | *Eubacterium ventriosum* | Non-classical Monocytes | Innate Immune Cells | 1.17E-09 | 2.13E-10 | | 2.61E-04 |
| Superpathway of 5-aminoimidazole ribonucleotide biosynthesis | Nucleotide Metabolism & Biosynthesis | *Eubacterium* | *Eubacterium ventriosum* | Non-classical Monocytes | Innate Immune Cells | 1.17E-09 | 2.13E-10 | | 2.61E-04 |
| Queuosine biosynthesis I (de novo) | Nucleotide Metabolism & Biosynthesis | *Coprobacter* | *Coprobacter fastidiosus* | CD4^+^ TEMRA | CD4^+^ T Cells | 7.21E-11 | 1.32E-11 | | 2.61E-04 |
| Methylerythritol phosphate pathway I | General Biosynthetic Pathways | *Holdemanella* | *Holdemanella biformis* | IL-17A-producing Activated NK Cells | Natural Killer (NK) Cells | 1.66E-08 | 3.04E-09 | | 2.65E-04 |
| Glycogen biosynthesis I (from ADP-D-Glucose) | General Biosynthetic Pathways | *Holdemanella* | *Holdemanella biformis* | Mature LDNs | Innate Immune Cells | 1.62E-09 | 3.06E-10 | | 2.77E-04 |
| L-valine biosynthesis | Amino Acid Biosynthesis | *Streptococcus* | *Streptococcus thermophilus* | IFNγ/TNFα-producing CD161^+^ CD4^+^ T Cells | CD4^+^ T Cells | 5.94E-11 | 1.05E-11 | | 2.82E-04 |
| S-adenosyl-L-methionine salvage I | Amino Acid Biosynthesis | *Streptococcus* | *Streptococcus thermophilus* | IFNγ-producing CD4^+^ EM T Cells | CD4^+^ T Cells | 3.88E-11 | 7.19E-12 | | 2.94E-04 |
| Sucrose biosynthesis II | General Biosynthetic Pathways | *Holdemanella* | *Holdemanella biformis* | Mature LDNs | Innate Immune Cells | 1.42E-09 | 2.69E-10 | | 2.98E-04 |
| 5-aminoimidazole ribonucleotide biosynthesis I | Nucleotide Metabolism & Biosynthesis | *Holdemanella* | *Holdemanella biformis* | TNFα-producing T regs | Adaptive Immune Cells | 7.95E-10 | 1.53E-10 | | 2.99E-04 |
| 5-aminoimidazole ribonucleotide biosynthesis I | Nucleotide Metabolism & Biosynthesis | *Holdemanella* | *Holdemanella biformis* | PMDSCs | Innate Immune Cells | 9.07E-10 | 1.75E-10 | | 3.19E-04 |
| UMP biosynthesis I | Nucleotide Metabolism & Biosynthesis | *Holdemanella* | *Holdemanella biformis* | IL-17A-producing CD4^+^ T Cells | CD4^+^ T Cells | 1.22E-09 | 2.30E-10 | | 3.20E-04 |
| UMP biosynthesis II | Nucleotide Metabolism & Biosynthesis | *Holdemanella* | *Holdemanella biformis* | IL-17A-producing CD4^+^ T Cells | CD4^+^ T Cells | 1.22E-09 | 2.30E-10 | | 3.20E-04 |
| UMP biosynthesis III | Nucleotide Metabolism & Biosynthesis | *Holdemanella* | *Holdemanella biformis* | IL-17A-producing CD4^+^ T Cells | CD4^+^ T Cells | 1.22E-09 | 2.30E-10 | | 3.20E-04 |
| 5-aminoimidazole ribonucleotide biosynthesis II | Nucleotide Metabolism & Biosynthesis | *Holdemanella* | *Holdemanella biformis* | PD-1^+^ Activated HLA-DR^+^ CD4^+^ T Cells | CD4^+^ T Cells | 1.85E-09 | 3.56E-10 | | 3.33E-04 |
| Superpathway of 5-aminoimidazole ribonucleotide biosynthesis | Nucleotide Metabolism & Biosynthesis | *Holdemanella* | *Holdemanella biformis* | PD-1^+^ Activated HLA-DR^+^ CD4^+^ T Cells | CD4^+^ T Cells | 1.85E-09 | 3.56E-10 | | 3.33E-04 |
| S-adenosyl-L-methionine salvage I | Amino Acid Biosynthesis | *Streptococcus* | *Streptococcus thermophilus* | CD8^+^ TEMRA | CD8^+^ T Cells | 1.60E-11 | 2.98E-12 | | 3.48E-04 |
| Guanosine ribonucleotides de novo biosynthesis | Nucleotide Metabolism & Biosynthesis | *Holdemanella* | *Holdemanella biformis* | TNFα-producing CD161^+^ NK Cells | Natural Killer (NK) Cells | 1.11E-09 | 2.12E-10 | | 3.77E-04 |
| Chorismate biosynthesis I | General Biosynthetic Pathways | *Holdemanella* | *Holdemanella biformis* | PD-1^+^ CD8^+^ T Cells | CD8^+^ T Cells | 6.30E-10 | 1.23E-10 | | 3.92E-04 |
| Purine ribonucleosides degradation | Nucleotide Metabolism & Biosynthesis | *Streptococcus* | *Streptococcus thermophilus* | IFNγ/TNFα-producing CD8^+^ T Cells | CD8^+^ T Cells | 4.55E-11 | 8.38E-12 | | 4.10E-04 |
| 5-aminoimidazole ribonucleotide biosynthesis I | Nucleotide Metabolism & Biosynthesis | *Holdemanella* | *Holdemanella biformis* | TNFα-producing gd T Cells | Adaptive Immune Cells | 3.43E-10 | 6.74E-11 | | 4.23E-04 |
| Peptidoglycan biosynthesis I (meso-diaminopimelate containing) | Cell wall & Peptidoglycan Synthesis | *Holdemanella* | *Holdemanella biformis* | CCR7^+^ Activated NK Cells | Natural Killer (NK) Cells | 4.88E-09 | 9.40E-10 | | 4.25E-04 |
| UDP-N-acetylmuramoyl-pentapeptide biosynthesis I (meso-diaminopimelate containing) | Cell wall & Peptidoglycan Synthesis | *Holdemanella* | *Holdemanella biformis* | CCR7^+^ Activated NK Cells | Natural Killer (NK) Cells | 4.79E-09 | 9.24E-10 | | 4.28E-04 |
| Glycogen biosynthesis I (from ADP-D-Glucose) | General Biosynthetic Pathways | *Holdemanella* | *Holdemanella biformis* | TNFα-producing T regs | Adaptive Immune Cells | 5.97E-10 | 1.18E-10 | | 4.32E-04 |
| Purine ribonucleosides degradation | Nucleotide Metabolism & Biosynthesis | *Streptococcus* | *Streptococcus thermophilus* | IFNγ-producing Activated HLA-DR^+^ CD4^+^ T Cells | CD4^+^ T Cells | 1.33E-10 | 2.50E-11 | | 4.38E-04 |
| Queuosine biosynthesis I (de novo) | Nucleotide Metabolism & Biosynthesis | *Coprobacter* | *Coprobacter fastidiosus* | IFNγ-producing Activated CD69^+^ CD8^+^ T Cells | CD8^+^ T Cells | 1.72E-11 | 3.10E-12 | | 4.44E-04 |
| Chorismate biosynthesis from 3-dehydroquinate | General Biosynthetic Pathways | *Holdemanella* | *Holdemanella biformis* | PD-1^+^ CD8^+^ T Cells | CD8^+^ T Cells | 5.73E-10 | 1.13E-10 | | 4.72E-04 |
| Sucrose biosynthesis II | General Biosynthetic Pathways | *Holdemanella* | *Holdemanella biformis* | TNFα-producing T regs | Adaptive Immune Cells | 5.21E-10 | 1.04E-10 | | 4.80E-04 |
| 5-aminoimidazole ribonucleotide biosynthesis I | Nucleotide Metabolism & Biosynthesis | *Holdemanella* | *Holdemanella biformis* | PD-1^+^ CD4^+^ T Cells | CD4^+^ T Cells | 3.68E-10 | 7.30E-11 | | 4.80E-04 |
| Glycogen biosynthesis I (from ADP-D-Glucose) | General Biosynthetic Pathways | *Holdemanella* | *Holdemanella biformis* | PMDSCs | Innate Immune Cells | 6.79E-10 | 1.35E-10 | | 4.94E-04 |
| 5-aminoimidazole ribonucleotide biosynthesis I | Nucleotide Metabolism & Biosynthesis | *Holdemanella* | *Holdemanella biformis* | IL-17A-producing Activated CD69^+^ CD8^+^ T Cells | CD8^+^ T Cells | 3.90E-09 | 7.74E-10 | | 5.03E-04 |
| UDP-N-acetylmuramoyl-pentapeptide biosynthesis II (lysine-containing) | Cell wall & Peptidoglycan Synthesis | *Holdemanella* | *Holdemanella biformis* | CCR7^+^ Activated NK Cells | Natural Killer (NK) Cells | 5.01E-09 | 9.77E-10 | | 5.03E-04 |
| Glycogen biosynthesis I (from ADP-D-Glucose) | General Biosynthetic Pathways | *Holdemanella* | *Holdemanella biformis* | TNFα-producing gd T Cells | Adaptive Immune Cells | 2.59E-10 | 5.16E-11 | | 5.14E-04 |
| UDP-N-acetylmuramoyl-pentapeptide biosynthesis II (lysine-containing) | Cell wall & Peptidoglycan Synthesis | *Holdemanella* | *Holdemanella biformis* | IL-17A-producing CD161^+^ CD8^+^ T Cells | CD8^+^ T Cells | 9.86E-09 | 1.86E-09 | | 5.20E-04 |
| Sucrose biosynthesis II | General Biosynthetic Pathways | *Holdemanella* | *Holdemanella biformis* | PMDSCs | Innate Immune Cells | 5.92E-10 | 1.19E-10 | | 5.48E-04 |
| Queuosine biosynthesis I (de novo) | Nucleotide Metabolism & Biosynthesis | *Coprobacter* | *Coprobacter fastidiosus* | IFNγ-producing CD8^+^ T Cells | CD8^+^ T Cells | 1.62E-11 | 2.95E-12 | | 5.55E-04 |
| Coenzyme A biosynthesis II (eukaryotic) | Cofactor & Vitamin Biosynthesis | *Coprobacter* | *Coprobacter fastidiosus* | IFNγ-producing CD161^+^ NK Cells | Natural Killer (NK) Cells | 3.78E-10 | 6.02E-11 | | 5.58E-04 |
| S-adenosyl-L-methionine salvage I | Amino Acid Biosynthesis | *Streptococcus* | *Streptococcus thermophilus* | IFNγ/TNFα-producing CD161^+^ CD4^+^ T Cells | CD4^+^ T Cells | 6.51E-11 | 1.20E-11 | | 5.62E-04 |
| Coenzyme A biosynthesis I (prokaryotic) | Cofactor & Vitamin Biosynthesis | *Holdemanella* | *Holdemanella biformis* | IL-17A-producing CD4^+^ T Cells | CD4^+^ T Cells | 9.69E-10 | 1.92E-10 | | 6.03E-04 |
| UMP biosynthesis I | Nucleotide Metabolism & Biosynthesis | *Holdemanella* | *Holdemanella biformis* | TNFα-producing NK Cells | Natural Killer (NK) Cells | 1.70E-09 | 3.36E-10 | | 6.23E-04 |
| UMP biosynthesis II | Nucleotide Metabolism & Biosynthesis | *Holdemanella* | *Holdemanella biformis* | TNFα-producing NK Cells | Natural Killer (NK) Cells | 1.70E-09 | 3.36E-10 | | 6.23E-04 |
| UMP biosynthesis III | Nucleotide Metabolism & Biosynthesis | *Holdemanella* | *Holdemanella biformis* | TNFα-producing NK Cells | Natural Killer (NK) Cells | 1.70E-09 | 3.36E-10 | | 6.23E-04 |
| S-adenosyl-L-methionine salvage I | Amino Acid Biosynthesis | *Holdemanella* | *Holdemanella biformis* | TNFα-producing T regs | Adaptive Immune Cells | 7.18E-10 | 1.46E-10 | | 6.45E-04 |
| L-valine biosynthesis | Amino Acid Biosynthesis | *Streptococcus* | *Streptococcus thermophilus* | IFNγ/TNFα-producing CD8^+^ T Cells | CD8^+^ T Cells | 4.24E-11 | 8.08E-12 | | 6.54E-04 |
| Queuosine biosynthesis I (de novo) | Nucleotide Metabolism & Biosynthesis | *Coprobacter* | *Coprobacter fastidiosus* | TNFα-producing Activated CD69^+^ CD8^+^ T Cells | CD8^+^ T Cells | 4.51E-11 | 8.49E-12 | | 6.71E-04 |
| Glycogen biosynthesis I (from ADP-D-Glucose) | General Biosynthetic Pathways | *Holdemanella* | *Holdemanella biformis* | PD-1^+^ CD4^+^ T Cells | CD4^+^ T Cells | 2.76E-10 | 5.61E-11 | | 6.74E-04 |
| Purine ribonucleosides degradation | Nucleotide Metabolism & Biosynthesis | *Streptococcus* | *Streptococcus thermophilus* | IFNγ/TNFα-producing CD69^+^ CD8^+^ T Cells | CD8^+^ T Cells | 1.16E-10 | 2.21E-11 | | 6.85E-04 |
| Sucrose biosynthesis II | General Biosynthetic Pathways | *Holdemanella* | *Holdemanella biformis* | TNFα-producing gd T Cells | Adaptive Immune Cells | 2.24E-10 | 4.56E-11 | | 6.86E-04 |
| Peptidoglycan biosynthesis I (meso-diaminopimelate containing) | Cell wall & Peptidoglycan Synthesis | *Holdemanella* | *Holdemanella biformis* | IL-17A-producing CD161^+^ CD8^+^ T Cells | CD8^+^ T Cells | 9.43E-09 | 1.81E-09 | | 6.87E-04 |
| Queuosine biosynthesis I (de novo) | Nucleotide Metabolism & Biosynthesis | *Coprobacter* | *Coprobacter fastidiosus* | IFNγ/TNFα-producing HLA-DR^+^ CD8^+^ T Cells | CD8^+^ T Cells | 5.47E-10 | 1.04E-10 | | 6.94E-04 |
| UDP-N-acetylmuramoyl-pentapeptide biosynthesis I (meso-diaminopimelate containing) | Cell wall & Peptidoglycan Synthesis | *Holdemanella* | *Holdemanella biformis* | IL-17A-producing CD161^+^ CD8^+^ T Cells | CD8^+^ T Cells | 9.26E-09 | 1.78E-09 | | 6.94E-04 |
| S-adenosyl-L-methionine salvage I | Amino Acid Biosynthesis | *Holdemanella* | *Holdemanella biformis* | PMDSCs | Innate Immune Cells | 8.18E-10 | 1.67E-10 | | 6.99E-04 |
| 5-aminoimidazole ribonucleotide biosynthesis I | Nucleotide Metabolism & Biosynthesis | *Holdemanella* | *Holdemanella biformis* | PD-1^+^ Activated HLA-DR^+^ CD4^+^ T Cells | CD4^+^ T Cells | 1.67E-09 | 3.37E-10 | | 7.13E-04 |
| Coenzyme A biosynthesis II (eukaryotic) | Cofactor & Vitamin Biosynthesis | *Holdemanella* | *Holdemanella biformis* | CCR7^+^ Activated NK Cells | Natural Killer (NK) Cells | 5.02E-09 | 1.00E-09 | | 7.23E-04 |
| tRNA charging | Amino Acid Biosynthesis | *Holdemanella* | *Holdemanella biformis* | PD-1^+^ CD8^+^ T Cells | CD8^+^ T Cells | 5.14E-10 | 1.05E-10 | | 7.30E-04 |
| Sucrose biosynthesis II | General Biosynthetic Pathways | *Holdemanella* | *Holdemanella biformis* | PD-1^+^ CD4^+^ T Cells | CD4^+^ T Cells | 2.41E-10 | 4.93E-11 | | 7.32E-04 |
| Coenzyme A biosynthesis I (prokaryotic) | Cofactor & Vitamin Biosynthesis | *Coprobacter* | *Coprobacter fastidiosus* | IFNγ-producing CD161^+^ NK Cells | Natural Killer (NK) Cells | 3.93E-10 | 6.66E-11 | | 7.34E-04 |
| Folate transformations II (plants) | Cofactor & Vitamin Biosynthesis | *Coprobacter* | *Coprobacter fastidiosus* | IFNγ-producing CD161^+^ NK Cells | Natural Killer (NK) Cells | 3.55E-10 | 6.03E-11 | | 7.34E-04 |
| Peptidoglycan biosynthesis III (mycobacteria) | Cell wall & Peptidoglycan Synthesis | *Coprobacter* | *Coprobacter fastidiosus* | IFNγ-producing CD161^+^ NK Cells | Natural Killer (NK) Cells | 3.87E-10 | 6.59E-11 | | 7.34E-04 |
| S-adenosyl-L-methionine salvage I | Amino Acid Biosynthesis | *Holdemanella* | *Holdemanella biformis* | IL-17A-producing Activated CD69^+^ CD8^+^ T Cells | CD8^+^ T Cells | 3.58E-09 | 7.28E-10 | | 7.35E-04 |
| Superpathway of coenzyme A biosynthesis III (mammals) | Cofactor & Vitamin Biosynthesis | *Holdemanella* | *Holdemanella biformis* | CCR7^+^ Activated NK Cells | Natural Killer (NK) Cells | 5.13E-09 | 1.03E-09 | | 7.40E-04 |
| Purine ribonucleosides degradation | Nucleotide Metabolism & Biosynthesis | *Streptococcus* | *Streptococcus thermophilus* | IFNγ/TNFα-producing HLA-DR^+^ CD8^+^ T Cells | CD8^+^ T Cells | 3.98E-10 | 7.64E-11 | | 7.43E-04 |
| Guanosine ribonucleotides de novo biosynthesis | Nucleotide Metabolism & Biosynthesis | *Holdemanella* | *Holdemanella biformis* | IL-17A-producing Activated CD69^+^ CD4^+^ T Cells | CD4^+^ T Cells | 8.31E-10 | 1.66E-10 | | 7.57E-04 |
| L-valine biosynthesis | Amino Acid Biosynthesis | *Streptococcus* | *Streptococcus thermophilus* | IFNγ-producing Activated HLA-DR^+^ CD4^+^ T Cells | CD4^+^ T Cells | 1.24E-10 | 2.42E-11 | | 7.99E-04 |
| Queuosine biosynthesis I (de novo) | Nucleotide Metabolism & Biosynthesis | *Coprobacter* | *Coprobacter fastidiosus* | TNFα-producing CD8^+^ T Cells | CD8^+^ T Cells | 5.57E-11 | 1.07E-11 | | 8.07E-04 |
| S-adenosyl-L-methionine salvage I | Amino Acid Biosynthesis | *Streptococcus* | *Streptococcus thermophilus* | IFNγ/TNFα-producing CD8^+^ T Cells | CD8^+^ T Cells | 4.70E-11 | 9.15E-12 | | 8.20E-04 |
| Coenzyme A biosynthesis I (prokaryotic) | Cofactor & Vitamin Biosynthesis | *Holdemanella* | *Holdemanella biformis* | TNFα-producing NK Cells | Natural Killer (NK) Cells | 1.37E-09 | 2.78E-10 | | 8.59E-04 |
| Inosine 5-phosphate degradation | Nucleotide Metabolism & Biosynthesis | *Coprobacter* | *Coprobacter fastidiosus* | IFNγ-producing CD161^+^ NK Cells | Natural Killer (NK) Cells | 3.18E-10 | 5.55E-11 | | 8.91E-04 |
| UDP-N-acetylmuramoyl-pentapeptide biosynthesis I (meso-diaminopimelate containing) | Cell wall & Peptidoglycan Synthesis | *Coprobacter* | *Coprobacter fastidiosus* | IFNγ-producing CD161^+^ NK Cells | Natural Killer (NK) Cells | 3.71E-10 | 6.45E-11 | | 8.91E-04 |
| Superpathway of L-threonine biosynthesis | Amino Acid Biosynthesis | *Holdemanella* | *Holdemanella biformis* | CCR7^+^ Activated NK Cells | Natural Killer (NK) Cells | 4.75E-09 | 9.66E-10 | | 9.18E-04 |
| S-adenosyl-L-methionine salvage I | Amino Acid Biosynthesis | *Holdemanella* | *Holdemanella biformis* | TNFα-producing gd T Cells | Adaptive Immune Cells | 3.09E-10 | 6.41E-11 | | 9.26E-04 |
| Chorismate biosynthesis I | General Biosynthetic Pathways | *Holdemanella* | *Holdemanella biformis* | IL-17A-producing CD4^+^ T Cells | CD4^+^ T Cells | 9.29E-10 | 1.89E-10 | | 9.29E-04 |
| UMP biosynthesis I | Nucleotide Metabolism & Biosynthesis | *Dialister* | *Dialister invisus* | IL-2/TNFα-producing CD161^+^ CD4^+^ T Cells | CD4^+^ T Cells | 4.94E-10 | 8.13E-11 | | 9.36E-04 |
| 5-aminoimidazole ribonucleotide biosynthesis I | Nucleotide Metabolism & Biosynthesis | *Dialister* | *Dialister invisus* | IL-2/TNFα-producing CD161^+^ CD4^+^ T Cells | CD4^+^ T Cells | 5.72E-10 | 1.01E-10 | | 9.36E-04 |
| 5-aminoimidazole ribonucleotide biosynthesis II | Nucleotide Metabolism & Biosynthesis | *Dialister* | *Dialister invisus* | IL-2/TNFα-producing CD161^+^ CD4^+^ T Cells | CD4^+^ T Cells | 5.89E-10 | 1.05E-10 | | 9.36E-04 |
| Superpathway of 5-aminoimidazole ribonucleotide biosynthesis | Nucleotide Metabolism & Biosynthesis | *Dialister* | *Dialister invisus* | IL-2/TNFα-producing CD161^+^ CD4^+^ T Cells | CD4^+^ T Cells | 5.89E-10 | 1.05E-10 | | 9.36E-04 |
| UMP biosynthesis II | Nucleotide Metabolism & Biosynthesis | *Dialister* | *Dialister invisus* | IL-2/TNFα-producing CD161^+^ CD4^+^ T Cells | CD4^+^ T Cells | 4.94E-10 | 8.13E-11 | | 9.36E-04 |
| UMP biosynthesis III | Nucleotide Metabolism & Biosynthesis | *Dialister* | *Dialister invisus* | IL-2/TNFα-producing CD161^+^ CD4^+^ T Cells | CD4^+^ T Cells | 4.94E-10 | 8.13E-11 | | 9.36E-04 |
| Coenzyme A biosynthesis I (prokaryotic) | Cofactor & Vitamin Biosynthesis | *Dialister* | *Dialister invisus* | IL-2/TNFα-producing CD161^+^ CD4^+^ T Cells | CD4^+^ T Cells | 4.76E-10 | 8.72E-11 | | 9.48E-04 |
| Superpathway of coenzyme A biosynthesis III (mammals) | Cofactor & Vitamin Biosynthesis | *Dialister* | *Dialister invisus* | IL-2/TNFα-producing CD161^+^ CD4^+^ T Cells | CD4^+^ T Cells | 4.51E-10 | 8.49E-11 | | 9.48E-04 |
| Folate transformations II (plants) | Cofactor & Vitamin Biosynthesis | *Dialister* | *Dialister invisus* | IL-2/TNFα-producing CD161^+^ CD4^+^ T Cells | CD4^+^ T Cells | 4.79E-10 | 8.72E-11 | | 9.48E-04 |
| Guanosine ribonucleotides de novo biosynthesis | Nucleotide Metabolism & Biosynthesis | *Dialister* | *Dialister invisus* | IL-2/TNFα-producing CD161^+^ CD4^+^ T Cells | CD4^+^ T Cells | 5.03E-10 | 9.23E-11 | | 9.48E-04 |
| Coenzyme A biosynthesis II (eukaryotic) | Cofactor & Vitamin Biosynthesis | *Dialister* | *Dialister invisus* | IL-2/TNFα-producing CD161^+^ CD4^+^ T Cells | CD4^+^ T Cells | 4.38E-10 | 8.23E-11 | | 9.48E-04 |
| S-adenosyl-L-methionine salvage I | Amino Acid Biosynthesis | *Holdemanella* | *Holdemanella biformis* | PD-1^+^ CD4^+^ T Cells | CD4^+^ T Cells | 3.32E-10 | 6.93E-11 | | 9.67E-04 |
| Glycogen biosynthesis I (from ADP-D-Glucose) | General Biosynthetic Pathways | *Holdemanella* | *Holdemanella biformis* | PD-1^+^ Activated HLA-DR^+^ CD4^+^ T Cells | CD4^+^ T Cells | 1.25E-09 | 2.59E-10 | | 9.89E-04 |
| L-valine biosynthesis | Amino Acid Biosynthesis | *Streptococcus* | *Streptococcus thermophilus* | IFNγ/TNFα-producing CD69^+^ CD8^+^ T Cells | CD8^+^ T Cells | 1.09E-10 | 2.13E-11 | | 0.00101 |
| 5-aminoimidazole ribonucleotide biosynthesis II | Nucleotide Metabolism & Biosynthesis | *Holdemanella* | *Holdemanella biformis* | PD-1^+^ CD8^+^ T Cells | CD8^+^ T Cells | 8.46E-10 | 1.76E-10 | | 0.00101 |
| Superpathway of 5-aminoimidazole ribonucleotide biosynthesis | Nucleotide Metabolism & Biosynthesis | *Holdemanella* | *Holdemanella biformis* | PD-1^+^ CD8^+^ T Cells | CD8^+^ T Cells | 8.46E-10 | 1.76E-10 | | 0.00101 |
| UMP biosynthesis I | Nucleotide Metabolism & Biosynthesis | *Eubacterium* | *Eubacterium ventriosum* | Non-classical Monocytes | Innate Immune Cells | 9.23E-10 | 1.85E-10 | | 0.00102 |
| UMP biosynthesis II | Nucleotide Metabolism & Biosynthesis | *Eubacterium* | *Eubacterium ventriosum* | Non-classical Monocytes | Innate Immune Cells | 9.23E-10 | 1.85E-10 | | 0.00102 |
| UMP biosynthesis III | Nucleotide Metabolism & Biosynthesis | *Eubacterium* | *Eubacterium ventriosum* | Non-classical Monocytes | Innate Immune Cells | 9.23E-10 | 1.85E-10 | | 0.00102 |
| Purine ribonucleosides degradation | Nucleotide Metabolism & Biosynthesis | *Streptococcus* | *Streptococcus thermophilus* | CD4^+^ TEMRA | CD4^+^ T Cells | 4.96E-11 | 1.00E-11 | | 0.00104 |
| Sucrose biosynthesis II | General Biosynthetic Pathways | *Holdemanella* | *Holdemanella biformis* | PD-1^+^ Activated HLA-DR^+^ CD4^+^ T Cells | CD4^+^ T Cells | 1.09E-09 | 2.28E-10 | | 0.00106 |
| S-adenosyl-L-methionine salvage I | Amino Acid Biosynthesis | *Streptococcus* | *Streptococcus thermophilus* | IFNγ/TNFα-producing CD69^+^ CD8^+^ T Cells | CD8^+^ T Cells | 1.22E-10 | 2.39E-11 | | 0.00109 |
| Glycogen biosynthesis I (from ADP-D-Glucose) | General Biosynthetic Pathways | *Holdemanella* | *Holdemanella biformis* | IL-17A-producing Activated CD69^+^ CD8^+^ T Cells | CD8^+^ T Cells | 2.88E-09 | 6.01E-10 | | 0.00109 |
| UMP biosynthesis I | Nucleotide Metabolism & Biosynthesis | *Holdemanella* | *Holdemanella biformis* | TNFα-producing CD161^+^ NK Cells | Natural Killer (NK) Cells | 1.59E-09 | 3.25E-10 | | 0.00110 |
| UMP biosynthesis II | Nucleotide Metabolism & Biosynthesis | *Holdemanella* | *Holdemanella biformis* | TNFα-producing CD161^+^ NK Cells | Natural Killer (NK) Cells | 1.59E-09 | 3.25E-10 | | 0.00110 |
| UMP biosynthesis III | Nucleotide Metabolism & Biosynthesis | *Holdemanella* | *Holdemanella biformis* | TNFα-producing CD161^+^ NK Cells | Natural Killer (NK) Cells | 1.59E-09 | 3.25E-10 | | 0.00110 |
| Queuosine biosynthesis I (de novo) | Nucleotide Metabolism & Biosynthesis | *Coprobacter* | *Coprobacter fastidiosus* | TNFα-producing Activated HLA-DR^+^ CD8^+^ T Cells | CD8^+^ T Cells | 1.69E-10 | 3.36E-11 | | 0.00111 |
| Purine ribonucleosides degradation | Nucleotide Metabolism & Biosynthesis | *Streptococcus* | *Streptococcus thermophilus* | TNFα-producing Activated HLA-DR^+^ CD8^+^ T Cells | CD8^+^ T Cells | 1.23E-10 | 2.45E-11 | | 0.00111 |
| L-valine biosynthesis | Amino Acid Biosynthesis | *Streptococcus* | *Streptococcus thermophilus* | IFNγ/TNFα-producing HLA-DR^+^ CD8^+^ T Cells | CD8^+^ T Cells | 3.70E-10 | 7.37E-11 | | 0.00118 |
| UDP-N-acetylmuramoyl-pentapeptide biosynthesis II (lysine-containing) | Cell wall & Peptidoglycan Synthesis | *Holdemanella* | *Holdemanella biformis* | IL-17A-producing CD8^+^ T Cells | CD8^+^ T Cells | 2.65E-09 | 5.04E-10 | | 0.00118 |
| Folate transformations II (plants) | Cofactor & Vitamin Biosynthesis | *Eubacterium* | *Eubacterium ventriosum* | Non-classical Monocytes | Innate Immune Cells | 9.50E-10 | 1.93E-10 | | 0.00121 |
| S-adenosyl-L-methionine salvage I | Amino Acid Biosynthesis | *Holdemanella* | *Holdemanella biformis* | PD-1^+^ Activated HLA-DR^+^ CD4^+^ T Cells | CD4^+^ T Cells | 1.51E-09 | 3.19E-10 | | 0.00122 |
| Peptidoglycan biosynthesis I (meso-diaminopimelate containing) | Cell wall & Peptidoglycan Synthesis | *Dialister* | *Dialister invisus* | IL-2/TNFα-producing CD161^+^ CD4^+^ T Cells | CD4^+^ T Cells | 4.39E-10 | 8.55E-11 | | 0.00123 |
| Inosine 5-phosphate degradation | Nucleotide Metabolism & Biosynthesis | *Dialister* | *Dialister invisus* | IL-2/TNFα-producing CD161^+^ CD4^+^ T Cells | CD4^+^ T Cells | 4.15E-10 | 8.07E-11 | | 0.00123 |
| Purine ribonucleosides degradation | Nucleotide Metabolism & Biosynthesis | *Streptococcus* | *Streptococcus thermophilus* | IFNγ-producing Activated CD69^+^ CD8^+^ T Cells | CD8^+^ T Cells | 1.20E-11 | 2.34E-12 | | 0.00124 |
| UMP biosynthesis I | Nucleotide Metabolism & Biosynthesis | *Holdemanella* | *Holdemanella biformis* | IL-17A-producing CD161^+^ CD8^+^ T Cells | CD8^+^ T Cells | 1.28E-08 | 2.57E-09 | | 0.00126 |
| UMP biosynthesis II | Nucleotide Metabolism & Biosynthesis | *Holdemanella* | *Holdemanella biformis* | IL-17A-producing CD161^+^ CD8^+^ T Cells | CD8^+^ T Cells | 1.28E-08 | 2.57E-09 | | 0.00126 |
| UMP biosynthesis III | Nucleotide Metabolism & Biosynthesis | *Holdemanella* | *Holdemanella biformis* | IL-17A-producing CD161^+^ CD8^+^ T Cells | CD8^+^ T Cells | 1.28E-08 | 2.57E-09 | | 0.00126 |
| CDP diacylglycerol biosynthesis I | Lipid & Membrane Metabolism | *Dialister* | *Dialister invisus* | IL-2/TNFα-producing CD161^+^ CD4^+^ T Cells | CD4^+^ T Cells | 4.93E-10 | 9.65E-11 | | 0.00127 |
| CDP diacylglycerol biosynthesis II | Lipid & Membrane Metabolism | *Dialister* | *Dialister invisus* | IL-2/TNFα-producing CD161^+^ CD4^+^ T Cells | CD4^+^ T Cells | 4.93E-10 | 9.65E-11 | | 0.00127 |
| Superpathway of coenzyme A biosynthesis III (mammals) | Cofactor & Vitamin Biosynthesis | *Holdemanella* | *Holdemanella biformis* | IL-17A-producing CD161^+^ CD8^+^ T Cells | CD8^+^ T Cells | 9.89E-09 | 1.99E-09 | | 0.00128 |
| Chorismate biosynthesis from 3-dehydroquinate | General Biosynthetic Pathways | *Holdemanella* | *Holdemanella biformis* | IL-17A-producing CD4^+^ T Cells | CD4^+^ T Cells | 8.39E-10 | 1.75E-10 | | 0.00130 |
| Queuosine biosynthesis I (de novo) | Nucleotide Metabolism & Biosynthesis | *Coprobacter* | *Coprobacter fastidiosus* | IFNγ-producing Activated HLA-DR^+^ CD8^+^ T Cells | CD8^+^ T Cells | 6.13E-11 | 1.21E-11 | | 0.00131 |
| Purine ribonucleosides degradation | Nucleotide Metabolism & Biosynthesis | *Streptococcus* | *Streptococcus thermophilus* | IFNγ-producing Activated HLA-DR^+^ CD8^+^ T Cells | CD8^+^ T Cells | 4.47E-11 | 8.83E-12 | | 0.00132 |
| UMP biosynthesis I | Nucleotide Metabolism & Biosynthesis | *Holdemanella* | *Holdemanella biformis* | IL-17A-producing CD8^+^ T Cells | CD8^+^ T Cells | 3.54E-09 | 6.83E-10 | | 0.00133 |
| UMP biosynthesis II | Nucleotide Metabolism & Biosynthesis | *Holdemanella* | *Holdemanella biformis* | IL-17A-producing CD8^+^ T Cells | CD8^+^ T Cells | 3.54E-09 | 6.83E-10 | | 0.00133 |
| UMP biosynthesis III | Nucleotide Metabolism & Biosynthesis | *Holdemanella* | *Holdemanella biformis* | IL-17A-producing CD8^+^ T Cells | CD8^+^ T Cells | 3.54E-09 | 6.83E-10 | | 0.00133 |
| UDP-N-acetylmuramoyl-pentapeptide biosynthesis I (meso-diaminopimelate containing) | Cell wall & Peptidoglycan Synthesis | *Dialister* | *Dialister invisus* | IL-2/TNFα-producing CD161^+^ CD4^+^ T Cells | CD4^+^ T Cells | 4.51E-10 | 8.89E-11 | | 0.00134 |
| Superpathway of L-threonine biosynthesis | Amino Acid Biosynthesis | *Dialister* | *Dialister invisus* | IL-2/TNFα-producing CD161^+^ CD4^+^ T Cells | CD4^+^ T Cells | 3.99E-10 | 7.87E-11 | | 0.00134 |
| Sucrose biosynthesis II | General Biosynthetic Pathways | *Holdemanella* | *Holdemanella biformis* | IL-17A-producing Activated CD69^+^ CD8^+^ T Cells | CD8^+^ T Cells | 2.50E-09 | 5.29E-10 | | 0.00136 |
| Methylerythritol phosphate pathway I | General Biosynthetic Pathways | *Holdemanella* | *Holdemanella biformis* | IL-17A-producing CD8^+^ T Cells | CD8^+^ T Cells | 2.78E-09 | 5.39E-10 | | 0.00140 |
| Coenzyme A biosynthesis I (prokaryotic) | Cofactor & Vitamin Biosynthesis | *Holdemanella* | *Holdemanella biformis* | TNFα-producing CD161^+^ NK Cells | Natural Killer (NK) Cells | 1.28E-09 | 2.68E-10 | | 0.00147 |
| Peptidoglycan biosynthesis I (meso-diaminopimelate containing) | Cell wall & Peptidoglycan Synthesis | *Holdemanella* | *Holdemanella biformis* | IL-17A-producing CD8^+^ T Cells | CD8^+^ T Cells | 2.52E-09 | 4.92E-10 | | 0.00147 |
| UDP-N-acetylmuramoyl-pentapeptide biosynthesis I (meso-diaminopimelate containing) | Cell wall & Peptidoglycan Synthesis | *Holdemanella* | *Holdemanella biformis* | IL-17A-producing CD8^+^ T Cells | CD8^+^ T Cells | 2.48E-09 | 4.84E-10 | | 0.00148 |
| Peptidoglycan biosynthesis III (mycobacteria) | Cell wall & Peptidoglycan Synthesis | *Dialister* | *Dialister invisus* | IL-2/TNFα-producing CD161^+^ CD4^+^ T Cells | CD4^+^ T Cells | 4.38E-10 | 8.73E-11 | | 0.00152 |
| Purine ribonucleosides degradation | Nucleotide Metabolism & Biosynthesis | *Streptococcus* | *Streptococcus thermophilus* | IFNγ-producing CD8^+^ T Cells | CD8^+^ T Cells | 1.12E-11 | 2.23E-12 | | 0.00154 |
| Flavin biosynthesis I (bacteria and plants) | Cofactor & Vitamin Biosynthesis | *Holdemanella* | *Holdemanella biformis* | IL-17A-producing CD161^+^ CD8^+^ T Cells | CD8^+^ T Cells | 9.43E-09 | 1.92E-09 | | 0.00157 |
| UDP-N-acetylmuramoyl-pentapeptide biosynthesis II (lysine-containing) | Cell wall & Peptidoglycan Synthesis | *Dialister* | *Dialister invisus* | IL-2/TNFα-producing CD161^+^ CD4^+^ T Cells | CD4^+^ T Cells | 4.43E-10 | 8.88E-11 | | 0.00166 |
| Chorismate biosynthesis I | General Biosynthetic Pathways | *Holdemanella* | *Holdemanella biformis* | TNFα-producing NK Cells | Natural Killer (NK) Cells | 1.29E-09 | 2.75E-10 | | 0.00167 |
| Purine ribonucleosides degradation | Nucleotide Metabolism & Biosynthesis | *Streptococcus* | *Streptococcus thermophilus* | TNFα-producing Activated CD69^+^ CD8^+^ T Cells | CD8^+^ T Cells | 3.13E-11 | 6.40E-12 | | 0.00171 |
| Adenine and adenosine salvage III | Nucleotide Metabolism & Biosynthesis | *Coprobacter* | *Coprobacter fastidiosus* | IFNγ-producing CD161^+^ NK Cells | Natural Killer (NK) Cells | 3.76E-10 | 6.85E-11 | | 0.00172 |
| tRNA charging | Amino Acid Biosynthesis | *Holdemanella* | *Holdemanella biformis* | IL-17A-producing CD4^+^ T Cells | CD4^+^ T Cells | 7.56E-10 | 1.61E-10 | | 0.00173 |
| L-valine biosynthesis | Amino Acid Biosynthesis | *Streptococcus* | *Streptococcus thermophilus* | IFNγ-producing Activated CD69^+^ CD8^+^ T Cells | CD8^+^ T Cells | 1.11E-11 | 2.26E-12 | | 0.00187 |
| L-valine biosynthesis | Amino Acid Biosynthesis | *Streptococcus* | *Streptococcus thermophilus* | CD4^+^ TEMRA | CD4^+^ T Cells | 4.57E-11 | 9.67E-12 | | 0.00194 |
| UMP biosynthesis I | Nucleotide Metabolism & Biosynthesis | *Holdemanella* | *Holdemanella biformis* | IL-17A-producing Activated CD69^+^ CD4^+^ T Cells | CD4^+^ T Cells | 1.19E-09 | 2.54E-10 | | 0.00195 |
| UMP biosynthesis II | Nucleotide Metabolism & Biosynthesis | *Holdemanella* | *Holdemanella biformis* | IL-17A-producing Activated CD69^+^ CD4^+^ T Cells | CD4^+^ T Cells | 1.19E-09 | 2.54E-10 | | 0.00195 |
| UMP biosynthesis III | Nucleotide Metabolism & Biosynthesis | *Holdemanella* | *Holdemanella biformis* | IL-17A-producing Activated CD69^+^ CD4^+^ T Cells | CD4^+^ T Cells | 1.19E-09 | 2.54E-10 | | 0.00195 |
| L-valine biosynthesis | Amino Acid Biosynthesis | *Streptococcus* | *Streptococcus thermophilus* | TNFα-producing Activated HLA-DR^+^ CD8^+^ T Cells | CD8^+^ T Cells | 1.14E-10 | 2.37E-11 | | 0.00196 |
| Guanosine ribonucleotides de novo biosynthesis | Nucleotide Metabolism & Biosynthesis | *Holdemanella* | *Holdemanella biformis* | CCR7^+^ Activated NK Cells | Natural Killer (NK) Cells | 4.22E-09 | 9.11E-10 | | 0.00198 |
| 5-aminoimidazole ribonucleotide biosynthesis I | Nucleotide Metabolism & Biosynthesis | *Holdemanella* | *Holdemanella biformis* | PD-1^+^ CD8^+^ T Cells | CD8^+^ T Cells | 7.61E-10 | 1.66E-10 | | 0.00200 |
| Coenzyme A biosynthesis II (eukaryotic) | Cofactor & Vitamin Biosynthesis | *Holdemanella* | *Holdemanella biformis* | IL-17A-producing CD161^+^ CD8^+^ T Cells | CD8^+^ T Cells | 9.47E-09 | 1.97E-09 | | 0.00201 |
| L-lysine biosynthesis VI | Amino Acid Biosynthesis | *Dialister* | *Dialister invisus* | IL-2/TNFα-producing CD161^+^ CD4^+^ T Cells | CD4^+^ T Cells | 3.94E-10 | 8.02E-11 | | 0.00202 |
| S-adenosyl-L-methionine salvage I | Amino Acid Biosynthesis | *Streptococcus* | *Streptococcus thermophilus* | IFNγ/TNFα-producing HLA-DR^+^ CD8^+^ T Cells | CD8^+^ T Cells | 4.05E-10 | 8.41E-11 | | 0.00209 |
| Queuosine biosynthesis I (de novo) | Nucleotide Metabolism & Biosynthesis | *Dialister* | *Dialister invisus* | IL-2/TNFα-producing CD161^+^ CD4^+^ T Cells | CD4^+^ T Cells | 4.64E-10 | 9.49E-11 | | 0.00212 |
| 5-aminoimidazole ribonucleotide biosynthesis II | Nucleotide Metabolism & Biosynthesis | *Holdemanella* | *Holdemanella biformis* | IL-17A-producing CD4^+^ T Cells | CD4^+^ T Cells | 1.25E-09 | 2.69E-10 | | 0.00212 |
| Superpathway of 5-aminoimidazole ribonucleotide biosynthesis | Nucleotide Metabolism & Biosynthesis | *Holdemanella* | *Holdemanella biformis* | IL-17A-producing CD4^+^ T Cells | CD4^+^ T Cells | 1.25E-09 | 2.69E-10 | | 0.00212 |
| L-valine biosynthesis | Amino Acid Biosynthesis | *Streptococcus* | *Streptococcus thermophilus* | IFNγ-producing Activated HLA-DR^+^ CD8^+^ T Cells | CD8^+^ T Cells | 4.14E-11 | 8.52E-12 | | 0.00213 |
| Coenzyme A biosynthesis I (prokaryotic) | Cofactor & Vitamin Biosynthesis | *Coprobacter* | *Coprobacter fastidiosus* | IFNγ-producing Activated NK Cells | Natural Killer (NK) Cells | 1.85E-10 | 3.28E-11 | | 0.00214 |
| Folate transformations II (plants) | Cofactor & Vitamin Biosynthesis | *Coprobacter* | *Coprobacter fastidiosus* | IFNγ-producing Activated NK Cells | Natural Killer (NK) Cells | 1.67E-10 | 2.97E-11 | | 0.00214 |
| Peptidoglycan biosynthesis III (mycobacteria) | Cell wall & Peptidoglycan Synthesis | *Coprobacter* | *Coprobacter fastidiosus* | IFNγ-producing Activated NK Cells | Natural Killer (NK) Cells | 1.81E-10 | 3.25E-11 | | 0.00214 |
| Coenzyme A biosynthesis II (eukaryotic) | Cofactor & Vitamin Biosynthesis | *Coprobacter* | *Coprobacter fastidiosus* | IFNγ-producing Activated NK Cells | Natural Killer (NK) Cells | 1.81E-10 | 2.93E-11 | | 0.00214 |
| Purine ribonucleosides degradation | Nucleotide Metabolism & Biosynthesis | *Streptococcus* | *Streptococcus thermophilus* | TNFα-producing CD8^+^ T Cells | CD8^+^ T Cells | 3.87E-11 | 8.04E-12 | | 0.00215 |
| 5-aminoimidazole ribonucleotide biosynthesis II | Nucleotide Metabolism & Biosynthesis | *Holdemanella* | *Holdemanella biformis* | IL-17A-producing CD8^+^ T Cells | CD8^+^ T Cells | 3.85E-09 | 7.75E-10 | | 0.00219 |
| Superpathway of 5-aminoimidazole ribonucleotide biosynthesis | Nucleotide Metabolism & Biosynthesis | *Holdemanella* | *Holdemanella biformis* | IL-17A-producing CD8^+^ T Cells | CD8^+^ T Cells | 3.85E-09 | 7.75E-10 | | 0.00219 |
| Chorismate biosynthesis from 3-dehydroquinate | General Biosynthetic Pathways | *Holdemanella* | *Holdemanella biformis* | TNFα-producing NK Cells | Natural Killer (NK) Cells | 1.17E-09 | 2.54E-10 | | 0.00220 |
| Guanosine ribonucleotides de novo biosynthesis | Nucleotide Metabolism & Biosynthesis | *Holdemanella* | *Holdemanella biformis* | IL-17A-producing CD161^+^ CD8^+^ T Cells | CD8^+^ T Cells | 8.30E-09 | 1.74E-09 | | 0.00223 |
| Superpathway of L-threonine biosynthesis | Amino Acid Biosynthesis | *Holdemanella* | *Holdemanella biformis* | IL-17A-producing CD161^+^ CD8^+^ T Cells | CD8^+^ T Cells | 8.98E-09 | 1.89E-09 | | 0.00224 |
| S-adenosyl-L-methionine salvage I | Amino Acid Biosynthesis | *Streptococcus* | *Streptococcus thermophilus* | IFNγ-producing Activated HLA-DR^+^ CD4^+^ T Cells | CD4^+^ T Cells | 1.33E-10 | 2.78E-11 | | 0.00226 |
| Chorismate biosynthesis I | General Biosynthetic Pathways | *Holdemanella* | *Holdemanella biformis* | IL-17A-producing CD8^+^ T Cells | CD8^+^ T Cells | 2.73E-09 | 5.56E-10 | | 0.00234 |
| Adenine and adenosine salvage III | Nucleotide Metabolism & Biosynthesis | *Dialister* | *Dialister invisus* | IL-2/TNFα-producing CD161^+^ CD4^+^ T Cells | CD4^+^ T Cells | 3.83E-10 | 7.90E-11 | | 0.00237 |
| S-adenosyl-L-methionine salvage I | Amino Acid Biosynthesis | *Streptococcus* | *Streptococcus thermophilus* | CD4^+^ TEMRA | CD4^+^ T Cells | 5.08E-11 | 1.09E-11 | | 0.00239 |
| Coenzyme A biosynthesis I (prokaryotic) | Cofactor & Vitamin Biosynthesis | *Holdemanella* | *Holdemanella biformis* | IL-17A-producing Activated CD69^+^ CD4^+^ T Cells | CD4^+^ T Cells | 9.66E-10 | 2.09E-10 | | 0.00239 |
| Inosine 5-phosphate degradation | Nucleotide Metabolism & Biosynthesis | *Coprobacter* | *Coprobacter fastidiosus* | IFNγ-producing Activated NK Cells | Natural Killer (NK) Cells | 1.49E-10 | 2.73E-11 | | 0.00241 |
| UDP-N-acetylmuramoyl-pentapeptide biosynthesis I (meso-diaminopimelate containing) | Cell wall & Peptidoglycan Synthesis | *Coprobacter* | *Coprobacter fastidiosus* | IFNγ-producing Activated NK Cells | Natural Killer (NK) Cells | 1.73E-10 | 3.18E-11 | | 0.00241 |
| L-valine biosynthesis | Amino Acid Biosynthesis | *Streptococcus* | *Streptococcus thermophilus* | IFNγ-producing CD8^+^ T Cells | CD8^+^ T Cells | 1.04E-11 | 2.15E-12 | | 0.00241 |
| Guanosine ribonucleotides de novo biosynthesis | Nucleotide Metabolism & Biosynthesis | *Holdemanella* | *Holdemanella biformis* | IL-17A-producing CD8^+^ T Cells | CD8^+^ T Cells | 2.28E-09 | 4.66E-10 | | 0.00246 |
| S-adenosyl-L-methionine salvage I | Amino Acid Biosynthesis | *Streptococcus* | *Streptococcus thermophilus* | IFNγ-producing Activated CD69^+^ CD8^+^ T Cells | CD8^+^ T Cells | 1.23E-11 | 2.56E-12 | | 0.00250 |
| Chorismate biosynthesis I | General Biosynthetic Pathways | *Holdemanella* | *Holdemanella biformis* | IL-17A-producing CD161^+^ CD8^+^ T Cells | CD8^+^ T Cells | 9.84E-09 | 2.09E-09 | | 0.00260 |
| L-valine biosynthesis | Amino Acid Biosynthesis | *Eubacterium* | *Eubacterium ventriosum* | Non-classical Monocytes | Innate Immune Cells | 1.08E-09 | 2.32E-10 | | 0.00262 |
| 5-aminoimidazole ribonucleotide biosynthesis II | Nucleotide Metabolism & Biosynthesis | *Holdemanella* | *Holdemanella biformis* | IL-17A-producing CD161^+^ CD8^+^ T Cells | CD8^+^ T Cells | 1.37E-08 | 2.93E-09 | | 0.00266 |
| Superpathway of 5-aminoimidazole ribonucleotide biosynthesis | Nucleotide Metabolism & Biosynthesis | *Holdemanella* | *Holdemanella biformis* | IL-17A-producing CD161^+^ CD8^+^ T Cells | CD8^+^ T Cells | 1.37E-08 | 2.93E-09 | | 0.00266 |
| Superpathway of coenzyme A biosynthesis III (mammals) | Cofactor & Vitamin Biosynthesis | *Holdemanella* | *Holdemanella biformis* | IL-17A-producing CD8^+^ T Cells | CD8^+^ T Cells | 2.62E-09 | 5.41E-10 | | 0.00268 |
| S-adenosyl-L-methionine salvage I | Amino Acid Biosynthesis | *Holdemanella* | *Holdemanella biformis* | IL-17A-producing CD8^+^ T Cells | CD8^+^ T Cells | 3.28E-09 | 6.77E-10 | | 0.00268 |
| tRNA charging | Amino Acid Biosynthesis | *Holdemanella* | *Holdemanella biformis* | IL-17A-producing CD8^+^ T Cells | CD8^+^ T Cells | 2.27E-09 | 4.68E-10 | | 0.00268 |
| Coenzyme A biosynthesis I (prokaryotic) | Cofactor & Vitamin Biosynthesis | *Coprobacter* | *Coprobacter fastidiosus* | IFNγ-producing NK Cells | Natural Killer (NK) Cells | 1.81E-10 | 3.26E-11 | | 0.00272 |
| Folate transformations II (plants) | Cofactor & Vitamin Biosynthesis | *Coprobacter* | *Coprobacter fastidiosus* | IFNγ-producing NK Cells | Natural Killer (NK) Cells | 1.64E-10 | 2.95E-11 | | 0.00272 |
| Peptidoglycan biosynthesis III (mycobacteria) | Cell wall & Peptidoglycan Synthesis | *Coprobacter* | *Coprobacter fastidiosus* | IFNγ-producing NK Cells | Natural Killer (NK) Cells | 1.77E-10 | 3.24E-11 | | 0.00272 |
| Coenzyme A biosynthesis II (eukaryotic) | Cofactor & Vitamin Biosynthesis | *Coprobacter* | *Coprobacter fastidiosus* | IFNγ-producing NK Cells | Natural Killer (NK) Cells | 1.78E-10 | 2.91E-11 | | 0.00272 |
| Chorismate biosynthesis from 3-dehydroquinate | General Biosynthetic Pathways | *Holdemanella* | *Holdemanella biformis* | IL-17A-producing CD8^+^ T Cells | CD8^+^ T Cells | 2.48E-09 | 5.14E-10 | | 0.00278 |
| L-valine biosynthesis | Amino Acid Biosynthesis | *Streptococcus* | *Streptococcus thermophilus* | TNFα-producing Activated CD69^+^ CD8^+^ T Cells | CD8^+^ T Cells | 2.91E-11 | 6.16E-12 | | 0.00278 |
| Queuosine biosynthesis I (de novo) | Nucleotide Metabolism & Biosynthesis | *Coprobacter* | *Coprobacter fastidiosus* | IFNγ-producing CD161^+^ CD8^+^ T Cells | CD8^+^ T Cells | 9.59E-11 | 1.88E-11 | | 0.00282 |
| Glycogen biosynthesis I (from ADP-D-Glucose) | General Biosynthetic Pathways | *Holdemanella* | *Holdemanella biformis* | PD-1^+^ CD8^+^ T Cells | CD8^+^ T Cells | 5.70E-10 | 1.28E-10 | | 0.00283 |
| Sucrose biosynthesis II | General Biosynthetic Pathways | *Holdemanella* | *Holdemanella biformis* | PD-1^+^ CD8^+^ T Cells | CD8^+^ T Cells | 5.00E-10 | 1.12E-10 | | 0.00284 |
| Queuosine biosynthesis I (de novo) | Nucleotide Metabolism & Biosynthesis | *Coprobacter* | *Coprobacter fastidiosus* | IFNγ/TNFα-producing CD69^+^ CD4^+^ T Cells | CD4^+^ T Cells | 3.40E-11 | 6.83E-12 | | 0.00285 |
| Superpathway of coenzyme A biosynthesis III (mammals) | Cofactor & Vitamin Biosynthesis | *Coprobacter* | *Coprobacter fastidiosus* | IFNγ-producing CD161^+^ NK Cells | Natural Killer (NK) Cells | 3.16E-10 | 5.99E-11 | | 0.00286 |
| S-adenosyl-L-methionine salvage I | Amino Acid Biosynthesis | *Coprobacter* | *Coprobacter fastidiosus* | IFNγ-producing CD161^+^ NK Cells | Natural Killer (NK) Cells | 2.94E-10 | 5.57E-11 | | 0.00286 |
| Chorismate biosynthesis I | General Biosynthetic Pathways | *Holdemanella* | *Holdemanella biformis* | TNFα-producing CD161^+^ NK Cells | Natural Killer (NK) Cells | 1.21E-09 | 2.66E-10 | | 0.00289 |
| Coenzyme A biosynthesis I (prokaryotic) | Cofactor & Vitamin Biosynthesis | *Holdemanella* | *Holdemanella biformis* | IL-17A-producing CD161^+^ CD8^+^ T Cells | CD8^+^ T Cells | 9.99E-09 | 2.16E-09 | | 0.00295 |
| Chorismate biosynthesis from 3-dehydroquinate | General Biosynthetic Pathways | *Holdemanella* | *Holdemanella biformis* | IL-17A-producing CD161^+^ CD8^+^ T Cells | CD8^+^ T Cells | 8.93E-09 | 1.93E-09 | | 0.00296 |
| tRNA charging | Amino Acid Biosynthesis | *Holdemanella* | *Holdemanella biformis* | IL-17A-producing CD161^+^ CD8^+^ T Cells | CD8^+^ T Cells | 8.15E-09 | 1.76E-09 | | 0.00296 |
| 5-aminoimidazole ribonucleotide biosynthesis I | Nucleotide Metabolism & Biosynthesis | *Eubacterium* | *Eubacterium ventriosum* | CD8^+^ CM | CD8^+^ T Cells | 8.74E-11 | 1.60E-11 | | 0.00300 |
| 5-aminoimidazole ribonucleotide biosynthesis II | Nucleotide Metabolism & Biosynthesis | *Eubacterium* | *Eubacterium ventriosum* | CD8^+^ CM | CD8^+^ T Cells | 8.07E-11 | 1.55E-11 | | 0.00300 |
| Superpathway of 5-aminoimidazole ribonucleotide biosynthesis | Nucleotide Metabolism & Biosynthesis | *Eubacterium* | *Eubacterium ventriosum* | CD8^+^ CM | CD8^+^ T Cells | 8.07E-11 | 1.55E-11 | | 0.00300 |
| Fatty acid biosynthesis initiation (mitochondria) | Lipid & Membrane Metabolism | *Eubacterium* | *Eubacterium ventriosum* | CD8^+^ CM | CD8^+^ T Cells | 6.93E-11 | 1.37E-11 | | 0.00300 |
| S-adenosyl-L-methionine salvage I | Amino Acid Biosynthesis | *Streptococcus* | *Streptococcus thermophilus* | IFNγ-producing CD8^+^ T Cells | CD8^+^ T Cells | 1.15E-11 | 2.44E-12 | | 0.00304 |
| Inosine 5-phosphate degradation | Nucleotide Metabolism & Biosynthesis | *Coprobacter* | *Coprobacter fastidiosus* | IFNγ-producing NK Cells | Natural Killer (NK) Cells | 1.46E-10 | 2.72E-11 | | 0.00305 |
| UDP-N-acetylmuramoyl-pentapeptide biosynthesis I (meso-diaminopimelate containing) | Cell wall & Peptidoglycan Synthesis | *Coprobacter* | *Coprobacter fastidiosus* | IFNγ-producing NK Cells | Natural Killer (NK) Cells | 1.70E-10 | 3.17E-11 | | 0.00305 |
| tRNA charging | Amino Acid Biosynthesis | *Holdemanella* | *Holdemanella biformis* | TNFα-producing NK Cells | Natural Killer (NK) Cells | 1.05E-09 | 2.34E-10 | | 0.00311 |
| Flavin biosynthesis I (bacteria and plants) | Cofactor & Vitamin Biosynthesis | *Clostridium* | *Clostridium sp AM22 11AC* | IFNγ-producing T regs | Adaptive Immune Cells | 1.38E-09 | 2.74E-10 | | 0.00321 |
| Peptidoglycan biosynthesis I (meso-diaminopimelate containing) | Cell wall & Peptidoglycan Synthesis | *Holdemanella* | *Holdemanella biformis* | Non-classical Monocytes | Innate Immune Cells | 5.39E-09 | 1.17E-09 | | 0.00327 |
| UDP-N-acetylmuramoyl-pentapeptide biosynthesis I (meso-diaminopimelate containing) | Cell wall & Peptidoglycan Synthesis | *Holdemanella* | *Holdemanella biformis* | Non-classical Monocytes | Innate Immune Cells | 5.30E-09 | 1.15E-09 | | 0.00334 |
| S-adenosyl-L-methionine salvage I | Amino Acid Biosynthesis | *Streptococcus* | *Streptococcus thermophilus* | TNFα-producing Activated CD69^+^ CD8^+^ T Cells | CD8^+^ T Cells | 3.24E-11 | 6.95E-12 | | 0.00336 |
| L-valine biosynthesis | Amino Acid Biosynthesis | *Streptococcus* | *Streptococcus thermophilus* | TNFα-producing CD8^+^ T Cells | CD8^+^ T Cells | 3.60E-11 | 7.74E-12 | | 0.00348 |
| UDP-N-acetylmuramoyl-pentapeptide biosynthesis II (lysine-containing) | Cell wall & Peptidoglycan Synthesis | *Holdemanella* | *Holdemanella biformis* | Non-classical Monocytes | Innate Immune Cells | 5.56E-09 | 1.22E-09 | | 0.00355 |
| S-adenosyl-L-methionine salvage I | Amino Acid Biosynthesis | *Streptococcus* | *Streptococcus thermophilus* | TNFα-producing Activated HLA-DR^+^ CD8^+^ T Cells | CD8^+^ T Cells | 1.25E-10 | 2.70E-11 | | 0.00358 |
| Sucrose biosynthesis II | General Biosynthetic Pathways | *Eubacterium* | *Eubacterium ventriosum* | Non-classical Monocytes | Innate Immune Cells | 8.19E-10 | 1.80E-10 | | 0.00364 |
| Chorismate biosynthesis from 3-dehydroquinate | General Biosynthetic Pathways | *Holdemanella* | *Holdemanella biformis* | TNFα-producing CD161^+^ NK Cells | Natural Killer (NK) Cells | 1.09E-09 | 2.46E-10 | | 0.00375 |
| Flavin biosynthesis I (bacteria and plants) | Cofactor & Vitamin Biosynthesis | *Clostridium* | *Clostridium sp AM22 11AC* | CD8^+^ CM | CD8^+^ T Cells | 1.53E-10 | 3.09E-11 | | 0.00381 |
| 5-aminoimidazole ribonucleotide biosynthesis I | Nucleotide Metabolism & Biosynthesis | *Holdemanella* | *Holdemanella biformis* | IL-17A-producing CD8^+^ T Cells | CD8^+^ T Cells | 3.45E-09 | 7.34E-10 | | 0.00381 |
| S-adenosyl-L-methionine salvage I | Amino Acid Biosynthesis | *Holdemanella* | *Holdemanella biformis* | PD-1^+^ CD8^+^ T Cells | CD8^+^ T Cells | 6.87E-10 | 1.58E-10 | | 0.00382 |
| S-adenosyl-L-methionine salvage I | Amino Acid Biosynthesis | *Holdemanella* | *Holdemanella biformis* | IL-17A-producing CD161^+^ CD8^+^ T Cells | CD8^+^ T Cells | 1.16E-08 | 2.57E-09 | | 0.00394 |
| 5-aminoimidazole ribonucleotide biosynthesis I | Nucleotide Metabolism & Biosynthesis | *Holdemanella* | *Holdemanella biformis* | IL-17A-producing CD4^+^ T Cells | CD4^+^ T Cells | 1.12E-09 | 2.54E-10 | | 0.00401 |
| 5-aminoimidazole ribonucleotide biosynthesis II | Nucleotide Metabolism & Biosynthesis | *Holdemanella* | *Holdemanella biformis* | TNFα-producing NK Cells | Natural Killer (NK) Cells | 1.73E-09 | 3.94E-10 | | 0.00411 |
| Superpathway of 5-aminoimidazole ribonucleotide biosynthesis | Nucleotide Metabolism & Biosynthesis | *Holdemanella* | *Holdemanella biformis* | TNFα-producing NK Cells | Natural Killer (NK) Cells | 1.73E-09 | 3.94E-10 | | 0.00411 |
| S-adenosyl-L-methionine salvage I | Amino Acid Biosynthesis | *Streptococcus* | *Streptococcus thermophilus* | TNFα-producing CD8^+^ T Cells | CD8^+^ T Cells | 4.01E-11 | 8.73E-12 | | 0.00413 |
| Coenzyme A biosynthesis I (prokaryotic) | Cofactor & Vitamin Biosynthesis | *Holdemanella* | *Holdemanella biformis* | IL-17A-producing CD8^+^ T Cells | CD8^+^ T Cells | 2.70E-09 | 5.81E-10 | | 0.00426 |
| Coenzyme A biosynthesis II (eukaryotic) | Cofactor & Vitamin Biosynthesis | *Holdemanella* | *Holdemanella biformis* | IL-17A-producing CD8^+^ T Cells | CD8^+^ T Cells | 2.50E-09 | 5.37E-10 | | 0.00426 |
| Superpathway of L-threonine biosynthesis | Amino Acid Biosynthesis | *Holdemanella* | *Holdemanella biformis* | IL-17A-producing CD8^+^ T Cells | CD8^+^ T Cells | 2.38E-09 | 5.13E-10 | | 0.00426 |
| S-adenosyl-L-methionine salvage I | Amino Acid Biosynthesis | *Streptococcus* | *Streptococcus thermophilus* | IFNγ-producing Activated HLA-DR^+^ CD8^+^ T Cells | CD8^+^ T Cells | 4.48E-11 | 9.76E-12 | | 0.00427 |
| Purine ribonucleosides degradation | Nucleotide Metabolism & Biosynthesis | *Streptococcus* | *Streptococcus thermophilus* | IFNγ/TNFα-producing CD69^+^ CD4^+^ T Cells | CD4^+^ T Cells | 2.42E-11 | 5.07E-12 | | 0.00430 |
| Coenzyme A biosynthesis II (eukaryotic) | Cofactor & Vitamin Biosynthesis | *Coprobacter* | *Coprobacter fastidiosus* | IFNγ-producing T regs | Adaptive Immune Cells | 8.83E-10 | 1.82E-10 | | 0.00469 |
| Coenzyme A biosynthesis II (eukaryotic) | Cofactor & Vitamin Biosynthesis | *Coprobacter* | *Coprobacter fastidiosus* | IFNγ/TNFα-producing CD161^+^ CD8^+^ T Cells | CD8^+^ T Cells | 4.51E-10 | 1.01E-10 | | 0.00469 |
| 5-aminoimidazole ribonucleotide biosynthesis I | Nucleotide Metabolism & Biosynthesis | *Holdemanella* | *Holdemanella biformis* | IL-17A-producing CD161^+^ CD8^+^ T Cells | CD8^+^ T Cells | 1.23E-08 | 2.77E-09 | | 0.00481 |
| Fatty acid biosynthesis initiation (mitochondria) | Lipid & Membrane Metabolism | *Escherichia* | *Escherichia coli* | IL-2-producing CD4^+^ EM T Cells | CD4^+^ T Cells | 1.88E-10 | 3.88E-11 | | 0.00494 |
| Chorismate biosynthesis I | General Biosynthetic Pathways | *Holdemanella* | *Holdemanella biformis* | IL-17A-producing Activated CD69^+^ CD4^+^ T Cells | CD4^+^ T Cells | 9.07E-10 | 2.08E-10 | | 0.00497 |
| Adenine and adenosine salvage III | Nucleotide Metabolism & Biosynthesis | *Coprobacter* | *Coprobacter fastidiosus* | IFNγ-producing Activated NK Cells | Natural Killer (NK) Cells | 1.74E-10 | 3.39E-11 | | 0.00506 |
| UMP biosynthesis I | Nucleotide Metabolism & Biosynthesis | *Holdemanella* | *Holdemanella biformis* | CCR7^+^ Activated NK Cells | Natural Killer (NK) Cells | 6.00E-09 | 1.40E-09 | | 0.00514 |
| UMP biosynthesis II | Nucleotide Metabolism & Biosynthesis | *Holdemanella* | *Holdemanella biformis* | CCR7^+^ Activated NK Cells | Natural Killer (NK) Cells | 6.00E-09 | 1.40E-09 | | 0.00514 |
| UMP biosynthesis III | Nucleotide Metabolism & Biosynthesis | *Holdemanella* | *Holdemanella biformis* | CCR7^+^ Activated NK Cells | Natural Killer (NK) Cells | 6.00E-09 | 1.40E-09 | | 0.00514 |
| tRNA charging | Amino Acid Biosynthesis | *Holdemanella* | *Holdemanella biformis* | TNFα-producing CD161^+^ NK Cells | Natural Killer (NK) Cells | 9.77E-10 | 2.27E-10 | | 0.00516 |
| Superpathway of L-threonine biosynthesis | Amino Acid Biosynthesis | *Holdemanella* | *Holdemanella biformis* | Non-classical Monocytes | Innate Immune Cells | 5.29E-09 | 1.20E-09 | | 0.00519 |
| Coenzyme A biosynthesis II (eukaryotic) | Cofactor & Vitamin Biosynthesis | *Holdemanella* | *Holdemanella biformis* | Non-classical Monocytes | Innate Immune Cells | 5.53E-09 | 1.25E-09 | | 0.00531 |
| Superpathway of coenzyme A biosynthesis III (mammals) | Cofactor & Vitamin Biosynthesis | *Holdemanella* | *Holdemanella biformis* | Non-classical Monocytes | Innate Immune Cells | 5.65E-09 | 1.28E-09 | | 0.00533 |
| Purine ribonucleosides degradation | Nucleotide Metabolism & Biosynthesis | *Streptococcus* | *Streptococcus thermophilus* | IFNγ-producing CD161^+^ CD8^+^ T Cells | CD8^+^ T Cells | 6.61E-11 | 1.42E-11 | | 0.00541 |
| CDP diacylglycerol biosynthesis I | Lipid & Membrane Metabolism | *Coprobacter* | *Coprobacter fastidiosus* | NK T Cells | Adaptive Immune Cells | 2.96E-11 | 6.76E-12 | | 0.00542 |
| CDP diacylglycerol biosynthesis II | Lipid & Membrane Metabolism | *Coprobacter* | *Coprobacter fastidiosus* | NK T Cells | Adaptive Immune Cells | 2.96E-11 | 6.76E-12 | | 0.00542 |
| Sucrose biosynthesis II | General Biosynthetic Pathways | *Eubacterium* | *Eubacterium ventriosum* | CD8^+^ CM | CD8^+^ T Cells | 6.04E-11 | 1.26E-11 | | 0.00556 |
| Superpathway of L-threonine biosynthesis | Amino Acid Biosynthesis | *Escherichia* | *Escherichia coli* | IL-2-producing CD4^+^ EM T Cells | CD4^+^ T Cells | 1.67E-10 | 3.49E-11 | | 0.00557 |
| Coenzyme A biosynthesis II (eukaryotic) | Cofactor & Vitamin Biosynthesis | *Coprobacter* | *Coprobacter fastidiosus* | IFNγ-producing CD161^+^ CD4^+^ T Cells | CD4^+^ T Cells | 5.58E-11 | 1.26E-11 | | 0.00573 |
| L-histidine biosynthesis | Amino Acid Biosynthesis | *Clostridium* | *Clostridium sp AM22 11AC* | IFNγ-producing T regs | Adaptive Immune Cells | 9.67E-10 | 2.05E-10 | | 0.00578 |
| CDP diacylglycerol biosynthesis I | Lipid & Membrane Metabolism | *Coprobacter* | *Coprobacter fastidiosus* | IFNγ-producing NK T Cells | Adaptive Immune Cells | 4.29E-11 | 9.85E-12 | | 0.00582 |
| CDP diacylglycerol biosynthesis II | Lipid & Membrane Metabolism | *Coprobacter* | *Coprobacter fastidiosus* | IFNγ-producing NK T Cells | Adaptive Immune Cells | 4.29E-11 | 9.85E-12 | | 0.00582 |
| 5-aminoimidazole ribonucleotide biosynthesis I | Nucleotide Metabolism & Biosynthesis | *Coprobacter* | *Coprobacter fastidiosus* | IFNγ-producing CD161^+^ NK Cells | Natural Killer (NK) Cells | 4.18E-10 | 8.30E-11 | | 0.00591 |
| L-valine biosynthesis | Amino Acid Biosynthesis | *Streptococcus* | *Streptococcus thermophilus* | IFNγ/TNFα-producing CD69^+^ CD4^+^ T Cells | CD4^+^ T Cells | 2.25E-11 | 4.87E-12 | | 0.00591 |
| Methylerythritol phosphate pathway I | General Biosynthetic Pathways | *Holdemanella* | *Holdemanella biformis* | IL-17A-producing CD161^+^ CD4^+^ T Cells | CD4^+^ T Cells | 2.49E-09 | 5.46E-10 | | 0.00602 |
| Flavin biosynthesis I (bacteria and plants) | Cofactor & Vitamin Biosynthesis | *Holdemanella* | *Holdemanella biformis* | IL-17A-producing CD161^+^ CD4^+^ T Cells | CD4^+^ T Cells | 2.36E-09 | 5.17E-10 | | 0.00602 |
| Adenine and adenosine salvage III | Nucleotide Metabolism & Biosynthesis | *Coprobacter* | *Coprobacter fastidiosus* | IFNγ-producing NK Cells | Natural Killer (NK) Cells | 1.71E-10 | 3.37E-11 | | 0.00618 |
| L-lysine biosynthesis VI | Amino Acid Biosynthesis | *Clostridium* | *Clostridium sp AM22 11AC* | CD8^+^ CM | CD8^+^ T Cells | 1.22E-10 | 2.57E-11 | | 0.00620 |
| Chorismate biosynthesis from 3-dehydroquinate | General Biosynthetic Pathways | *Holdemanella* | *Holdemanella biformis* | IL-17A-producing Activated CD69^+^ CD4^+^ T Cells | CD4^+^ T Cells | 8.22E-10 | 1.92E-10 | | 0.00632 |
| Fatty acid biosynthesis initiation (mitochondria) | Lipid & Membrane Metabolism | *Escherichia* | *Escherichia coli* | γδ T Cells | Adaptive Immune Cells | 4.05E-11 | 8.40E-12 | | 0.00632 |
| S-adenosyl-L-methionine salvage I | Amino Acid Biosynthesis | *Holdemanella* | *Holdemanella biformis* | IL-17A-producing CD4^+^ T Cells | CD4^+^ T Cells | 1.02E-09 | 2.40E-10 | | 0.00643 |
| Queuosine biosynthesis I (de novo) | Nucleotide Metabolism & Biosynthesis | *Escherichia* | *Escherichia coli* | IL-2-producing CD4^+^ EM T Cells | CD4^+^ T Cells | 1.49E-10 | 3.16E-11 | | 0.00645 |
| 5-aminoimidazole ribonucleotide biosynthesis II | Nucleotide Metabolism & Biosynthesis | *Holdemanella* | *Holdemanella biformis* | TNFα-producing CD161^+^ NK Cells | Natural Killer (NK) Cells | 1.61E-09 | 3.81E-10 | | 0.00656 |
| Superpathway of 5-aminoimidazole ribonucleotide biosynthesis | Nucleotide Metabolism & Biosynthesis | *Holdemanella* | *Holdemanella biformis* | TNFα-producing CD161^+^ NK Cells | Natural Killer (NK) Cells | 1.61E-09 | 3.81E-10 | | 0.00656 |
| Queuosine biosynthesis I (de novo) | Nucleotide Metabolism & Biosynthesis | *Coprobacter* | *Coprobacter fastidiosus* | IFNγ-producing CD4^+^ T Cells | CD4^+^ T Cells | 1.44E-11 | 3.10E-12 | | 0.00662 |
| 5-aminoimidazole ribonucleotide biosynthesis II | Nucleotide Metabolism & Biosynthesis | *Coprobacter* | *Coprobacter fastidiosus* | IFNγ-producing CD161^+^ NK Cells | Natural Killer (NK) Cells | 3.86E-10 | 7.77E-11 | | 0.00663 |
| Superpathway of 5-aminoimidazole ribonucleotide biosynthesis | Nucleotide Metabolism & Biosynthesis | *Coprobacter* | *Coprobacter fastidiosus* | IFNγ-producing CD161^+^ NK Cells | Natural Killer (NK) Cells | 3.86E-10 | 7.77E-11 | | 0.00663 |
| S-adenosyl-L-methionine salvage I | Amino Acid Biosynthesis | *Coprobacter* | *Coprobacter fastidiosus* | IFNγ-producing Activated NK Cells | Natural Killer (NK) Cells | 1.38E-10 | 2.73E-11 | | 0.00666 |
| L-lysine biosynthesis VI | Amino Acid Biosynthesis | *Clostridium* | *Clostridium sp AM22 11AC* | IFNγ-producing T regs | Adaptive Immune Cells | 1.06E-09 | 2.32E-10 | | 0.00675 |
| UDP-N-acetylmuramoyl-pentapeptide biosynthesis I (meso-diaminopimelate containing) | Cell wall & Peptidoglycan Synthesis | *Holdemanella* | *Holdemanella biformis* | IL-17A-producing Activated NK Cells | Natural Killer (NK) Cells | 1.27E-08 | 2.96E-09 | | 0.00682 |
| Fatty acid biosynthesis initiation (mitochondria) | Lipid & Membrane Metabolism | *Eubacterium* | *Eubacterium ventriosum* | Non-classical Monocytes | Innate Immune Cells | 8.73E-10 | 2.03E-10 | | 0.00686 |
| Peptidoglycan biosynthesis I (meso-diaminopimelate containing) | Cell wall & Peptidoglycan Synthesis | *Holdemanella* | *Holdemanella biformis* | IL-17A-producing Activated NK Cells | Natural Killer (NK) Cells | 1.29E-08 | 3.02E-09 | | 0.00689 |
| UDP-N-acetylmuramoyl-pentapeptide biosynthesis II (lysine-containing) | Cell wall & Peptidoglycan Synthesis | *Holdemanella* | *Holdemanella biformis* | IL-17A-producing Activated NK Cells | Natural Killer (NK) Cells | 1.34E-08 | 3.12E-09 | | 0.00689 |
| L-valine biosynthesis | Amino Acid Biosynthesis | *Streptococcus* | *Streptococcus thermophilus* | IFNγ-producing CD161^+^ CD8^+^ T Cells | CD8^+^ T Cells | 6.15E-11 | 1.36E-11 | | 0.00709 |
| Flavin biosynthesis I (bacteria and plants) | Cofactor & Vitamin Biosynthesis | *Holdemanella* | *Holdemanella biformis* | IL-17A-producing CD8^+^ T Cells | CD8^+^ T Cells | 2.36E-09 | 5.39E-10 | | 0.00713 |
| L-histidine biosynthesis | Amino Acid Biosynthesis | *Coprobacter* | *Coprobacter fastidiosus* | IFNγ-producing CD161^+^ NK Cells | Natural Killer (NK) Cells | 3.43E-10 | 6.97E-11 | | 0.00733 |
| 5-aminoimidazole ribonucleotide biosynthesis I | Nucleotide Metabolism & Biosynthesis | *Holdemanella* | *Holdemanella biformis* | TNFα-producing NK Cells | Natural Killer (NK) Cells | 1.55E-09 | 3.72E-10 | | 0.00743 |
| S-adenosyl-L-methionine salvage I | Amino Acid Biosynthesis | *Streptococcus* | *Streptococcus thermophilus* | IFNγ/TNFα-producing CD69^+^ CD4^+^ T Cells | CD4^+^ T Cells | 2.48E-11 | 5.53E-12 | | 0.00744 |
| Glycogen biosynthesis I (from ADP-D-Glucose) | General Biosynthetic Pathways | *Holdemanella* | *Holdemanella biformis* | IL-17A-producing CD4^+^ T Cells | CD4^+^ T Cells | 8.26E-10 | 1.97E-10 | | 0.00751 |
| Coenzyme A biosynthesis I (prokaryotic) | Cofactor & Vitamin Biosynthesis | *Holdemanella* | *Holdemanella biformis* | CCR7^+^ Activated NK Cells | Natural Killer (NK) Cells | 4.79E-09 | 1.16E-09 | | 0.00773 |
| L-histidine biosynthesis | Amino Acid Biosynthesis | *Clostridium* | *Clostridium sp AM22 11AC* | CD8^+^ CM | CD8^+^ T Cells | 1.07E-10 | 2.32E-11 | | 0.00775 |
| Sucrose biosynthesis II | General Biosynthetic Pathways | *Holdemanella* | *Holdemanella biformis* | IL-17A-producing CD4^+^ T Cells | CD4^+^ T Cells | 7.22E-10 | 1.73E-10 | | 0.00785 |
| Coenzyme A biosynthesis II (eukaryotic) | Cofactor & Vitamin Biosynthesis | *Coprobacter* | *Coprobacter fastidiosus* | IFNγ/TNFα-producing CD161^+^ CD4^+^ T Cells | CD4^+^ T Cells | 9.43E-11 | 2.16E-11 | | 0.00788 |
| Purine ribonucleosides degradation | Nucleotide Metabolism & Biosynthesis | *Eubacterium* | *Eubacterium ventriosum* | PD-1^+^ CD8^+^ T Cells | CD8^+^ T Cells | 1.01E-10 | 2.45E-11 | | 0.00793 |
| Purine ribonucleosides degradation | Nucleotide Metabolism & Biosynthesis | *Eubacterium* | *Eubacterium ventriosum* | PD-1^+^ CD4^+^ T Cells | CD4^+^ T Cells | 4.55E-11 | 1.11E-11 | | 0.00818 |
| Folate transformations II (plants) | Cofactor & Vitamin Biosynthesis | *Eubacterium* | *Eubacterium ventriosum* | IL-17A-producing Activated CD69^+^ CD4^+^ T Cells | CD4^+^ T Cells | 1.33E-10 | 3.18E-11 | | 0.00818 |
| tRNA charging | Amino Acid Biosynthesis | *Holdemanella* | *Holdemanella biformis* | IL-17A-producing Activated CD69^+^ CD4^+^ T Cells | CD4^+^ T Cells | 7.39E-10 | 1.76E-10 | | 0.00818 |
| L-histidine biosynthesis | Amino Acid Biosynthesis | *Clostridium* | *Clostridium sp AM22 11AC* | CCR7^+^ CD8^+^ T Cells | CD8^+^ T Cells | 4.05E-11 | 7.21E-12 | | 0.00828 |
| UMP biosynthesis I | Nucleotide Metabolism & Biosynthesis | *Clostridium* | *Clostridium sp AM22 11AC* | CCR7^+^ CD8^+^ T Cells | CD8^+^ T Cells | 5.96E-11 | 1.11E-11 | | 0.00828 |
| UMP biosynthesis II | Nucleotide Metabolism & Biosynthesis | *Clostridium* | *Clostridium sp AM22 11AC* | CCR7^+^ CD8^+^ T Cells | CD8^+^ T Cells | 5.96E-11 | 1.11E-11 | | 0.00828 |
| UMP biosynthesis II | Nucleotide Metabolism & Biosynthesis | *Clostridium* | *Clostridium sp AM22 11AC* | CCR7^+^ CD8^+^ T Cells | CD8^+^ T Cells | 5.96E-11 | 1.11E-11 | | 0.00828 |
| S-adenosyl-L-methionine salvage I | Amino Acid Biosynthesis | *Coprobacter* | *Coprobacter fastidiosus* | IFNγ-producing NK Cells | Natural Killer (NK) Cells | 1.35E-10 | 2.72E-11 | | 0.00828 |
| Glycogen biosynthesis I (from ADP-D-Glucose) | General Biosynthetic Pathways | *Escherichia* | *Escherichia coli* | IL-2-producing CD4^+^ EM T Cells | CD4^+^ T Cells | 1.34E-10 | 2.95E-11 | | 0.00836 |
| Inosine 5-phosphate degradation | Nucleotide Metabolism & Biosynthesis | *Escherichia* | *Escherichia coli* | IL-2-producing CD4^+^ EM T Cells | CD4^+^ T Cells | 2.00E-10 | 4.37E-11 | | 0.00836 |
| Coenzyme A biosynthesis II (eukaryotic) | Cofactor & Vitamin Biosynthesis | *Coprobacter* | *Coprobacter fastidiosus* | IFNγ/TNFα-producing CD69^+^ CD8^+^ T Cells | CD8^+^ T Cells | 1.82E-10 | 4.21E-11 | | 0.00850 |
| Purine ribonucleosides degradation | Nucleotide Metabolism & Biosynthesis | *Eubacterium* | *Eubacterium ventriosum* | TNFα-producing T regs | Adaptive Immune Cells | 9.61E-11 | 2.37E-11 | | 0.00870 |
| Sucrose biosynthesis II | General Biosynthetic Pathways | *Clostridium* | *Clostridium sp AM22 11AC* | IFNγ-producing T regs | Adaptive Immune Cells | 1.80E-09 | 4.17E-10 | | 0.00873 |
| Folate transformations II (plants) | Cofactor & Vitamin Biosynthesis | *Eubacterium* | *Eubacterium ventriosum* | IL-17A-producing CD4^+^ T Cells | CD4^+^ T Cells | 1.25E-10 | 3.02E-11 | | 0.00884 |
| Coenzyme A biosynthesis II (eukaryotic) | Cofactor & Vitamin Biosynthesis | *Coprobacter* | *Coprobacter fastidiosus* | IFNγ-producing CD4^+^ T Cells | CD4^+^ T Cells | 1.78E-11 | 4.12E-12 | | 0.00885 |
| Coenzyme A biosynthesis I (prokaryotic) | Cofactor & Vitamin Biosynthesis | *Coprobacter* | *Coprobacter fastidiosus* | IFNγ-producing T regs | Adaptive Immune Cells | 8.76E-10 | 2.04E-10 | | 0.00912 |
| S-adenosyl-L-methionine salvage I | Amino Acid Biosynthesis | *Streptococcus* | *Streptococcus thermophilus* | IFNγ-producing CD161^+^ CD8^+^ T Cells | CD8^+^ T Cells | 6.75E-11 | 1.55E-11 | | 0.00922 |
| Purine ribonucleosides degradation | Nucleotide Metabolism & Biosynthesis | *Streptococcus* | *Streptococcus thermophilus* | IFNγ-producing CD4^+^ T Cells | CD4^+^ T Cells | 9.96E-12 | 2.32E-12 | | 0.00927 |
| Superpathway of L-threonine biosynthesis | Amino Acid Biosynthesis | *Escherichia* | *Escherichia coli* | γδ T Cells | Adaptive Immune Cells | 3.54E-11 | 7.62E-12 | | 0.00943 |
| Purine ribonucleosides degradation | Nucleotide Metabolism & Biosynthesis | *Eubacterium* | *Eubacterium ventriosum* | PMDSCs | Innate Immune Cells | 1.09E-10 | 2.72E-11 | | 0.00992 |
| Coenzyme A biosynthesis I (prokaryotic) | Cofactor & Vitamin Biosynthesis | *Coprobacter* | *Coprobacter fastidiosus* | IFNγ-producing CD161^+^ CD4^+^ T Cells | CD4^+^ T Cells | 5.80E-11 | 1.37E-11 | | 0.01006 |
| 5-aminoimidazole ribonucleotide biosynthesis II | Nucleotide Metabolism & Biosynthesis | *Holdemanella* | *Holdemanella biformis* | IL-17A-producing Activated CD69^+^ CD4^+^ T Cells | CD4^+^ T Cells | 1.21E-09 | 2.96E-10 | | 0.01035 |
| Superpathway of 5-aminoimidazole ribonucleotide biosynthesis | Nucleotide Metabolism & Biosynthesis | *Holdemanella* | *Holdemanella biformis* | IL-17A-producing Activated CD69^+^ CD4^+^ T Cells | CD4^+^ T Cells | 1.21E-09 | 2.96E-10 | | 0.01035 |
| Glycogen biosynthesis I (from ADP-D-Glucose) | General Biosynthetic Pathways | *Holdemanella* | *Holdemanella biformis* | TNFα-producing NK Cells | Natural Killer (NK) Cells | 1.15E-09 | 2.86E-10 | | 0.01044 |
| Purine ribonucleosides degradation | Nucleotide Metabolism & Biosynthesis | *Eubacterium* | *Eubacterium ventriosum* | Non-classical Monocytes | Innate Immune Cells | 1.04E-09 | 2.50E-10 | | 0.01054 |
| Guanosine ribonucleotides de novo biosynthesis | Nucleotide Metabolism & Biosynthesis | *Holdemanella* | *Holdemanella biformis* | Non-classical Monocytes | Innate Immune Cells | 4.67E-09 | 1.13E-09 | | 0.01054 |
| Sucrose biosynthesis II | General Biosynthetic Pathways | *Holdemanella* | *Holdemanella biformis* | TNFα-producing NK Cells | Natural Killer (NK) Cells | 1.01E-09 | 2.51E-10 | | 0.01075 |
| Superpathway of coenzyme A biosynthesis III (mammals) | Cofactor & Vitamin Biosynthesis | *Holdemanella* | *Holdemanella biformis* | IL-17A-producing Activated NK Cells | Natural Killer (NK) Cells | 1.36E-08 | 3.29E-09 | | 0.01084 |
| Purine ribonucleosides degradation | Nucleotide Metabolism & Biosynthesis | *Eubacterium* | *Eubacterium ventriosum* | PD-1^+^ Activated HLA-DR^+^ CD4^+^ T Cells | CD4^+^ T Cells | 2.05E-10 | 5.14E-11 | | 0.01085 |
| Chorismate biosynthesis I | General Biosynthetic Pathways | *Holdemanella* | *Holdemanella biformis* | CCR7^+^ Activated NK Cells | Natural Killer (NK) Cells | 4.57E-09 | 1.14E-09 | | 0.01102 |
| 5-aminoimidazole ribonucleotide biosynthesis I | Nucleotide Metabolism & Biosynthesis | *Holdemanella* | *Holdemanella biformis* | TNFα-producing CD161^+^ NK Cells | Natural Killer (NK) Cells | 1.44E-09 | 3.59E-10 | | 0.01107 |
| 5-aminoimidazole ribonucleotide biosynthesis I | Nucleotide Metabolism & Biosynthesis | *Escherichia* | *Escherichia coli* | IL-2-producing CD4^+^ EM T Cells | CD4^+^ T Cells | 1.40E-10 | 3.17E-11 | | 0.01121 |
| Adenine and adenosine salvage III | Nucleotide Metabolism & Biosynthesis | *Coprobacter* | *Coprobacter fastidiosus* | IFNγ-producing CD161^+^ CD4^+^ T Cells | CD4^+^ T Cells | 5.73E-11 | 1.37E-11 | | 0.01145 |
| UMP biosynthesis I | Nucleotide Metabolism & Biosynthesis | *Clostridium* | *Clostridium sp AM22 11AC* | IFNγ-producing T regs | Adaptive Immune Cells | 1.35E-09 | 3.23E-10 | | 0.01162 |
| UMP biosynthesis II | Nucleotide Metabolism & Biosynthesis | *Clostridium* | *Clostridium sp AM22 11AC* | IFNγ-producing T regs | Adaptive Immune Cells | 1.35E-09 | 3.23E-10 | | 0.01162 |
| UMP biosynthesis II | Nucleotide Metabolism & Biosynthesis | *Clostridium* | *Clostridium sp AM22 11AC* | IFNγ-producing T regs | Adaptive Immune Cells | 1.35E-09 | 3.23E-10 | | 0.01162 |
| Superpathway of L-threonine biosynthesis | Amino Acid Biosynthesis | *Holdemanella* | *Holdemanella biformis* | IL-17A-producing Activated NK Cells | Natural Killer (NK) Cells | 1.26E-08 | 3.08E-09 | | 0.01166 |
| L-valine biosynthesis | Amino Acid Biosynthesis | *Streptococcus* | *Streptococcus thermophilus* | IFNγ-producing CD4^+^ T Cells | CD4^+^ T Cells | 9.24E-12 | 2.23E-12 | | 0.01168 |
| S-adenosyl-L-methionine salvage I | Amino Acid Biosynthesis | *Holdemanella* | *Holdemanella biformis* | TNFα-producing NK Cells | Natural Killer (NK) Cells | 1.40E-09 | 3.51E-10 | | 0.01190 |
| L-lysine biosynthesis VI | Amino Acid Biosynthesis | *Coprobacter* | *Coprobacter fastidiosus* | IFNγ-producing CD161^+^ NK Cells | Natural Killer (NK) Cells | 3.36E-10 | 7.08E-11 | | 0.01227 |
| Coenzyme A biosynthesis II (eukaryotic) | Cofactor & Vitamin Biosynthesis | *Holdemanella* | *Holdemanella biformis* | IL-17A-producing Activated NK Cells | Natural Killer (NK) Cells | 1.31E-08 | 3.23E-09 | | 0.01251 |
| Coenzyme A biosynthesis II (eukaryotic) | Cofactor & Vitamin Biosynthesis | *Coprobacter* | *Coprobacter fastidiosus* | IFNγ-producing Activated CD69^+^ CD4^+^ T Cells | CD4^+^ T Cells | 2.39E-11 | 5.78E-12 | | 0.01267 |
| Queuosine biosynthesis I (de novo) | Nucleotide Metabolism & Biosynthesis | *Escherichia* | *Escherichia coli* | γδ T Cells | Adaptive Immune Cells | 3.09E-11 | 6.96E-12 | | 0.01282 |
| 5-aminoimidazole ribonucleotide biosynthesis I | Nucleotide Metabolism & Biosynthesis | *Dialister* | *Dialister invisus* | IFNγ-producing T regs | Adaptive Immune Cells | 2.54E-09 | 6.14E-10 | | 0.01285 |
| Guanosine ribonucleotides de novo biosynthesis | Nucleotide Metabolism & Biosynthesis | *Clostridium* | *Clostridium sp AM22 11AC* | IFNγ-producing T regs | Adaptive Immune Cells | 1.13E-09 | 2.74E-10 | | 0.01296 |
| Sucrose biosynthesis II | General Biosynthetic Pathways | *Holdemanella* | *Holdemanella biformis* | IL-17A-producing CD8^+^ T Cells | CD8^+^ T Cells | 2.12E-09 | 5.10E-10 | | 0.01306 |
| L-valine biosynthesis | Amino Acid Biosynthesis | *Coprobacter* | *Coprobacter fastidiosus* | IFNγ-producing CD161^+^ NK Cells | Natural Killer (NK) Cells | 2.76E-10 | 5.91E-11 | | 0.01343 |
| Inosine 5-phosphate degradation | Nucleotide Metabolism & Biosynthesis | *Escherichia* | *Escherichia coli* | γδ T Cells | Adaptive Immune Cells | 4.20E-11 | 9.56E-12 | | 0.01348 |
| Glycogen biosynthesis I (from ADP-D-Glucose) | General Biosynthetic Pathways | *Holdemanella* | *Holdemanella biformis* | IL-17A-producing CD8^+^ T Cells | CD8^+^ T Cells | 2.41E-09 | 5.82E-10 | | 0.01355 |
| Chorismate biosynthesis from 3-dehydroquinate | General Biosynthetic Pathways | *Clostridium* | *Clostridium sp AM22 11AC* | CCR7^+^ CD8^+^ T Cells | CD8^+^ T Cells | 4.87E-11 | 1.04E-11 | | 0.01375 |
| Chorismate biosynthesis I | General Biosynthetic Pathways | *Clostridium* | *Clostridium sp AM22 11AC* | CCR7^+^ CD8^+^ T Cells | CD8^+^ T Cells | 4.60E-11 | 9.75E-12 | | 0.01375 |
| Fatty acid biosynthesis initiation (mitochondria) | Lipid & Membrane Metabolism | *Clostridium* | *Clostridium sp AM22 11AC* | CCR7^+^ CD8^+^ T Cells | CD8^+^ T Cells | 5.79E-11 | 1.27E-11 | | 0.01375 |
| Flavin biosynthesis I (bacteria and plants) | Cofactor & Vitamin Biosynthesis | *Clostridium* | *Clostridium sp AM22 11AC* | CCR7^+^ CD8^+^ T Cells | CD8^+^ T Cells | 5.02E-11 | 1.05E-11 | | 0.01375 |
| L-lysine biosynthesis VI | Amino Acid Biosynthesis | *Clostridium* | *Clostridium sp AM22 11AC* | CCR7^+^ CD8^+^ T Cells | CD8^+^ T Cells | 3.86E-11 | 8.85E-12 | | 0.01375 |
| Queuosine biosynthesis I (de novo) | Nucleotide Metabolism & Biosynthesis | *Clostridium* | *Clostridium sp AM22 11AC* | CCR7^+^ CD8^+^ T Cells | CD8^+^ T Cells | 4.44E-11 | 1.02E-11 | | 0.01375 |
| Sucrose biosynthesis II | General Biosynthetic Pathways | *Clostridium* | *Clostridium sp AM22 11AC* | CCR7^+^ CD8^+^ T Cells | CD8^+^ T Cells | 6.92E-11 | 1.55E-11 | | 0.01375 |
| Superpathway of coenzyme A biosynthesis III (mammals) | Cofactor & Vitamin Biosynthesis | *Clostridium* | *Clostridium sp AM22 11AC* | CCR7^+^ CD8^+^ T Cells | CD8^+^ T Cells | 3.75E-11 | 8.74E-12 | | 0.01375 |
| 5-aminoimidazole ribonucleotide biosynthesis I | Nucleotide Metabolism & Biosynthesis | *Eubacterium* | *Eubacterium ventriosum* | CCR7^+^ CD8^+^ T Cells | CD8^+^ T Cells | 2.48E-11 | 5.85E-12 | | 0.01375 |
| 5-aminoimidazole ribonucleotide biosynthesis II | Nucleotide Metabolism & Biosynthesis | *Eubacterium* | *Eubacterium ventriosum* | CCR7^+^ CD8^+^ T Cells | CD8^+^ T Cells | 2.35E-11 | 5.58E-12 | | 0.01375 |
| Superpathway of 5-aminoimidazole ribonucleotide biosynthesis | Nucleotide Metabolism & Biosynthesis | *Eubacterium* | *Eubacterium ventriosum* | CCR7^+^ CD8^+^ T Cells | CD8^+^ T Cells | 2.35E-11 | 5.58E-12 | | 0.01375 |
| Guanosine ribonucleotides de novo biosynthesis | Nucleotide Metabolism & Biosynthesis | *Eubacterium* | *Eubacterium ventriosum* | CCR7^+^ CD8^+^ T Cells | CD8^+^ T Cells | 2.24E-11 | 5.01E-12 | | 0.01375 |
| Chorismate biosynthesis I | General Biosynthetic Pathways | *Parabacteroides* | *Parabacteroides goldsteinii* | CCR7^+^ CD8^+^ T Cells | CD8^+^ T Cells | 3.15E-11 | 7.33E-12 | | 0.01375 |
| Superpathway of coenzyme A biosynthesis III (mammals) | Cofactor & Vitamin Biosynthesis | *Parabacteroides* | *Parabacteroides goldsteinii* | CCR7^+^ CD8^+^ T Cells | CD8^+^ T Cells | 3.03E-11 | 7.09E-12 | | 0.01375 |
| Methylerythritol phosphate pathway I | General Biosynthetic Pathways | *Parabacteroides* | *Parabacteroides goldsteinii* | CCR7^+^ CD8^+^ T Cells | CD8^+^ T Cells | 2.78E-11 | 6.61E-12 | | 0.01375 |
| Phosphopantothenate biosynthesis I | Lipid & Membrane Metabolism | *Parabacteroides* | *Parabacteroides goldsteinii* | CCR7^+^ CD8^+^ T Cells | CD8^+^ T Cells | 3.21E-11 | 7.59E-12 | | 0.01375 |
| Peptidoglycan biosynthesis I (meso-diaminopimelate containing) | Cell wall & Peptidoglycan Synthesis | *Parabacteroides* | *Parabacteroides goldsteinii* | CCR7^+^ CD8^+^ T Cells | CD8^+^ T Cells | 3.16E-11 | 7.36E-12 | | 0.01375 |
| Folate transformations II (plants) | Cofactor & Vitamin Biosynthesis | *Parabacteroides* | *Parabacteroides goldsteinii* | CCR7^+^ CD8^+^ T Cells | CD8^+^ T Cells | 3.39E-11 | 8.07E-12 | | 0.01375 |
| UMP biosynthesis I | Nucleotide Metabolism & Biosynthesis | *Parabacteroides* | *Parabacteroides goldsteinii* | CCR7^+^ CD8^+^ T Cells | CD8^+^ T Cells | 3.68E-11 | 8.74E-12 | | 0.01375 |
| 5-aminoimidazole ribonucleotide biosynthesis I | Nucleotide Metabolism & Biosynthesis | *Parabacteroides* | *Parabacteroides goldsteinii* | CCR7^+^ CD8^+^ T Cells | CD8^+^ T Cells | 3.00E-11 | 7.04E-12 | | 0.01375 |
| 5-aminoimidazole ribonucleotide biosynthesis II | Nucleotide Metabolism & Biosynthesis | *Parabacteroides* | *Parabacteroides goldsteinii* | CCR7^+^ CD8^+^ T Cells | CD8^+^ T Cells | 3.16E-11 | 7.35E-12 | | 0.01375 |
| Chorismate biosynthesis from 3-dehydroquinate | General Biosynthetic Pathways | *Parabacteroides* | *Parabacteroides goldsteinii* | CCR7^+^ CD8^+^ T Cells | CD8^+^ T Cells | 3.56E-11 | 8.07E-12 | | 0.01375 |
| Superpathway of 5-aminoimidazole ribonucleotide biosynthesis | Nucleotide Metabolism & Biosynthesis | *Parabacteroides* | *Parabacteroides goldsteinii* | CCR7^+^ CD8^+^ T Cells | CD8^+^ T Cells | 3.16E-11 | 7.35E-12 | | 0.01375 |
| Peptidoglycan biosynthesis III (mycobacteria) | Cell wall & Peptidoglycan Synthesis | *Parabacteroides* | *Parabacteroides goldsteinii* | CCR7^+^ CD8^+^ T Cells | CD8^+^ T Cells | 3.36E-11 | 7.75E-12 | | 0.01375 |
| Adenine and adenosine salvage III | Nucleotide Metabolism & Biosynthesis | *Parabacteroides* | *Parabacteroides goldsteinii* | CCR7^+^ CD8^+^ T Cells | CD8^+^ T Cells | 3.25E-11 | 7.52E-12 | | 0.01375 |
| Queuosine biosynthesis I (de novo) | Nucleotide Metabolism & Biosynthesis | *Parabacteroides* | *Parabacteroides goldsteinii* | CCR7^+^ CD8^+^ T Cells | CD8^+^ T Cells | 3.53E-11 | 8.38E-12 | | 0.01375 |
| UMP biosynthesis II | Nucleotide Metabolism & Biosynthesis | *Parabacteroides* | *Parabacteroides goldsteinii* | CCR7^+^ CD8^+^ T Cells | CD8^+^ T Cells | 3.68E-11 | 8.74E-12 | | 0.01375 |
| Coenzyme A biosynthesis II (eukaryotic) | Cofactor & Vitamin Biosynthesis | *Parabacteroides* | *Parabacteroides goldsteinii* | CCR7^+^ CD8^+^ T Cells | CD8^+^ T Cells | 2.90E-11 | 6.84E-12 | | 0.01375 |
| Flavin biosynthesis I (bacteria and plants) | Cofactor & Vitamin Biosynthesis | *Parabacteroides* | *Parabacteroides goldsteinii* | CCR7^+^ CD8^+^ T Cells | CD8^+^ T Cells | 2.93E-11 | 6.90E-12 | | 0.01375 |
| Guanosine ribonucleotides de novo biosynthesis | Nucleotide Metabolism & Biosynthesis | *Clostridium* | *Clostridium sp AM22 11AC* | CCR7^+^ CD8^+^ T Cells | CD8^+^ T Cells | 4.29E-11 | 1.02E-11 | | 0.01382 |
| UMP biosynthesis I | Nucleotide Metabolism & Biosynthesis | *Holdemanella* | *Holdemanella biformis* | Non-classical Monocytes | Innate Immune Cells | 6.86E-09 | 1.70E-09 | | 0.01382 |
| UMP biosynthesis II | Nucleotide Metabolism & Biosynthesis | *Holdemanella* | *Holdemanella biformis* | Non-classical Monocytes | Innate Immune Cells | 6.86E-09 | 1.70E-09 | | 0.01382 |
| UMP biosynthesis III | Nucleotide Metabolism & Biosynthesis | *Holdemanella* | *Holdemanella biformis* | Non-classical Monocytes | Innate Immune Cells | 6.86E-09 | 1.70E-09 | | 0.01382 |
| L-valine biosynthesis | Amino Acid Biosynthesis | *Escherichia* | *Escherichia coli* | IL-2-producing CD4^+^ EM T Cells | CD4^+^ T Cells | 1.47E-10 | 3.42E-11 | | 0.01410 |
| Coenzyme A biosynthesis I (prokaryotic) | Cofactor & Vitamin Biosynthesis | *Holdemanella* | *Holdemanella biformis* | Non-classical Monocytes | Innate Immune Cells | 5.60E-09 | 1.39E-09 | | 0.01427 |
| Coenzyme A biosynthesis II (eukaryotic) | Cofactor & Vitamin Biosynthesis | *Clostridium* | *Clostridium sp AM22 11AC* | CCR7^+^ CD8^+^ T Cells | CD8^+^ T Cells | 3.64E-11 | 8.75E-12 | | 0.01429 |
| Coenzyme A biosynthesis I (prokaryotic) | Cofactor & Vitamin Biosynthesis | *Parabacteroides* | *Parabacteroides goldsteinii* | CCR7^+^ CD8^+^ T Cells | CD8^+^ T Cells | 2.88E-11 | 6.91E-12 | | 0.01429 |
| L-lysine biosynthesis VI | Amino Acid Biosynthesis | *Parabacteroides* | *Parabacteroides goldsteinii* | CCR7^+^ CD8^+^ T Cells | CD8^+^ T Cells | 2.64E-11 | 6.33E-12 | | 0.01429 |
| UMP biosynthesis I | Nucleotide Metabolism & Biosynthesis | *Holdemanella* | *Holdemanella biformis* | IL-17A-producing CD4^+^ EM T Cells | CD4^+^ T Cells | 3.73E-09 | 8.35E-10 | | 0.01430 |
| UMP biosynthesis II | Nucleotide Metabolism & Biosynthesis | *Holdemanella* | *Holdemanella biformis* | IL-17A-producing CD4^+^ EM T Cells | CD4^+^ T Cells | 3.73E-09 | 8.35E-10 | | 0.01430 |
| UMP biosynthesis III | Nucleotide Metabolism & Biosynthesis | *Holdemanella* | *Holdemanella biformis* | IL-17A-producing CD4^+^ EM T Cells | CD4^+^ T Cells | 3.73E-09 | 8.35E-10 | | 0.01430 |
| Chorismate biosynthesis from 3-dehydroquinate | General Biosynthetic Pathways | *Holdemanella* | *Holdemanella biformis* | CCR7^+^ Activated NK Cells | Natural Killer (NK) Cells | 4.12E-09 | 1.05E-09 | | 0.01439 |
| Coenzyme A biosynthesis II (eukaryotic) | Cofactor & Vitamin Biosynthesis | *Coprobacter* | *Coprobacter fastidiosus* | IFNγ-producing CD161^+^ CD8^+^ T Cells | CD8^+^ T Cells | 1.09E-10 | 2.60E-11 | | 0.01445 |
| UMP biosynthesis III | Nucleotide Metabolism & Biosynthesis | *Parabacteroides* | *Parabacteroides goldsteinii* | CCR7^+^ CD8^+^ T Cells | CD8^+^ T Cells | 3.70E-11 | 8.91E-12 | | 0.01459 |
| S-adenosyl-L-methionine salvage I | Amino Acid Biosynthesis | *Streptococcus* | *Streptococcus thermophilus* | IFNγ-producing CD4^+^ T Cells | CD4^+^ T Cells | 1.02E-11 | 2.53E-12 | | 0.01465 |
| tRNA charging | Amino Acid Biosynthesis | *Parabacteroides* | *Parabacteroides goldsteinii* | CCR7^+^ CD8^+^ T Cells | CD8^+^ T Cells | 2.73E-11 | 6.58E-12 | | 0.01482 |
| Coenzyme A biosynthesis I (prokaryotic) | Cofactor & Vitamin Biosynthesis | *Dialister* | *Dialister invisus* | IFNγ-producing T regs | Adaptive Immune Cells | 2.13E-09 | 5.26E-10 | | 0.01485 |
| UMP biosynthesis I | Nucleotide Metabolism & Biosynthesis | *Dialister* | *Dialister invisus* | IFNγ-producing T regs | Adaptive Immune Cells | 2.08E-09 | 5.14E-10 | | 0.01485 |
| UMP biosynthesis II | Nucleotide Metabolism & Biosynthesis | *Dialister* | *Dialister invisus* | IFNγ-producing T regs | Adaptive Immune Cells | 2.08E-09 | 5.14E-10 | | 0.01485 |
| UMP biosynthesis III | Nucleotide Metabolism & Biosynthesis | *Dialister* | *Dialister invisus* | IFNγ-producing T regs | Adaptive Immune Cells | 2.08E-09 | 5.14E-10 | | 0.01485 |
| Fatty acid biosynthesis initiation (mitochondria) | Lipid & Membrane Metabolism | *Parabacteroides* | *Parabacteroides goldsteinii* | CCR7^+^ CD8^+^ T Cells | CD8^+^ T Cells | 2.62E-11 | 6.33E-12 | | 0.01486 |
| Coenzyme A biosynthesis I (prokaryotic) | Cofactor & Vitamin Biosynthesis | *Coprobacter* | *Coprobacter fastidiosus* | IFNγ/TNFα-producing CD161^+^ CD8^+^ T Cells | CD8^+^ T Cells | 4.54E-10 | 1.11E-10 | | 0.01539 |
| Guanosine ribonucleotides de novo biosynthesis | Nucleotide Metabolism & Biosynthesis | *Parabacteroides* | *Parabacteroides goldsteinii* | CCR7^+^ CD8^+^ T Cells | CD8^+^ T Cells | 3.62E-11 | 8.79E-12 | | 0.01552 |
| Sucrose biosynthesis II | General Biosynthetic Pathways | *Eubacterium* | *Eubacterium ventriosum* | CCR7^+^ CD8^+^ T Cells | CD8^+^ T Cells | 1.82E-11 | 4.41E-12 | | 0.01556 |
| Coenzyme A biosynthesis II (eukaryotic) | Cofactor & Vitamin Biosynthesis | *Coprobacter* | *Coprobacter fastidiosus* | IFNγ/TNFα-producing CD8^+^ T Cells | CD8^+^ T Cells | 6.72E-11 | 1.64E-11 | | 0.01556 |
| Superpathway of coenzyme A biosynthesis III (mammals) | Cofactor & Vitamin Biosynthesis | *Coprobacter* | *Coprobacter fastidiosus* | IFNγ-producing Activated NK Cells | Natural Killer (NK) Cells | 1.42E-10 | 3.01E-11 | | 0.01562 |
| Sucrose biosynthesis II | General Biosynthetic Pathways | *Holdemanella* | *Holdemanella biformis* | TNFα-producing CD161^+^ NK Cells | Natural Killer (NK) Cells | 9.39E-10 | 2.42E-10 | | 0.01568 |
| Adenine and adenosine salvage III | Nucleotide Metabolism & Biosynthesis | *Coprobacter* | *Coprobacter fastidiosus* | IFNγ-producing T regs | Adaptive Immune Cells | 8.35E-10 | 2.07E-10 | | 0.01574 |
| Sucrose biosynthesis II | General Biosynthetic Pathways | *Holdemanella* | *Holdemanella biformis* | IL-17A-producing CD161^+^ CD8^+^ T Cells | CD8^+^ T Cells | 7.66E-09 | 1.91E-09 | | 0.01575 |
| Glycogen biosynthesis I (from ADP-D-Glucose) | General Biosynthetic Pathways | *Holdemanella* | *Holdemanella biformis* | TNFα-producing CD161^+^ NK Cells | Natural Killer (NK) Cells | 1.07E-09 | 2.76E-10 | | 0.01578 |
| L-valine biosynthesis | Amino Acid Biosynthesis | *Eubacterium* | *Eubacterium ventriosum* | TNFα-producing CD161^+^ NK Cells | Natural Killer (NK) Cells | 1.92E-10 | 4.98E-11 | | 0.01623 |
| L-histidine biosynthesis | Amino Acid Biosynthesis | *Parabacteroides* | *Parabacteroides goldsteinii* | CCR7^+^ CD8^+^ T Cells | CD8^+^ T Cells | 2.44E-11 | 5.95E-12 | | 0.01632 |
| 5-aminoimidazole ribonucleotide biosynthesis II | Nucleotide Metabolism & Biosynthesis | *Escherichia* | *Escherichia coli* | IL-2-producing CD4^+^ EM T Cells | CD4^+^ T Cells | 1.50E-10 | 3.55E-11 | | 0.01638 |
| Superpathway of 5-aminoimidazole ribonucleotide biosynthesis | Nucleotide Metabolism & Biosynthesis | *Escherichia* | *Escherichia coli* | IL-2-producing CD4^+^ EM T Cells | CD4^+^ T Cells | 1.50E-10 | 3.55E-11 | | 0.01638 |
| 5-aminoimidazole ribonucleotide biosynthesis I | Nucleotide Metabolism & Biosynthesis | *Holdemanella* | *Holdemanella biformis* | IL-17A-producing Activated CD69^+^ CD4^+^ T Cells | CD4^+^ T Cells | 1.09E-09 | 2.78E-10 | | 0.01640 |
| Glycogen biosynthesis I (from ADP-D-Glucose) | General Biosynthetic Pathways | *Holdemanella* | *Holdemanella biformis* | IL-17A-producing CD161^+^ CD8^+^ T Cells | CD8^+^ T Cells | 8.70E-09 | 2.18E-09 | | 0.01668 |
| Folate transformations II (plants) | Cofactor & Vitamin Biosynthesis | *Coprobacter* | *Coprobacter fastidiosus* | IFNγ-producing T regs | Adaptive Immune Cells | 7.55E-10 | 1.88E-10 | | 0.01671 |
| L-valine biosynthesis | Amino Acid Biosynthesis | *Eubacterium* | *Eubacterium ventriosum* | IL-17A-producing Activated CD69^+^ CD4^+^ T Cells | CD4^+^ T Cells | 1.49E-10 | 3.83E-11 | | 0.01689 |
| Coenzyme A biosynthesis II (eukaryotic) | Cofactor & Vitamin Biosynthesis | *Coprobacter* | *Coprobacter fastidiosus* | IFNγ/TNFα-producing CD69^+^ CD4^+^ T Cells | CD4^+^ T Cells | 3.90E-11 | 9.43E-12 | | 0.01694 |
| CDP diacylglycerol biosynthesis I | Lipid & Membrane Metabolism | *Clostridium* | *Clostridium sp AM22 11AC* | IFNγ-producing T regs | Adaptive Immune Cells | 9.08E-10 | 2.27E-10 | | 0.01696 |
| CDP diacylglycerol biosynthesis II | Lipid & Membrane Metabolism | *Clostridium* | *Clostridium sp AM22 11AC* | IFNγ-producing T regs | Adaptive Immune Cells | 9.08E-10 | 2.27E-10 | | 0.01696 |
| Guanosine ribonucleotides de novo biosynthesis | Nucleotide Metabolism & Biosynthesis | *Eubacterium* | *Eubacterium ventriosum* | CD8^+^ CM | CD8^+^ T Cells | 6.53E-11 | 1.51E-11 | | 0.01705 |
| L-valine biosynthesis | Amino Acid Biosynthesis | *Parabacteroides* | *Parabacteroides goldsteinii* | CCR7^+^ CD8^+^ T Cells | CD8^+^ T Cells | 2.82E-11 | 6.91E-12 | | 0.01719 |
| Inosine 5-phosphate degradation | Nucleotide Metabolism & Biosynthesis | *Eubacterium* | *Eubacterium ventriosum* | Non-classical Monocytes | Innate Immune Cells | 7.57E-10 | 1.93E-10 | | 0.01724 |
| S-adenosyl-L-methionine salvage I | Amino Acid Biosynthesis | *Holdemanella* | *Holdemanella biformis* | TNFα-producing CD161^+^ NK Cells | Natural Killer (NK) Cells | 1.30E-09 | 3.39E-10 | | 0.01736 |
| Chorismate biosynthesis I | General Biosynthetic Pathways | *Clostridium* | *Clostridium sp AM22 11AC* | IFNγ-producing T regs | Adaptive Immune Cells | 1.09E-09 | 2.74E-10 | | 0.01764 |
| Coenzyme A biosynthesis I (prokaryotic) | Cofactor & Vitamin Biosynthesis | *Coprobacter* | *Coprobacter fastidiosus* | IFNγ-producing CD4^+^ T Cells | CD4^+^ T Cells | 1.79E-11 | 4.55E-12 | | 0.01790 |
| Coenzyme A biosynthesis I (prokaryotic) | Cofactor & Vitamin Biosynthesis | *Coprobacter* | *Coprobacter fastidiosus* | IFNγ/TNFα-producing CD161^+^ CD4^+^ T Cells | CD4^+^ T Cells | 9.66E-11 | 2.37E-11 | | 0.01805 |
| UMP biosynthesis I | Nucleotide Metabolism & Biosynthesis | *Eubacterium* | *Eubacterium ventriosum* | IL-17A-producing Activated CD69^+^ CD4^+^ T Cells | CD4^+^ T Cells | 1.21E-10 | 3.14E-11 | | 0.01807 |
| UMP biosynthesis II | Nucleotide Metabolism & Biosynthesis | *Eubacterium* | *Eubacterium ventriosum* | IL-17A-producing Activated CD69^+^ CD4^+^ T Cells | CD4^+^ T Cells | 1.21E-10 | 3.14E-11 | | 0.01807 |
| UMP biosynthesis III | Nucleotide Metabolism & Biosynthesis | *Eubacterium* | *Eubacterium ventriosum* | IL-17A-producing Activated CD69^+^ CD4^+^ T Cells | CD4^+^ T Cells | 1.21E-10 | 3.14E-11 | | 0.01807 |
| Purine ribonucleosides degradation | Nucleotide Metabolism & Biosynthesis | *Eubacterium* | *Eubacterium ventriosum* | TNFα-producing gd T Cells | Adaptive Immune Cells | 4.00E-11 | 1.05E-11 | | 0.01815 |
| Inosine 5-phosphate degradation | Nucleotide Metabolism & Biosynthesis | *Coprobacter* | *Coprobacter fastidiosus* | IFNγ-producing CD161^+^ CD4^+^ T Cells | CD4^+^ T Cells | 4.58E-11 | 1.15E-11 | | 0.01843 |
| Queuosine biosynthesis I (de novo) | Nucleotide Metabolism & Biosynthesis | *Clostridium* | *Clostridium sp AM22 11AC* | IFNγ-producing T regs | Adaptive Immune Cells | 1.10E-09 | 2.79E-10 | | 0.01881 |
| Guanosine ribonucleotides de novo biosynthesis | Nucleotide Metabolism & Biosynthesis | *Holdemanella* | *Holdemanella biformis* | IL-17A-producing Activated NK Cells | Natural Killer (NK) Cells | 1.13E-08 | 2.88E-09 | | 0.01881 |
| Superpathway of coenzyme A biosynthesis III (mammals) | Cofactor & Vitamin Biosynthesis | *Coprobacter* | *Coprobacter fastidiosus* | IFNγ-producing NK Cells | Natural Killer (NK) Cells | 1.40E-10 | 2.99E-11 | | 0.01885 |
| L-valine biosynthesis | Amino Acid Biosynthesis | *Eubacterium* | *Eubacterium ventriosum* | IL-17A-producing CD4^+^ T Cells | CD4^+^ T Cells | 1.39E-10 | 3.64E-11 | | 0.01886 |
| 5-aminoimidazole ribonucleotide biosynthesis II | Nucleotide Metabolism & Biosynthesis | *Dialister* | *Dialister invisus* | IFNγ-producing T regs | Adaptive Immune Cells | 2.54E-09 | 6.45E-10 | | 0.01888 |
| Superpathway of 5-aminoimidazole ribonucleotide biosynthesis | Nucleotide Metabolism & Biosynthesis | *Dialister* | *Dialister invisus* | IFNγ-producing T regs | Adaptive Immune Cells | 2.54E-09 | 6.45E-10 | | 0.01888 |
| UMP biosynthesis I | Nucleotide Metabolism & Biosynthesis | *Clostridium* | *Clostridium sp AM22 11AC* | CD8^+^ CM | CD8^+^ T Cells | 1.54E-10 | 3.60E-11 | | 0.01892 |
| UMP biosynthesis II | Nucleotide Metabolism & Biosynthesis | *Clostridium* | *Clostridium sp AM22 11AC* | CD8^+^ CM | CD8^+^ T Cells | 1.54E-10 | 3.60E-11 | | 0.01892 |
| UMP biosynthesis II | Nucleotide Metabolism & Biosynthesis | *Clostridium* | *Clostridium sp AM22 11AC* | CD8^+^ CM | CD8^+^ T Cells | 1.54E-10 | 3.60E-11 | | 0.01892 |
| Coenzyme A biosynthesis I (prokaryotic) | Cofactor & Vitamin Biosynthesis | *Eubacterium* | *Eubacterium ventriosum* | Non-classical Monocytes | Innate Immune Cells | 8.70E-10 | 2.24E-10 | | 0.01892 |
| Inosine 5-phosphate degradation | Nucleotide Metabolism & Biosynthesis | *Dialister* | *Dialister invisus* | IFNγ-producing T regs | Adaptive Immune Cells | 1.89E-09 | 4.79E-10 | | 0.01916 |
| Superpathway of coenzyme A biosynthesis III (mammals) | Cofactor & Vitamin Biosynthesis | *Coprobacter* | *Coprobacter fastidiosus* | IFNγ-producing NK T Cells | Adaptive Immune Cells | 3.71E-11 | 9.34E-12 | | 0.01924 |
| Glycogen biosynthesis I (from ADP-D-Glucose) | General Biosynthetic Pathways | *Eubacterium* | *Eubacterium ventriosum* | IL-17A-producing CD4^+^ T Cells | CD4^+^ T Cells | 1.16E-10 | 3.05E-11 | | 0.01965 |
| UMP biosynthesis I | Nucleotide Metabolism & Biosynthesis | *Dialister* | *Dialister invisus* | IL-2/TNFα-producing CD4^+^ T Cells | CD4^+^ T Cells | 1.76E-10 | 3.97E-11 | | 0.01973 |
| UMP biosynthesis II | Nucleotide Metabolism & Biosynthesis | *Dialister* | *Dialister invisus* | IL-2/TNFα-producing CD4^+^ T Cells | CD4^+^ T Cells | 1.76E-10 | 3.97E-11 | | 0.01973 |
| UMP biosynthesis III | Nucleotide Metabolism & Biosynthesis | *Dialister* | *Dialister invisus* | IL-2/TNFα-producing CD4^+^ T Cells | CD4^+^ T Cells | 1.76E-10 | 3.97E-11 | | 0.01973 |
| Adenine and adenosine salvage III | Nucleotide Metabolism & Biosynthesis | *Escherichia* | *Escherichia coli* | IL-2-producing CD4^+^ EM T Cells | CD4^+^ T Cells | 1.34E-10 | 3.27E-11 | | 0.01975 |
| UMP biosynthesis I | Nucleotide Metabolism & Biosynthesis | *Eubacterium* | *Eubacterium ventriosum* | IL-17A-producing CD4^+^ T Cells | CD4^+^ T Cells | 1.13E-10 | 2.98E-11 | | 0.01983 |
| UMP biosynthesis II | Nucleotide Metabolism & Biosynthesis | *Eubacterium* | *Eubacterium ventriosum* | IL-17A-producing CD4^+^ T Cells | CD4^+^ T Cells | 1.13E-10 | 2.98E-11 | | 0.01983 |
| UMP biosynthesis III | Nucleotide Metabolism & Biosynthesis | *Eubacterium* | *Eubacterium ventriosum* | IL-17A-producing CD4^+^ T Cells | CD4^+^ T Cells | 1.13E-10 | 2.98E-11 | | 0.01983 |
| Folate transformations II (plants) | Cofactor & Vitamin Biosynthesis | *Coprobacter* | *Coprobacter fastidiosus* | IFNγ-producing CD161^+^ CD4^+^ T Cells | CD4^+^ T Cells | 5.01E-11 | 1.26E-11 | | 0.02007 |
| tRNA charging | Amino Acid Biosynthesis | *Holdemanella* | *Holdemanella biformis* | CCR7^+^ Activated NK Cells | Natural Killer (NK) Cells | 3.67E-09 | 9.67E-10 | | 0.02014 |
| Purine ribonucleosides degradation | Nucleotide Metabolism & Biosynthesis | *Eubacterium* | *Eubacterium ventriosum* | IL-17A-producing Activated CD69^+^ CD4^+^ T Cells | CD4^+^ T Cells | 1.53E-10 | 4.00E-11 | | 0.02019 |
| Glycogen biosynthesis I (from ADP-D-Glucose) | General Biosynthetic Pathways | *Eubacterium* | *Eubacterium ventriosum* | IL-17A-producing Activated CD69^+^ CD4^+^ T Cells | CD4^+^ T Cells | 1.23E-10 | 3.22E-11 | | 0.02021 |
| Inosine 5-phosphate degradation | Nucleotide Metabolism & Biosynthesis | *Eubacterium* | *Eubacterium ventriosum* | CD8^+^ CM | CD8^+^ T Cells | 5.69E-11 | 1.34E-11 | | 0.02033 |
| 5-aminoimidazole ribonucleotide biosynthesis II | Nucleotide Metabolism & Biosynthesis | *Holdemanella* | *Holdemanella biformis* | IL-17A-producing CD4^+^ EM T Cells | CD4^+^ T Cells | 4.08E-09 | 9.41E-10 | | 0.02044 |
| Superpathway of 5-aminoimidazole ribonucleotide biosynthesis | Nucleotide Metabolism & Biosynthesis | *Holdemanella* | *Holdemanella biformis* | IL-17A-producing CD4^+^ EM T Cells | CD4^+^ T Cells | 4.08E-09 | 9.41E-10 | | 0.02044 |
| Glycogen biosynthesis I (from ADP-D-Glucose) | General Biosynthetic Pathways | *Holdemanella* | *Holdemanella biformis* | IL-17A-producing Activated CD69^+^ CD4^+^ T Cells | CD4^+^ T Cells | 8.15E-10 | 2.14E-10 | | 0.02044 |
| Folate transformations II (plants) | Cofactor & Vitamin Biosynthesis | *Clostridium* | *Clostridium sp AM22 11AC* | IFNγ-producing T regs | Adaptive Immune Cells | 1.19E-09 | 3.06E-10 | | 0.02063 |
| Superpathway of coenzyme A biosynthesis III (mammals) | Cofactor & Vitamin Biosynthesis | *Clostridium* | *Clostridium sp AM22 11AC* | IFNγ-producing T regs | Adaptive Immune Cells | 9.33E-10 | 2.39E-10 | | 0.02063 |
| Superpathway of coenzyme A biosynthesis III (mammals) | Cofactor & Vitamin Biosynthesis | *Dialister* | *Dialister invisus* | IFNγ-producing T regs | Adaptive Immune Cells | 1.99E-09 | 5.12E-10 | | 0.02063 |
| UMP biosynthesis I | Nucleotide Metabolism & Biosynthesis | *Eubacterium* | *Eubacterium ventriosum* | CD8^+^ CM | CD8^+^ T Cells | 5.87E-11 | 1.39E-11 | | 0.02075 |
| UMP biosynthesis II | Nucleotide Metabolism & Biosynthesis | *Eubacterium* | *Eubacterium ventriosum* | CD8^+^ CM | CD8^+^ T Cells | 5.87E-11 | 1.39E-11 | | 0.02075 |
| UMP biosynthesis III | Nucleotide Metabolism & Biosynthesis | *Eubacterium* | *Eubacterium ventriosum* | CD8^+^ CM | CD8^+^ T Cells | 5.87E-11 | 1.39E-11 | | 0.02075 |
| Coenzyme A biosynthesis I (prokaryotic) | Cofactor & Vitamin Biosynthesis | *Coprobacter* | *Coprobacter fastidiosus* | IFNγ-producing Activated CD69^+^ CD4^+^ T Cells | CD4^+^ T Cells | 2.49E-11 | 6.28E-12 | | 0.02106 |
| Folate transformations II (plants) | Cofactor & Vitamin Biosynthesis | *Dialister* | *Dialister invisus* | IFNγ-producing T regs | Adaptive Immune Cells | 2.07E-09 | 5.33E-10 | | 0.02111 |
| Purine ribonucleosides degradation | Nucleotide Metabolism & Biosynthesis | *Eubacterium* | *Eubacterium ventriosum* | IL-17A-producing CD4^+^ T Cells | CD4^+^ T Cells | 1.43E-10 | 3.79E-11 | | 0.02129 |
| Coenzyme A biosynthesis II (eukaryotic) | Cofactor & Vitamin Biosynthesis | *Dialister* | *Dialister invisus* | IFNγ-producing T regs | Adaptive Immune Cells | 1.92E-09 | 4.98E-10 | | 0.02168 |
| Sucrose biosynthesis II | General Biosynthetic Pathways | *Holdemanella* | *Holdemanella biformis* | IL-17A-producing Activated CD69^+^ CD4^+^ T Cells | CD4^+^ T Cells | 7.10E-10 | 1.88E-10 | | 0.02177 |
| 5-aminoimidazole ribonucleotide biosynthesis I | Nucleotide Metabolism & Biosynthesis | *Coprobacter* | *Coprobacter fastidiosus* | IFNγ-producing Activated NK Cells | Natural Killer (NK) Cells | 1.90E-10 | 4.13E-11 | | 0.02213 |
| Glycogen biosynthesis I (from ADP-D-Glucose) | General Biosynthetic Pathways | *Eubacterium* | *Eubacterium ventriosum* | Non-classical Monocytes | Innate Immune Cells | 7.85E-10 | 2.06E-10 | | 0.02215 |
| Fatty acid biosynthesis initiation (mitochondria) | Lipid & Membrane Metabolism | *Eubacterium* | *Eubacterium ventriosum* | CCR7^+^ CD8^+^ T Cells | CD8^+^ T Cells | 1.95E-11 | 4.95E-12 | | 0.02222 |
| Adenine and adenosine salvage III | Nucleotide Metabolism & Biosynthesis | *Clostridium* | *Clostridium sp AM22 11AC* | CCR7^+^ CD8^+^ T Cells | CD8^+^ T Cells | 4.82E-11 | 1.23E-11 | | 0.02239 |
| CDP diacylglycerol biosynthesis I | Lipid & Membrane Metabolism | *Clostridium* | *Clostridium sp AM22 11AC* | CCR7^+^ CD8^+^ T Cells | CD8^+^ T Cells | 3.36E-11 | 8.56E-12 | | 0.02239 |
| CDP diacylglycerol biosynthesis II | Lipid & Membrane Metabolism | *Clostridium* | *Clostridium sp AM22 11AC* | CCR7^+^ CD8^+^ T Cells | CD8^+^ T Cells | 3.36E-11 | 8.56E-12 | | 0.02239 |
| Methylerythritol phosphate pathway I | General Biosynthetic Pathways | *Clostridium* | *Clostridium sp AM22 11AC* | CCR7^+^ CD8^+^ T Cells | CD8^+^ T Cells | 3.66E-11 | 9.31E-12 | | 0.02239 |
| CDP diacylglycerol biosynthesis I | Lipid & Membrane Metabolism | *Coprobacter* | *Coprobacter fastidiosus* | IFNγ/TNFα-producing CD161^+^ CD8^+^ T Cells | CD8^+^ T Cells | 4.12E-10 | 1.05E-10 | | 0.02242 |
| CDP diacylglycerol biosynthesis II | Lipid & Membrane Metabolism | *Coprobacter* | *Coprobacter fastidiosus* | IFNγ/TNFα-producing CD161^+^ CD8^+^ T Cells | CD8^+^ T Cells | 4.12E-10 | 1.05E-10 | | 0.02242 |
| CDP diacylglycerol biosynthesis I | Lipid & Membrane Metabolism | *Coprobacter* | *Coprobacter fastidiosus* | IFNγ-producing CD161^+^ CD4^+^ T Cells | CD4^+^ T Cells | 5.10E-11 | 1.30E-11 | | 0.02274 |
| CDP diacylglycerol biosynthesis II | Lipid & Membrane Metabolism | *Coprobacter* | *Coprobacter fastidiosus* | IFNγ-producing CD161^+^ CD4^+^ T Cells | CD4^+^ T Cells | 5.10E-11 | 1.30E-11 | | 0.02274 |
| Adenine and adenosine salvage III | Nucleotide Metabolism & Biosynthesis | *Coprobacter* | *Coprobacter fastidiosus* | IFNγ/TNFα-producing CD161^+^ CD8^+^ T Cells | CD8^+^ T Cells | 4.41E-10 | 1.12E-10 | | 0.02289 |
| UMP biosynthesis I | Nucleotide Metabolism & Biosynthesis | *Dialister* | *Dialister invisus* | IFNγ-producing CD4^+^ T Cells | CD4^+^ T Cells | 4.35E-11 | 1.14E-11 | | 0.02289 |
| UMP biosynthesis II | Nucleotide Metabolism & Biosynthesis | *Dialister* | *Dialister invisus* | IFNγ-producing CD4^+^ T Cells | CD4^+^ T Cells | 4.35E-11 | 1.14E-11 | | 0.02289 |
| UMP biosynthesis III | Nucleotide Metabolism & Biosynthesis | *Dialister* | *Dialister invisus* | IFNγ-producing CD4^+^ T Cells | CD4^+^ T Cells | 4.35E-11 | 1.14E-11 | | 0.02289 |
| Glycogen biosynthesis I (from ADP-D-Glucose) | General Biosynthetic Pathways | *Clostridium* | *Clostridium sp AM22 11AC* | Non-classical Monocytes | Innate Immune Cells | 3.49E-09 | 9.17E-10 | | 0.02293 |
| 5-aminoimidazole ribonucleotide biosynthesis I | Nucleotide Metabolism & Biosynthesis | *Eubacterium* | *Eubacterium ventriosum* | IL-17A-producing Activated CD69^+^ CD4^+^ T Cells | CD4^+^ T Cells | 1.49E-10 | 3.97E-11 | | 0.02297 |
| Guanosine ribonucleotides de novo biosynthesis | Nucleotide Metabolism & Biosynthesis | *Eubacterium* | *Eubacterium ventriosum* | Non-classical Monocytes | Innate Immune Cells | 8.36E-10 | 2.20E-10 | | 0.02313 |
| Purine ribonucleosides degradation | Nucleotide Metabolism & Biosynthesis | *Eubacterium* | *Eubacterium ventriosum* | TNFα-producing CD161^+^ NK Cells | Natural Killer (NK) Cells | 1.95E-10 | 5.22E-11 | | 0.02321 |
| 5-aminoimidazole ribonucleotide biosynthesis I | Nucleotide Metabolism & Biosynthesis | *Dialister* | *Dialister invisus* | IFNγ-producing CD4^+^ T Cells | CD4^+^ T Cells | 5.22E-11 | 1.37E-11 | | 0.02331 |
| S-adenosyl-L-methionine salvage I | Amino Acid Biosynthesis | *Holdemanella* | *Holdemanella biformis* | IL-17A-producing Activated CD69^+^ CD4^+^ T Cells | CD4^+^ T Cells | 9.85E-10 | 2.62E-10 | | 0.02342 |
| Coenzyme A biosynthesis I (prokaryotic) | Cofactor & Vitamin Biosynthesis | *Clostridium* | *Clostridium sp AM22 11AC* | CCR7^+^ CD8^+^ T Cells | CD8^+^ T Cells | 3.66E-11 | 9.37E-12 | | 0.02359 |
| Methylerythritol phosphate pathway I | General Biosynthetic Pathways | *Clostridium* | *Clostridium sp AM22 11AC* | IFNγ-producing T regs | Adaptive Immune Cells | 9.54E-10 | 2.50E-10 | | 0.02377 |
| 5-aminoimidazole ribonucleotide biosynthesis II | Nucleotide Metabolism & Biosynthesis | *Coprobacter* | *Coprobacter fastidiosus* | IFNγ-producing Activated NK Cells | Natural Killer (NK) Cells | 1.76E-10 | 3.87E-11 | | 0.02387 |
| Superpathway of 5-aminoimidazole ribonucleotide biosynthesis | Nucleotide Metabolism & Biosynthesis | *Coprobacter* | *Coprobacter fastidiosus* | IFNγ-producing Activated NK Cells | Natural Killer (NK) Cells | 1.76E-10 | 3.87E-11 | | 0.02387 |
| Adenine and adenosine salvage III | Nucleotide Metabolism & Biosynthesis | *Coprobacter* | *Coprobacter fastidiosus* | IFNγ-producing Activated CD69^+^ CD4^+^ T Cells | CD4^+^ T Cells | 2.46E-11 | 6.29E-12 | | 0.02403 |
| Guanosine ribonucleotides de novo biosynthesis | Nucleotide Metabolism & Biosynthesis | *Dialister* | *Dialister invisus* | IFNγ-producing T regs | Adaptive Immune Cells | 2.16E-09 | 5.65E-10 | | 0.02413 |
| Fatty acid biosynthesis initiation (mitochondria) | Lipid & Membrane Metabolism | *Eubacterium* | *Eubacterium ventriosum* | IFNγ-producing T regs | Adaptive Immune Cells | 5.07E-10 | 1.33E-10 | | 0.02413 |
| Adenine and adenosine salvage III | Nucleotide Metabolism & Biosynthesis | *Coprobacter* | *Coprobacter fastidiosus* | IFNγ/TNFα-producing CD161^+^ CD4^+^ T Cells | CD4^+^ T Cells | 9.44E-11 | 2.38E-11 | | 0.02419 |
| Glycogen biosynthesis I (from ADP-D-Glucose) | General Biosynthetic Pathways | *Eubacterium* | *Eubacterium ventriosum* | TNFα-producing T regs | Adaptive Immune Cells | 7.21E-11 | 1.96E-11 | | 0.02462 |
| UMP biosynthesis I | Nucleotide Metabolism & Biosynthesis | *Dialister* | *Dialister invisus* | IL-2-producing CD161^+^ CD8^+^ T Cells | CD8^+^ T Cells | 4.87E-09 | 1.13E-09 | | 0.02466 |
| UMP biosynthesis II | Nucleotide Metabolism & Biosynthesis | *Dialister* | *Dialister invisus* | IL-2-producing CD161^+^ CD8^+^ T Cells | CD8^+^ T Cells | 4.87E-09 | 1.13E-09 | | 0.02466 |
| UMP biosynthesis III | Nucleotide Metabolism & Biosynthesis | *Dialister* | *Dialister invisus* | IL-2-producing CD161^+^ CD8^+^ T Cells | CD8^+^ T Cells | 4.87E-09 | 1.13E-09 | | 0.02466 |
| Glycogen biosynthesis I (from ADP-D-Glucose) | General Biosynthetic Pathways | *Eubacterium* | *Eubacterium ventriosum* | PD-1^+^ CD4^+^ T Cells | CD4^+^ T Cells | 3.40E-11 | 9.22E-12 | | 0.02499 |
| Glycogen biosynthesis I (from ADP-D-Glucose) | General Biosynthetic Pathways | *Escherichia* | *Escherichia coli* | IL-2-producing CD161^+^ CD4^+^ T Cells | CD4^+^ T Cells | 1.96E-10 | 4.23E-11 | | 0.02507 |
| Fatty acid biosynthesis initiation (mitochondria) | Lipid & Membrane Metabolism | *Escherichia* | *Escherichia coli* | IL-2-producing CD161^+^ CD4^+^ T Cells | CD4^+^ T Cells | 2.57E-10 | 5.75E-11 | | 0.02507 |
| Superpathway of L-threonine biosynthesis | Amino Acid Biosynthesis | *Escherichia* | *Escherichia coli* | IL-2-producing CD161^+^ CD4^+^ T Cells | CD4^+^ T Cells | 2.28E-10 | 5.17E-11 | | 0.02507 |
| Adenine and adenosine salvage III | Nucleotide Metabolism & Biosynthesis | *Escherichia* | *Escherichia coli* | γδ T Cells | Adaptive Immune Cells | 2.85E-11 | 7.10E-12 | | 0.02536 |
| Queuosine biosynthesis I (de novo) | Nucleotide Metabolism & Biosynthesis | *Escherichia* | *Escherichia coli* | IL-2-producing CD161^+^ CD4^+^ T Cells | CD4^+^ T Cells | 2.02E-10 | 4.69E-11 | | 0.02551 |
| Inosine 5-phosphate degradation | Nucleotide Metabolism & Biosynthesis | *Coprobacter* | *Coprobacter fastidiosus* | IFNγ-producing T regs | Adaptive Immune Cells | 6.58E-10 | 1.74E-10 | | 0.02564 |
| Glycogen biosynthesis I (from ADP-D-Glucose) | General Biosynthetic Pathways | *Eubacterium* | *Eubacterium ventriosum* | PMDSCs | Innate Immune Cells | 8.22E-11 | 2.24E-11 | | 0.02595 |
| Chorismate biosynthesis I | General Biosynthetic Pathways | *Holdemanella* | *Holdemanella biformis* | IL-17A-producing CD4^+^ EM T Cells | CD4^+^ T Cells | 2.86E-09 | 6.78E-10 | | 0.02606 |
| 5-aminoimidazole ribonucleotide biosynthesis I | Nucleotide Metabolism & Biosynthesis | *Holdemanella* | *Holdemanella biformis* | IL-17A-producing CD4^+^ EM T Cells | CD4^+^ T Cells | 3.64E-09 | 8.89E-10 | | 0.02606 |
| S-adenosyl-L-methionine salvage I | Amino Acid Biosynthesis | *Holdemanella* | *Holdemanella biformis* | IL-17A-producing CD4^+^ EM T Cells | CD4^+^ T Cells | 3.46E-09 | 8.23E-10 | | 0.02606 |
| Chorismate biosynthesis from 3-dehydroquinate | General Biosynthetic Pathways | *Holdemanella* | *Holdemanella biformis* | IL-17A-producing CD4^+^ EM T Cells | CD4^+^ T Cells | 2.55E-09 | 6.29E-10 | | 0.02606 |
| UDP-N-acetylmuramoyl-pentapeptide biosynthesis II (lysine-containing) | Cell wall & Peptidoglycan Synthesis | *Holdemanella* | *Holdemanella biformis* | IL-17A-producing CD4^+^ EM T Cells | CD4^+^ T Cells | 2.66E-09 | 6.31E-10 | | 0.02606 |
| Guanosine ribonucleotides de novo biosynthesis | Nucleotide Metabolism & Biosynthesis | *Holdemanella* | *Holdemanella biformis* | IL-17A-producing CD4^+^ EM T Cells | CD4^+^ T Cells | 2.39E-09 | 5.68E-10 | | 0.02606 |
| tRNA charging | Amino Acid Biosynthesis | *Holdemanella* | *Holdemanella biformis* | IL-17A-producing CD4^+^ EM T Cells | CD4^+^ T Cells | 2.35E-09 | 5.73E-10 | | 0.02606 |
| 5-aminoimidazole ribonucleotide biosynthesis II | Nucleotide Metabolism & Biosynthesis | *Holdemanella* | *Holdemanella biformis* | CCR7^+^ Activated NK Cells | Natural Killer (NK) Cells | 6.02E-09 | 1.62E-09 | | 0.02611 |
| Superpathway of 5-aminoimidazole ribonucleotide biosynthesis | Nucleotide Metabolism & Biosynthesis | *Holdemanella* | *Holdemanella biformis* | CCR7^+^ Activated NK Cells | Natural Killer (NK) Cells | 6.02E-09 | 1.62E-09 | | 0.02611 |
| Superpathway of coenzyme A biosynthesis III (mammals) | Cofactor & Vitamin Biosynthesis | *Coprobacter* | *Coprobacter fastidiosus* | NK T Cells | Adaptive Immune Cells | 2.50E-11 | 6.46E-12 | | 0.02615 |
| Coenzyme A biosynthesis I (prokaryotic) | Cofactor & Vitamin Biosynthesis | *Coprobacter* | *Coprobacter fastidiosus* | IFNγ/TNFα-producing CD69^+^ CD8^+^ T Cells | CD8^+^ T Cells | 1.82E-10 | 4.66E-11 | | 0.02628 |
| Adenine and adenosine salvage III | Nucleotide Metabolism & Biosynthesis | *Coprobacter* | *Coprobacter fastidiosus* | IFNγ-producing CD4^+^ T Cells | CD4^+^ T Cells | 1.72E-11 | 4.59E-12 | | 0.02644 |
| L-histidine biosynthesis | Amino Acid Biosynthesis | *Coprobacter* | *Coprobacter fastidiosus* | IFNγ-producing Activated NK Cells | Natural Killer (NK) Cells | 1.56E-10 | 3.46E-11 | | 0.02645 |
| Glycogen biosynthesis I (from ADP-D-Glucose) | General Biosynthetic Pathways | *Eubacterium* | *Eubacterium ventriosum* | PD-1^+^ CD8^+^ T Cells | CD8^+^ T Cells | 7.48E-11 | 2.03E-11 | | 0.02664 |
| 5-aminoimidazole ribonucleotide biosynthesis I | Nucleotide Metabolism & Biosynthesis | *Coprobacter* | *Coprobacter fastidiosus* | IFNγ-producing NK Cells | Natural Killer (NK) Cells | 1.87E-10 | 4.11E-11 | | 0.02666 |
| L-valine biosynthesis | Amino Acid Biosynthesis | *Eubacterium* | *Eubacterium ventriosum* | TNFα-producing NK Cells | Natural Killer (NK) Cells | 1.95E-10 | 5.27E-11 | | 0.02670 |
| Queuosine biosynthesis I (de novo) | Nucleotide Metabolism & Biosynthesis | *Coprobacter* | *Coprobacter fastidiosus* | IFNγ-producing T regs | Adaptive Immune Cells | 5.69E-10 | 1.51E-10 | | 0.02677 |
| Peptidoglycan biosynthesis I (meso-diaminopimelate containing) | Cell wall & Peptidoglycan Synthesis | *Holdemanella* | *Holdemanella biformis* | IL-17A-producing CD4^+^ EM T Cells | CD4^+^ T Cells | 2.49E-09 | 6.17E-10 | | 0.02685 |
| 5-aminoimidazole ribonucleotide biosynthesis I | Nucleotide Metabolism & Biosynthesis | *Dialister* | *Dialister invisus* | IL-2-producing CD161^+^ CD8^+^ T Cells | CD8^+^ T Cells | 5.80E-09 | 1.36E-09 | | 0.02701 |
| Fatty acid biosynthesis initiation (mitochondria) | Lipid & Membrane Metabolism | *Escherichia* | *Escherichia coli* | IL-2-producing Activated CD69^+^ CD4^+^ T Cells | CD4^+^ T Cells | 3.92E-10 | 9.18E-11 | | 0.02735 |
| 5-aminoimidazole ribonucleotide biosynthesis I | Nucleotide Metabolism & Biosynthesis | *Eubacterium* | *Eubacterium ventriosum* | IL-17A-producing CD4^+^ T Cells | CD4^+^ T Cells | 1.38E-10 | 3.78E-11 | | 0.02802 |
| Coenzyme A biosynthesis II (eukaryotic) | Cofactor & Vitamin Biosynthesis | *Coprobacter* | *Coprobacter fastidiosus* | TNFα-producing Activated CD69^+^ CD8^+^ T Cells | CD8^+^ T Cells | 4.75E-11 | 1.22E-11 | | 0.02802 |
| UDP-N-acetylmuramoyl-pentapeptide biosynthesis I (meso-diaminopimelate containing) | Cell wall & Peptidoglycan Synthesis | *Holdemanella* | *Holdemanella biformis* | IL-17A-producing CD4^+^ EM T Cells | CD4^+^ T Cells | 2.43E-09 | 6.08E-10 | | 0.02803 |
| Adenine and adenosine salvage III | Nucleotide Metabolism & Biosynthesis | *Dialister* | *Dialister invisus* | IFNγ-producing T regs | Adaptive Immune Cells | 1.73E-09 | 4.66E-10 | | 0.02806 |
| Coenzyme A biosynthesis I (prokaryotic) | Cofactor & Vitamin Biosynthesis | *Dialister* | *Dialister invisus* | IFNγ-producing CD4^+^ T Cells | CD4^+^ T Cells | 4.36E-11 | 1.17E-11 | | 0.02823 |
| CDP diacylglycerol biosynthesis I | Lipid & Membrane Metabolism | *Coprobacter* | *Coprobacter fastidiosus* | TNFα-producing NK T Cells | Adaptive Immune Cells | 1.01E-10 | 2.65E-11 | | 0.02834 |
| CDP diacylglycerol biosynthesis II | Lipid & Membrane Metabolism | *Coprobacter* | *Coprobacter fastidiosus* | TNFα-producing NK T Cells | Adaptive Immune Cells | 1.01E-10 | 2.65E-11 | | 0.02834 |
| L-valine biosynthesis | Amino Acid Biosynthesis | *Eubacterium* | *Eubacterium ventriosum* | CD8^+^ CM | CD8^+^ T Cells | 7.01E-11 | 1.72E-11 | | 0.02838 |
| Flavin biosynthesis I (bacteria and plants) | Cofactor & Vitamin Biosynthesis | *Clostridium* | *Clostridium sp AM22 11AC* | IFNγ-producing CD4^+^ T Cells | CD4^+^ T Cells | 2.44E-11 | 6.54E-12 | | 0.02841 |
| 5-aminoimidazole ribonucleotide biosynthesis II | Nucleotide Metabolism & Biosynthesis | *Coprobacter* | *Coprobacter fastidiosus* | IFNγ-producing NK Cells | Natural Killer (NK) Cells | 1.72E-10 | 3.84E-11 | | 0.02867 |
| Superpathway of 5-aminoimidazole ribonucleotide biosynthesis | Nucleotide Metabolism & Biosynthesis | *Coprobacter* | *Coprobacter fastidiosus* | IFNγ-producing NK Cells | Natural Killer (NK) Cells | 1.72E-10 | 3.84E-11 | | 0.02867 |
| 5-aminoimidazole ribonucleotide biosynthesis II | Nucleotide Metabolism & Biosynthesis | *Eubacterium* | *Eubacterium ventriosum* | IL-17A-producing Activated CD69^+^ CD4^+^ T Cells | CD4^+^ T Cells | 1.39E-10 | 3.80E-11 | | 0.02891 |
| Superpathway of 5-aminoimidazole ribonucleotide biosynthesis | Nucleotide Metabolism & Biosynthesis | *Eubacterium* | *Eubacterium ventriosum* | IL-17A-producing Activated CD69^+^ CD4^+^ T Cells | CD4^+^ T Cells | 1.39E-10 | 3.80E-11 | | 0.02891 |
| Adenine and adenosine salvage III | Nucleotide Metabolism & Biosynthesis | *Holdemanella* | *Holdemanella biformis* | CCR7^+^ CD8^+^ T Cells | CD8^+^ T Cells | 5.53E-11 | 1.45E-11 | | 0.02893 |
| Glycogen biosynthesis I (from ADP-D-Glucose) | General Biosynthetic Pathways | *Eubacterium* | *Eubacterium ventriosum* | PD-1^+^ Activated HLA-DR^+^ CD4^+^ T Cells | CD4^+^ T Cells | 1.54E-10 | 4.24E-11 | | 0.02933 |
| Fatty acid biosynthesis initiation (mitochondria) | Lipid & Membrane Metabolism | *Clostridium* | *Clostridium sp AM22 11AC* | IFNγ-producing T regs | Adaptive Immune Cells | 1.33E-09 | 3.59E-10 | | 0.02957 |
| Superpathway of L-threonine biosynthesis | Amino Acid Biosynthesis | *Dialister* | *Dialister invisus* | IFNγ-producing T regs | Adaptive Immune Cells | 1.74E-09 | 4.72E-10 | | 0.03018 |
| 5-aminoimidazole ribonucleotide biosynthesis I | Nucleotide Metabolism & Biosynthesis | *Dialister* | *Dialister invisus* | IL-2/TNFα-producing CD4^+^ T Cells | CD4^+^ T Cells | 2.06E-10 | 4.82E-11 | | 0.03030 |
| Purine ribonucleosides degradation | Nucleotide Metabolism & Biosynthesis | *Eubacterium* | *Eubacterium ventriosum* | TNFα-producing NK Cells | Natural Killer (NK) Cells | 2.00E-10 | 5.49E-11 | | 0.03070 |
| Inosine 5-phosphate degradation | Nucleotide Metabolism & Biosynthesis | *Coprobacter* | *Coprobacter fastidiosus* | IFNγ/TNFα-producing CD161^+^ CD4^+^ T Cells | CD4^+^ T Cells | 7.62E-11 | 1.98E-11 | | 0.03113 |
| Folate transformations II (plants) | Cofactor & Vitamin Biosynthesis | *Coprobacter* | *Coprobacter fastidiosus* | IFNγ-producing CD4^+^ T Cells | CD4^+^ T Cells | 1.54E-11 | 4.19E-12 | | 0.03155 |
| 5-aminoimidazole ribonucleotide biosynthesis II | Nucleotide Metabolism & Biosynthesis | *Dialister* | *Dialister invisus* | IFNγ-producing CD4^+^ T Cells | CD4^+^ T Cells | 5.25E-11 | 1.43E-11 | | 0.03155 |
| Superpathway of 5-aminoimidazole ribonucleotide biosynthesis | Nucleotide Metabolism & Biosynthesis | *Dialister* | *Dialister invisus* | IFNγ-producing CD4^+^ T Cells | CD4^+^ T Cells | 5.25E-11 | 1.43E-11 | | 0.03155 |
| L-histidine biosynthesis | Amino Acid Biosynthesis | *Coprobacter* | *Coprobacter fastidiosus* | IFNγ-producing NK Cells | Natural Killer (NK) Cells | 1.53E-10 | 3.44E-11 | | 0.03163 |
| Chorismate biosynthesis I | General Biosynthetic Pathways | *Holdemanella* | *Holdemanella biformis* | Non-classical Monocytes | Innate Immune Cells | 5.11E-09 | 1.39E-09 | | 0.03173 |
| CDP diacylglycerol biosynthesis I | Lipid & Membrane Metabolism | *Coprobacter* | *Coprobacter fastidiosus* | IFNγ-producing CD161^+^ NK Cells | Natural Killer (NK) Cells | 3.01E-10 | 6.95E-11 | | 0.03177 |
| CDP diacylglycerol biosynthesis II | Lipid & Membrane Metabolism | *Coprobacter* | *Coprobacter fastidiosus* | IFNγ-producing CD161^+^ NK Cells | Natural Killer (NK) Cells | 3.01E-10 | 6.95E-11 | | 0.03177 |
| Folate transformations II (plants) | Cofactor & Vitamin Biosynthesis | *Coprobacter* | *Coprobacter fastidiosus* | IFNγ/TNFα-producing CD161^+^ CD4^+^ T Cells | CD4^+^ T Cells | 8.36E-11 | 2.18E-11 | | 0.03205 |
| Inosine 5-phosphate degradation | Nucleotide Metabolism & Biosynthesis | *Coprobacter* | *Coprobacter fastidiosus* | IFNγ/TNFα-producing CD161^+^ CD8^+^ T Cells | CD8^+^ T Cells | 3.55E-10 | 9.34E-11 | | 0.03256 |
| UMP biosynthesis I | Nucleotide Metabolism & Biosynthesis | *Eubacterium* | *Eubacterium ventriosum* | PD-1^+^ CD8^+^ T Cells | CD8^+^ T Cells | 7.19E-11 | 2.00E-11 | | 0.03286 |
| UMP biosynthesis II | Nucleotide Metabolism & Biosynthesis | *Eubacterium* | *Eubacterium ventriosum* | PD-1^+^ CD8^+^ T Cells | CD8^+^ T Cells | 7.19E-11 | 2.00E-11 | | 0.03286 |
| UMP biosynthesis III | Nucleotide Metabolism & Biosynthesis | *Eubacterium* | *Eubacterium ventriosum* | PD-1^+^ CD8^+^ T Cells | CD8^+^ T Cells | 7.19E-11 | 2.00E-11 | | 0.03286 |
| Coenzyme A biosynthesis II (eukaryotic) | Cofactor & Vitamin Biosynthesis | *Coprobacter* | *Coprobacter fastidiosus* | TNFα-producing CD161^+^ CD4^+^ T Cells | CD4^+^ T Cells | 6.95E-11 | 1.66E-11 | | 0.03296 |
| Coenzyme A biosynthesis I (prokaryotic) | Cofactor & Vitamin Biosynthesis | *Dialister* | *Dialister invisus* | TNFα-producing CD161^+^ CD4^+^ T Cells | CD4^+^ T Cells | 1.88E-10 | 4.52E-11 | | 0.03296 |
| Folate transformations II (plants) | Cofactor & Vitamin Biosynthesis | *Dialister* | *Dialister invisus* | TNFα-producing CD161^+^ CD4^+^ T Cells | CD4^+^ T Cells | 1.89E-10 | 4.54E-11 | | 0.03296 |
| UMP biosynthesis I | Nucleotide Metabolism & Biosynthesis | *Dialister* | *Dialister invisus* | TNFα-producing CD161^+^ CD4^+^ T Cells | CD4^+^ T Cells | 1.95E-10 | 4.31E-11 | | 0.03296 |
| 5-aminoimidazole ribonucleotide biosynthesis I | Nucleotide Metabolism & Biosynthesis | *Dialister* | *Dialister invisus* | TNFα-producing CD161^+^ CD4^+^ T Cells | CD4^+^ T Cells | 2.27E-10 | 5.26E-11 | | 0.03296 |
| 5-aminoimidazole ribonucleotide biosynthesis II | Nucleotide Metabolism & Biosynthesis | *Dialister* | *Dialister invisus* | TNFα-producing CD161^+^ CD4^+^ T Cells | CD4^+^ T Cells | 2.32E-10 | 5.48E-11 | | 0.03296 |
| Superpathway of 5-aminoimidazole ribonucleotide biosynthesis | Nucleotide Metabolism & Biosynthesis | *Dialister* | *Dialister invisus* | TNFα-producing CD161^+^ CD4^+^ T Cells | CD4^+^ T Cells | 2.32E-10 | 5.48E-11 | | 0.03296 |
| Guanosine ribonucleotides de novo biosynthesis | Nucleotide Metabolism & Biosynthesis | *Dialister* | *Dialister invisus* | TNFα-producing CD161^+^ CD4^+^ T Cells | CD4^+^ T Cells | 1.98E-10 | 4.80E-11 | | 0.03296 |
| UMP biosynthesis II | Nucleotide Metabolism & Biosynthesis | *Dialister* | *Dialister invisus* | TNFα-producing CD161^+^ CD4^+^ T Cells | CD4^+^ T Cells | 1.95E-10 | 4.31E-11 | | 0.03296 |
| UMP biosynthesis III | Nucleotide Metabolism & Biosynthesis | *Dialister* | *Dialister invisus* | TNFα-producing CD161^+^ CD4^+^ T Cells | CD4^+^ T Cells | 1.95E-10 | 4.31E-11 | | 0.03296 |
| L-valine biosynthesis | Amino Acid Biosynthesis | *Eubacterium* | *Eubacterium ventriosum* | TNFα-producing T regs | Adaptive Immune Cells | 8.43E-11 | 2.35E-11 | | 0.03317 |
| UMP biosynthesis I | Nucleotide Metabolism & Biosynthesis | *Coprobacter* | *Coprobacter fastidiosus* | IFNγ-producing CD161^+^ NK Cells | Natural Killer (NK) Cells | 3.02E-10 | 7.30E-11 | | 0.03336 |
| UMP biosynthesis II | Nucleotide Metabolism & Biosynthesis | *Coprobacter* | *Coprobacter fastidiosus* | IFNγ-producing CD161^+^ NK Cells | Natural Killer (NK) Cells | 3.02E-10 | 7.30E-11 | | 0.03336 |
| UMP biosynthesis III | Nucleotide Metabolism & Biosynthesis | *Coprobacter* | *Coprobacter fastidiosus* | IFNγ-producing CD161^+^ NK Cells | Natural Killer (NK) Cells | 3.02E-10 | 7.30E-11 | | 0.03336 |
| Folate transformations II (plants) | Cofactor & Vitamin Biosynthesis | *Eubacterium* | *Eubacterium ventriosum* | TNFα-producing T regs | Adaptive Immune Cells | 7.14E-11 | 2.00E-11 | | 0.03340 |
| 5-aminoimidazole ribonucleotide biosynthesis I | Nucleotide Metabolism & Biosynthesis | *Eubacterium* | *Eubacterium ventriosum* | IL-17A-producing CD161^+^ CD4^+^ T Cells | CD4^+^ T Cells | 4.11E-10 | 1.07E-10 | | 0.03353 |
| UMP biosynthesis I | Nucleotide Metabolism & Biosynthesis | *Holdemanella* | *Holdemanella biformis* | IL-17A-producing Activated NK Cells | Natural Killer (NK) Cells | 1.63E-08 | 4.37E-09 | | 0.03371 |
| UMP biosynthesis II | Nucleotide Metabolism & Biosynthesis | *Holdemanella* | *Holdemanella biformis* | IL-17A-producing Activated NK Cells | Natural Killer (NK) Cells | 1.63E-08 | 4.37E-09 | | 0.03371 |
| UMP biosynthesis III | Nucleotide Metabolism & Biosynthesis | *Holdemanella* | *Holdemanella biformis* | IL-17A-producing Activated NK Cells | Natural Killer (NK) Cells | 1.63E-08 | 4.37E-09 | | 0.03371 |
| Glycogen biosynthesis I (from ADP-D-Glucose) | General Biosynthetic Pathways | *Escherichia* | *Escherichia coli* | γδ T Cells | Adaptive Immune Cells | 2.58E-11 | 6.68E-12 | | 0.03389 |
| 5-aminoimidazole ribonucleotide biosynthesis II | Nucleotide Metabolism & Biosynthesis | *Dialister* | *Dialister invisus* | IL-2/TNFα-producing CD4^+^ T Cells | CD4^+^ T Cells | 2.10E-10 | 5.04E-11 | | 0.03425 |
| Superpathway of 5-aminoimidazole ribonucleotide biosynthesis | Nucleotide Metabolism & Biosynthesis | *Dialister* | *Dialister invisus* | IL-2/TNFα-producing CD4^+^ T Cells | CD4^+^ T Cells | 2.10E-10 | 5.04E-11 | | 0.03425 |
| Coenzyme A biosynthesis I (prokaryotic) | Cofactor & Vitamin Biosynthesis | *Dialister* | *Dialister invisus* | IL-2-producing CD161^+^ CD8^+^ T Cells | CD8^+^ T Cells | 4.83E-09 | 1.17E-09 | | 0.03460 |
| 5-aminoimidazole ribonucleotide biosynthesis II | Nucleotide Metabolism & Biosynthesis | *Dialister* | *Dialister invisus* | IL-2-producing CD161^+^ CD8^+^ T Cells | CD8^+^ T Cells | 5.88E-09 | 1.42E-09 | | 0.03460 |
| Superpathway of 5-aminoimidazole ribonucleotide biosynthesis | Nucleotide Metabolism & Biosynthesis | *Dialister* | *Dialister invisus* | IL-2-producing CD161^+^ CD8^+^ T Cells | CD8^+^ T Cells | 5.88E-09 | 1.42E-09 | | 0.03460 |
| Folate transformations II (plants) | Cofactor & Vitamin Biosynthesis | *Coprobacter* | *Coprobacter fastidiosus* | IFNγ/TNFα-producing CD161^+^ CD8^+^ T Cells | CD8^+^ T Cells | 3.88E-10 | 1.03E-10 | | 0.03468 |
| Chorismate biosynthesis from 3-dehydroquinate | General Biosynthetic Pathways | *Coprobacter* | *Coprobacter fastidiosus* | IFNγ-producing CD161^+^ NK Cells | Natural Killer (NK) Cells | 3.37E-10 | 8.30E-11 | | 0.03497 |
| Guanosine ribonucleotides de novo biosynthesis | Nucleotide Metabolism & Biosynthesis | *Coprobacter* | *Coprobacter fastidiosus* | IFNγ-producing CD161^+^ NK Cells | Natural Killer (NK) Cells | 1.74E-10 | 4.28E-11 | | 0.03497 |
| 5-aminoimidazole ribonucleotide biosynthesis I | Nucleotide Metabolism & Biosynthesis | *Clostridium* | *Clostridium sp AM22 11AC* | CCR7^+^ CD8^+^ T Cells | CD8^+^ T Cells | 3.17E-11 | 8.51E-12 | | 0.03521 |
| Superpathway of L-threonine biosynthesis | Amino Acid Biosynthesis | *Escherichia* | *Escherichia coli* | IL-2-producing Activated CD69^+^ CD4^+^ T Cells | CD4^+^ T Cells | 3.44E-10 | 8.29E-11 | | 0.03523 |
| Superpathway of coenzyme A biosynthesis III (mammals) | Cofactor & Vitamin Biosynthesis | *Dialister* | *Dialister invisus* | TNFα-producing CD161^+^ CD4^+^ T Cells | CD4^+^ T Cells | 1.78E-10 | 4.39E-11 | | 0.03528 |
| Coenzyme A biosynthesis II (eukaryotic) | Cofactor & Vitamin Biosynthesis | *Dialister* | *Dialister invisus* | TNFα-producing CD161^+^ CD4^+^ T Cells | CD4^+^ T Cells | 1.73E-10 | 4.26E-11 | | 0.03528 |
| Coenzyme A biosynthesis I (prokaryotic) | Cofactor & Vitamin Biosynthesis | *Eubacterium* | *Eubacterium ventriosum* | CD8^+^ CM | CD8^+^ T Cells | 6.30E-11 | 1.59E-11 | | 0.03543 |
| UMP biosynthesis I | Nucleotide Metabolism & Biosynthesis | *Dialister* | *Dialister invisus* | IL-2/TNFα-producing CD69^+^ CD4^+^ T Cells | CD4^+^ T Cells | 1.17E-10 | 2.76E-11 | | 0.03560 |
| UMP biosynthesis II | Nucleotide Metabolism & Biosynthesis | *Dialister* | *Dialister invisus* | IL-2/TNFα-producing CD69^+^ CD4^+^ T Cells | CD4^+^ T Cells | 1.17E-10 | 2.76E-11 | | 0.03560 |
| UMP biosynthesis III | Nucleotide Metabolism & Biosynthesis | *Dialister* | *Dialister invisus* | IL-2/TNFα-producing CD69^+^ CD4^+^ T Cells | CD4^+^ T Cells | 1.17E-10 | 2.76E-11 | | 0.03560 |
| Chorismate biosynthesis from 3-dehydroquinate | General Biosynthetic Pathways | *Holdemanella* | *Holdemanella biformis* | Non-classical Monocytes | Innate Immune Cells | 4.64E-09 | 1.28E-09 | | 0.03562 |
| Folate transformations II (plants) | Cofactor & Vitamin Biosynthesis | *Dialister* | *Dialister invisus* | IFNγ-producing CD4^+^ T Cells | CD4^+^ T Cells | 4.27E-11 | 1.18E-11 | | 0.03583 |
| Inosine 5-phosphate degradation | Nucleotide Metabolism & Biosynthesis | *Escherichia* | *Escherichia coli* | IL-2-producing Activated CD69^+^ CD4^+^ T Cells | CD4^+^ T Cells | 4.21E-10 | 1.03E-10 | | 0.03586 |
| Purine ribonucleosides degradation | Nucleotide Metabolism & Biosynthesis | *Clostridium* | *Clostridium sp AM22 11AC* | IFNγ-producing T regs | Adaptive Immune Cells | 1.14E-09 | 3.18E-10 | | 0.03589 |
| Chorismate biosynthesis from 3-dehydroquinate | General Biosynthetic Pathways | *Clostridium* | *Clostridium sp AM22 11AC* | IFNγ-producing T regs | Adaptive Immune Cells | 1.07E-09 | 2.99E-10 | | 0.03589 |
| 5-aminoimidazole ribonucleotide biosynthesis II | Nucleotide Metabolism & Biosynthesis | *Eubacterium* | *Eubacterium ventriosum* | IL-17A-producing CD4^+^ T Cells | CD4^+^ T Cells | 1.29E-10 | 3.62E-11 | | 0.03599 |
| Superpathway of 5-aminoimidazole ribonucleotide biosynthesis | Nucleotide Metabolism & Biosynthesis | *Eubacterium* | *Eubacterium ventriosum* | IL-17A-producing CD4^+^ T Cells | CD4^+^ T Cells | 1.29E-10 | 3.62E-11 | | 0.03599 |
| Folate transformations II (plants) | Cofactor & Vitamin Biosynthesis | *Clostridium* | *Clostridium sp AM22 11AC* | CCR7^+^ CD8^+^ T Cells | CD8^+^ T Cells | 4.32E-11 | 1.16E-11 | | 0.03673 |
| Coenzyme A biosynthesis I (prokaryotic) | Cofactor & Vitamin Biosynthesis | *Dialister* | *Dialister invisus* | IL-2/TNFα-producing CD4^+^ T Cells | CD4^+^ T Cells | 1.71E-10 | 4.15E-11 | | 0.03673 |
| Folate transformations II (plants) | Cofactor & Vitamin Biosynthesis | *Dialister* | *Dialister invisus* | IL-2/TNFα-producing CD4^+^ T Cells | CD4^+^ T Cells | 1.70E-10 | 4.17E-11 | | 0.03673 |
| 5-aminoimidazole ribonucleotide biosynthesis II | Nucleotide Metabolism & Biosynthesis | *Holdemanella* | *Holdemanella biformis* | Non-classical Monocytes | Innate Immune Cells | 7.05E-09 | 1.95E-09 | | 0.03721 |
| Superpathway of 5-aminoimidazole ribonucleotide biosynthesis | Nucleotide Metabolism & Biosynthesis | *Holdemanella* | *Holdemanella biformis* | Non-classical Monocytes | Innate Immune Cells | 7.05E-09 | 1.95E-09 | | 0.03721 |
| Coenzyme A biosynthesis I (prokaryotic) | Cofactor & Vitamin Biosynthesis | *Coprobacter* | *Coprobacter fastidiosus* | IFNγ/TNFα-producing CD69^+^ CD4^+^ T Cells | CD4^+^ T Cells | 3.94E-11 | 1.04E-11 | | 0.03723 |
| Folate transformations II (plants) | Cofactor & Vitamin Biosynthesis | *Eubacterium* | *Eubacterium ventriosum* | PMDSCs | Innate Immune Cells | 8.08E-11 | 2.29E-11 | | 0.03725 |
| Superpathway of coenzyme A biosynthesis III (mammals) | Cofactor & Vitamin Biosynthesis | *Dialister* | *Dialister invisus* | IFNγ-producing CD4^+^ T Cells | CD4^+^ T Cells | 4.09E-11 | 1.14E-11 | | 0.03745 |
| Inosine 5-phosphate degradation | Nucleotide Metabolism & Biosynthesis | *Dialister* | *Dialister invisus* | IFNγ-producing CD4^+^ T Cells | CD4^+^ T Cells | 3.83E-11 | 1.07E-11 | | 0.03745 |
| UMP biosynthesis I | Nucleotide Metabolism & Biosynthesis | *Eubacterium* | *Eubacterium ventriosum* | TNFα-producing CD161^+^ NK Cells | Natural Killer (NK) Cells | 1.48E-10 | 4.15E-11 | | 0.03748 |
| UMP biosynthesis II | Nucleotide Metabolism & Biosynthesis | *Eubacterium* | *Eubacterium ventriosum* | TNFα-producing CD161^+^ NK Cells | Natural Killer (NK) Cells | 1.48E-10 | 4.15E-11 | | 0.03748 |
| UMP biosynthesis III | Nucleotide Metabolism & Biosynthesis | *Eubacterium* | *Eubacterium ventriosum* | TNFα-producing CD161^+^ NK Cells | Natural Killer (NK) Cells | 1.48E-10 | 4.15E-11 | | 0.03748 |
| Purine ribonucleosides degradation | Nucleotide Metabolism & Biosynthesis | *Eubacterium* | *Eubacterium ventriosum* | CD8^+^ CM | CD8^+^ T Cells | 7.12E-11 | 1.80E-11 | | 0.03750 |
| Coenzyme A biosynthesis II (eukaryotic) | Cofactor & Vitamin Biosynthesis | *Coprobacter* | *Coprobacter fastidiosus* | TNFα-producing CD8^+^ T Cells | CD8^+^ T Cells | 5.82E-11 | 1.54E-11 | | 0.03757 |
| Folate transformations II (plants) | Cofactor & Vitamin Biosynthesis | *Eubacterium* | *Eubacterium ventriosum* | PD-1^+^ CD4^+^ T Cells | CD4^+^ T Cells | 3.33E-11 | 9.43E-12 | | 0.03764 |
| Guanosine ribonucleotides de novo biosynthesis | Nucleotide Metabolism & Biosynthesis | *Dialister* | *Dialister invisus* | IL-2/TNFα-producing CD4^+^ T Cells | CD4^+^ T Cells | 1.78E-10 | 4.42E-11 | | 0.03805 |
| Inosine 5-phosphate degradation | Nucleotide Metabolism & Biosynthesis | *Coprobacter* | *Coprobacter fastidiosus* | IFNγ-producing Activated CD69^+^ CD4^+^ T Cells | CD4^+^ T Cells | 1.97E-11 | 5.24E-12 | | 0.03816 |
| Folate transformations II (plants) | Cofactor & Vitamin Biosynthesis | *Eubacterium* | *Eubacterium ventriosum* | TNFα-producing CD161^+^ NK Cells | Natural Killer (NK) Cells | 1.53E-10 | 4.31E-11 | | 0.03816 |
| Adenine and adenosine salvage III | Nucleotide Metabolism & Biosynthesis | *Coprobacter* | *Coprobacter fastidiosus* | IFNγ/TNFα-producing CD69^+^ CD8^+^ T Cells | CD8^+^ T Cells | 1.76E-10 | 4.69E-11 | | 0.03817 |
| Adenine and adenosine salvage III | Nucleotide Metabolism & Biosynthesis | *Escherichia* | *Escherichia coli* | IL-2-producing CD161^+^ CD4^+^ T Cells | CD4^+^ T Cells | 1.90E-10 | 4.74E-11 | | 0.03825 |
| L-valine biosynthesis | Amino Acid Biosynthesis | *Eubacterium* | *Eubacterium ventriosum* | PD-1^+^ CD4^+^ T Cells | CD4^+^ T Cells | 3.93E-11 | 1.11E-11 | | 0.03827 |
| L-valine biosynthesis | Amino Acid Biosynthesis | *Eubacterium* | *Eubacterium ventriosum* | PD-1^+^ CD8^+^ T Cells | CD8^+^ T Cells | 8.68E-11 | 2.45E-11 | | 0.03859 |
| Peptidoglycan biosynthesis I (meso-diaminopimelate containing) | Cell wall & Peptidoglycan Synthesis | *Dialister* | *Dialister invisus* | IFNγ-producing T regs | Adaptive Immune Cells | 1.85E-09 | 5.20E-10 | | 0.03873 |
| Coenzyme A biosynthesis II (eukaryotic) | Cofactor & Vitamin Biosynthesis | *Dialister* | *Dialister invisus* | IFNγ-producing CD4^+^ T Cells | CD4^+^ T Cells | 3.95E-11 | 1.11E-11 | | 0.03903 |
| Coenzyme A biosynthesis II (eukaryotic) | Cofactor & Vitamin Biosynthesis | *Clostridium* | *Clostridium sp AM22 11AC* | IFNγ-producing T regs | Adaptive Immune Cells | 8.62E-10 | 2.43E-10 | | 0.03907 |
| UDP-N-acetylmuramoyl-pentapeptide biosynthesis II (lysine-containing) | Cell wall & Peptidoglycan Synthesis | *Dialister* | *Dialister invisus* | IFNγ-producing T regs | Adaptive Immune Cells | 1.90E-09 | 5.34E-10 | | 0.03917 |
| 5-aminoimidazole ribonucleotide biosynthesis I | Nucleotide Metabolism & Biosynthesis | *Escherichia* | *Escherichia coli* | IL-2-producing Activated CD69^+^ CD4^+^ T Cells | CD4^+^ T Cells | 2.99E-10 | 7.37E-11 | | 0.03918 |
| Glycogen biosynthesis I (from ADP-D-Glucose) | General Biosynthetic Pathways | *Eubacterium* | *Eubacterium ventriosum* | TNFα-producing gd T Cells | Adaptive Immune Cells | 3.04E-11 | 8.64E-12 | | 0.03918 |
| Folate transformations II (plants) | Cofactor & Vitamin Biosynthesis | *Dialister* | *Dialister invisus* | IL-2-producing CD161^+^ CD8^+^ T Cells | CD8^+^ T Cells | 4.77E-09 | 1.18E-09 | | 0.03918 |
| tRNA charging | Amino Acid Biosynthesis | *Holdemanella* | *Holdemanella biformis* | Non-classical Monocytes | Innate Immune Cells | 4.21E-09 | 1.17E-09 | | 0.03924 |
| Superpathway of coenzyme A biosynthesis III (mammals) | Cofactor & Vitamin Biosynthesis | *Dialister* | *Dialister invisus* | IL-2-producing CD161^+^ CD8^+^ T Cells | CD8^+^ T Cells | 4.54E-09 | 1.14E-09 | | 0.03930 |
| Guanosine ribonucleotides de novo biosynthesis | Nucleotide Metabolism & Biosynthesis | *Dialister* | *Dialister invisus* | IL-2-producing CD161^+^ CD8^+^ T Cells | CD8^+^ T Cells | 4.99E-09 | 1.25E-09 | | 0.03930 |
| L-lysine biosynthesis VI | Amino Acid Biosynthesis | *Coprobacter* | *Coprobacter fastidiosus* | IFNγ-producing Activated NK Cells | Natural Killer (NK) Cells | 1.53E-10 | 3.51E-11 | | 0.03934 |
| Coenzyme A biosynthesis I (prokaryotic) | Cofactor & Vitamin Biosynthesis | *Coprobacter* | *Coprobacter fastidiosus* | IFNγ/TNFα-producing CD8^+^ T Cells | CD8^+^ T Cells | 6.79E-11 | 1.80E-11 | | 0.03940 |
| Guanosine ribonucleotides de novo biosynthesis | Nucleotide Metabolism & Biosynthesis | *Coprobacter* | *Coprobacter fastidiosus* | IFNγ-producing Activated NK Cells | Natural Killer (NK) Cells | 8.80E-11 | 2.02E-11 | | 0.03955 |
| Coenzyme A biosynthesis II (eukaryotic) | Cofactor & Vitamin Biosynthesis | *Dialister* | *Dialister invisus* | IL-2-producing CD161^+^ CD8^+^ T Cells | CD8^+^ T Cells | 4.39E-09 | 1.10E-09 | | 0.03959 |
| Folate transformations II (plants) | Cofactor & Vitamin Biosynthesis | *Eubacterium* | *Eubacterium ventriosum* | PD-1^+^ CD8^+^ T Cells | CD8^+^ T Cells | 7.32E-11 | 2.08E-11 | | 0.03988 |
| Guanosine ribonucleotides de novo biosynthesis | Nucleotide Metabolism & Biosynthesis | *Dialister* | *Dialister invisus* | IFNγ-producing CD4^+^ T Cells | CD4^+^ T Cells | 4.45E-11 | 1.25E-11 | | 0.03993 |
| CDP diacylglycerol biosynthesis I | Lipid & Membrane Metabolism | *Dialister* | *Dialister invisus* | IFNγ-producing T regs | Adaptive Immune Cells | 2.08E-09 | 5.86E-10 | | 0.03994 |
| CDP diacylglycerol biosynthesis II | Lipid & Membrane Metabolism | *Dialister* | *Dialister invisus* | IFNγ-producing T regs | Adaptive Immune Cells | 2.08E-09 | 5.86E-10 | | 0.03994 |
| Inosine 5-phosphate degradation | Nucleotide Metabolism & Biosynthesis | *Coprobacter* | *Coprobacter fastidiosus* | IFNγ-producing CD4^+^ T Cells | CD4^+^ T Cells | 1.36E-11 | 3.84E-12 | | 0.04012 |
| 5-aminoimidazole ribonucleotide biosynthesis II | Nucleotide Metabolism & Biosynthesis | *Clostridium* | *Clostridium sp AM22 11AC* | CCR7^+^ CD8^+^ T Cells | CD8^+^ T Cells | 3.34E-11 | 9.14E-12 | | 0.04013 |
| Superpathway of 5-aminoimidazole ribonucleotide biosynthesis | Nucleotide Metabolism & Biosynthesis | *Clostridium* | *Clostridium sp AM22 11AC* | CCR7^+^ CD8^+^ T Cells | CD8^+^ T Cells | 3.34E-11 | 9.14E-12 | | 0.04013 |
| Inosine 5-phosphate degradation | Nucleotide Metabolism & Biosynthesis | *Eubacterium* | *Eubacterium ventriosum* | CCR7^+^ CD8^+^ T Cells | CD8^+^ T Cells | 1.71E-11 | 4.68E-12 | | 0.04013 |
| Inosine 5-phosphate degradation | Nucleotide Metabolism & Biosynthesis | *Dialister* | *Dialister invisus* | IL-2-producing CD161^+^ CD8^+^ T Cells | CD8^+^ T Cells | 4.23E-09 | 1.07E-09 | | 0.04041 |
| Flavin biosynthesis I (bacteria and plants) | Cofactor & Vitamin Biosynthesis | *Holdemanella* | *Holdemanella biformis* | CCR7^+^ NK Cells | Natural Killer (NK) Cells | 6.09E-09 | 1.60E-09 | | 0.04079 |
| Folate transformations II (plants) | Cofactor & Vitamin Biosynthesis | *Eubacterium* | *Eubacterium ventriosum* | CD8^+^ CM | CD8^+^ T Cells | 5.75E-11 | 1.48E-11 | | 0.04162 |
| Phosphopantothenate biosynthesis I | Lipid & Membrane Metabolism | *Eubacterium* | *Eubacterium ventriosum* | CCR7^+^ CD8^+^ T Cells | CD8^+^ T Cells | 1.67E-11 | 4.58E-12 | | 0.04180 |
| Folate transformations II (plants) | Cofactor & Vitamin Biosynthesis | *Coprobacter* | *Coprobacter fastidiosus* | IFNγ-producing Activated CD69^+^ CD4^+^ T Cells | CD4^+^ T Cells | 2.15E-11 | 5.78E-12 | | 0.04186 |
| Fatty acid biosynthesis initiation (mitochondria) | Lipid & Membrane Metabolism | *Escherichia* | *Escherichia coli* | IL-2-producing gd T Cells | Adaptive Immune Cells | 3.69E-09 | 8.69E-10 | | 0.04204 |
| L-valine biosynthesis | Amino Acid Biosynthesis | *Eubacterium* | *Eubacterium ventriosum* | PMDSCs | Innate Immune Cells | 9.43E-11 | 2.71E-11 | | 0.04227 |
| Superpathway of coenzyme A biosynthesis III (mammals) | Cofactor & Vitamin Biosynthesis | *Dialister* | *Dialister invisus* | IL-2/TNFα-producing CD4^+^ T Cells | CD4^+^ T Cells | 1.60E-10 | 4.04E-11 | | 0.04230 |
| Coenzyme A biosynthesis II (eukaryotic) | Cofactor & Vitamin Biosynthesis | *Dialister* | *Dialister invisus* | IL-2/TNFα-producing CD4^+^ T Cells | CD4^+^ T Cells | 1.56E-10 | 3.92E-11 | | 0.04230 |
| UMP biosynthesis I | Nucleotide Metabolism & Biosynthesis | *Eubacterium* | *Eubacterium ventriosum* | TNFα-producing T regs | Adaptive Immune Cells | 6.74E-11 | 1.94E-11 | | 0.04252 |
| UMP biosynthesis II | Nucleotide Metabolism & Biosynthesis | *Eubacterium* | *Eubacterium ventriosum* | TNFα-producing T regs | Adaptive Immune Cells | 6.74E-11 | 1.94E-11 | | 0.04252 |
| UMP biosynthesis III | Nucleotide Metabolism & Biosynthesis | *Eubacterium* | *Eubacterium ventriosum* | TNFα-producing T regs | Adaptive Immune Cells | 6.74E-11 | 1.94E-11 | | 0.04252 |
| UDP-N-acetylmuramoyl-pentapeptide biosynthesis I (meso-diaminopimelate containing) | Cell wall & Peptidoglycan Synthesis | *Dialister* | *Dialister invisus* | IFNγ-producing T regs | Adaptive Immune Cells | 1.90E-09 | 5.40E-10 | | 0.04255 |
| Phosphopantothenate biosynthesis I | Lipid & Membrane Metabolism | *Coprobacter* | *Coprobacter fastidiosus* | IFNγ-producing CD161^+^ NK Cells | Natural Killer (NK) Cells | 2.21E-10 | 5.64E-11 | | 0.04257 |
| UDP-N-acetylmuramoyl-pentapeptide biosynthesis II (lysine-containing) | Cell wall & Peptidoglycan Synthesis | *Coprobacter* | *Coprobacter fastidiosus* | IFNγ-producing CD161^+^ NK Cells | Natural Killer (NK) Cells | 2.50E-10 | 6.38E-11 | | 0.04257 |
| Fatty acid biosynthesis initiation (mitochondria) | Lipid & Membrane Metabolism | *Coprobacter* | *Coprobacter fastidiosus* | IFNγ-producing CD161^+^ NK Cells | Natural Killer (NK) Cells | 2.82E-10 | 7.22E-11 | | 0.04257 |
| Inosine 5-phosphate degradation | Nucleotide Metabolism & Biosynthesis | *Dialister* | *Dialister invisus* | TNFα-producing CD161^+^ CD4^+^ T Cells | CD4^+^ T Cells | 1.64E-10 | 4.15E-11 | | 0.04263 |
| Coenzyme A biosynthesis I (prokaryotic) | Cofactor & Vitamin Biosynthesis | *Dialister* | *Dialister invisus* | IL-2/TNFα-producing CD4^+^ EM T Cells | CD4^+^ T Cells | 5.57E-10 | 1.44E-10 | | 0.04263 |
| Folate transformations II (plants) | Cofactor & Vitamin Biosynthesis | *Dialister* | *Dialister invisus* | IL-2/TNFα-producing CD4^+^ EM T Cells | CD4^+^ T Cells | 5.58E-10 | 1.45E-10 | | 0.04263 |
| UMP biosynthesis I | Nucleotide Metabolism & Biosynthesis | *Dialister* | *Dialister invisus* | IL-2/TNFα-producing CD4^+^ EM T Cells | CD4^+^ T Cells | 5.77E-10 | 1.38E-10 | | 0.04263 |
| 5-aminoimidazole ribonucleotide biosynthesis I | Nucleotide Metabolism & Biosynthesis | *Dialister* | *Dialister invisus* | IL-2/TNFα-producing CD4^+^ EM T Cells | CD4^+^ T Cells | 6.72E-10 | 1.68E-10 | | 0.04263 |
| 5-aminoimidazole ribonucleotide biosynthesis II | Nucleotide Metabolism & Biosynthesis | *Dialister* | *Dialister invisus* | IL-2/TNFα-producing CD4^+^ EM T Cells | CD4^+^ T Cells | 6.89E-10 | 1.74E-10 | | 0.04263 |
| Superpathway of 5-aminoimidazole ribonucleotide biosynthesis | Nucleotide Metabolism & Biosynthesis | *Dialister* | *Dialister invisus* | IL-2/TNFα-producing CD4^+^ EM T Cells | CD4^+^ T Cells | 6.89E-10 | 1.74E-10 | | 0.04263 |
| UMP biosynthesis II | Nucleotide Metabolism & Biosynthesis | *Dialister* | *Dialister invisus* | IL-2/TNFα-producing CD4^+^ EM T Cells | CD4^+^ T Cells | 5.77E-10 | 1.38E-10 | | 0.04263 |
| UMP biosynthesis III | Nucleotide Metabolism & Biosynthesis | *Dialister* | *Dialister invisus* | IL-2/TNFα-producing CD4^+^ EM T Cells | CD4^+^ T Cells | 5.77E-10 | 1.38E-10 | | 0.04263 |
| Glycogen biosynthesis I (from ADP-D-Glucose) | General Biosynthetic Pathways | *Escherichia* | *Escherichia coli* | IL-2/TNFα-producing CD4^+^ EM T Cells | CD4^+^ T Cells | 1.27E-10 | 3.13E-11 | | 0.04263 |
| Queuosine biosynthesis I (de novo) | Nucleotide Metabolism & Biosynthesis | *Escherichia* | *Escherichia coli* | IL-2/TNFα-producing CD4^+^ EM T Cells | CD4^+^ T Cells | 1.34E-10 | 3.42E-11 | | 0.04263 |
| Fatty acid biosynthesis initiation (mitochondria) | Lipid & Membrane Metabolism | *Escherichia* | *Escherichia coli* | IL-2/TNFα-producing CD4^+^ EM T Cells | CD4^+^ T Cells | 1.68E-10 | 4.22E-11 | | 0.04263 |
| Superpathway of L-threonine biosynthesis | Amino Acid Biosynthesis | *Escherichia* | *Escherichia coli* | IL-2/TNFα-producing CD4^+^ EM T Cells | CD4^+^ T Cells | 1.50E-10 | 3.79E-11 | | 0.04263 |
| 5-aminoimidazole ribonucleotide biosynthesis I | Nucleotide Metabolism & Biosynthesis | *Eubacterium* | *Eubacterium ventriosum* | IFNγ-producing T regs | Adaptive Immune Cells | 5.74E-10 | 1.64E-10 | | 0.04290 |
| 5-aminoimidazole ribonucleotide biosynthesis I | Nucleotide Metabolism & Biosynthesis | *Escherichia* | *Escherichia coli* | IL-2-producing gd T Cells | Adaptive Immune Cells | 2.91E-09 | 6.88E-10 | | 0.04293 |
| L-valine biosynthesis | Amino Acid Biosynthesis | *Eubacterium* | *Eubacterium ventriosum* | PD-1^+^ Activated HLA-DR^+^ CD4^+^ T Cells | CD4^+^ T Cells | 1.79E-10 | 5.11E-11 | | 0.04300 |
| Guanosine ribonucleotides de novo biosynthesis | Nucleotide Metabolism & Biosynthesis | *Dialister* | *Dialister invisus* | IL-2/TNFα-producing CD4^+^ EM T Cells | CD4^+^ T Cells | 5.85E-10 | 1.53E-10 | | 0.04319 |
| Folate transformations II (plants) | Cofactor & Vitamin Biosynthesis | *Eubacterium* | *Eubacterium ventriosum* | PD-1^+^ Activated HLA-DR^+^ CD4^+^ T Cells | CD4^+^ T Cells | 1.52E-10 | 4.33E-11 | | 0.04355 |
| UMP biosynthesis I | Nucleotide Metabolism & Biosynthesis | *Eubacterium* | *Eubacterium ventriosum* | PD-1^+^ CD4^+^ T Cells | CD4^+^ T Cells | 3.17E-11 | 9.13E-12 | | 0.04366 |
| UMP biosynthesis II | Nucleotide Metabolism & Biosynthesis | *Eubacterium* | *Eubacterium ventriosum* | PD-1^+^ CD4^+^ T Cells | CD4^+^ T Cells | 3.17E-11 | 9.13E-12 | | 0.04366 |
| UMP biosynthesis III | Nucleotide Metabolism & Biosynthesis | *Eubacterium* | *Eubacterium ventriosum* | PD-1^+^ CD4^+^ T Cells | CD4^+^ T Cells | 3.17E-11 | 9.13E-12 | | 0.04366 |
| Coenzyme A biosynthesis I (prokaryotic) | Cofactor & Vitamin Biosynthesis | *Coprobacter* | *Coprobacter fastidiosus* | IFNγ-producing CD161^+^ CD8^+^ T Cells | CD8^+^ T Cells | 1.07E-10 | 2.89E-11 | | 0.04368 |
| 5-aminoimidazole ribonucleotide biosynthesis I | Nucleotide Metabolism & Biosynthesis | *Escherichia* | *Escherichia coli* | IL-2-producing CD161^+^ CD4^+^ T Cells | CD4^+^ T Cells | 1.86E-10 | 4.72E-11 | | 0.04405 |
| tRNA charging | Amino Acid Biosynthesis | *Coprobacter* | *Coprobacter fastidiosus* | IFNγ-producing CD161^+^ NK Cells | Natural Killer (NK) Cells | 2.89E-10 | 7.45E-11 | | 0.04426 |
| Chorismate biosynthesis from 3-dehydroquinate | General Biosynthetic Pathways | *Clostridium* | *Clostridium sp AM22 11AC* | CD8^+^ Naive | CD8^+^ T Cells | 6.09E-11 | 1.40E-11 | | 0.04430 |
| Chorismate biosynthesis I | General Biosynthetic Pathways | *Clostridium* | *Clostridium sp AM22 11AC* | CD8^+^ Naive | CD8^+^ T Cells | 5.60E-11 | 1.33E-11 | | 0.04430 |
| L-histidine biosynthesis | Amino Acid Biosynthesis | *Clostridium* | *Clostridium sp AM22 11AC* | CD8^+^ Naive | CD8^+^ T Cells | 4.72E-11 | 1.02E-11 | | 0.04430 |
| UMP biosynthesis I | Nucleotide Metabolism & Biosynthesis | *Clostridium* | *Clostridium sp AM22 11AC* | CD8^+^ Naive | CD8^+^ T Cells | 7.08E-11 | 1.55E-11 | | 0.04430 |
| UMP biosynthesis II | Nucleotide Metabolism & Biosynthesis | *Clostridium* | *Clostridium sp AM22 11AC* | CD8^+^ Naive | CD8^+^ T Cells | 7.08E-11 | 1.55E-11 | | 0.04430 |
| UMP biosynthesis II | Nucleotide Metabolism & Biosynthesis | *Clostridium* | *Clostridium sp AM22 11AC* | CD8^+^ Naive | CD8^+^ T Cells | 7.08E-11 | 1.55E-11 | | 0.04430 |
| Guanosine ribonucleotides de novo biosynthesis | Nucleotide Metabolism & Biosynthesis | *Eubacterium* | *Eubacterium ventriosum* | CD8^+^ Naive | CD8^+^ T Cells | 2.70E-11 | 6.80E-12 | | 0.04430 |
| Chorismate biosynthesis I | General Biosynthetic Pathways | *Parabacteroides* | *Parabacteroides goldsteinii* | CD8^+^ Naive | CD8^+^ T Cells | 4.03E-11 | 9.73E-12 | | 0.04430 |
| Coenzyme A biosynthesis I (prokaryotic) | Cofactor & Vitamin Biosynthesis | *Parabacteroides* | *Parabacteroides goldsteinii* | CD8^+^ Naive | CD8^+^ T Cells | 3.64E-11 | 9.20E-12 | | 0.04430 |
| Superpathway of coenzyme A biosynthesis III (mammals) | Cofactor & Vitamin Biosynthesis | *Parabacteroides* | *Parabacteroides goldsteinii* | CD8^+^ Naive | CD8^+^ T Cells | 3.87E-11 | 9.41E-12 | | 0.04430 |
| L-histidine biosynthesis | Amino Acid Biosynthesis | *Parabacteroides* | *Parabacteroides goldsteinii* | CD8^+^ Naive | CD8^+^ T Cells | 3.13E-11 | 7.87E-12 | | 0.04430 |
| Methylerythritol phosphate pathway I | General Biosynthetic Pathways | *Parabacteroides* | *Parabacteroides goldsteinii* | CD8^+^ Naive | CD8^+^ T Cells | 3.56E-11 | 8.77E-12 | | 0.04430 |
| Phosphopantothenate biosynthesis I | Lipid & Membrane Metabolism | *Parabacteroides* | *Parabacteroides goldsteinii* | CD8^+^ Naive | CD8^+^ T Cells | 4.05E-11 | 1.01E-11 | | 0.04430 |
| Peptidoglycan biosynthesis I (meso-diaminopimelate containing) | Cell wall & Peptidoglycan Synthesis | *Parabacteroides* | *Parabacteroides goldsteinii* | CD8^+^ Naive | CD8^+^ T Cells | 4.02E-11 | 9.80E-12 | | 0.04430 |
| Folate transformations II (plants) | Cofactor & Vitamin Biosynthesis | *Parabacteroides* | *Parabacteroides goldsteinii* | CD8^+^ Naive | CD8^+^ T Cells | 4.29E-11 | 1.08E-11 | | 0.04430 |
| L-lysine biosynthesis VI | Amino Acid Biosynthesis | *Parabacteroides* | *Parabacteroides goldsteinii* | CD8^+^ Naive | CD8^+^ T Cells | 3.38E-11 | 8.39E-12 | | 0.04430 |
| UMP biosynthesis I | Nucleotide Metabolism & Biosynthesis | *Parabacteroides* | *Parabacteroides goldsteinii* | CD8^+^ Naive | CD8^+^ T Cells | 4.67E-11 | 1.16E-11 | | 0.04430 |
| 5-aminoimidazole ribonucleotide biosynthesis I | Nucleotide Metabolism & Biosynthesis | *Parabacteroides* | *Parabacteroides goldsteinii* | CD8^+^ Naive | CD8^+^ T Cells | 3.84E-11 | 9.34E-12 | | 0.04430 |
| 5-aminoimidazole ribonucleotide biosynthesis II | Nucleotide Metabolism & Biosynthesis | *Parabacteroides* | *Parabacteroides goldsteinii* | CD8^+^ Naive | CD8^+^ T Cells | 4.04E-11 | 9.75E-12 | | 0.04430 |
| Chorismate biosynthesis from 3-dehydroquinate | General Biosynthetic Pathways | *Parabacteroides* | *Parabacteroides goldsteinii* | CD8^+^ Naive | CD8^+^ T Cells | 4.48E-11 | 1.08E-11 | | 0.04430 |
| Superpathway of 5-aminoimidazole ribonucleotide biosynthesis | Nucleotide Metabolism & Biosynthesis | *Parabacteroides* | *Parabacteroides goldsteinii* | CD8^+^ Naive | CD8^+^ T Cells | 4.04E-11 | 9.75E-12 | | 0.04430 |
| Peptidoglycan biosynthesis III (mycobacteria) | Cell wall & Peptidoglycan Synthesis | *Parabacteroides* | *Parabacteroides goldsteinii* | CD8^+^ Naive | CD8^+^ T Cells | 4.26E-11 | 1.03E-11 | | 0.04430 |
| Adenine and adenosine salvage III | Nucleotide Metabolism & Biosynthesis | *Parabacteroides* | *Parabacteroides goldsteinii* | CD8^+^ Naive | CD8^+^ T Cells | 4.15E-11 | 9.99E-12 | | 0.04430 |
| Queuosine biosynthesis I (de novo) | Nucleotide Metabolism & Biosynthesis | *Parabacteroides* | *Parabacteroides goldsteinii* | CD8^+^ Naive | CD8^+^ T Cells | 4.47E-11 | 1.12E-11 | | 0.04430 |
| Guanosine ribonucleotides de novo biosynthesis | Nucleotide Metabolism & Biosynthesis | *Parabacteroides* | *Parabacteroides goldsteinii* | CD8^+^ Naive | CD8^+^ T Cells | 4.57E-11 | 1.17E-11 | | 0.04430 |
| UMP biosynthesis II | Nucleotide Metabolism & Biosynthesis | *Parabacteroides* | *Parabacteroides goldsteinii* | CD8^+^ Naive | CD8^+^ T Cells | 4.67E-11 | 1.16E-11 | | 0.04430 |
| UMP biosynthesis III | Nucleotide Metabolism & Biosynthesis | *Parabacteroides* | *Parabacteroides goldsteinii* | CD8^+^ Naive | CD8^+^ T Cells | 4.69E-11 | 1.18E-11 | | 0.04430 |
| Coenzyme A biosynthesis II (eukaryotic) | Cofactor & Vitamin Biosynthesis | *Parabacteroides* | *Parabacteroides goldsteinii* | CD8^+^ Naive | CD8^+^ T Cells | 3.71E-11 | 9.07E-12 | | 0.04430 |
| Fatty acid biosynthesis initiation (mitochondria) | Lipid & Membrane Metabolism | *Parabacteroides* | *Parabacteroides goldsteinii* | CD8^+^ Naive | CD8^+^ T Cells | 3.32E-11 | 8.41E-12 | | 0.04430 |
| Flavin biosynthesis I (bacteria and plants) | Cofactor & Vitamin Biosynthesis | *Parabacteroides* | *Parabacteroides goldsteinii* | CD8^+^ Naive | CD8^+^ T Cells | 3.75E-11 | 9.16E-12 | | 0.04430 |
| tRNA charging | Amino Acid Biosynthesis | *Parabacteroides* | *Parabacteroides goldsteinii* | CD8^+^ Naive | CD8^+^ T Cells | 3.47E-11 | 8.74E-12 | | 0.04430 |
| L-valine biosynthesis | Amino Acid Biosynthesis | *Parabacteroides* | *Parabacteroides goldsteinii* | CD8^+^ Naive | CD8^+^ T Cells | 3.58E-11 | 9.17E-12 | | 0.04430 |
| 5-aminoimidazole ribonucleotide biosynthesis I | Nucleotide Metabolism & Biosynthesis | *Holdemanella* | *Holdemanella biformis* | CCR7^+^ Activated NK Cells | Natural Killer (NK) Cells | 5.36E-09 | 1.53E-09 | | 0.04449 |
| Adenine and adenosine salvage III | Nucleotide Metabolism & Biosynthesis | *Coprobacter* | *Coprobacter fastidiosus* | IFNγ/TNFα-producing CD69^+^ CD4^+^ T Cells | CD4^+^ T Cells | 3.82E-11 | 1.04E-11 | | 0.04466 |
| Queuosine biosynthesis I (de novo) | Nucleotide Metabolism & Biosynthesis | *Escherichia* | *Escherichia coli* | IL-2-producing Activated CD69^+^ CD4^+^ T Cells | CD4^+^ T Cells | 3.01E-10 | 7.55E-11 | | 0.04507 |
| UMP biosynthesis I | Nucleotide Metabolism & Biosynthesis | *Coprobacter* | *Coprobacter fastidiosus* | IFNγ-producing Activated NK Cells | Natural Killer (NK) Cells | 1.48E-10 | 3.50E-11 | | 0.04548 |
| UMP biosynthesis II | Nucleotide Metabolism & Biosynthesis | *Coprobacter* | *Coprobacter fastidiosus* | IFNγ-producing Activated NK Cells | Natural Killer (NK) Cells | 1.48E-10 | 3.50E-11 | | 0.04548 |
| UMP biosynthesis III | Nucleotide Metabolism & Biosynthesis | *Coprobacter* | *Coprobacter fastidiosus* | IFNγ-producing Activated NK Cells | Natural Killer (NK) Cells | 1.48E-10 | 3.50E-11 | | 0.04548 |
| L-valine biosynthesis | Amino Acid Biosynthesis | *Coprobacter* | *Coprobacter fastidiosus* | IFNγ-producing Activated NK Cells | Natural Killer (NK) Cells | 1.24E-10 | 2.94E-11 | | 0.04548 |
| Peptidoglycan biosynthesis III (mycobacteria) | Cell wall & Peptidoglycan Synthesis | *Dialister* | *Dialister invisus* | IFNγ-producing T regs | Adaptive Immune Cells | 1.84E-09 | 5.29E-10 | | 0.04619 |
| CDP diacylglycerol biosynthesis I | Lipid & Membrane Metabolism | *Clostridium* | *Clostridium sp AM22 11AC* | CD8^+^ CM | CD8^+^ T Cells | 9.90E-11 | 2.57E-11 | | 0.04635 |
| CDP diacylglycerol biosynthesis II | Lipid & Membrane Metabolism | *Clostridium* | *Clostridium sp AM22 11AC* | CD8^+^ CM | CD8^+^ T Cells | 9.90E-11 | 2.57E-11 | | 0.04635 |
| Fatty acid biosynthesis initiation (mitochondria) | Lipid & Membrane Metabolism | *Clostridium* | *Clostridium sp AM22 11AC* | CD8^+^ CM | CD8^+^ T Cells | 1.54E-10 | 4.00E-11 | | 0.04636 |
| Fatty acid biosynthesis initiation (mitochondria) | Lipid & Membrane Metabolism | *Clostridium* | *Clostridium sp AM22 11AC* | CD8^+^ Naive | CD8^+^ T Cells | 6.74E-11 | 1.75E-11 | | 0.04656 |
| Queuosine biosynthesis I (de novo) | Nucleotide Metabolism & Biosynthesis | *Clostridium* | *Clostridium sp AM22 11AC* | CD8^+^ Naive | CD8^+^ T Cells | 5.32E-11 | 1.39E-11 | | 0.04656 |
| Inosine 5-phosphate degradation | Nucleotide Metabolism & Biosynthesis | *Escherichia* | *Escherichia coli* | IL-2/TNFα-producing CD4^+^ EM T Cells | CD4^+^ T Cells | 1.79E-10 | 4.72E-11 | | 0.04713 |
| Purine ribonucleosides degradation | Nucleotide Metabolism & Biosynthesis | *Clostridium* | *Clostridium sp AM22 11AC* | CCR7^+^ CD8^+^ T Cells | CD8^+^ T Cells | 4.30E-11 | 1.20E-11 | | 0.04751 |
| L-lysine biosynthesis VI | Amino Acid Biosynthesis | *Coprobacter* | *Coprobacter fastidiosus* | IFNγ-producing NK Cells | Natural Killer (NK) Cells | 1.50E-10 | 3.49E-11 | | 0.04832 |
| Guanosine ribonucleotides de novo biosynthesis | Nucleotide Metabolism & Biosynthesis | *Coprobacter* | *Coprobacter fastidiosus* | IFNγ-producing NK Cells | Natural Killer (NK) Cells | 8.61E-11 | 2.01E-11 | | 0.04832 |
| Inosine 5-phosphate degradation | Nucleotide Metabolism & Biosynthesis | *Dialister* | *Dialister invisus* | IL-2/TNFα-producing CD4^+^ T Cells | CD4^+^ T Cells | 1.48E-10 | 3.81E-11 | | 0.04842 |
| L-valine biosynthesis | Amino Acid Biosynthesis | *Clostridium* | *Clostridium sp AM22 11AC* | IFNγ-producing T regs | Adaptive Immune Cells | 1.21E-09 | 3.51E-10 | | 0.04852 |
| CDP diacylglycerol biosynthesis I | Lipid & Membrane Metabolism | *Coprobacter* | *Coprobacter fastidiosus* | IFNγ/TNFα-producing CD69^+^ CD8^+^ T Cells | CD8^+^ T Cells | 1.61E-10 | 4.40E-11 | | 0.04869 |
| CDP diacylglycerol biosynthesis II | Lipid & Membrane Metabolism | *Coprobacter* | *Coprobacter fastidiosus* | IFNγ/TNFα-producing CD69^+^ CD8^+^ T Cells | CD8^+^ T Cells | 1.61E-10 | 4.40E-11 | | 0.04869 |
| Superpathway of coenzyme A biosynthesis III (mammals) | Cofactor & Vitamin Biosynthesis | *Dialister* | *Dialister invisus* | IL-2/TNFα-producing CD4^+^ EM T Cells | CD4^+^ T Cells | 5.25E-10 | 1.40E-10 | | 0.04903 |
| Coenzyme A biosynthesis II (eukaryotic) | Cofactor & Vitamin Biosynthesis | *Dialister* | *Dialister invisus* | IL-2/TNFα-producing CD4^+^ EM T Cells | CD4^+^ T Cells | 5.10E-10 | 1.36E-10 | | 0.04903 |
| Folate transformations II (plants) | Cofactor & Vitamin Biosynthesis | *Eubacterium* | *Eubacterium ventriosum* | TNFα-producing gd T Cells | Adaptive Immune Cells | 3.01E-11 | 8.80E-12 | | 0.04905 |
| Folate transformations II (plants) | Cofactor & Vitamin Biosynthesis | *Coprobacter* | *Coprobacter fastidiosus* | IFNγ/TNFα-producing CD69^+^ CD4^+^ T Cells | CD4^+^ T Cells | 3.40E-11 | 9.51E-12 | | 0.04932 |
| L-lysine biosynthesis VI | Amino Acid Biosynthesis | *Dialister* | *Dialister invisus* | IFNγ-producing T regs | Adaptive Immune Cells | 1.67E-09 | 4.83E-10 | | 0.04945 |
| Superpathway of coenzyme A biosynthesis III (mammals) | Cofactor & Vitamin Biosynthesis | *Coprobacter* | *Coprobacter fastidiosus* | IFNγ-producing CD161^+^ CD4^+^ T Cells | CD4^+^ T Cells | 4.47E-11 | 1.22E-11 | | 0.04945 |
| UMP biosynthesis I | Nucleotide Metabolism & Biosynthesis | *Eubacterium* | *Eubacterium ventriosum* | PMDSCs | Innate Immune Cells | 7.6E-11 | 2.23E-11 | | 0.04945 |
| UMP biosynthesis II | Nucleotide Metabolism & Biosynthesis | *Eubacterium* | *Eubacterium ventriosum* | PMDSCs | Innate Immune Cells | 7.6E-11 | 2.23E-11 | | 0.04945 |
| UMP biosynthesis III | Nucleotide Metabolism & Biosynthesis | *Eubacterium* | *Eubacterium ventriosum* | PMDSCs | Innate Immune Cells | 7.6E-11 | 2.23E-11 | | 0.04945 |
| Sucrose biosynthesis II | General Biosynthetic Pathways | *Clostridium* | *Clostridium sp AM22 11AC* | CD8^+^ Naive | CD8^+^ T Cells | 8.09E-11 | 2.13E-11 | | 0.04959 |
| Phosphopantothenate biosynthesis I | Lipid & Membrane Metabolism | *Eubacterium* | *Eubacterium ventriosum* | CD8^+^ Naive | CD8^+^ T Cells | 2.26E-11 | 5.95E-12 | | 0.04959 |

Differential abundance analyses were conducted using the R package MaAsLin2 (v1.15.1), employing linear regressions adjusted for age, sex, and BMI. Relative abundances were normalised using the centred log-ratio (CLR) method. Adjustments were performed using the Benjamini–Hochberg False FDR method, and all results presented meet the significance threshold of q<0.05. CCR7, C-C chemokine receptor type 7; CM, central memory; EM, effector memory; HLA-DR, human leukocyte antigen-DR isotype; IFN-γ, interferon-γ; IL-, interleukin-; NK, natural killer; PD-1, programmed cell death protein 1; PMDSCs, polymorphonuclear myeloid-derived suppressor cells; T regs, T regulatory cells; TEMRA, terminally differentiated effector memory T cells; TNF-α, tumour necrosis factor-α.

**Table S9. Surface and intracellular antibody panel for participant peripheral blood mononuclear cell (PBMC) immunoprofiling.**

| **Antibody** | **Fluorochrome** | **Clone** | **Surface or Intracellular** | **Supplier** |
| --- | --- | --- | --- | --- |
| cFluor® CD161 | R780 | HP-3G10 | Surface | Cytek |
| cFluor® CD56 | YG584 | 22URTI | Surface | Cytek |
| cFluor® TCRγδ | BYG710 | B1 | Surface | Cytek |
| CD11c | BV510 | SHCL-3 | Surface | BD Biosciences |
| CD14 | BUV737 | M5E2 | Surface | BD Biosciences |
| CD15 | BUV661 | 7C3.rMAb | Surface | BD Biosciences |
| CD16 | BUV496 | 3G8 | Surface | BD Biosciences |
| CD19 | BV650 | HIB19 | Surface | BioLegend |
| CD27 | BUV563 | M-T271 | Surface | BD Biosciences |
| cFluor® CCR7 | R685 | G043H7 | Surface | Cytek |
| cFluor® CD45RA | B520 | HI100 | Surface | Cytek |
| HLA-DR | BUV395 | L243 | Surface | BD Biosciences |
| CD25 | BV421 | BC96 | Intracellular | BD Biosciences |
| CD366 (TIM-3) | BV711 | 7D3 | Intracellular | BD Biosciences |
| CD69 | PE-Cy7 | FN50 | Intracellular | BioLegend |
| cFluor® CD3 | B548 | SK7 | Intracellular | Cytek |
| cFluor® CD4 | R840 | SK3 | Intracellular | Cytek |
| cFluor® CD8 | B690 | RPA-T8 | Intracellular | Cytek |
| cFluor® FoxP3 | BYG667 | PCH101 | Intracellular | Cytek |
| cFluor® GM-CSF | BV450 | BVD2-21C11 | Intracellular | Cytek |
| cFluor® IL-10 | R659 | JES3-9D7 | Intracellular | Cytek |
| cFluor® IL-17A | BYG610 | eBio64DEC17 | Intracellular | Cytek |
| cFluor® IL-22 | BYG575 | 22URTI | Intracellular | Cytek |
| cFluor® TNF-α | R720 | MAb11 | Intracellular | Cytek |
| CD152 (CTLA-4) | BUV805 | BNI3 | Intracellular | BD Biosciences |
| CX3CR1 | BV480 | 2A9-1 | Intracellular | BD Biosciences |
| IFN-γ | BV605 | B27 | Intracellular | BioLegend |
| IL-2 | BV750 | MQ1-17H12 | Intracellular | BD Biosciences |
| CD279 (PD-1) | BV786 | MIH4 | Intracellular | BD Biosciences |

**Table S10. Summary of the immune cell populations, activated sub-types and cytokine production with corresponding gating definitions.**

| **Cell Type** | **Class** | **Markers/Gating** |
| --- | --- | --- |
| **Mature LDNs** | Myeloid | CD14^-^ CD15^+^ CD16^+^ |
| **cDCs** | Myeloid | CD3^-^ CD56^-^ CD14^-^ CD16^-^ CD11c^+^ HLA-DR^+^ |
| **Classical monocytes (CM)** | Myeloid | CD3^-^ CD56^-^ CD14^+^ CD16^-^ |
| **Intermediate monocytes (IM)** | Myeloid | CD3^-^ CD56^-^ CD14^+^ CD16^+^ |
| **Nonclassical monocytes (non-CM)** | Myeloid | CD3^-^ CD56^-^ CD14^-^ CD16^+^ |
| **Monocytic myeloid-derived suppressor cells (MDSCs)** | Myeloid | CD14^+^ CD15^-^ HLA-DR^-^ |
| **Polymorphonuclear MDSCs** | Myeloid | CD15^+^ CD14^-^ HLA-DR^-^ |
| **B cells** | Lymphoid | CD3^-^ CD19^+^ |
| TNFα-producing | Lymphoid | CD3^-^ CD19^+^ TNFα^+^ |
| *Activated B cells* | Lymphoid | CD3^-^ CD19^+^ CD27^+^ |
| TNFα-producing | Lymphoid | CD3^-^ CD19^+^ CD27^+^ TNFα^+^ |
| **Natural Killer (NK) T cells** | Lymphoid | CD3^+^ CD56^+^ |
| IFNγ-producing | Lymphoid | CD3^+^ CD56^+^ IFNγ^+^ |
| TNFα-producing | Lymphoid | CD3^+^ CD56^+^ TNFα^+^ |
| **NK cells** | Lymphoid | CD3^-^ CD56^+^ |
| CCR7^+^ | Lymphoid | CD3^-^ CD56^+^ CCR7^+^ |
| CX3CR1^+^ | Lymphoid | CD3^-^ CD56^+^ CX3CR1^+^ |
| IFNγ-producing | Lymphoid | CD3^-^ CD56^+^ IFNγ^+^ |
| IL17A-producing | Lymphoid | CD3^-^ CD56^+^ IL-17A^+^ |
| TNFα-producing | Lymphoid | CD3^-^ CD56^+^ TNFα^+^ |
| CD161^+^ NK cells | Lymphoid | CD3^-^ CD56^+^ CD161^+^ |
| *Activated NK cells* | Lymphoid | CD3^-^ CD56^+^ CD69^+^ |
| CCR7^+^ | Lymphoid | CD3^-^ CD56^+^ CD69^+^ CCR7^+^ |
| IFNγ-producing | Lymphoid | CD3^-^ CD56^+^ CD69^+^ IFNγ^+^ |
| IL17A-producing | Lymphoid | CD3^-^ CD56^+^ CD69^+^ IL-17A^+^ |
| TNFα-producing | Lymphoid | CD3^-^ CD56^+^ CD69^+^ TNFα^+^ |
| **Cytotoxic T cells** | Lymphoid | CD3^+^ CD56^-^ CD8^+^ |
| **CD8^+^ Naïve** | Lymphoid | CD3^+^ CD56^-^ CD8^+^ CD45RA^+^ CCR7^+^ |
| **CD8^+^ central memory** | Lymphoid | CD3^+^ CD56^-^ CD8^+^ CD45RA^-^ CCR7^+^ |
| CCR7^+^ CD8^+^ T cells | Lymphoid | CD3^+^ CD56^-^ CD8^+^ CCR7^+^ |
| CTLA-4^+^ CD8^+^ T cells | Lymphoid | CD3^+^ CD56^-^ CD8^+^ CTLA-4^+^ |
| PD-1^+^ CD8^+^ T cells | Lymphoid | CD3^+^ CD56^-^ CD8^+^ PD-1^+^ |
| IFNγ-producing CD8^+^ T cells | Lymphoid | CD3^+^ CD56^-^ CD8^+^ IFNγ^+^ |
| IL2-producing CD8^+^ T cells | Lymphoid | CD3^+^ CD56^-^ CD8^+^ IL-2^+^ |
| IL17A-producing CD8^+^ T cells | Lymphoid | CD3^+^ CD56^-^ CD8^+^ IL-17A^+^ |
| TNFα-producing CD8^+^ T cells | Lymphoid | CD3^+^ CD56^-^ CD8^+^ TNFα^+^ |
| *Activated CD8^+^ (early)* | Lymphoid | CD3^+^ CD56^-^ CD8^+^ CD69^+^ |
| IFNγ-producing | Lymphoid | CD3^+^ CD56^-^ CD8^+^ CD69^+^ IFNγ^+^ |
| IL2-producing | Lymphoid | CD3^+^ CD56^-^ CD8^+^ CD69^+^ IL-2^+^ |
| IL17A-producing | Lymphoid | CD3^+^ CD56^-^ CD8^+^ CD69^+^ IL-17A^+^ |
| TNFα-producing | Lymphoid | CD3^+^ CD56^-^ CD8^+^ CD69^+^ TNFα^+^ |
| CTLA-4^+^ | Lymphoid | CD3^+^ CD56^-^ CD8^+^ CD69^+^ CTLA-4^+^ |
| PD-1^+^ | Lymphoid | CD3^+^ CD56^-^ CD8^+^ CD69^+^ PD-1^+^ |
| *Activated CD8^+^ T Cells (late)* | Lymphoid | CD3^+^ CD56^-^ CD8^+^ HLA-DR^+^ |
| IFNγ-producing | Lymphoid | CD3^+^ CD56^-^ CD8^+^ HLA-DR^+^ IFNγ^+^ |
| IL2-producing | Lymphoid | CD3^+^ CD56^-^ CD8^+^ HLA-DR^+^ IL-2^+^ |
| IL17A-producing | Lymphoid | CD3^+^ CD56^-^ CD8^+^ HLA-DR^+^ IL-17A^+^ |
| TNFα-producing | Lymphoid | CD3^+^ CD56^-^ CD8^+^ HLA-DR^+^ TNFα^+^ |
| CTLA-4^+^ | Lymphoid | CD3^+^ CD56^-^ CD8^+^ HLA-DR^+^ CTLA-4^+^ |
| PD-1^+^ | Lymphoid | CD3^+^ CD56^-^ CD8^+^ HLA-DR^+^ PD-1^+^ |
| **Mucosal-Associated Invariant (MAIT) T cells** | Lymphoid | CD3^+^ CD56^-^ CD8^+^ CD161^+^ |
| IFNγ-producing | Lymphoid | CD3^+^ CD56^-^ CD8^+^ CD161^+^ IFNγ^+^ |
| IL2-producing | Lymphoid | CD3^+^ CD56^-^ CD8^+^ CD161^+^ IL-2^+^ |
| IL17A-producing | Lymphoid | CD3^+^ CD56^-^ CD8^+^ CD161^+^ IL-17A^+^ |
| TNFα-producing | Lymphoid | CD3^+^ CD56^-^ CD8^+^ CD161^+^ TNFα^+^ |
| **CD8^+^ EM (effector memory)** | Lymphoid | CD3^+^ CD56^-^ CD8^+^ CD45RA^-^ CCR7^-^ |
| IFNγ-producing | Lymphoid | CD3^+^ CD56^-^ CD8^+^ CD45RA^-^ CCR7^-^ IFNγ^+^ |
| IL2-producing | Lymphoid | CD3^+^ CD56^-^ CD8^+^ CD45RA^-^ CCR7^-^ IL-2^+^ |
| IL17A-producing | Lymphoid | CD3^+^ CD56^-^ CD8^+^ CD45RA^-^ CCR7^-^ IL-17A^+^ |
| TNFα-producing | Lymphoid | CD3^+^ CD56^-^ CD8^+^ CD45RA^-^ CCR7^-^ TNFα^+^ |
| **Helper T cells (Th)** | Lymphoid | CD3^+^ CD56^-^ CD4^+^ |
| **CD4^+^ TEMRA** | Lymphoid | CD3^+^ CD56^-^ CD4^+^ CD45RA^+^ CCR7^-^ |
| **CD4^+^ Naïve** | Lymphoid | CD3^+^ CD56^-^ CD4^+^ CD45RA^+^ CCR7^+^ |
| **CD4^+^ (CM) central memory** | Lymphoid | CD3^+^ CD56^-^ CD4^+^ CD45RA^-^ CCR7^+^ |
| CCR7^+^ CD4^+^ T cells | Lymphoid | CD3^+^ CD56^-^ CD4^+^ CCR7^+^ |
| IFNγ-producing | Lymphoid | CD3^+^ CD56^-^ CD4^+^ IFNγ^+^ |
| IL2-producing | Lymphoid | CD3^+^ CD56^-^ CD4^+^ IL-2^+^ |
| IL17A-producing | Lymphoid | CD3^+^ CD56^-^ CD4^+^ IL-17A^+^ |
| TNFα-producing | Lymphoid | CD3^+^ CD56^-^ CD4^+^ TNFα^+^ |
| CTLA-4^+^ | Lymphoid | CD3^+^ CD56^-^ CD4^+^ CTLA-4^+^ |
| PD-1^+^ | Lymphoid | CD3^+^ CD56^-^ CD4^+^ PD-1^+^ |
| *Activated CD4^+^ T cells (early)* | Lymphoid | CD3^+^ CD56^-^ CD4^+^ CD69^+^ |
| IFNγ-producing | Lymphoid | CD3^+^ CD56^-^ CD4^+^ CD69^+^ IFNγ^+^ |
| IL2-producing | Lymphoid | CD3^+^ CD56^-^ CD4^+^ CD69^+^ IL-2^+^ |
| IL17A-producing | Lymphoid | CD3^+^ CD56^-^ CD4^+^ CD69^+^ IL-17A^+^ |
| TNFα-producing | Lymphoid | CD3^+^ CD56^-^ CD4^+^ CD69^+^ TNFα^+^ |
| CTLA-4^+^ | Lymphoid | CD3^+^ CD56^-^ CD4^+^ CD69^+^ CTLA-4^+^ |
| PD-1^+^ | Lymphoid | CD3^+^ CD56^-^ CD4^+^ CD69^+^ PD-1^+^ |
| *Activated CD4^+^ T cells (late)* | Lymphoid | CD3^+^ CD56^-^ CD4^+^ HLA-DR^+^ |
| IFNγ-producing | Lymphoid | CD3^+^ CD56^-^ CD4^+^ HLA-DR^+^ IFNγ^+^ |
| IL2-producing | Lymphoid | CD3^+^ CD56^-^ CD4^+^ HLA-DR^+^ IL-2^+^ |
| IL17A-producing | Lymphoid | CD3^+^ CD56^-^ CD4^+^ HLA-DR^+^ IL-17A^+^ |
| TNFα-producing | Lymphoid | CD3^+^ CD56^-^ CD4^+^ HLA-DR^+^ TNFα^+^ |
| CTLA-4^+^ | Lymphoid | CD3^+^ CD56^-^ CD4^+^ HLA-DR^+^ CTLA-4^+^ |
| PD-1^+^ | Lymphoid | CD3^+^ CD56^-^ CD4^+^ HLA-DR^+^ PD-1^+^ |
| CD161^+^ CD4^+^ T cells | Lymphoid | CD3^+^ CD56^-^ CD4^+^ CD161^+^ |
| IFNγ-producing | Lymphoid | CD3^+^ CD56^-^ CD4^+^ CD161^+^ IFNγ^+^ |
| IL2-producing | Lymphoid | CD3^+^ CD56^-^ CD4^+^ CD161^+^ IL-2^+^ |
| IL17A-producing | Lymphoid | CD3^+^ CD56^-^ CD4^+^ CD161^+^ IL-17A^+^ |
| TNFα-producing | Lymphoid | CD3^+^ CD56^-^ CD4^+^ CD161^+^ TNFα^+^ |
| **CD4^+^ EM** | Lymphoid | CD3^+^ CD56^-^ CD4^+^ CD45RA^-^ CCR7^-^ |
| IFNγ-producing | Lymphoid | CD3^+^ CD56^-^ CD4^+^ CD45RA^-^ CCR7^-^ IFNγ^+^ |
| IL2-producing | Lymphoid | CD3^+^ CD56^-^ CD4^+^ CD45RA^-^ CCR7^-^ IL-2^+^ |
| IL17A-producing | Lymphoid | CD3^+^ CD56^-^ CD4^+^ CD45RA^-^ CCR7^-^ IL-17A^+^ |
| TNFα-producing | Lymphoid | CD3^+^ CD56^-^ CD4^+^ CD45RA^-^ CCR7^-^ TNFα^+^ |
| **T regs (T regulatory cells)** | Lymphoid | CD3^+^ CD56^-^ CD4^+^ FoxP3^+^ |
| IFNγ-producing | Lymphoid | CD3^+^ CD56^-^ CD4^+^ FoxP3^+^ IFNγ^+^ |
| IL2-producing | Lymphoid | CD3^+^ CD56^-^ CD4^+^ FoxP3^+^ IL-2^+^ |
| IL17A-producing | Lymphoid | CD3^+^ CD56^-^ CD4^+^ FoxP3^+^ IL-17A^+^ |
| TNFα-producing | Lymphoid | CD3^+^ CD56^-^ CD4^+^ FoxP3^+^ TNFα^+^ |
| **γδ T cells** | Lymphoid | CD3^+^ TCRγδ^+^ |
| IFNγ-producing | Lymphoid | CD3^+^ TCRγδ^+^ IFNγ^+^ |
| IL2-producing | Lymphoid | CD3^+^ TCRγδ^+^ IL-2^+^ |
| IL17A-producing | Lymphoid | CD3^+^ TCRγδ^+^ IL-17A^+^ |
| IL22-producing | Lymphoid | CD3^+^ TCRγδ^+^ IL-22^+^ |
| TNFα-producing | Lymphoid | CD3^+^ TCRγδ^+^ TNFα^+^ |
